# A Systematic Review and Meta-Analysis: Research Using the Autism Polygenic Score

**DOI:** 10.1101/2024.03.08.24303918

**Authors:** M.M. de Wit, M.J. Morgan, I. Libedinsky, C. Austerberry, S. Begeer, A. Abdellaoui, A. Ronald, T.J.C. Polderman

## Abstract

**Objective:** Genetic factors play a substantial role in the etiology of autism and its co-occurrence with other conditions and traits. The autism polygenic score, derived from the latest autism case-control meta-genome-wide association studies, captures some of the accumulated influence of common genetic variants on autism. We reviewed and meta-analyzed published studies that assessed the relationship between this autism polygenic score and autism diagnosis, and autistic, behavioral and neurobiological traits.

**Method:** Systematically searching public databases, we identified 72 studies and > 750 outcome measures. Included studies received a quality assessment.

**Results:** The majority of included studies were rated as good quality. The autism polygenic score was most strongly associated with autism diagnosis (meta-analytic *r* = .162, 95% CI .066 – .258). The autism polygenic score was also significantly associated with autistic traits but to a lesser degree than for autism (meta-analytic *r* = .042 (95% CI .004 – .081). Associations with other outcomes were inconsistent and meta-analytic effect sizes were generally small (median *r* = .03).

**Conclusion:** We conclude that the current autism polygenic score is consistently associated with autism diagnostic status and autistic traits, but overlap between autism and other traits and conditions is not, from publications to date, explained significantly by the autism polygenic score. When compared to other mental conditions, autism is phenotypically and etiologically heterogeneous, which might drive the relatively modest associations observed with the autism polygenic score to date.

## Introduction

Autism Spectrum Disorder (ASD; from here on autism as per Bottema-Beutel^1^) is a complex neurodevelopmental condition that is characterized by atypical social interaction and communication, repetitive behavior and focused interests, and sensory sensitivities. The prevalence of autism is estimated at 1-2% of the population worldwide^2^. Genetic factors substantially contribute to autism’s etiology. For example, a meta-analysis of twin and family studies indicated that autism is approximately 64 – 91% heritable^3^.

Autism shows considerable co-occurrence with developmental, psychiatric and somatic conditions and a range of other traits. The incidence of other psychiatric diagnoses in autistic individuals is estimated around 70%^4^. Autistic individuals^1^ also have increased liability for a range of physical conditions^7^. Shared genetic influences between autism, autistic traits and neurodevelopmental, psychiatric and physical conditions may explain part of this phenotypic co-occurrence^8,9^.

Genome-wide association studies (GWASs) assess the relationship between a large number of common genetic variants spread across the human genome and phenotypic outcomes of interest. The latest autism meta-GWAS by Grove et al. (2019)^10^ identified 5 independent common genetic loci that were significantly associated with autism and revealed a single nucleotide polymorphism (SNP)-based heritability, which reflects the proportion of phenotypic variance that is explained by additive effects of genomic variants included in the GWAS, of 11.8%. However, many common genetic loci beyond those that are genome-wide significant are involved in autism’s etiology^10^.

Polygenic scores use the summary statistics from GWAS to capture part of the combined influence of common genetic influences on an outcome. They reflect the effect of multiple genetic variants with small individual effects in a single value. They are calculated by summing together an individual’s common genetic variants, weighted by their effect size. Polygenic scores are one of the tools that can be employed to increase understanding of the genetic basis of complex traits such as autism, and overlap with other traits^11^, amongst many other popular methods in molecular genetic studies^12^. In addition to their scientific value, polygenic scores have potential clinical value in the future in addition to existing diagnostic tools^13^. However, at present, most psychiatric polygenic scores are not accurate for individuals, and bioethical issues relating to the application of polygenic scores need to be carefully planned^13^.

Although psychiatric phenotypes have long been among the most studied in genetics^14^, only few studies systematically studied reported polygenic score associations for a specific neuropsychiatric classification with related traits and conditions^15–18^. These studies report consistent associations between the ADHD polygenic score and ADHD diagnosis (OR range 1.22% to 1.76%), as well as other traits and diagnoses^15,16,18^ and between the schizophrenia polygenic score and other traits and disorders^17^.

The latest autism GWAS led to a wealth of studies assessing the relationship between the autism polygenic score and autism diagnosis, autistic traits, and a wide variety of other related traits. Here, we systematically reviewed and meta-analyzed all published studies that assessed the relationship between the autism polygenic scores and all available outcome measures, in order to understand whether the current autism polygenic score can explain phenotypic and genetic overlap between autism and co-occurring conditions and traits.

## Methods

This study was pre-registered with PROSPERO Framework under reference number CRD42022307993. Primary analyses were carried out according to the preregistration. We performed additional analyses, of which details are described under ‘secondary analyses’.

### Study Selection

We aimed to include all studies assessing the relationship between the autism polygenic score based on the most recent GWAS results^10^ and autism diagnostic status, autistic traits, and behavioral and neurobiological traits. A first systematic search of PubMed, Web of Science, Psychinfo and Scopus for published, peer-reviewed studies in the English language was performed on November 2, 2022 and a second search on January 6, 2023. The search terms per search engine are given in sTable 2, available online. Studies were excluded if: 1) The predictor was not an autism polygenic score based on the latest autism GWAS^10^; 2) The polygenic score was not based on genome-wide results, but included only a selection of SNPs; 3) The study was a review; 4) No direct associations between the autism polygenic score and an outcome variable were reported (e.g., polygenic transmission tests (pTDT), interaction effects, genetic correlations, multivariate associations). Abstracts were inspected by MdW. A second reviewer (TP) was involved in case of any doubt regarding inclusion. Figure 1 shows the full selection procedure.

**Figure 1.**
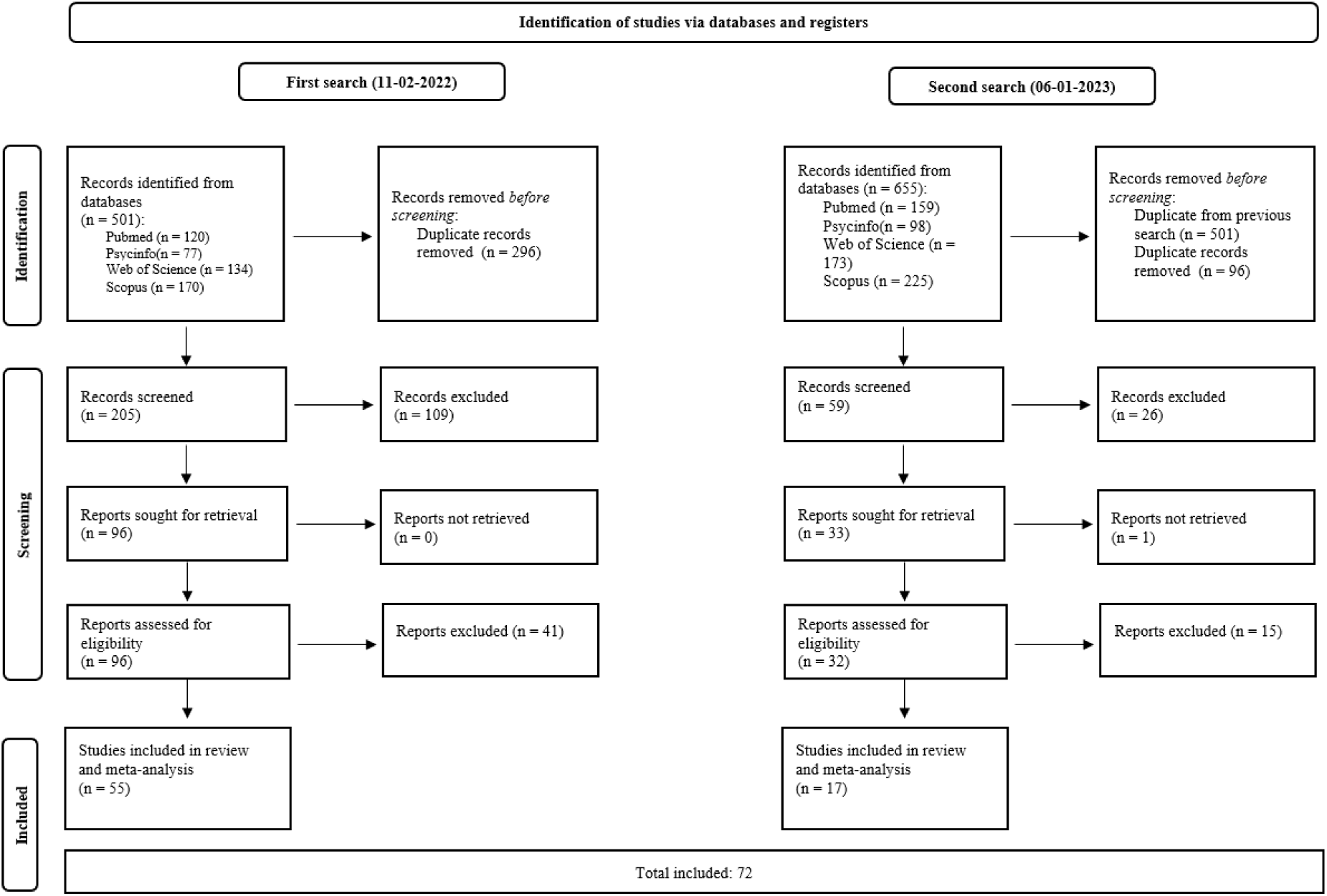
PRISMA Flow Diagram

### Quality assessment

To evaluate the quality, validity and generalizability of our included studies, we reviewed each study against a set of quality assessment criteria based on Hayden et al. (2006, 2013)^19,20^. Criteria were largely similar to Ronald et al. (2019), with one addition in the control of confounders domain, where we stated that the polygenic score p-value threshold should be reported. Detailed descriptions of the quality assessment criteria are provided in the Supplementary Methods. All criteria were rated by reviewers MdW and MM. Since these raters had insufficient specialism in brain-related measures, these specific outcomes were reviewed by an expert (IL). In case of any inconsistencies, criteria were assessed by a third rater (TP). A quality assessment domain was considered to contain a bias if > 50% of the criteria in that domain were scored negatively.

### Systematic review and meta-analyses

#### Data extraction

Results were extracted by authors MdW, and for brain measures by IL. Effect sizes that were not provided in the included papers were requested from corresponding authors via email. If authors did not respond to our request but effect sizes were displayed in Figures, we extracted the results using an online tool (https://automeris.io/WebPlotDigitizer/). All reported effect sizes were transformed to correlation coefficients.

If studies reported results for polygenic scores based on multiple GWAS p-value thresholds, we selected the threshold with the strongest association. Where studies reported multiple models, we selected the effect sizes from models that included covariates that most closely resembled those in other included papers and the requirements for the quality assessment (sex, age, socio-economic status (SES)).

#### Primary analyses

First, all included studies were systematically reviewed with the significance of individual effect sizes based on how they were reported in the respective studies.

Second, for the meta-analysis, all the extracted effect sizes were transformed into correlation coefficients. Since several studies assessed multiple effect sizes, and samples overlapped between studies, we applied three-level random effects meta-analyses per outcome category using the R package Metafor^21^. These models account for dependency between effect sizes by incorporating three variance components; sampling variance of extracted effect sizes (level 1), within-sample variance in effect sizes (level 2) and between sample variance (level 3) ^22^. We tested for overall significant heterogeneity, and heterogeneity within and between studies using the *I*^2^ statistic^23^. We assessed potential publication bias using funnel plots, and by performing and Rosenthal’s Fail Safe N test^24^.

#### Secondary analyses

In our systematic review, we additionally describe sex differences. We further performed a series of secondary meta-analyses: 1) differences in the association between the autism polygenic score and autism diagnosis for specific populations (Europe-based versus US-based), and 2) subgroup analyses for the associations between the autism polygenic score and ADHD, psychotic spectrum, eating disorders and self-harm and suicide ideation (in specific psychiatric classifications category).

## Results

### Study characteristics

Studies showed relatively large sample overlap: A total of 57 different samples were used across the 72 studies (pooled N = 720,087). The average percentage of males in the included study samples was 48.3% (^25–31^) omitted from this percentage due to missing or unclear sex information). Of the included studies 47.2% were performed on child or adolescent (<18 years) participants, 19.4% on adult (> 18 years) participants, 29.2% on a mixed sample of children and adults, and for 2,8% the sample age information was unavailable or unclear. Studies were most often based on participants living in Europe (63.8%), followed by Northern America (25.8%), Asia (5.6%), and Australia (1.4%). The remaining studies (6.9%) were based on participants from multiple continents. Genetic ancestry was mostly European (86.1%), with a limited number of papers including East Asian (5.6%) or trans-ancestry sample analyses (9.7%). Results can be considered to be based on participants of European ancestry, unless described otherwise. Individual study characteristics are presented in Table 1.

**Table 1.**
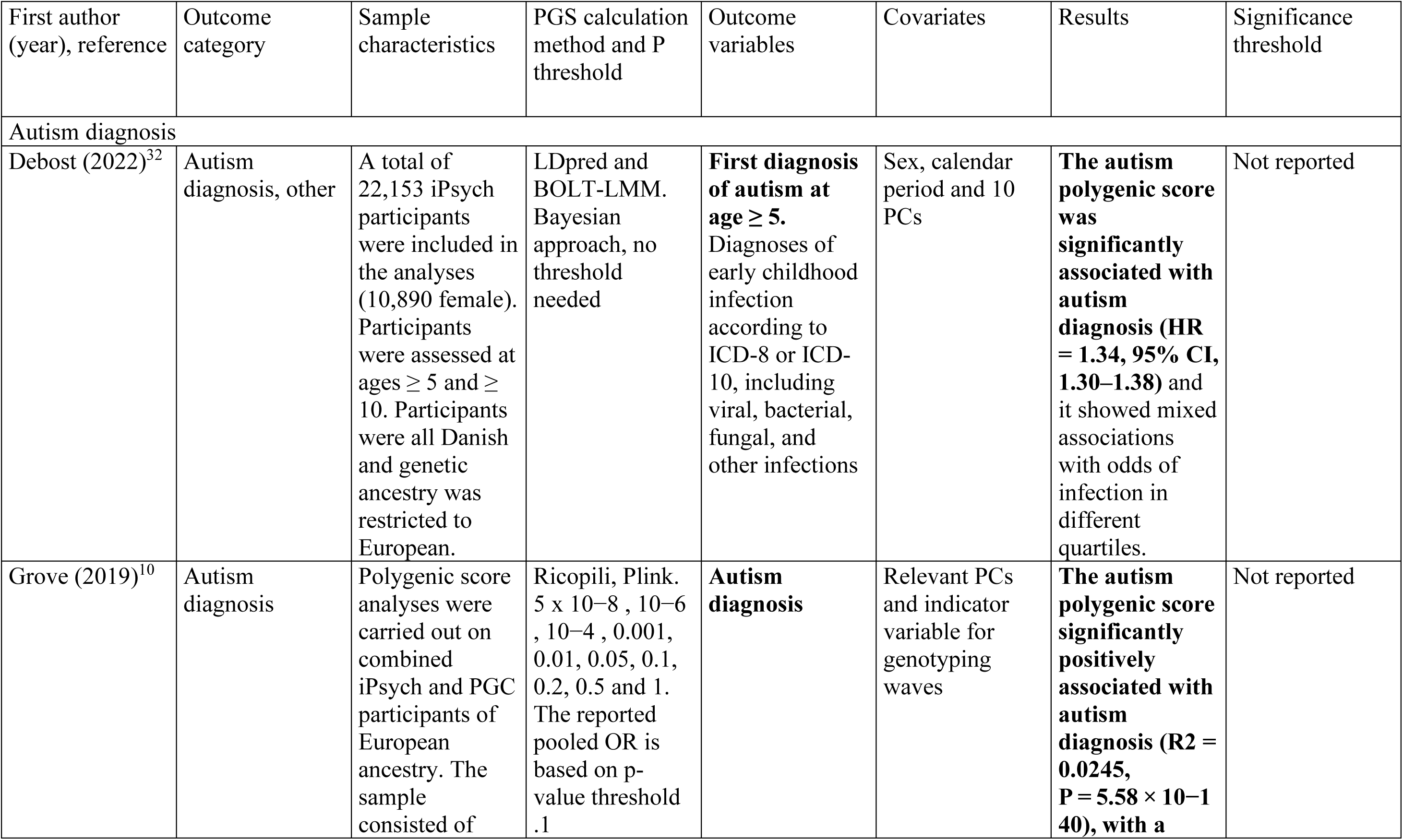
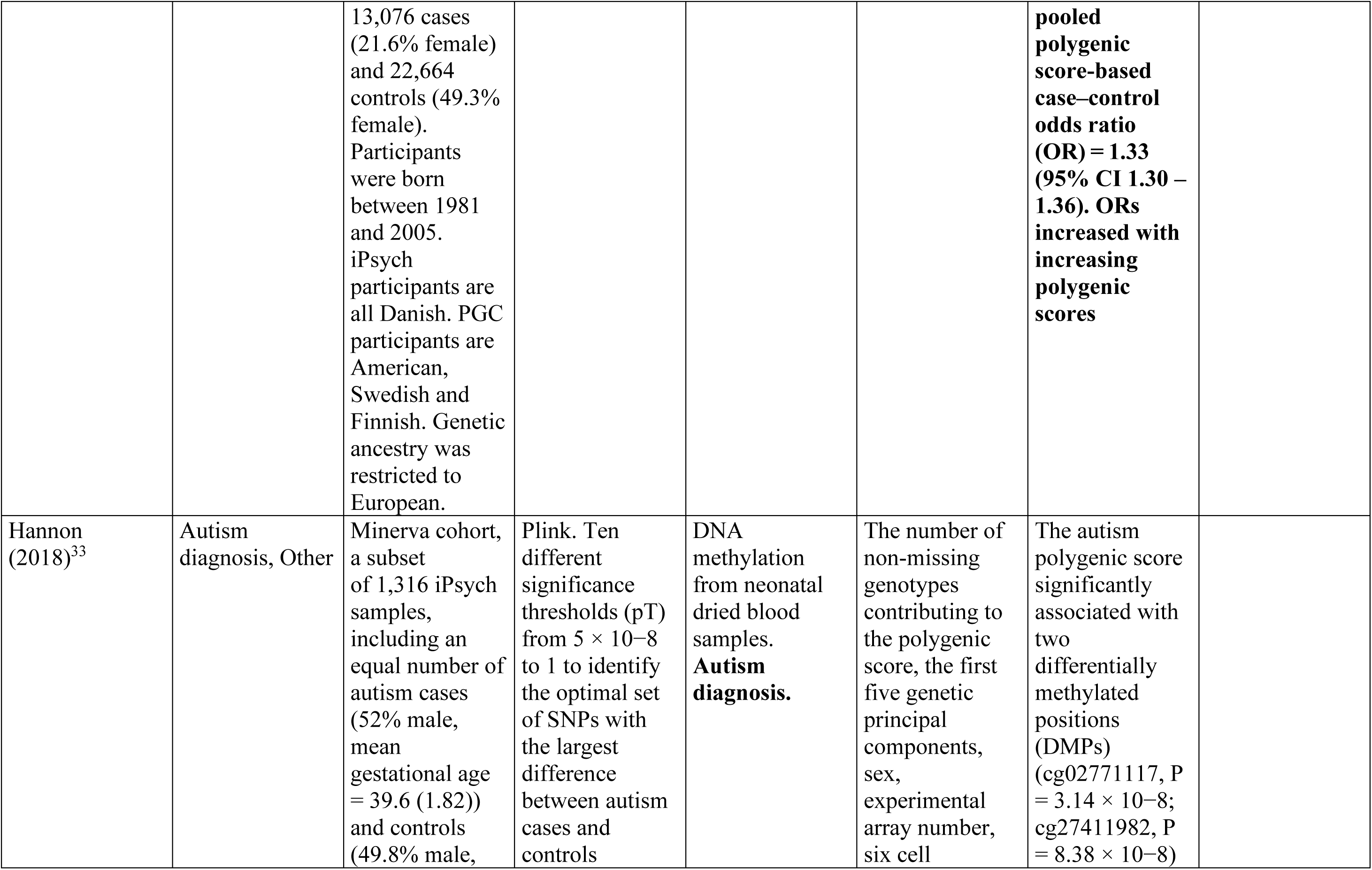
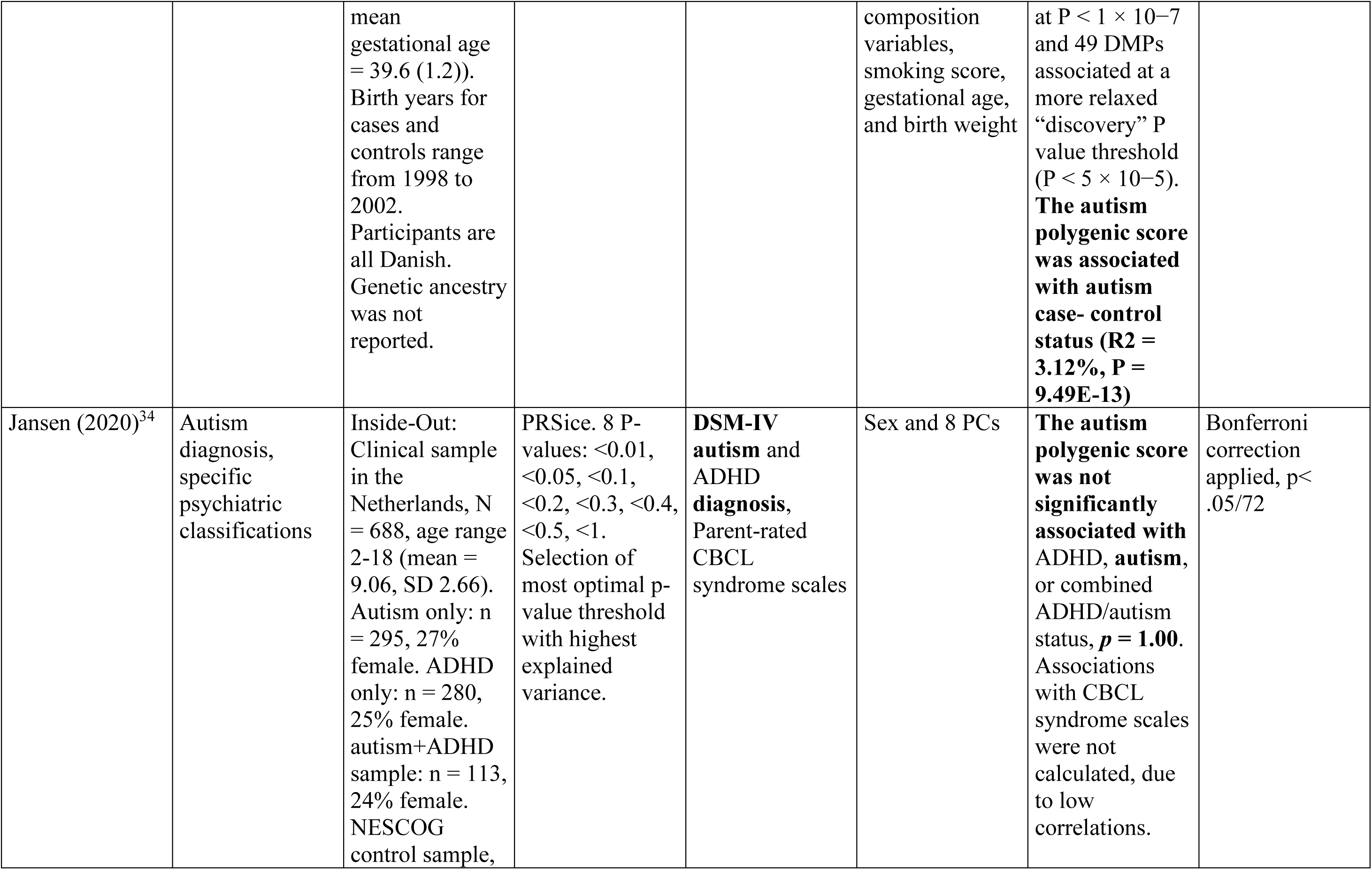
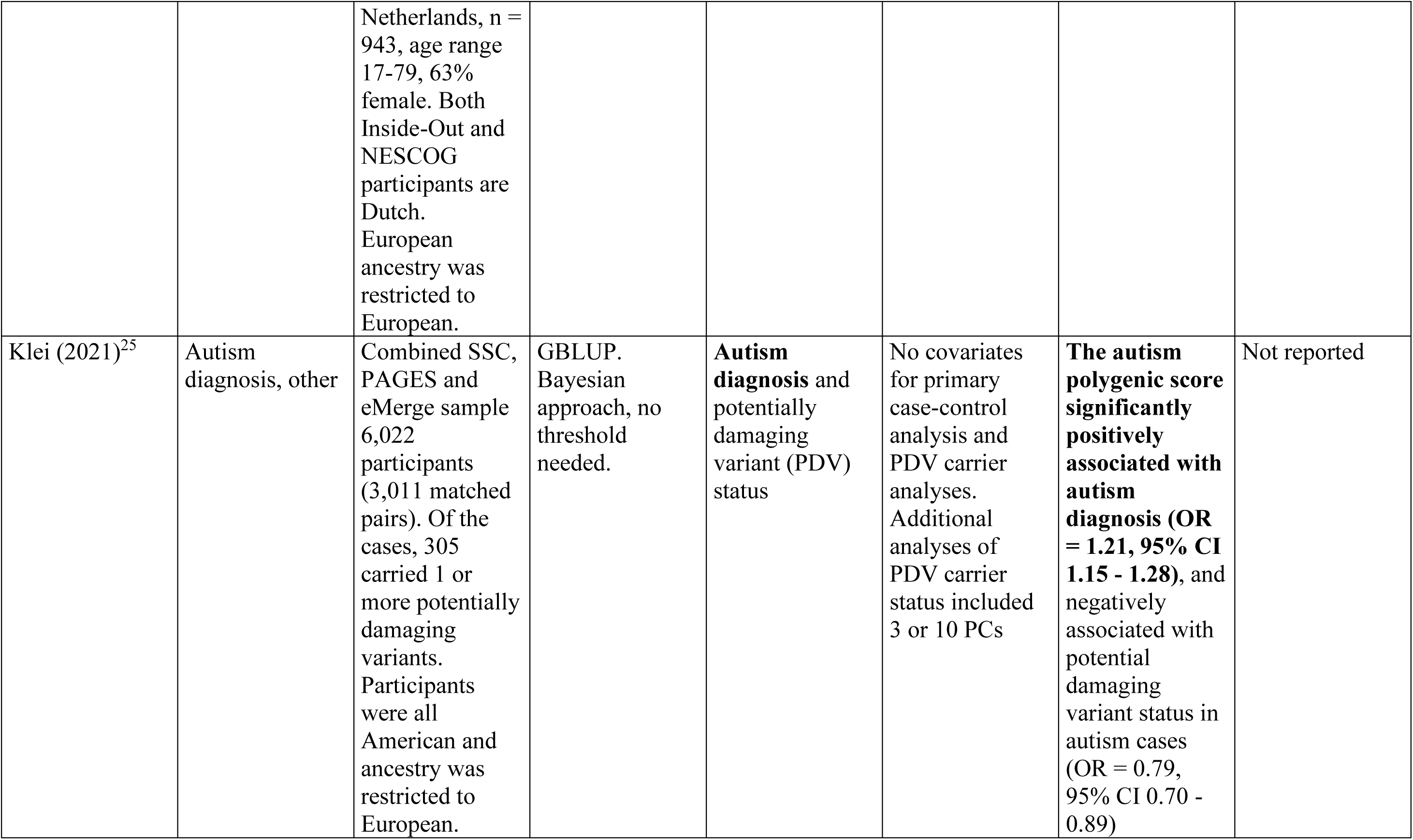
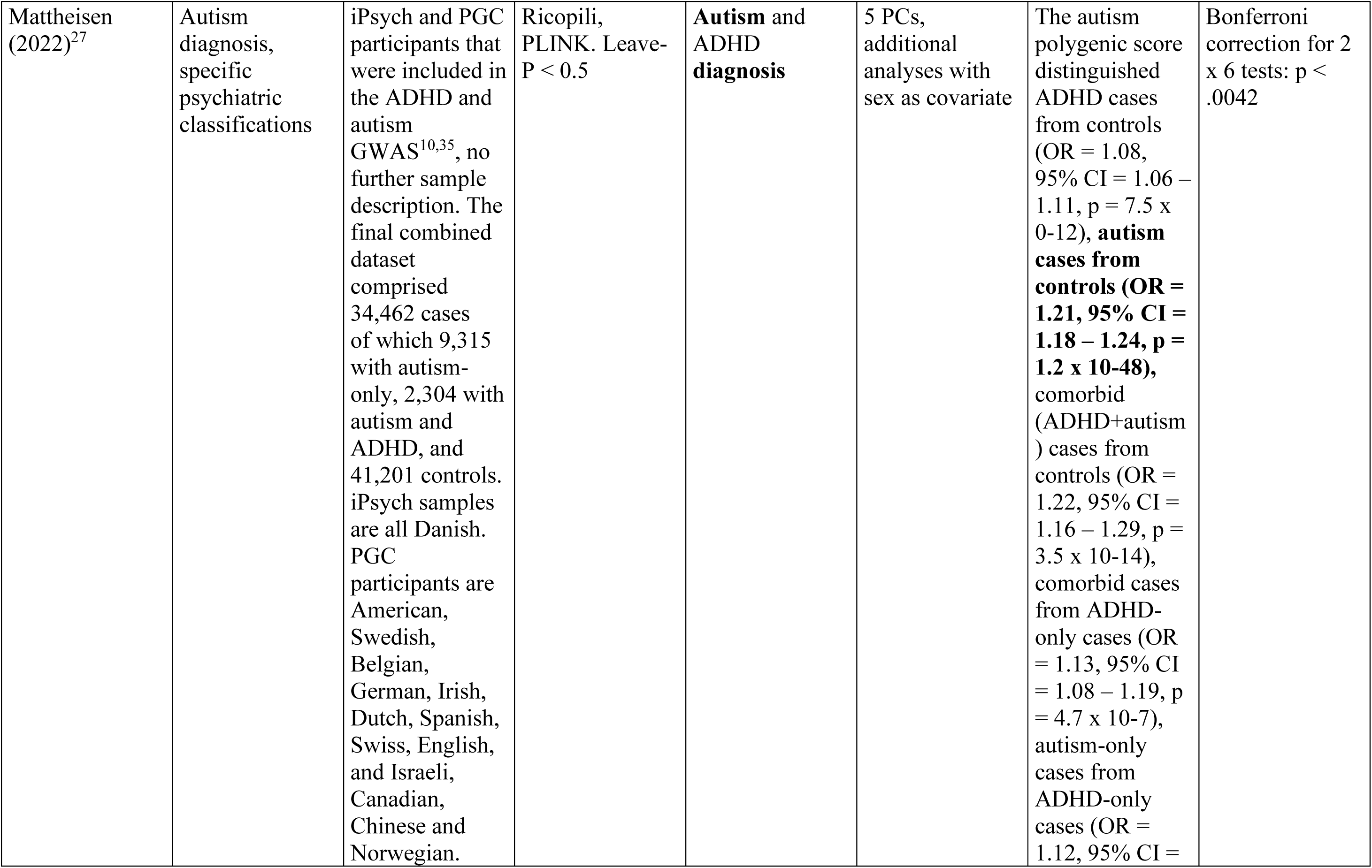
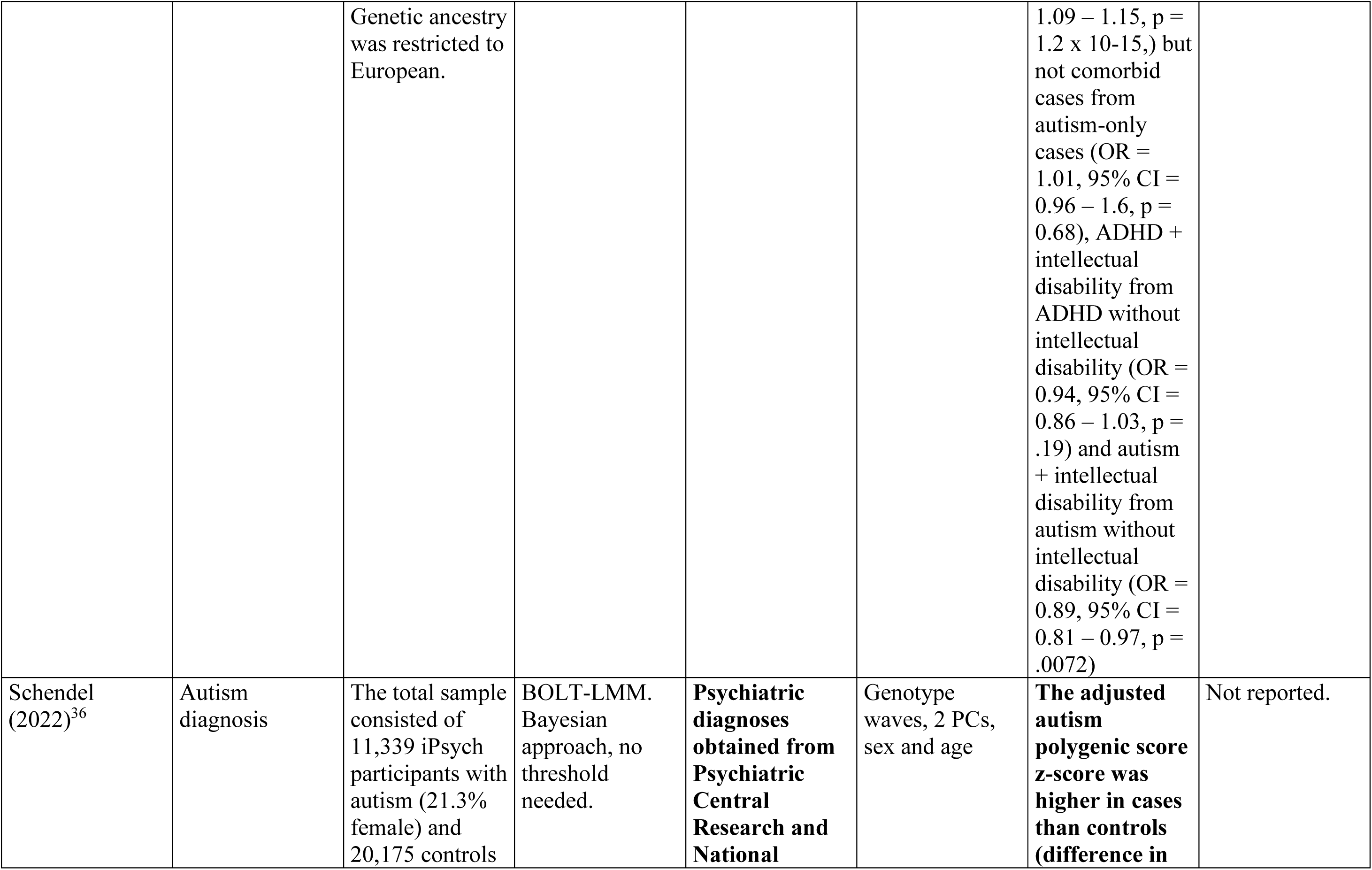
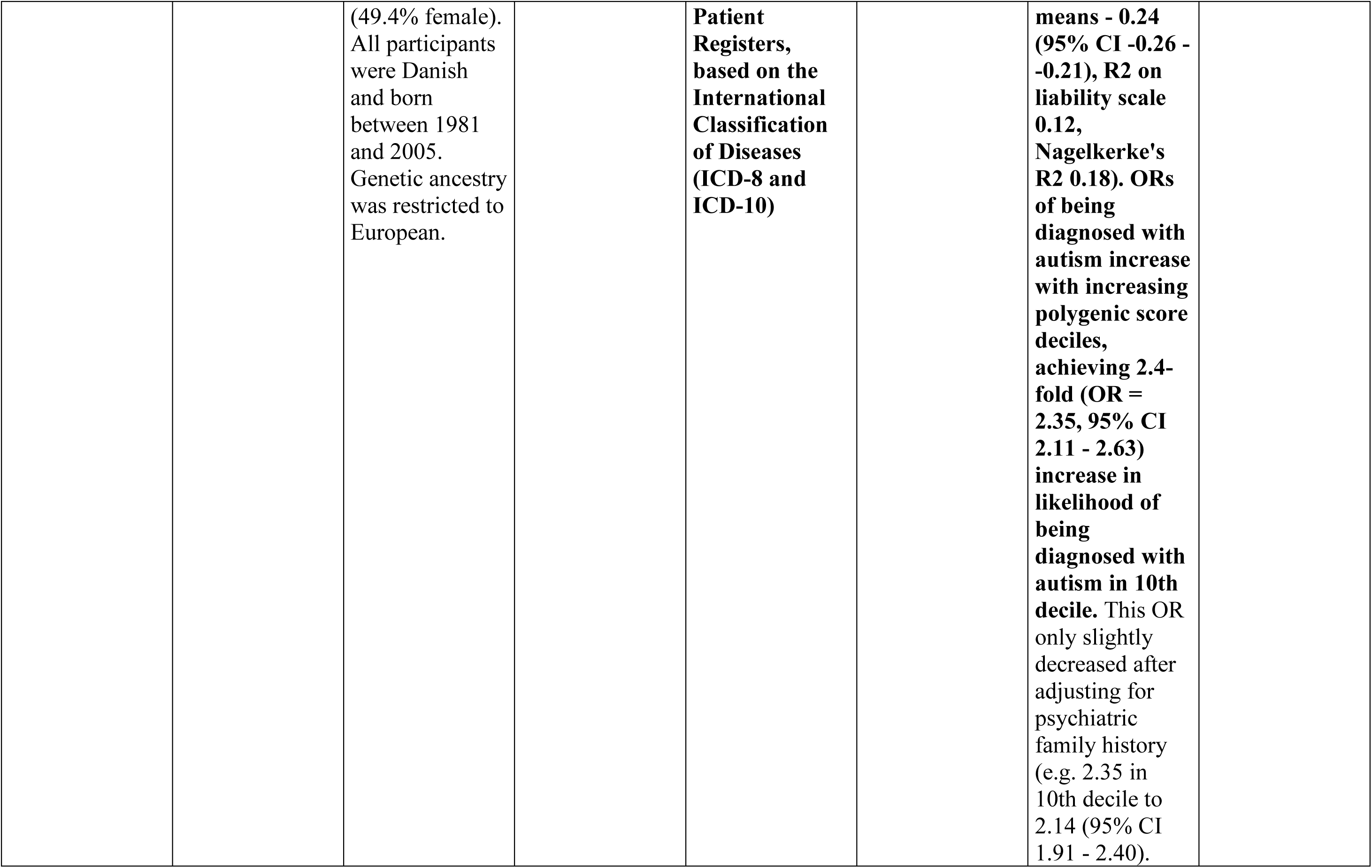
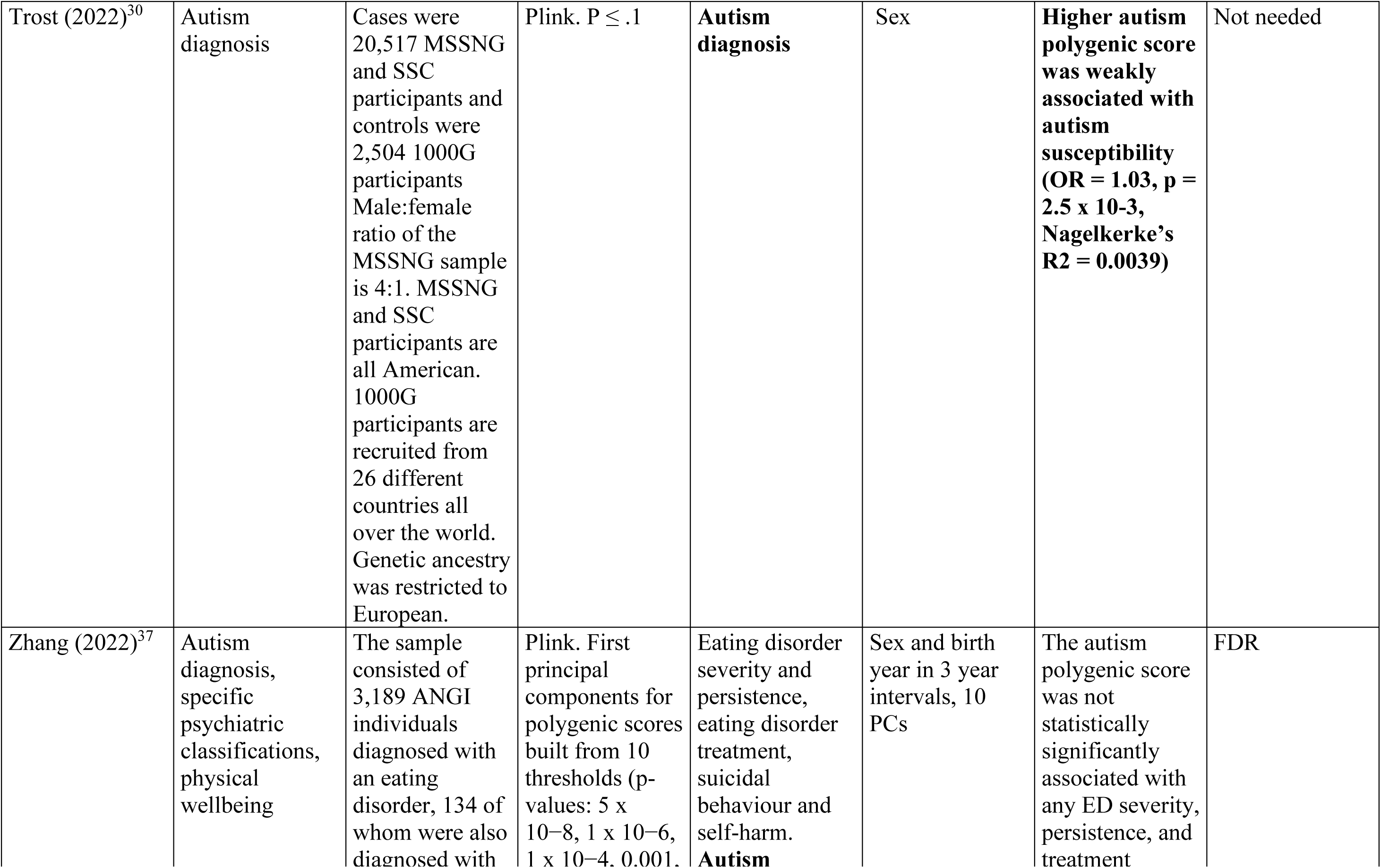
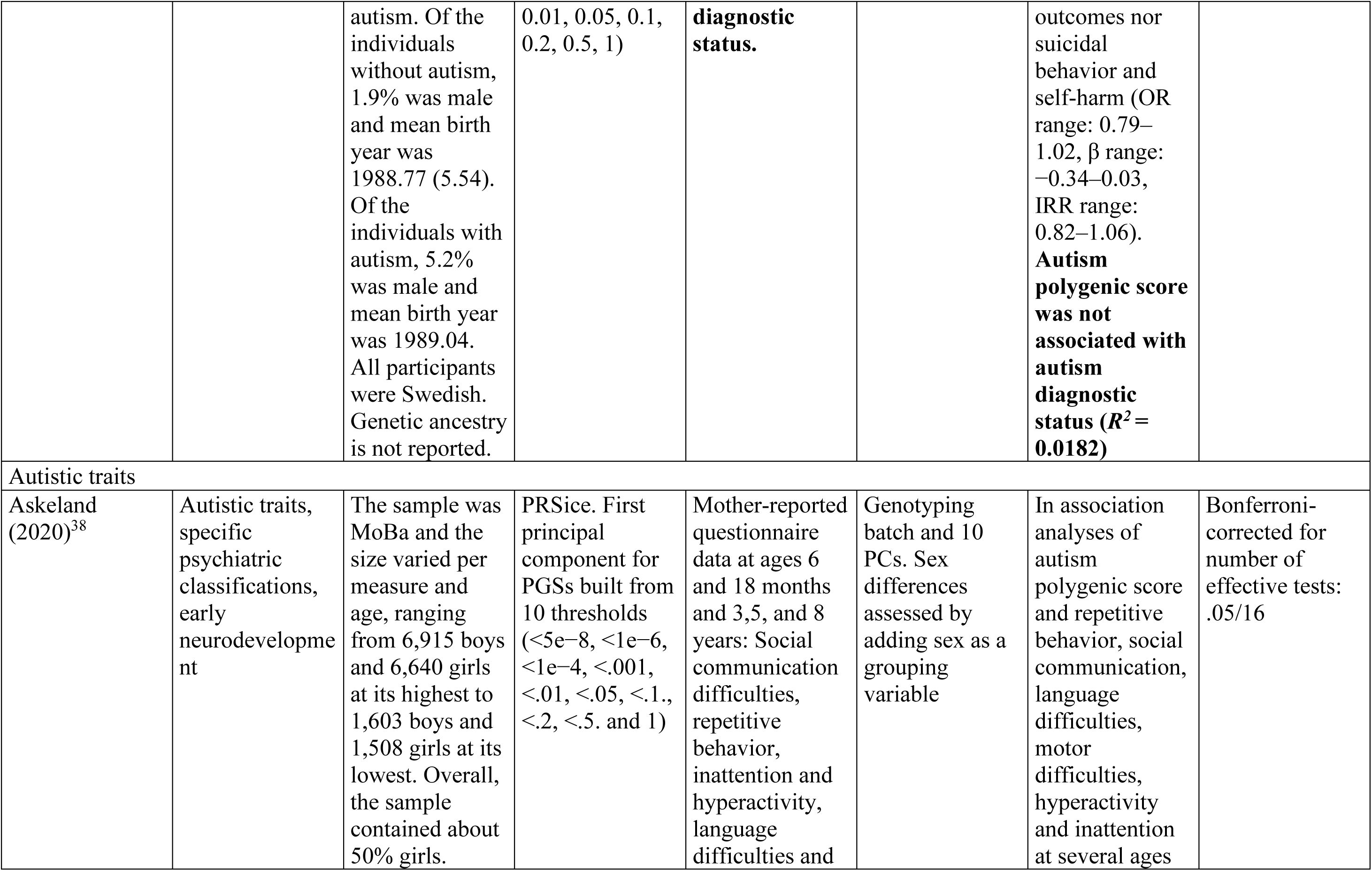
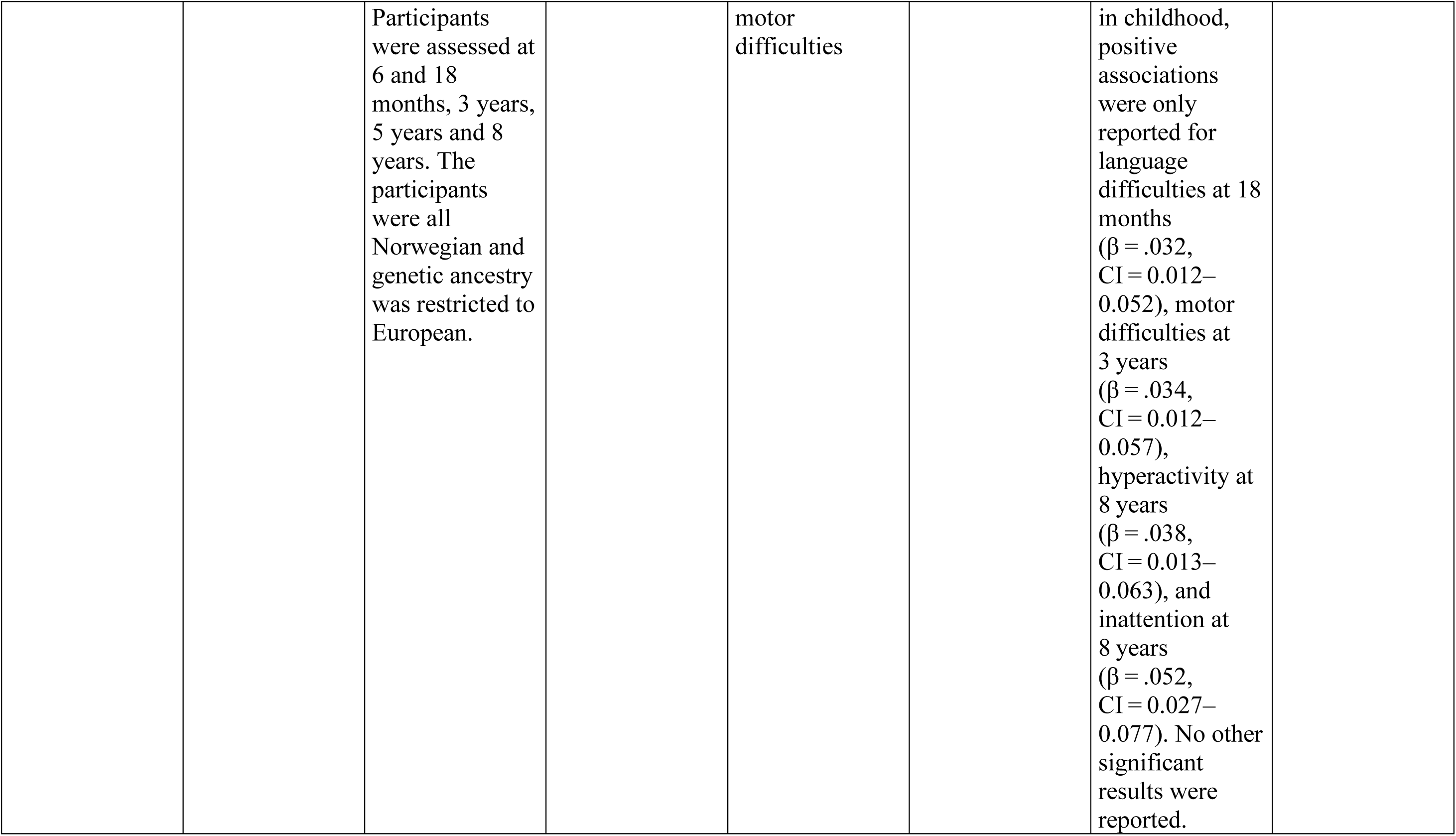
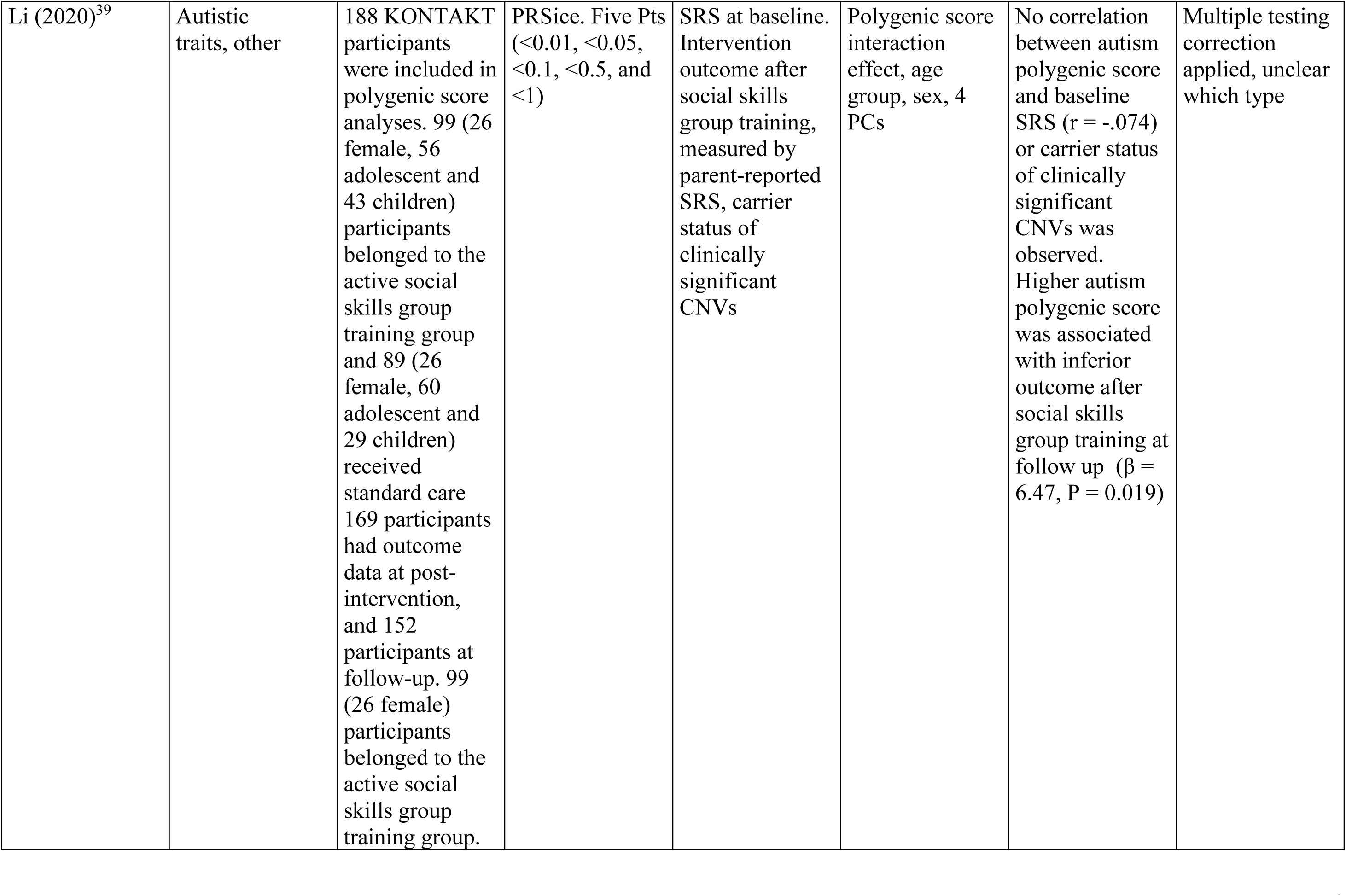
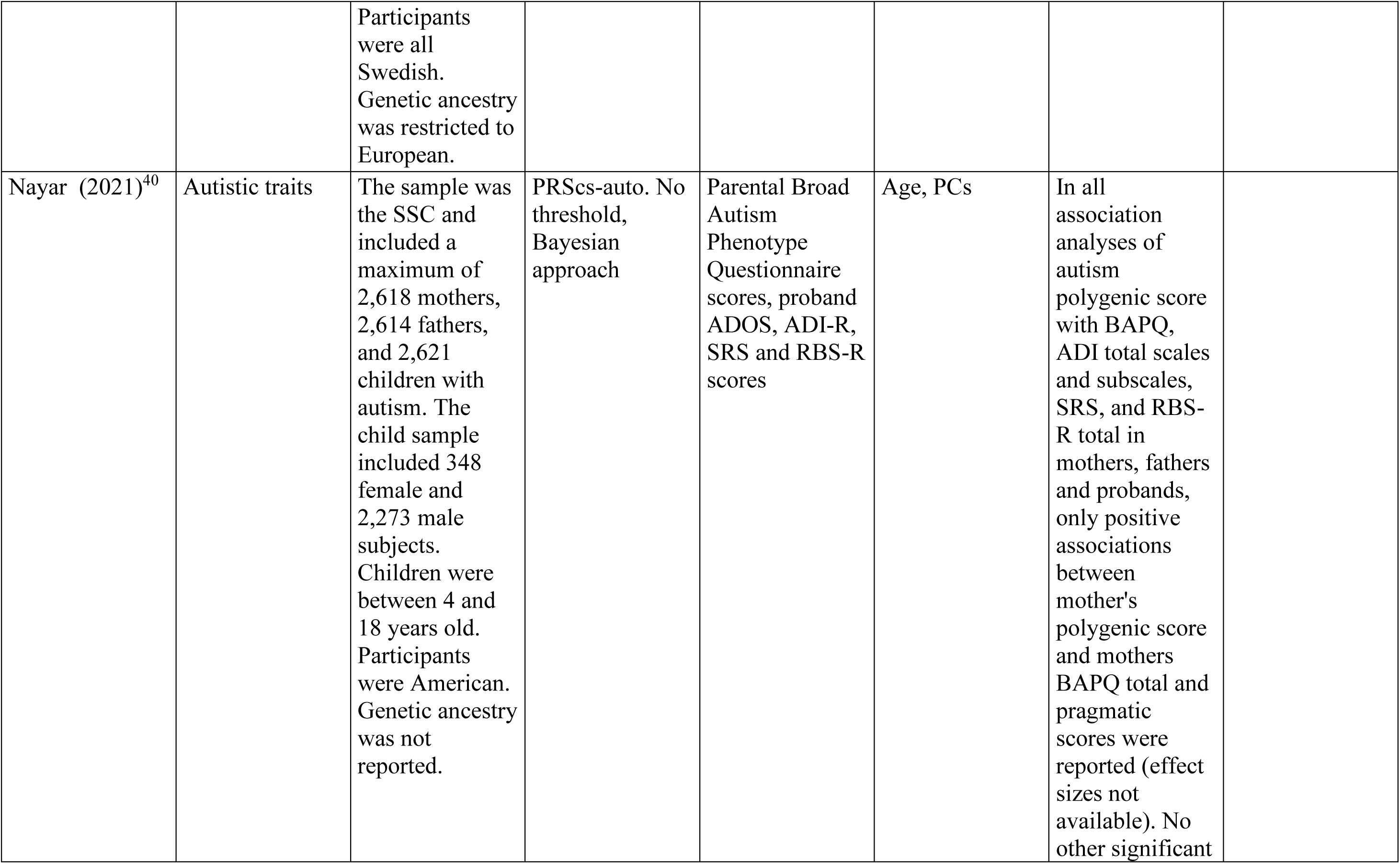
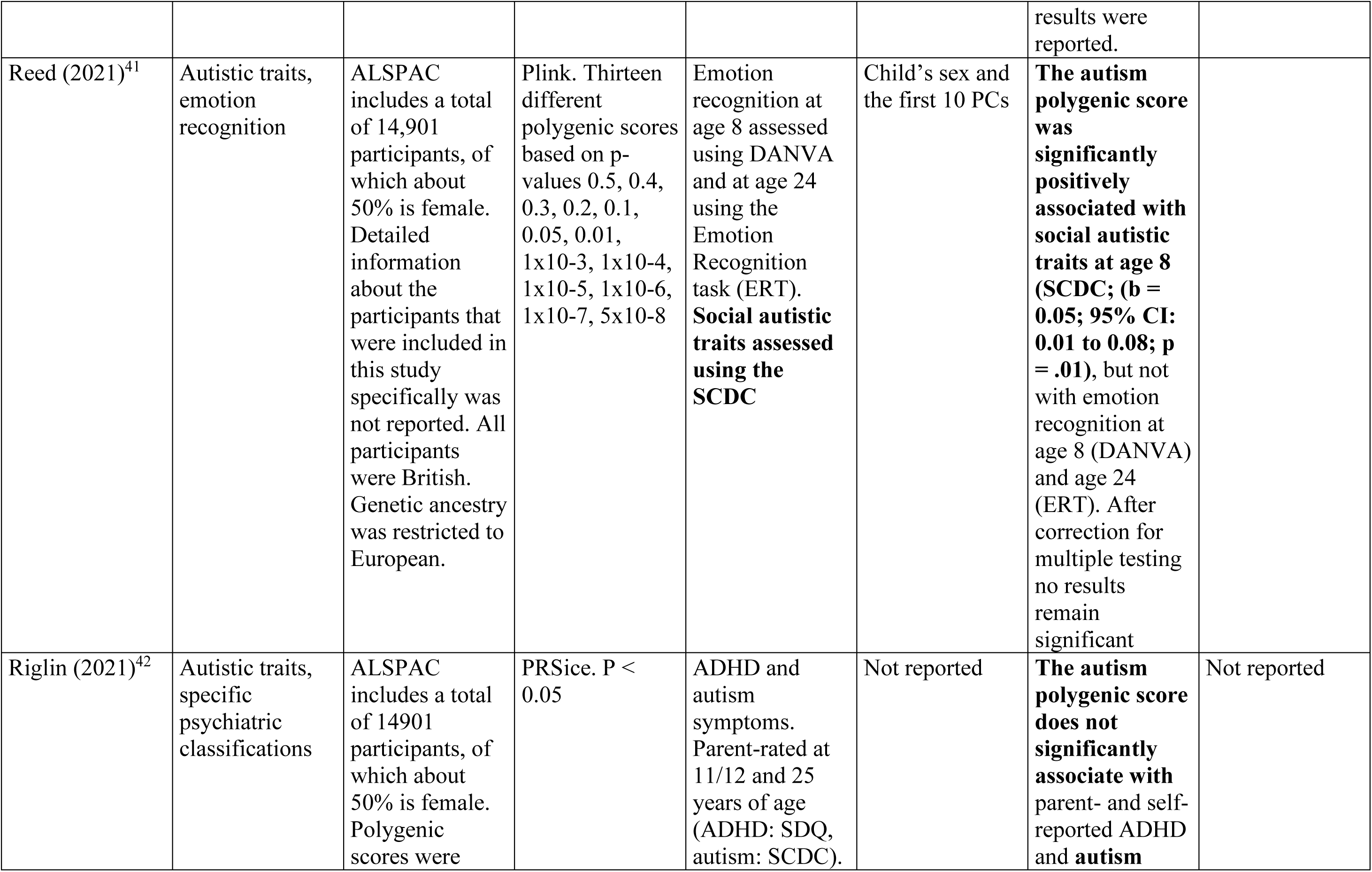
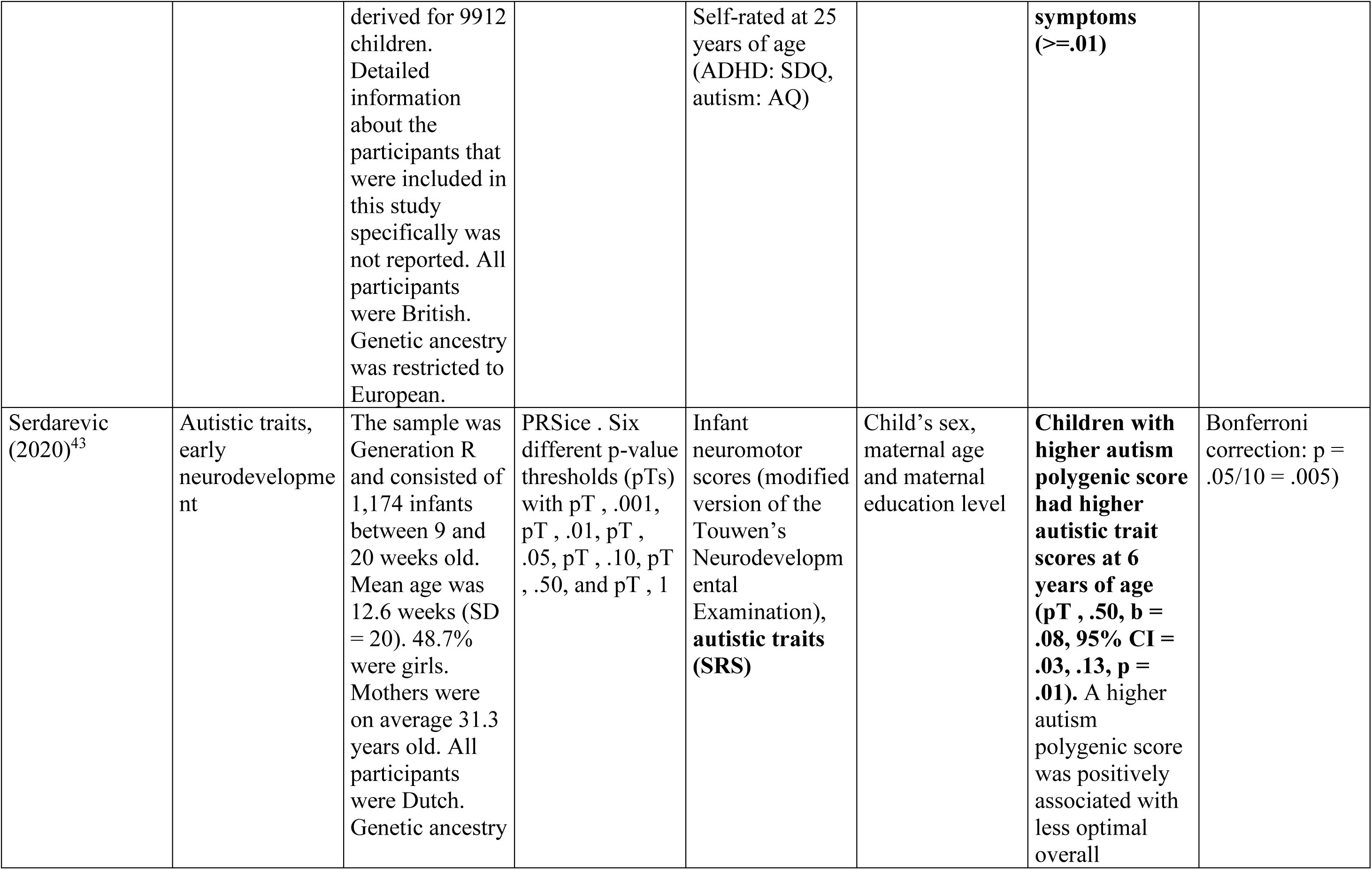
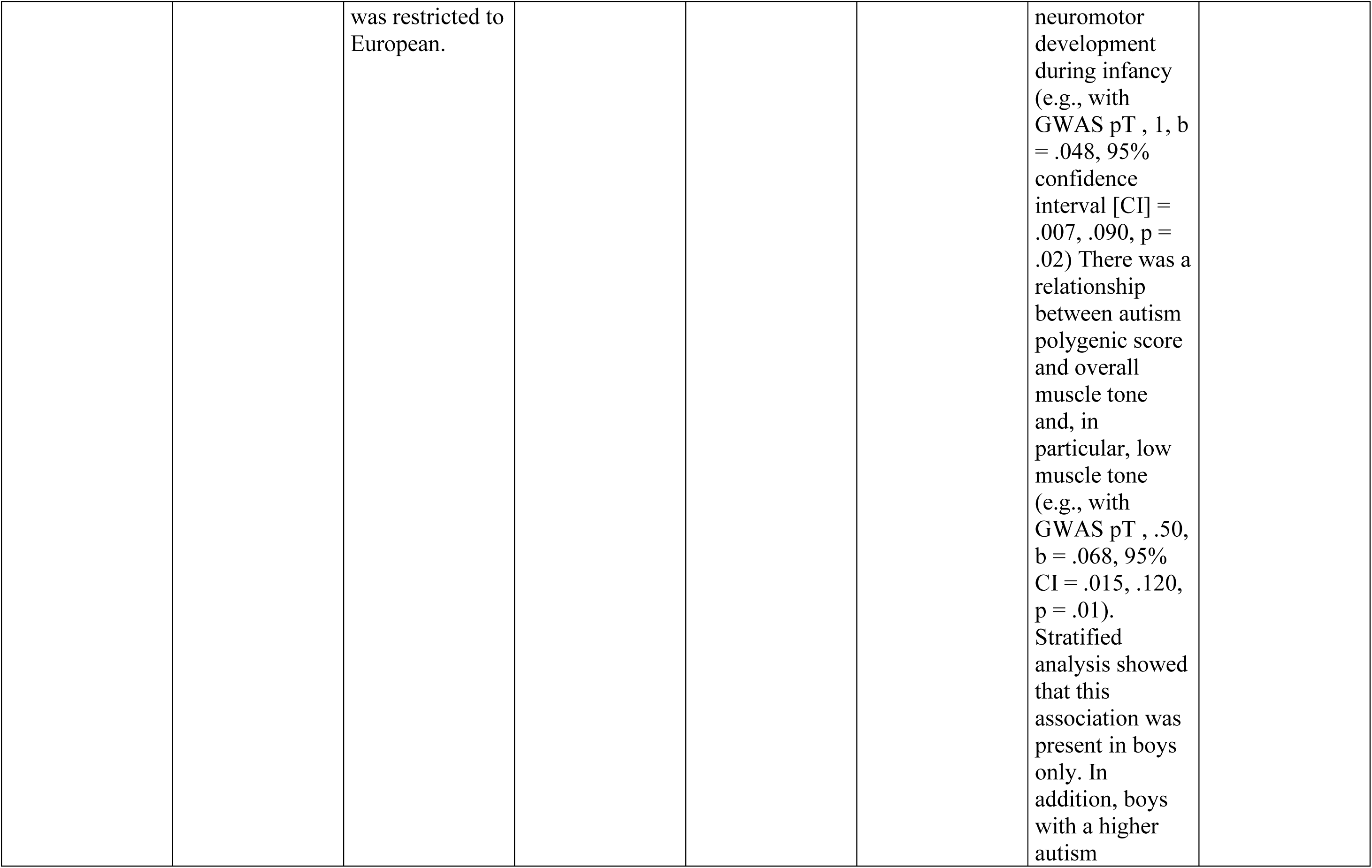
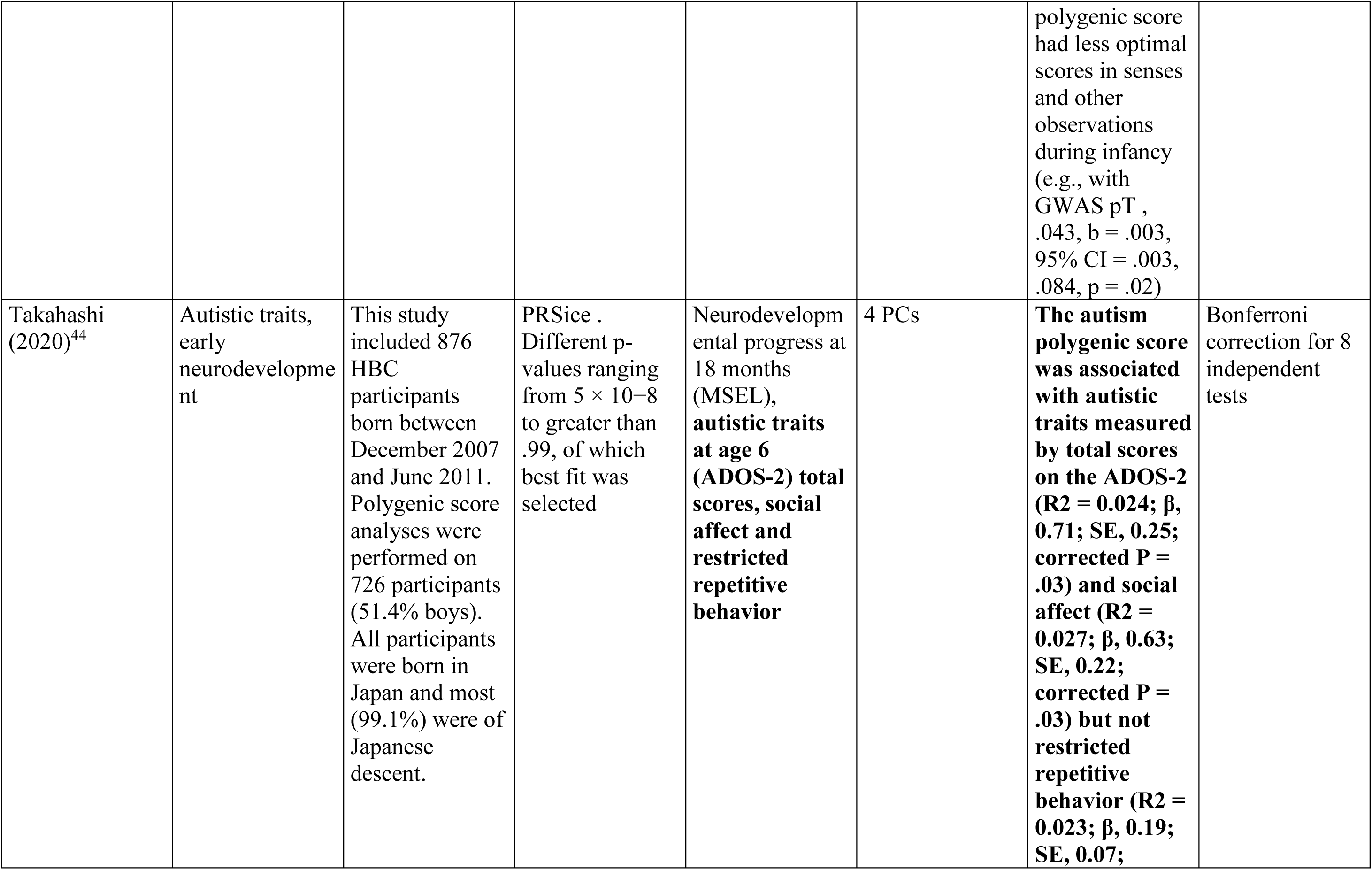
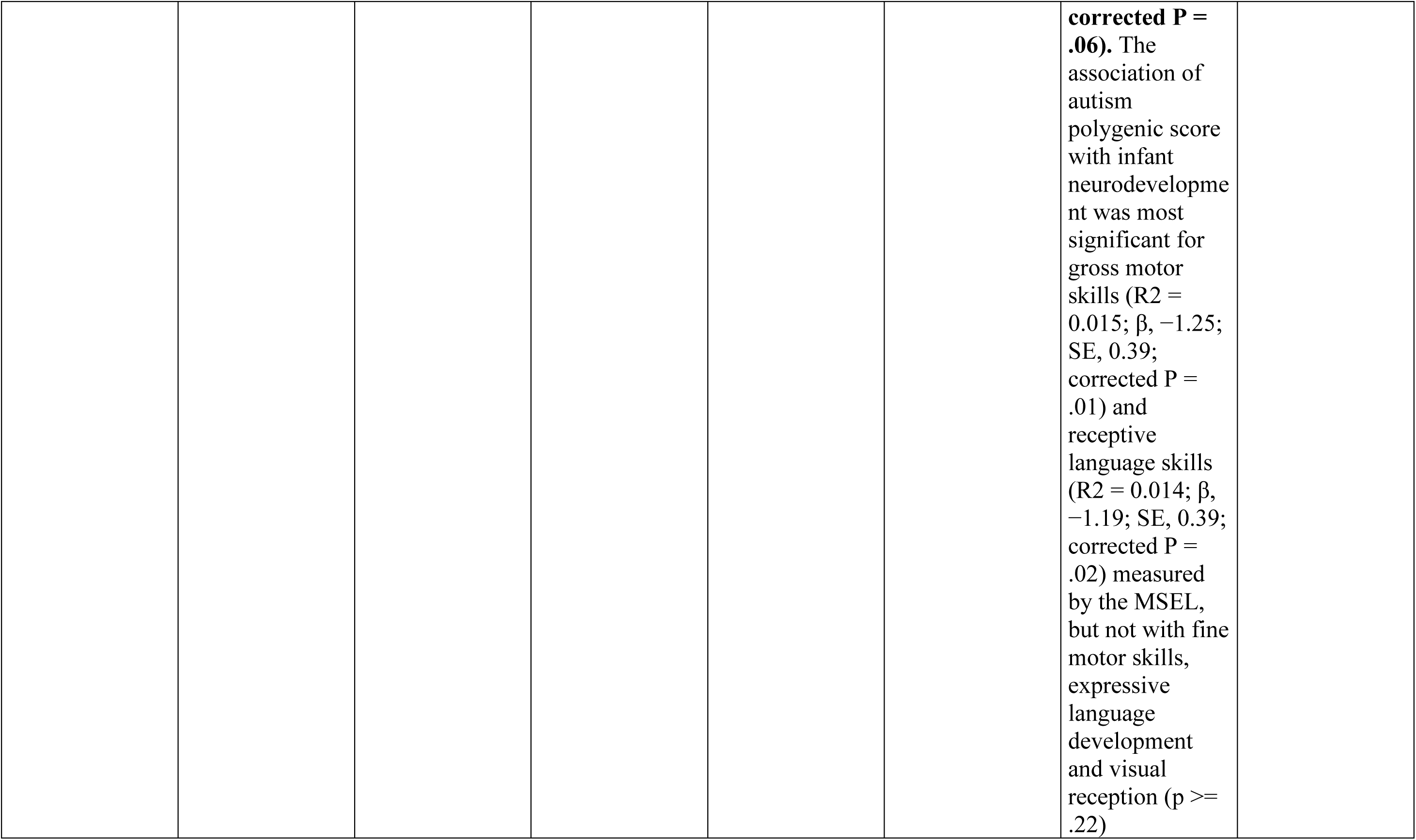
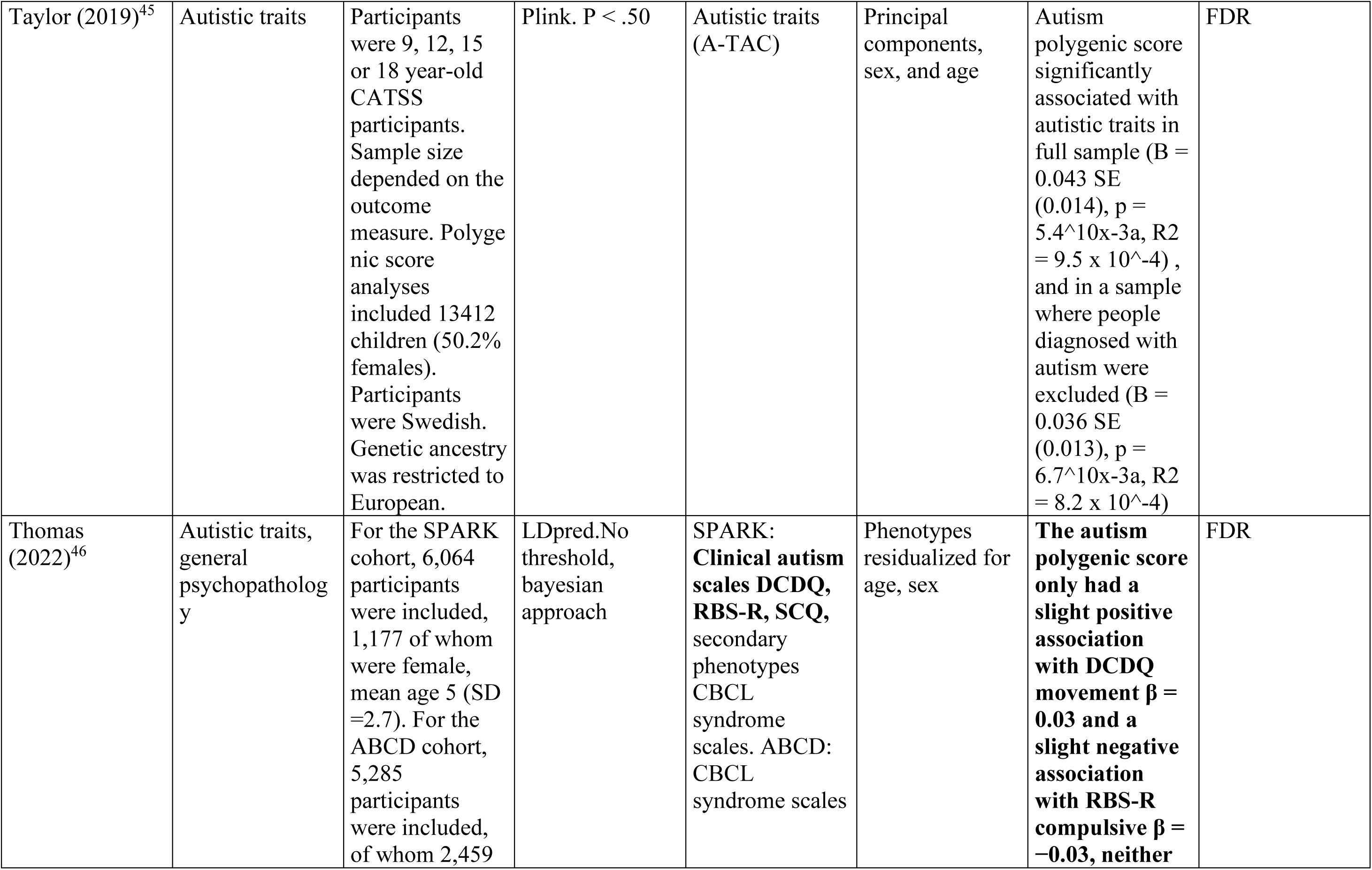
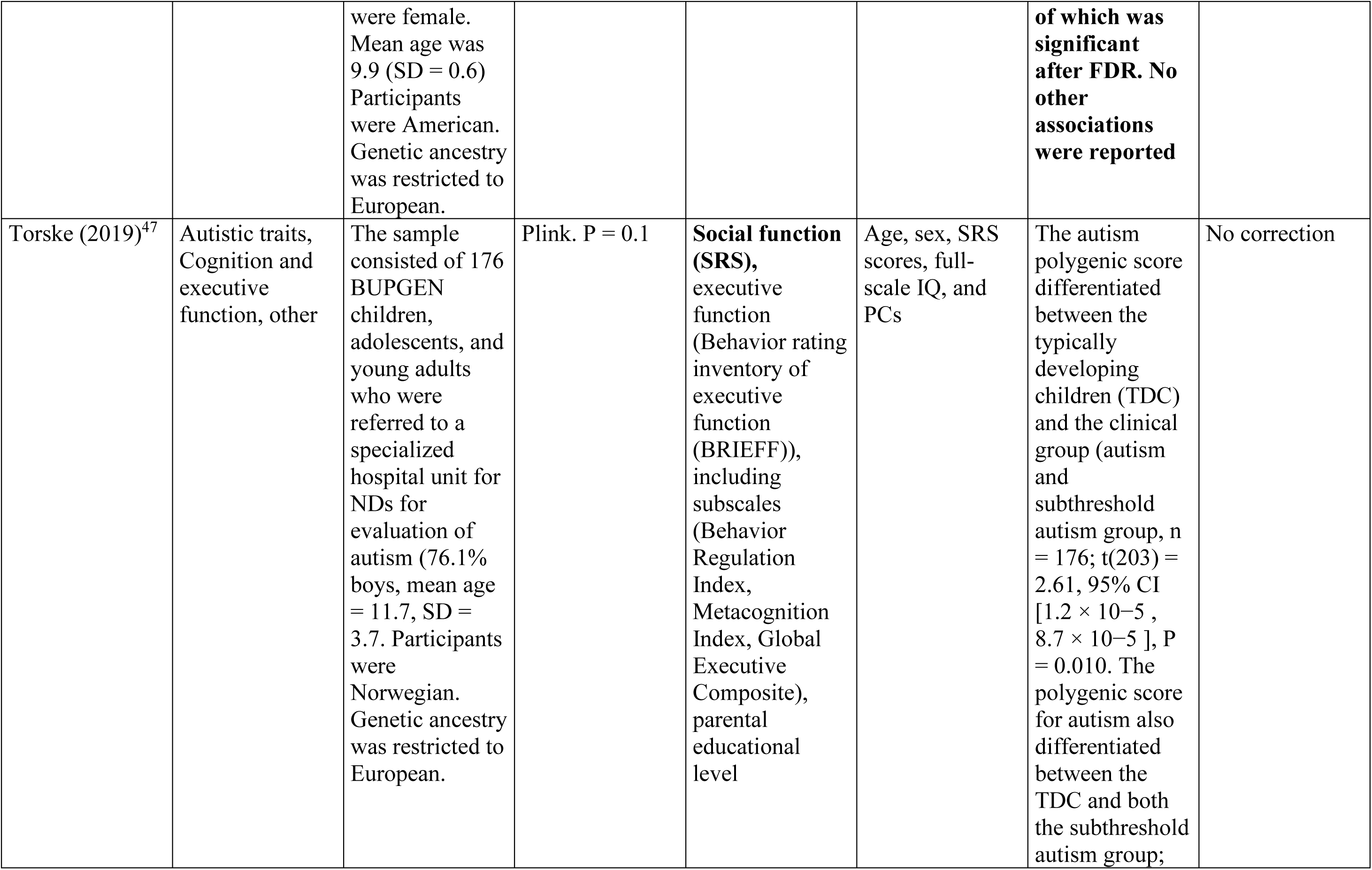
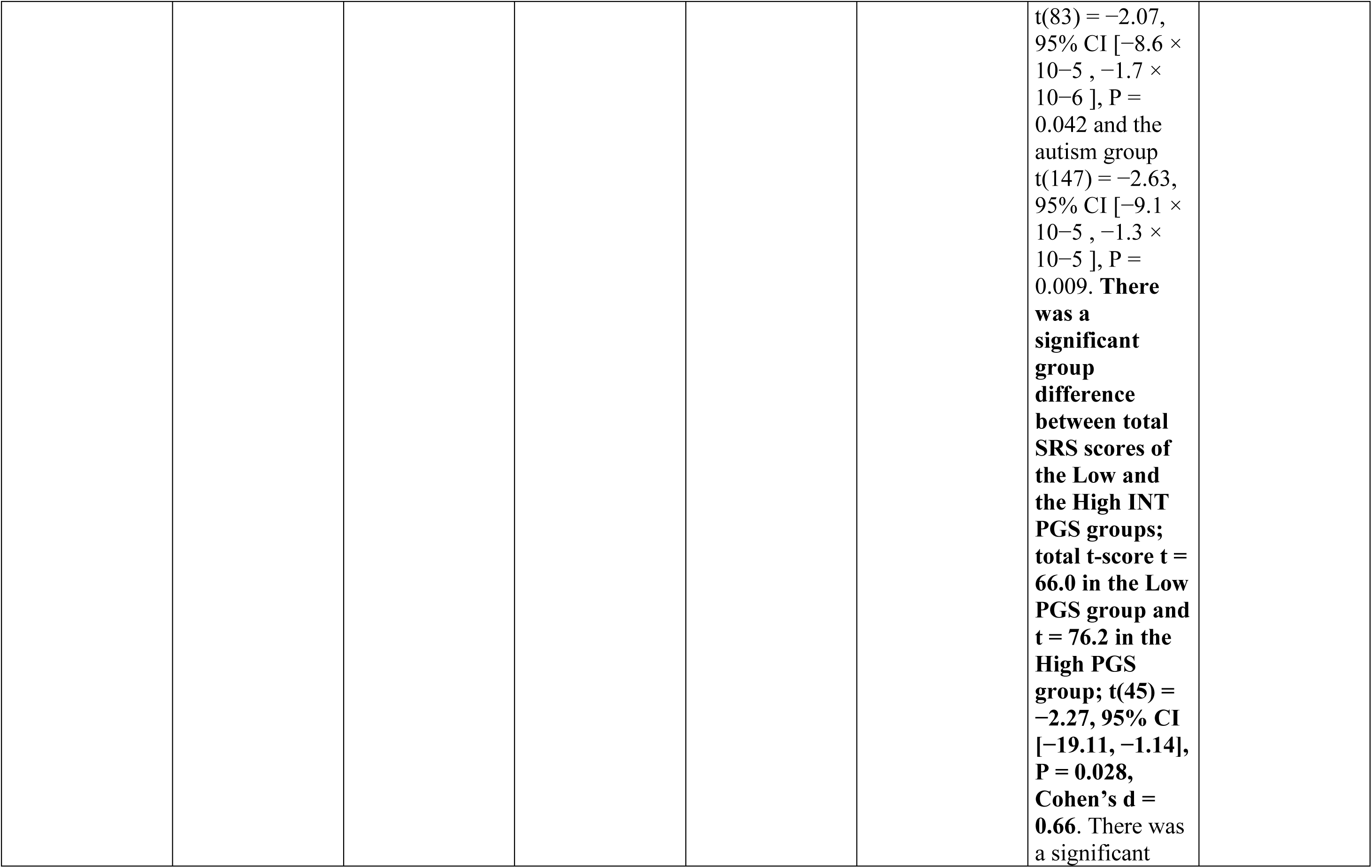
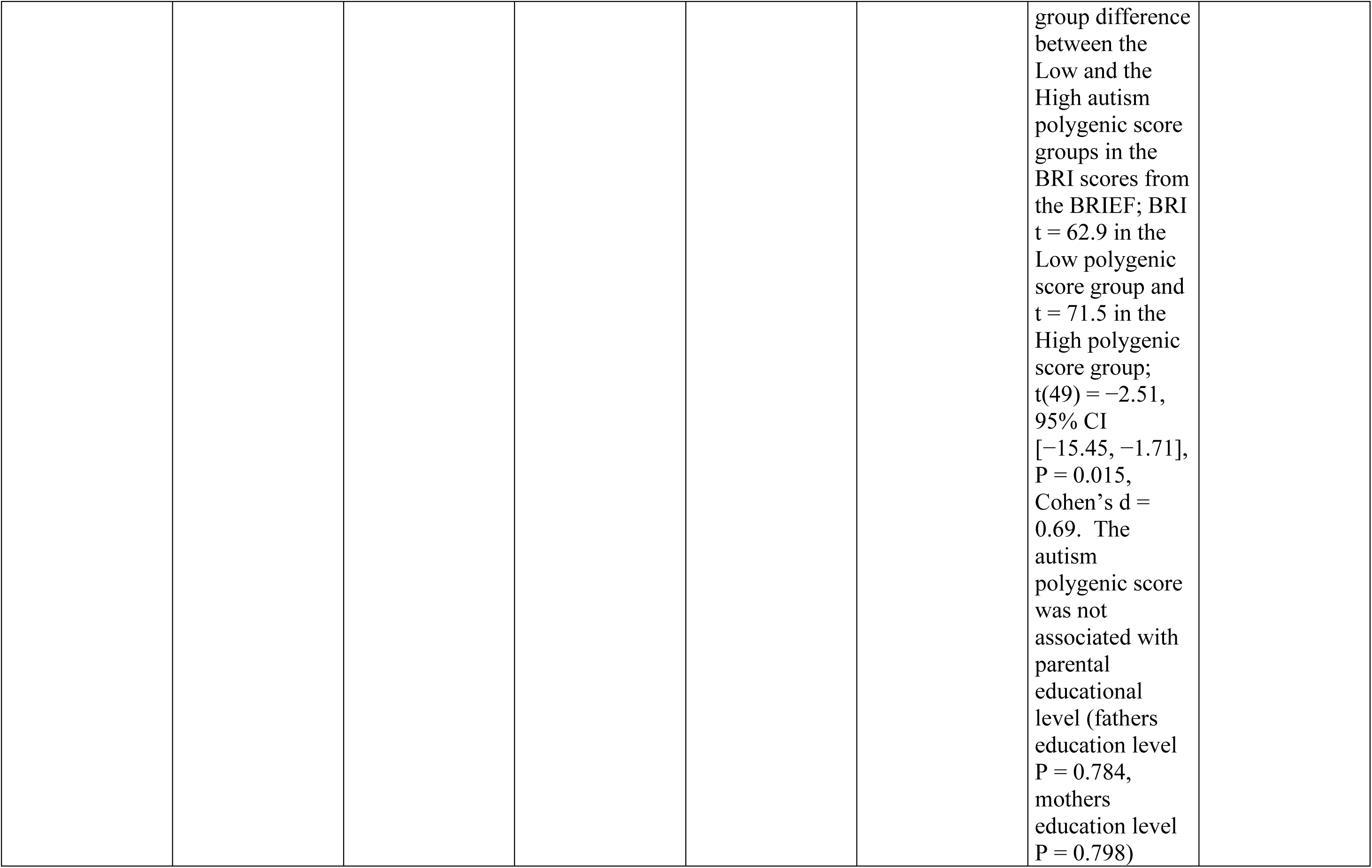
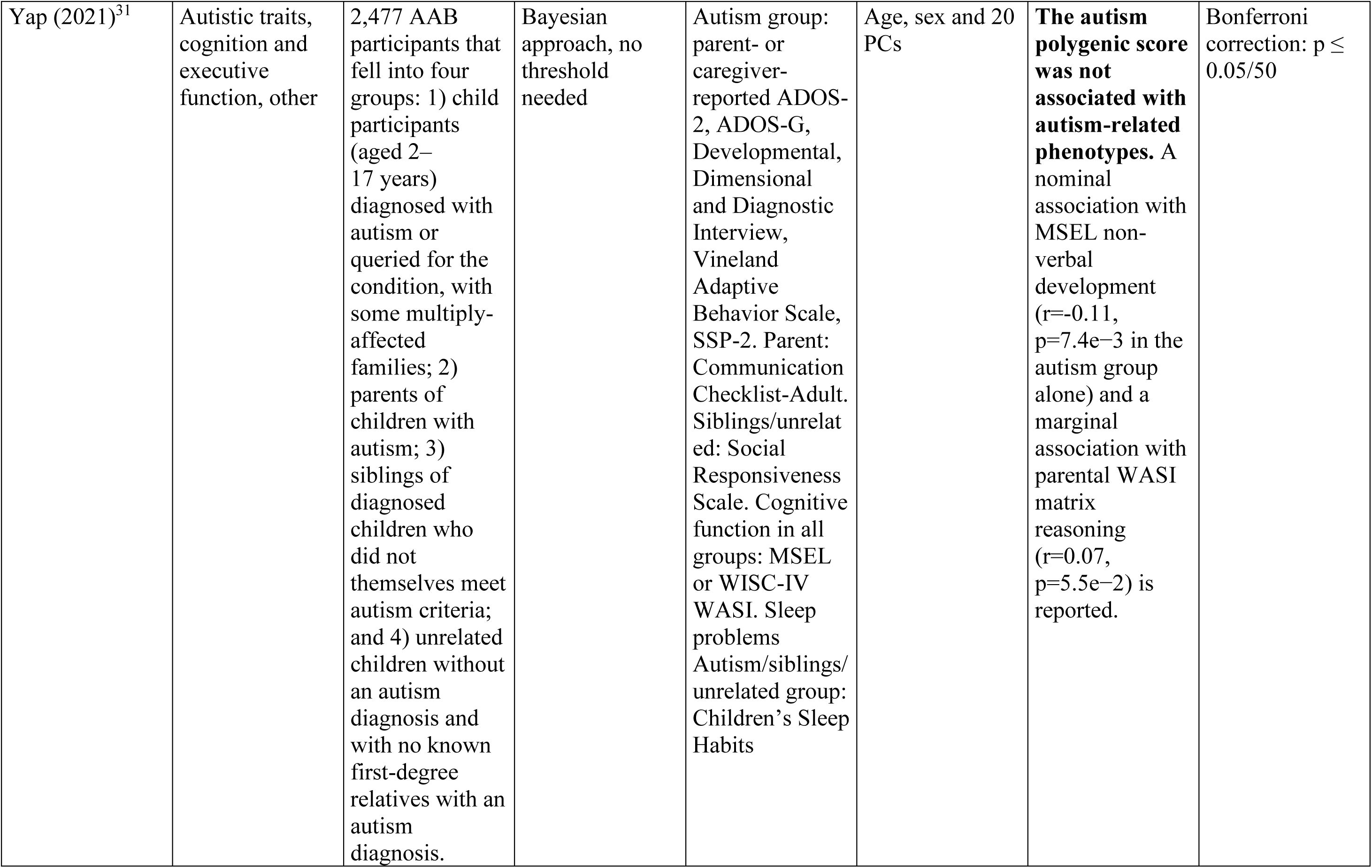
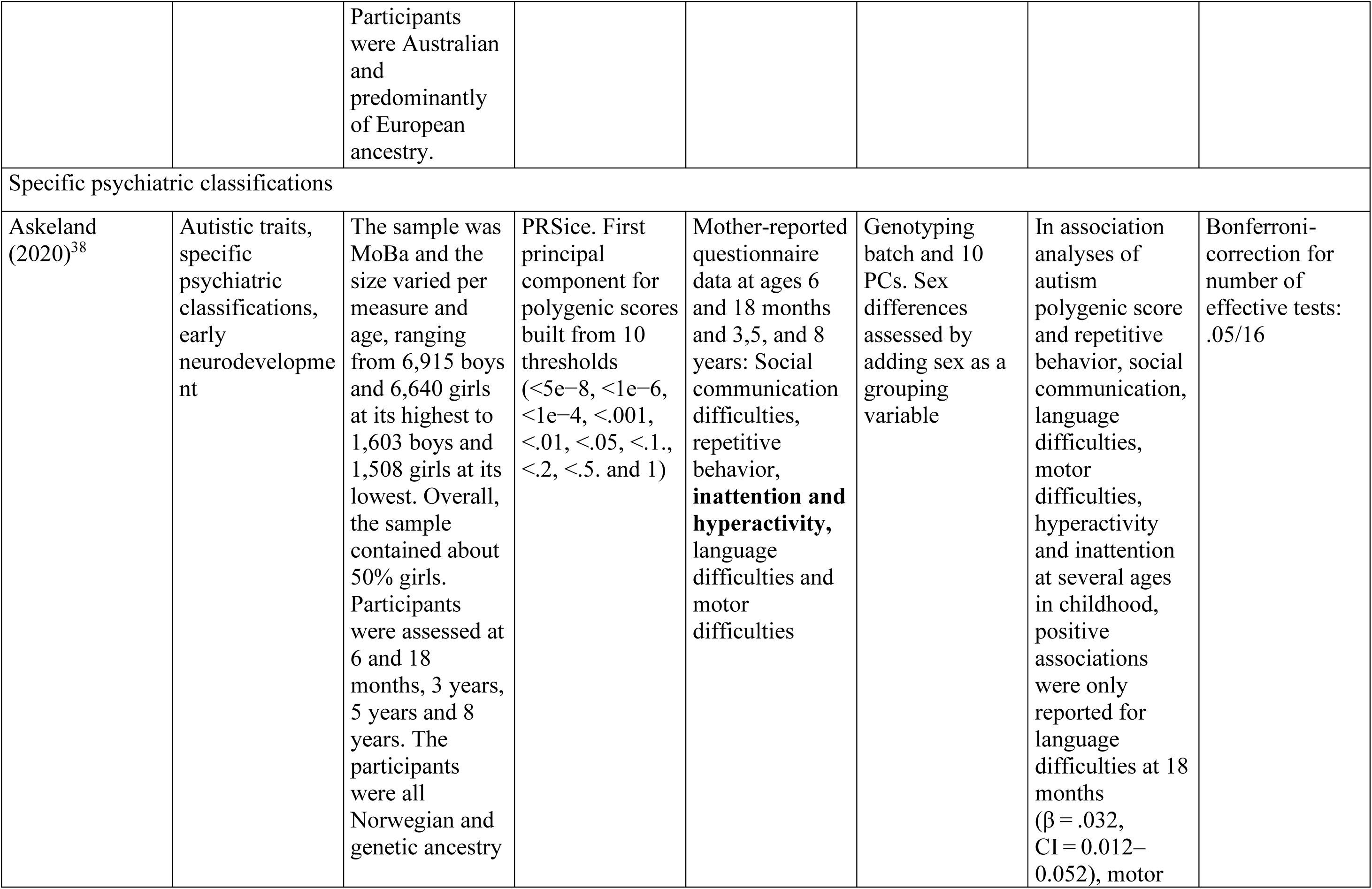
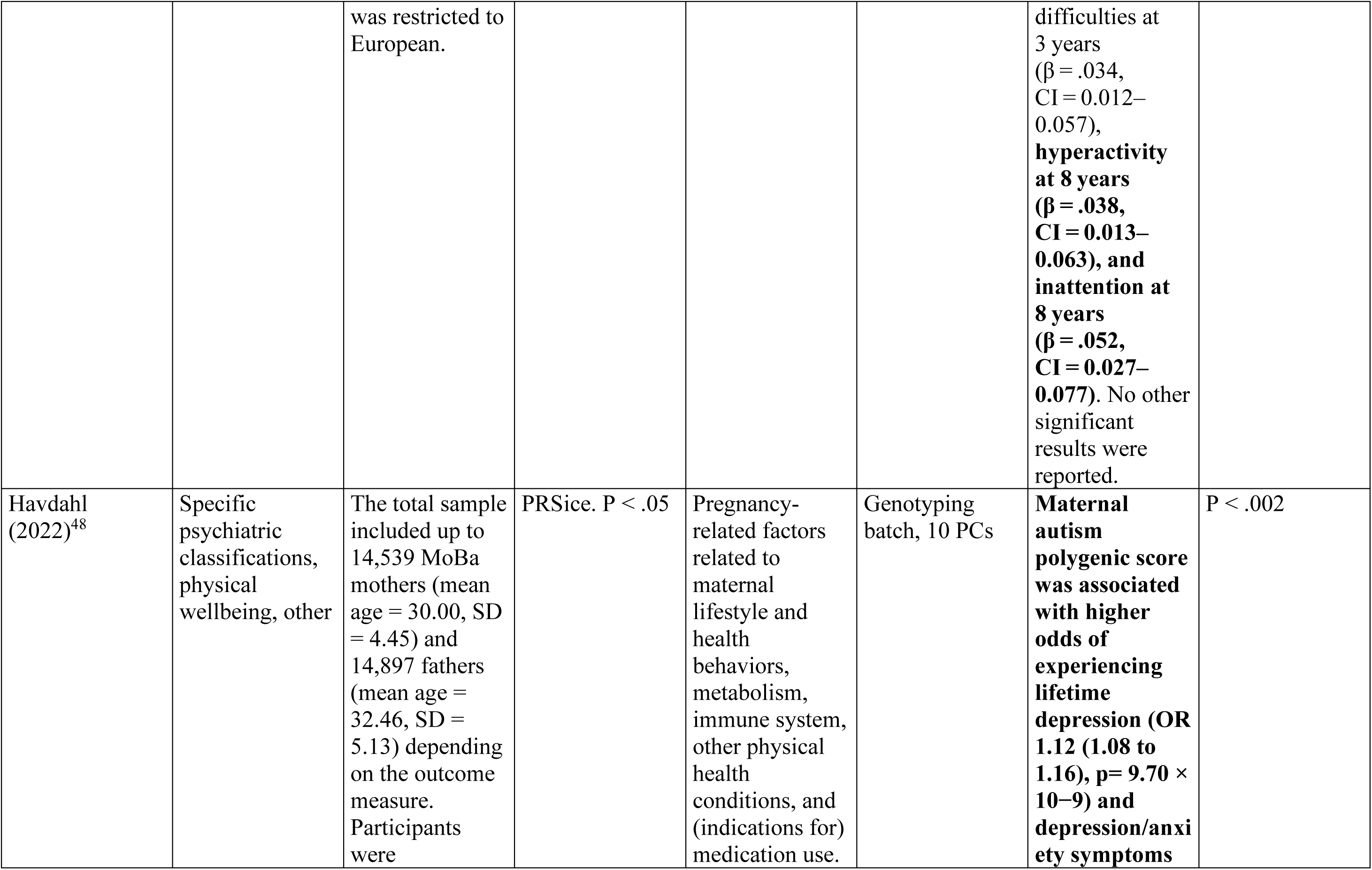
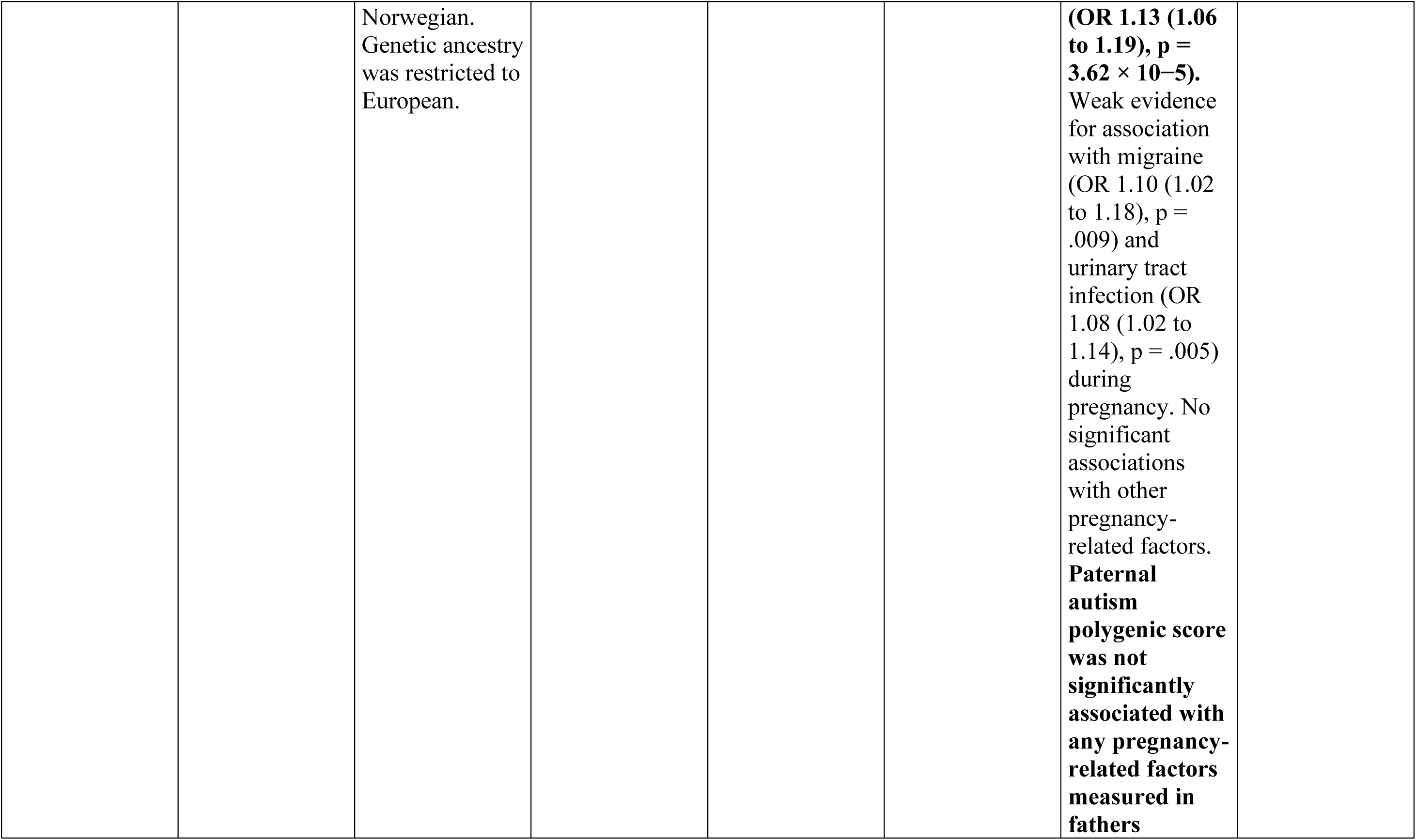
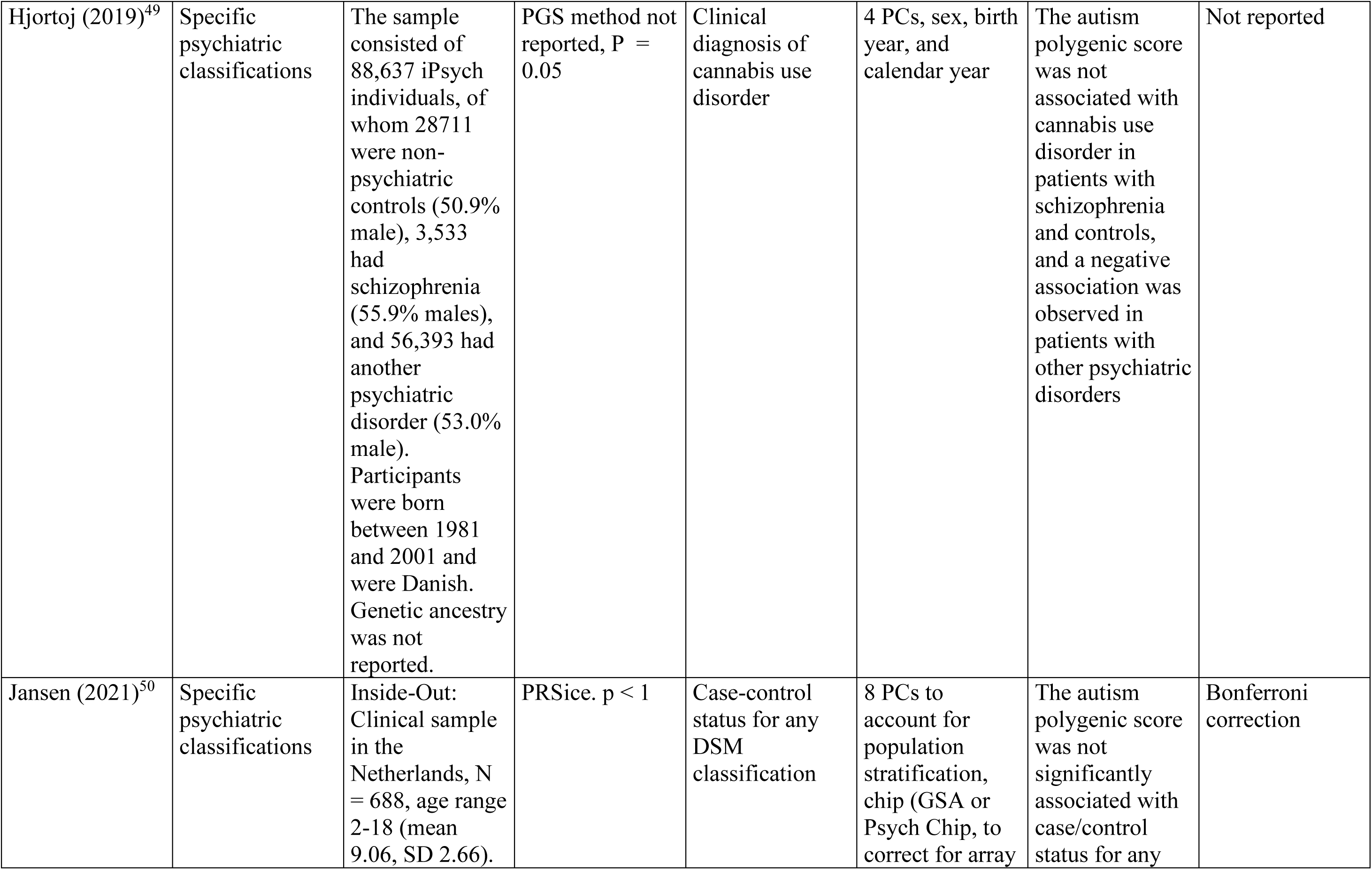
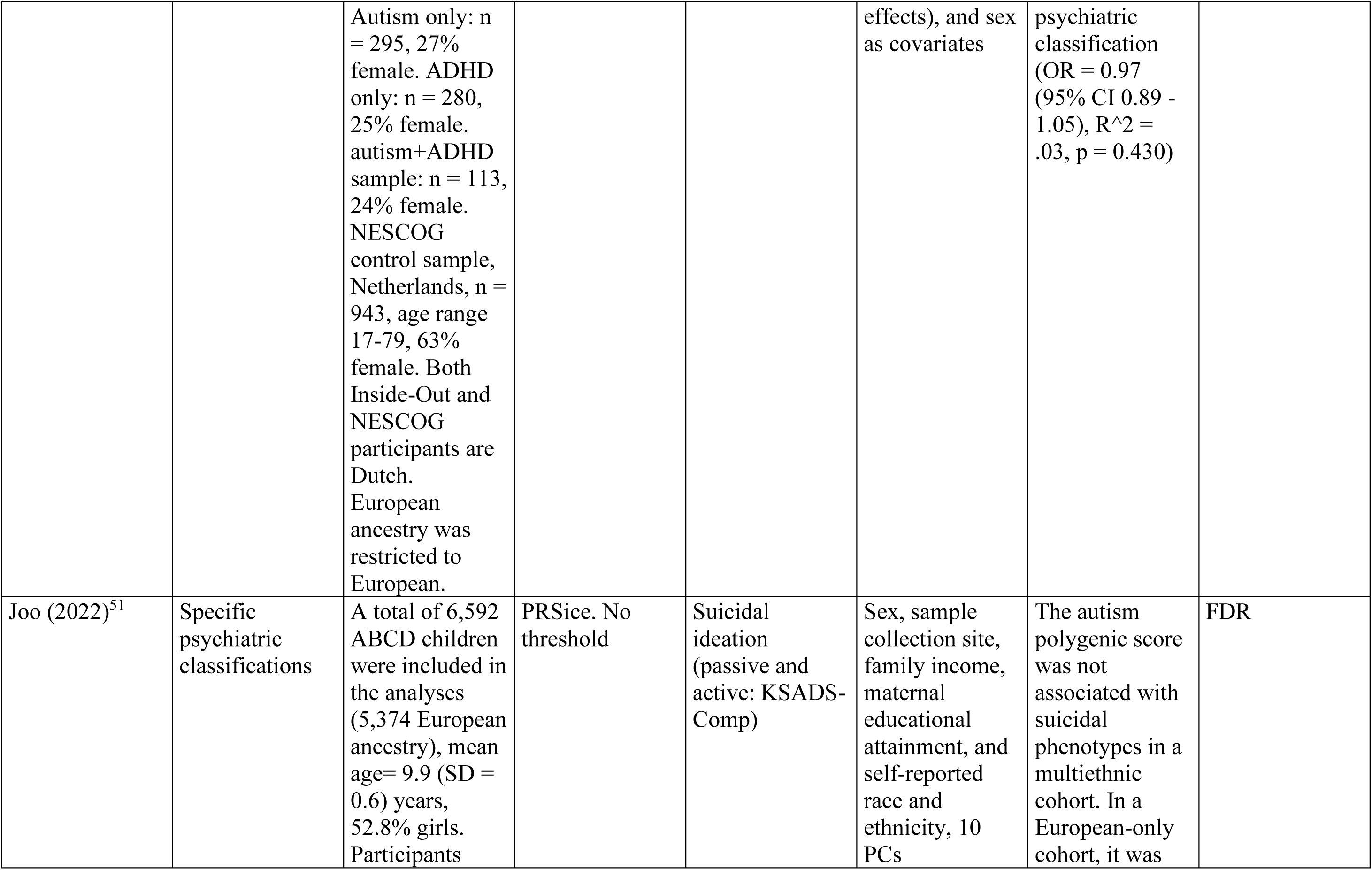
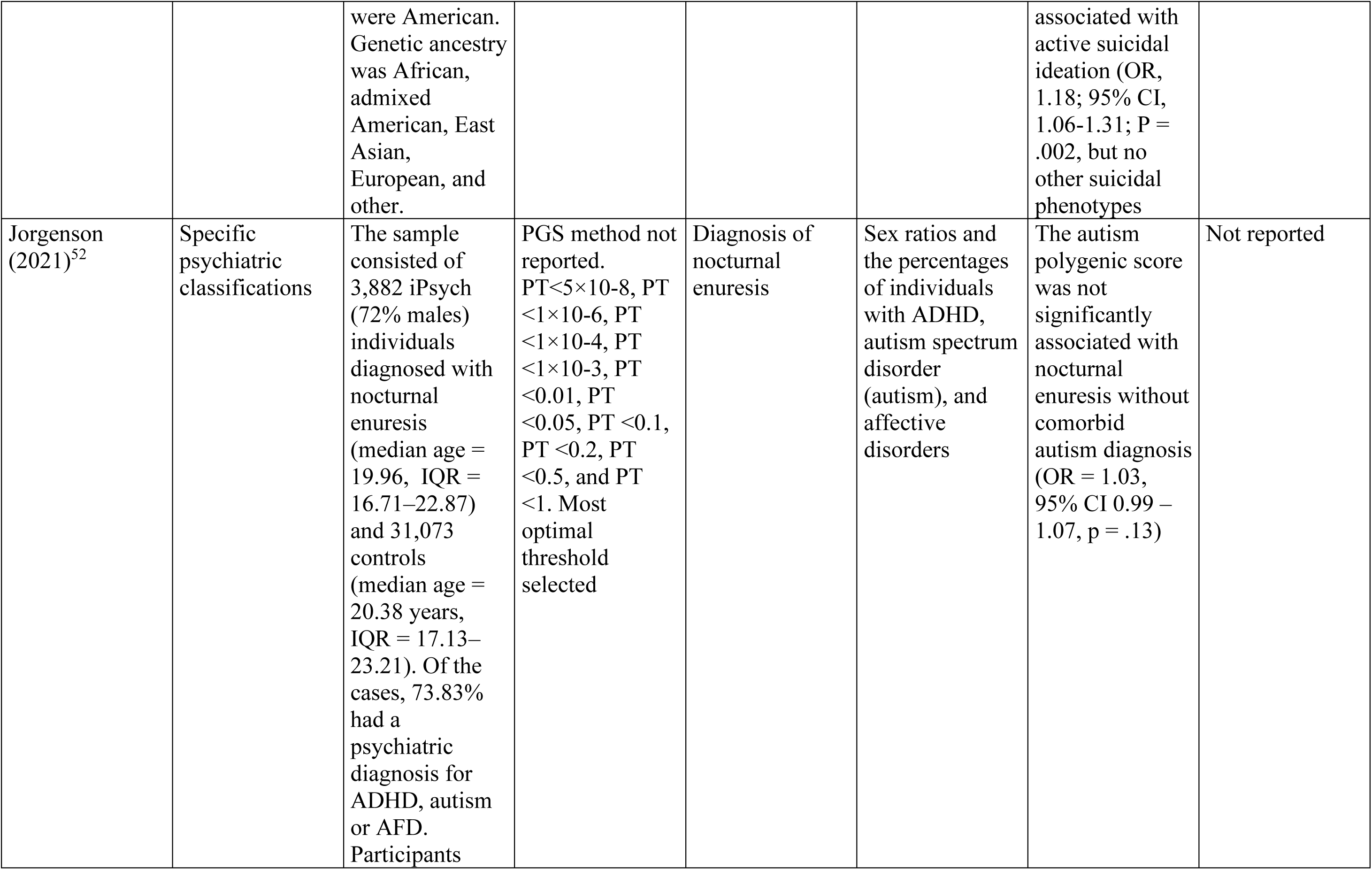
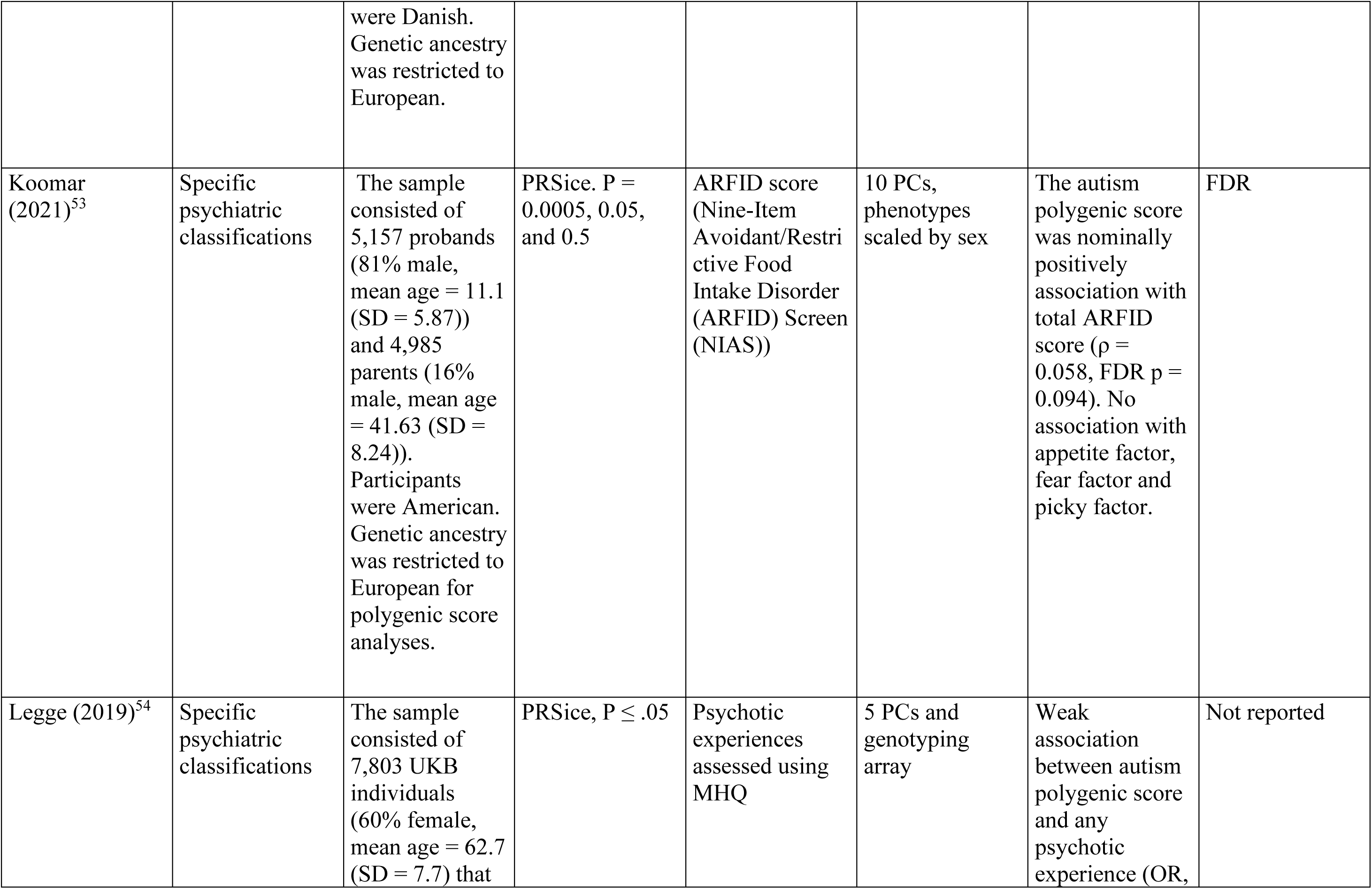
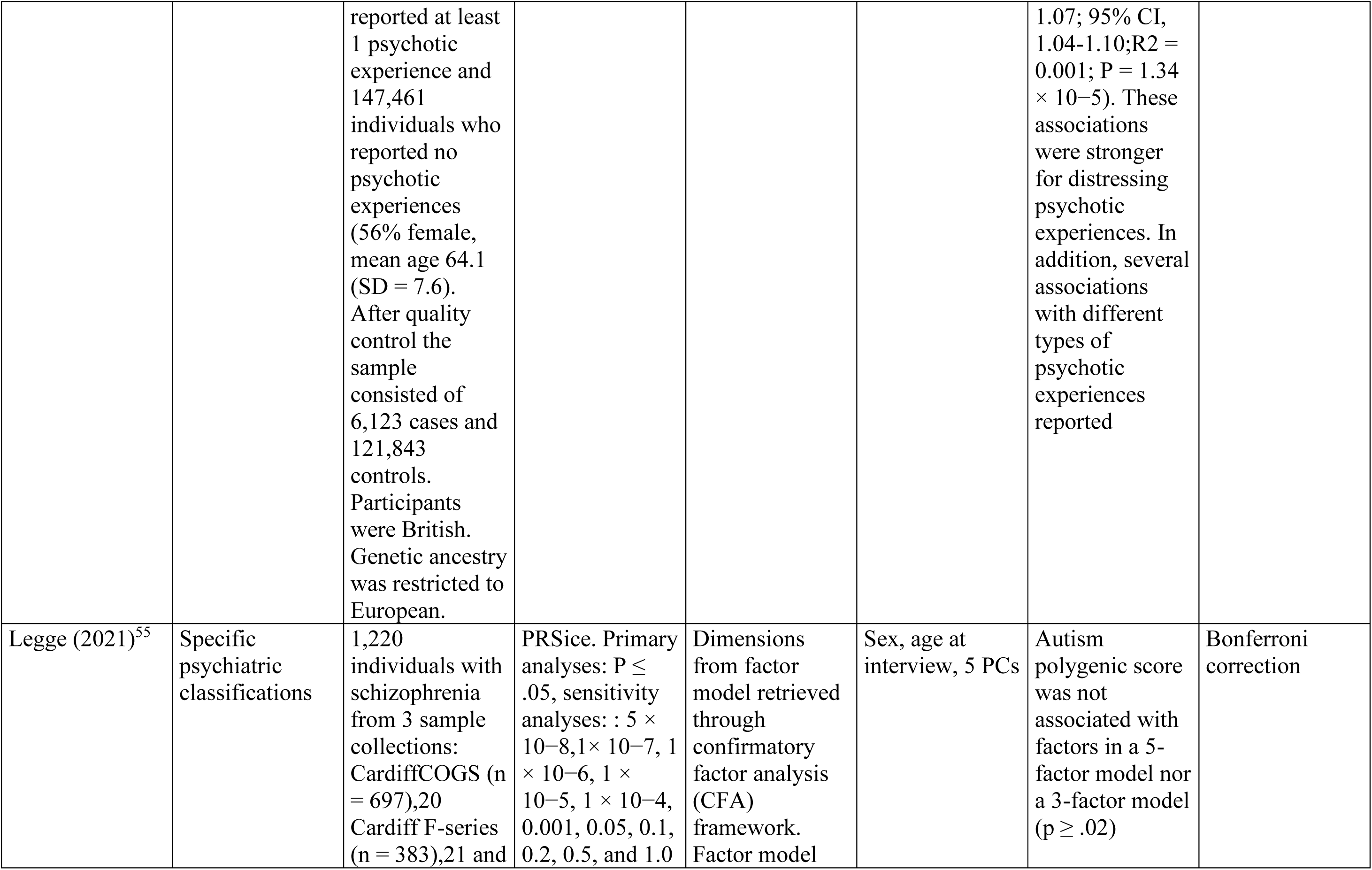
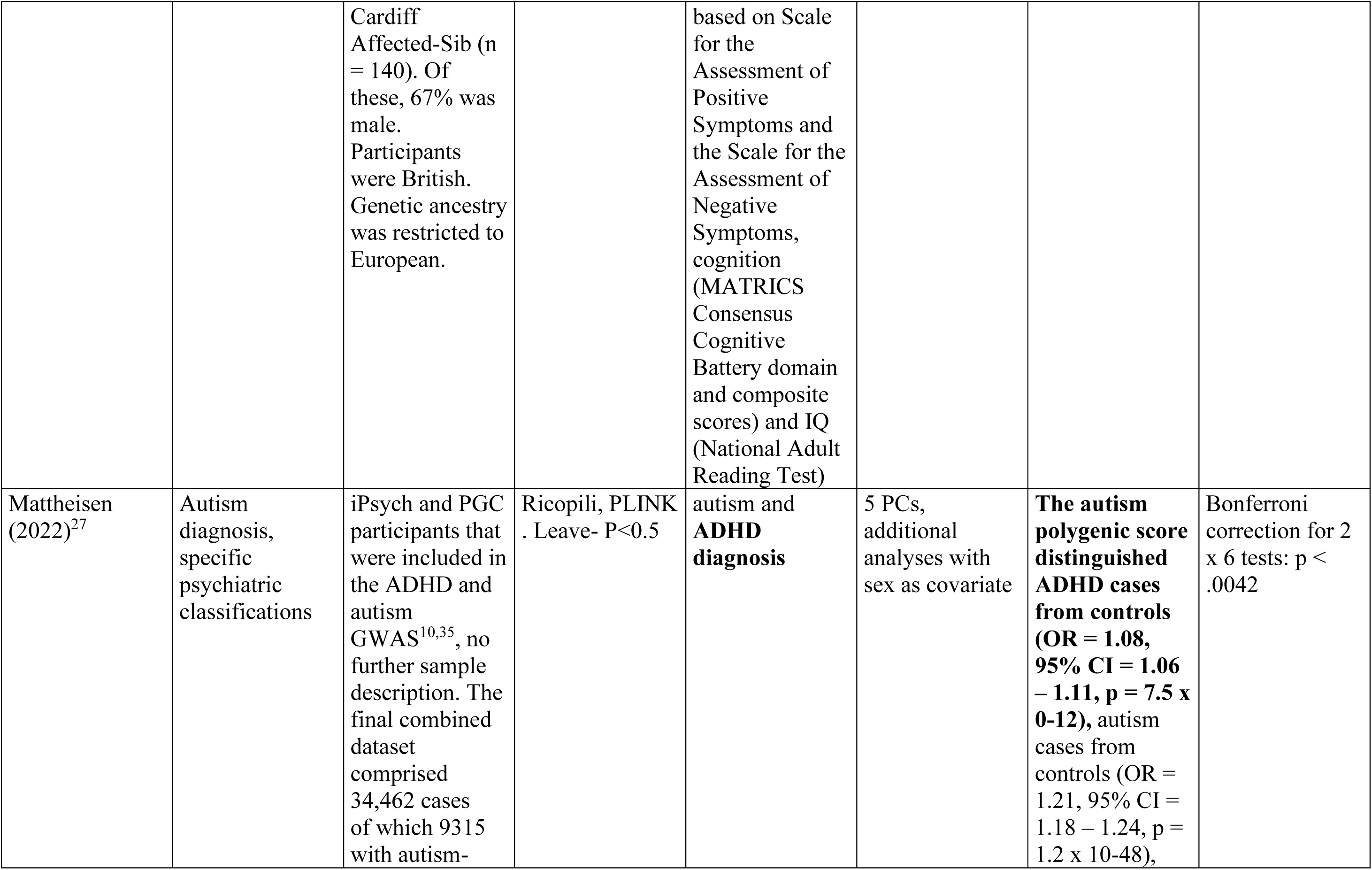
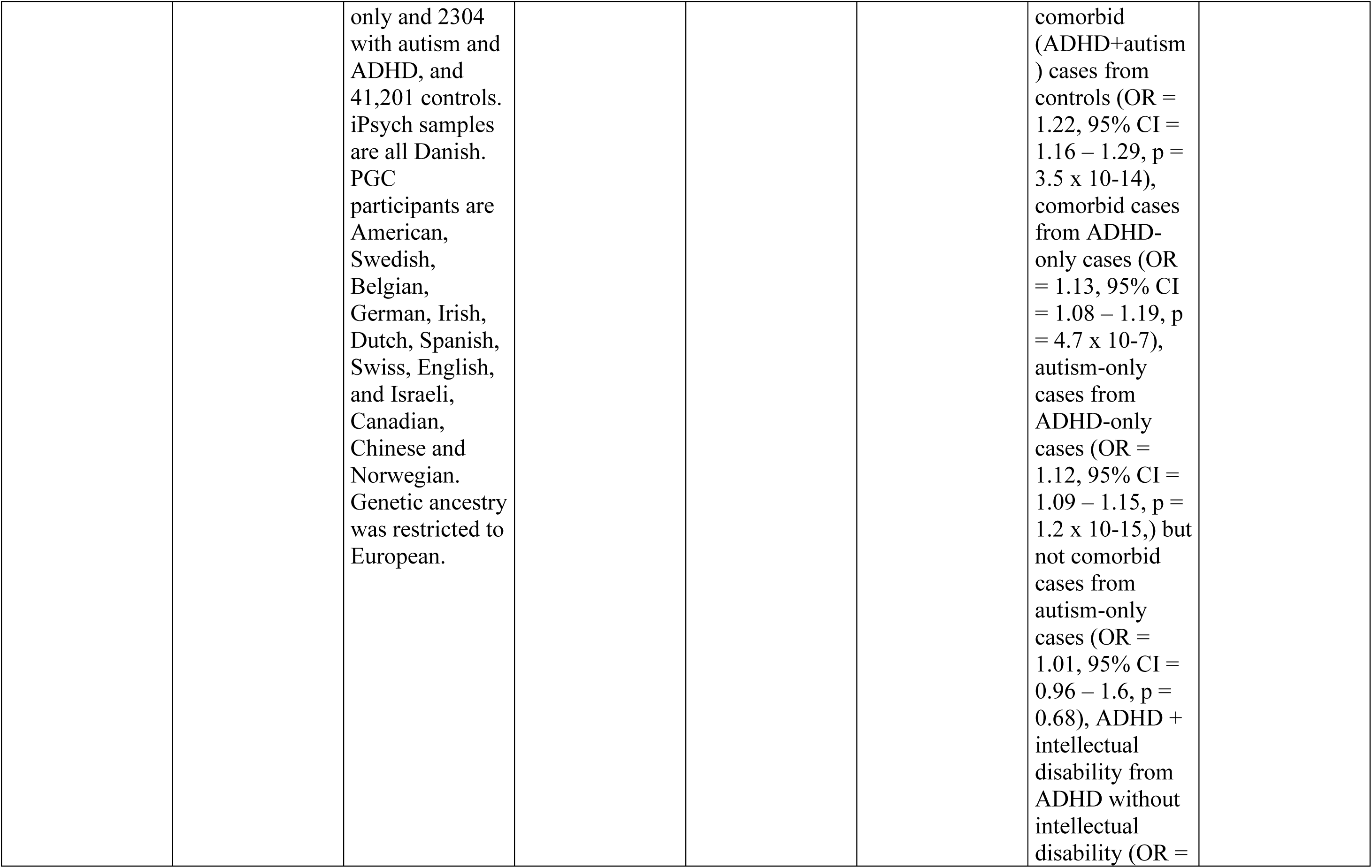
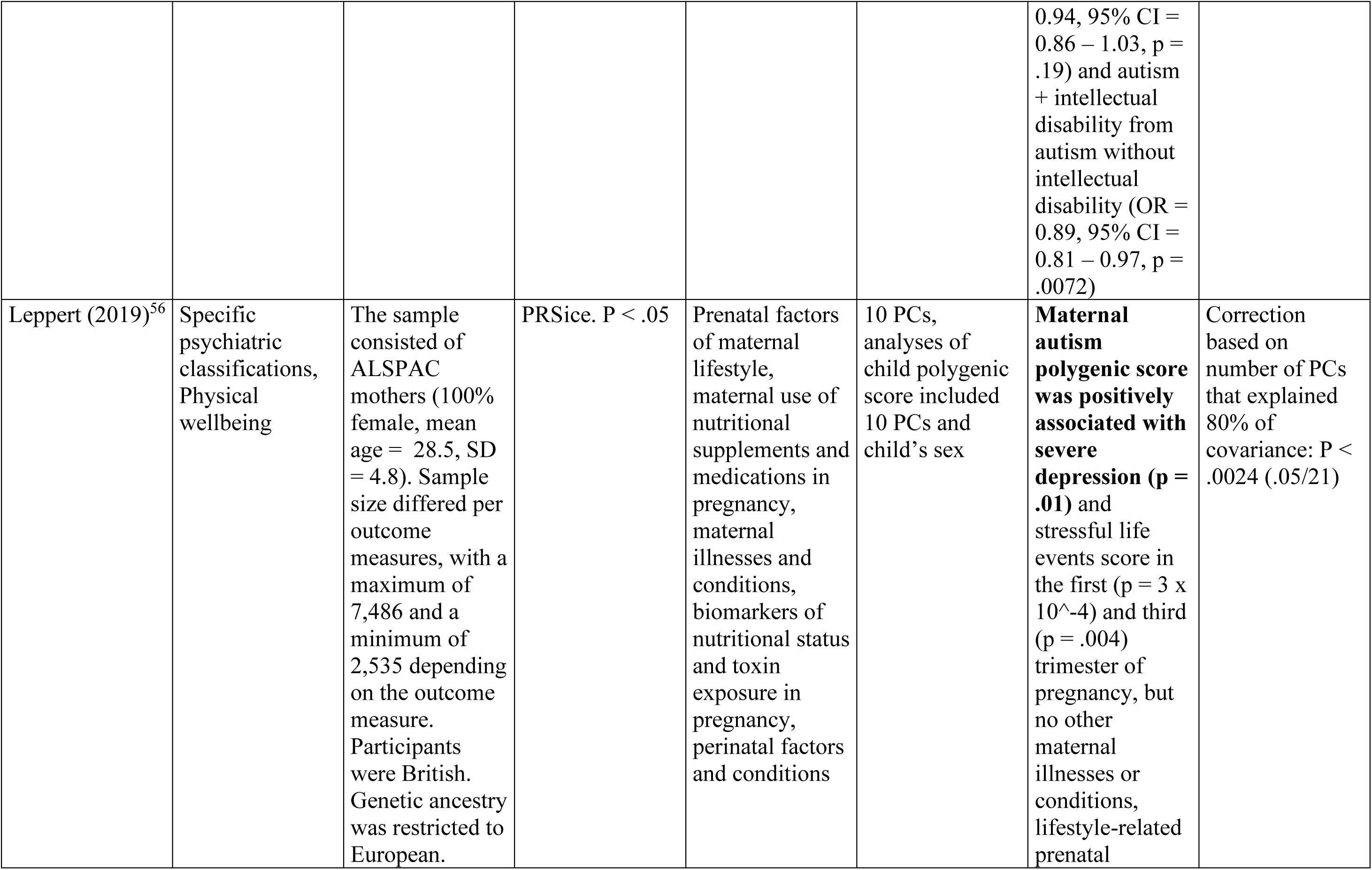
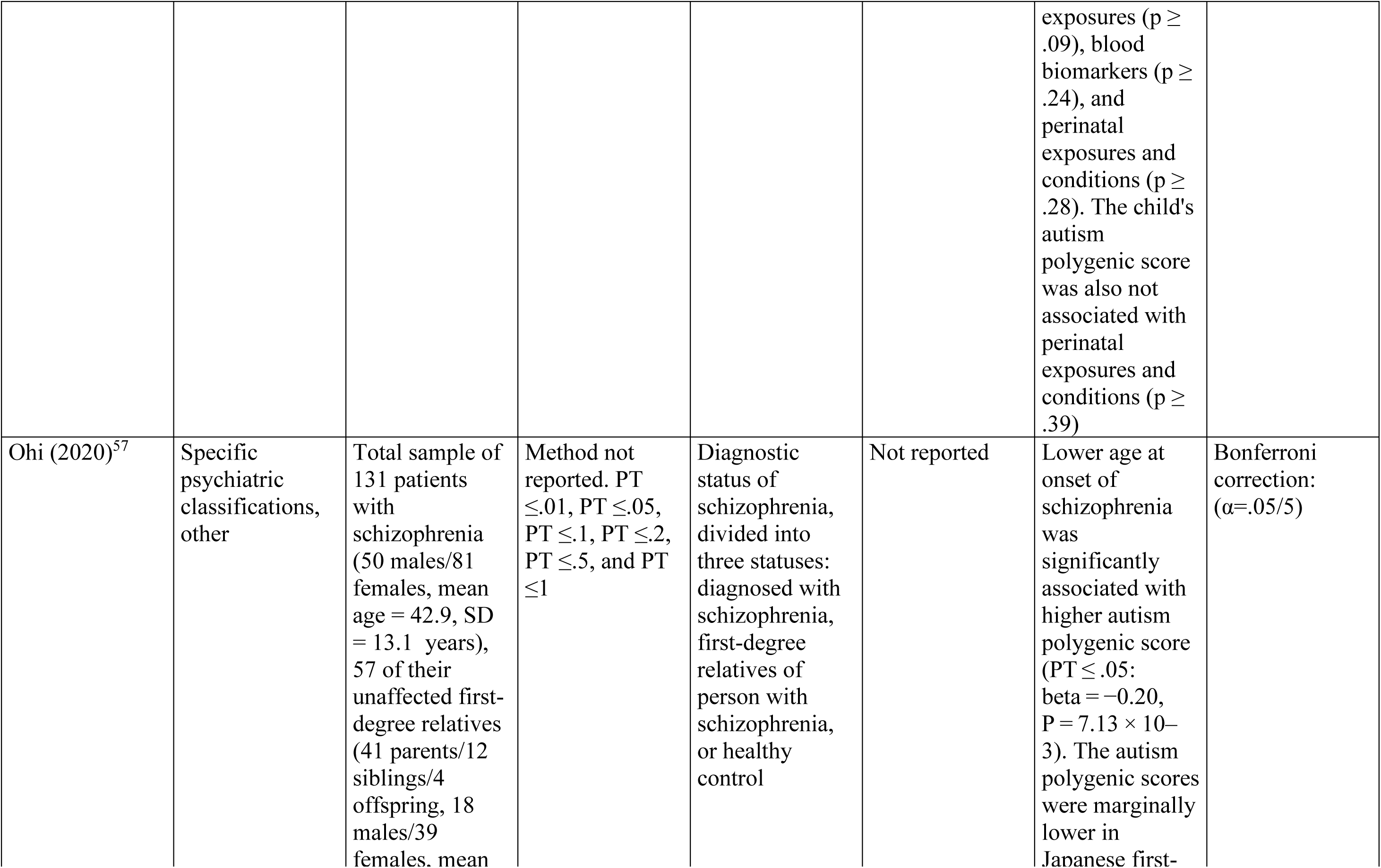
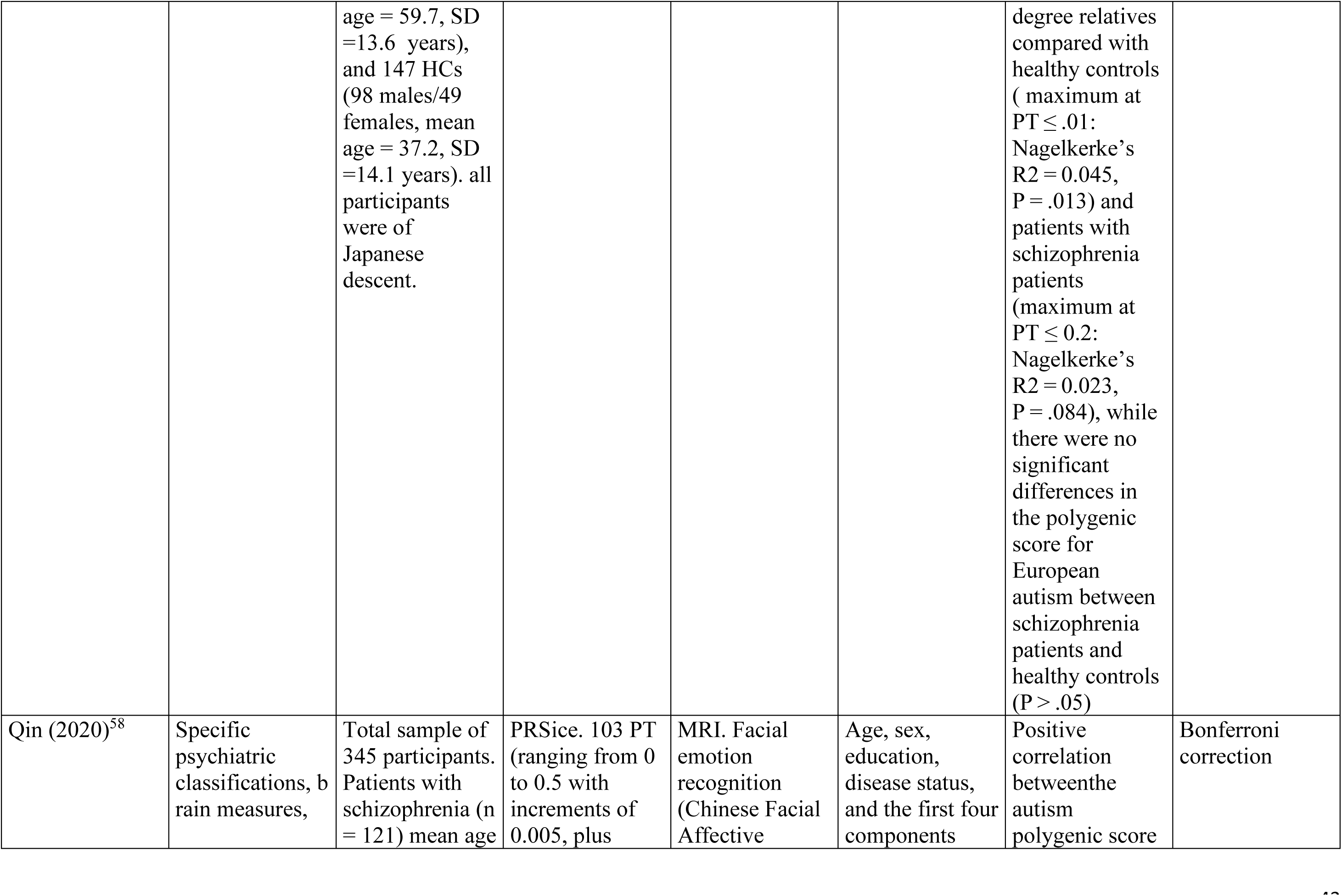
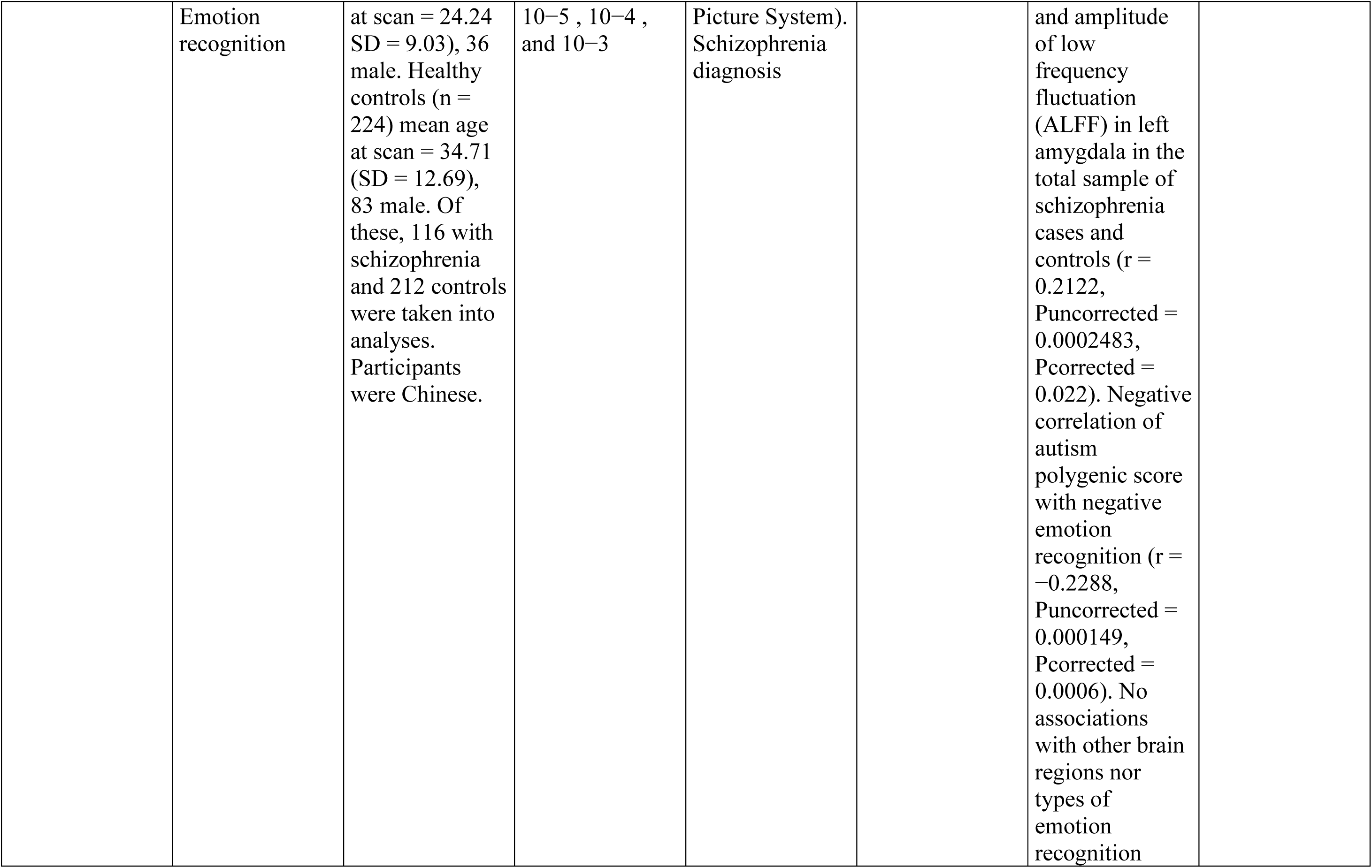
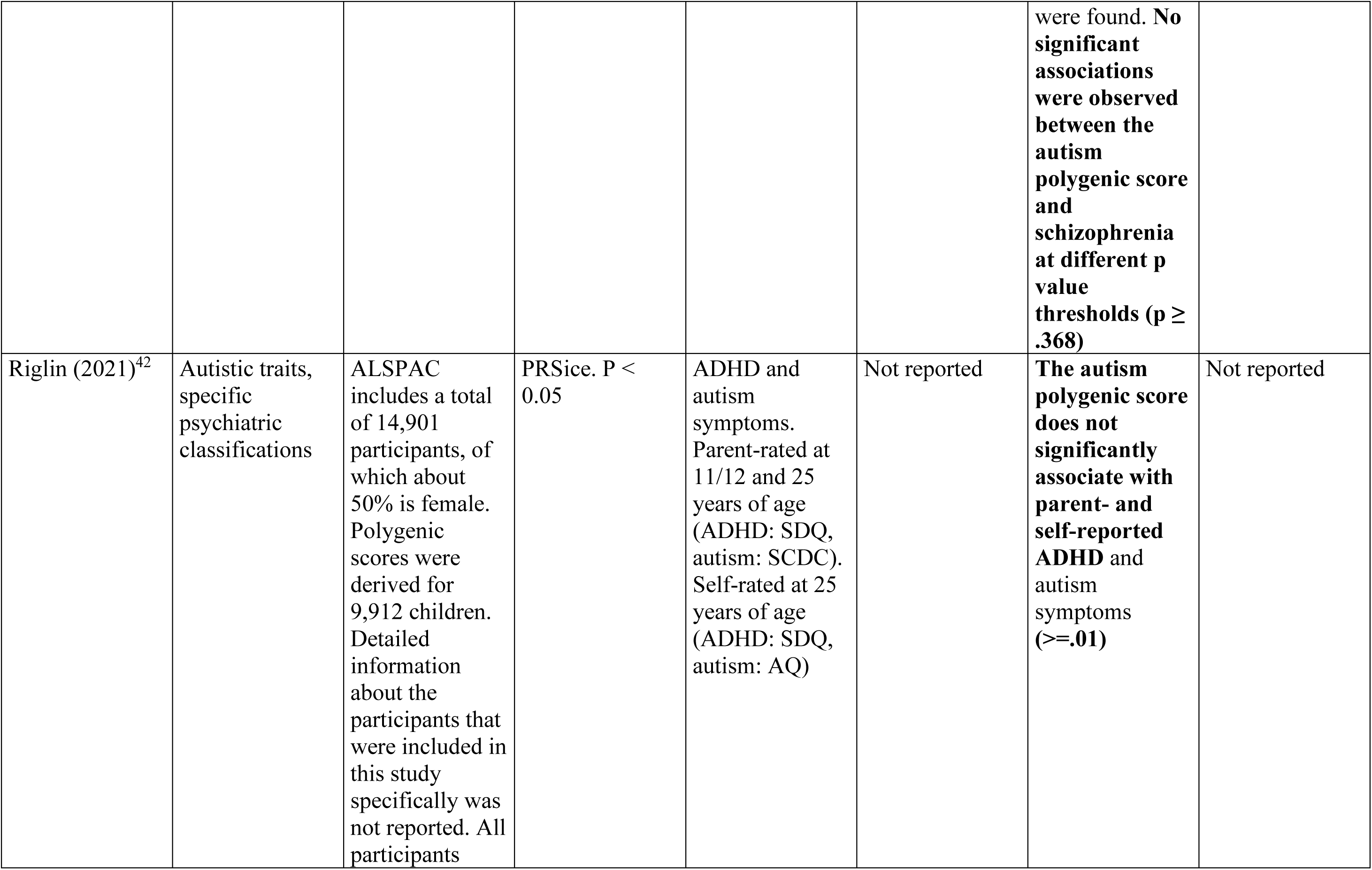
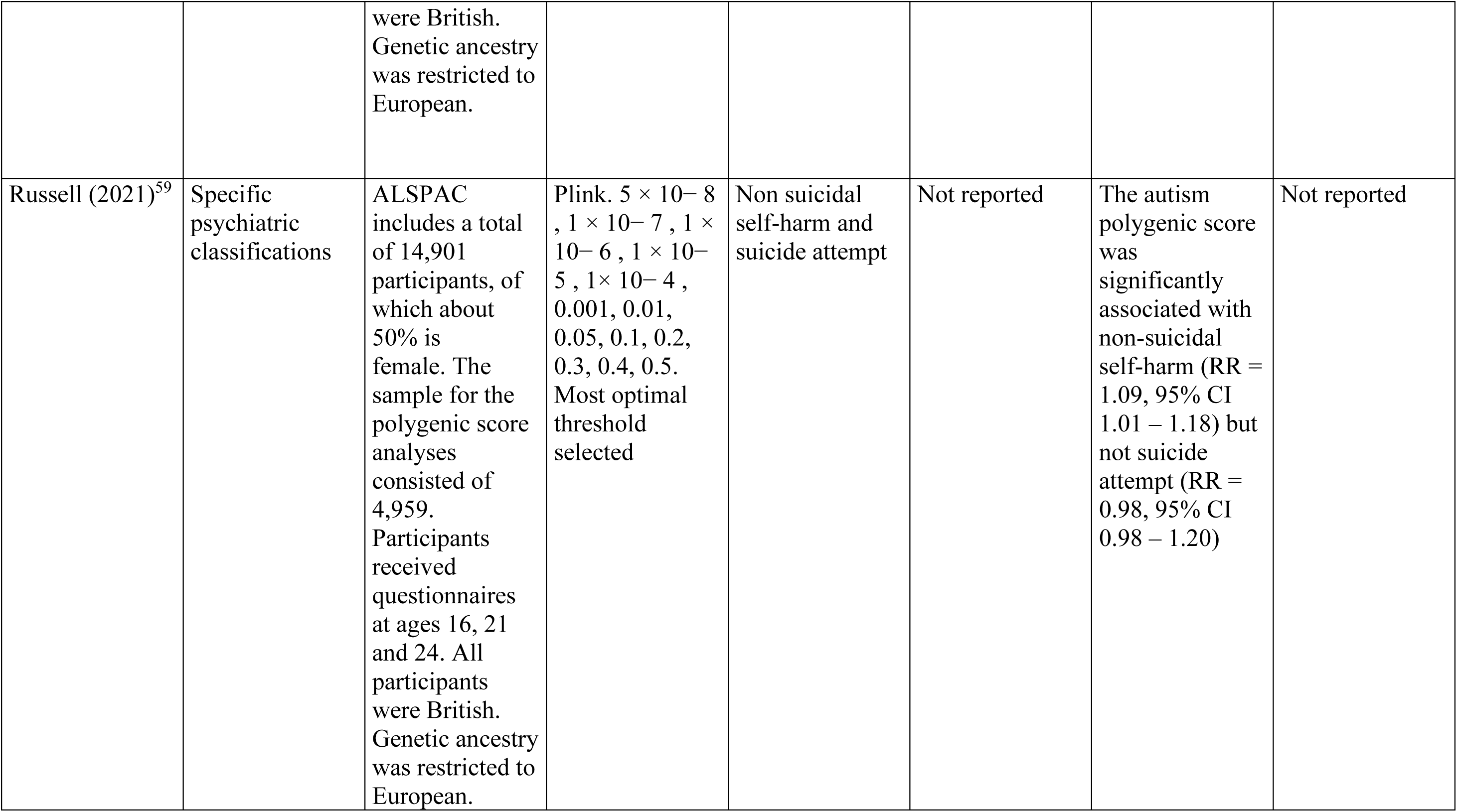
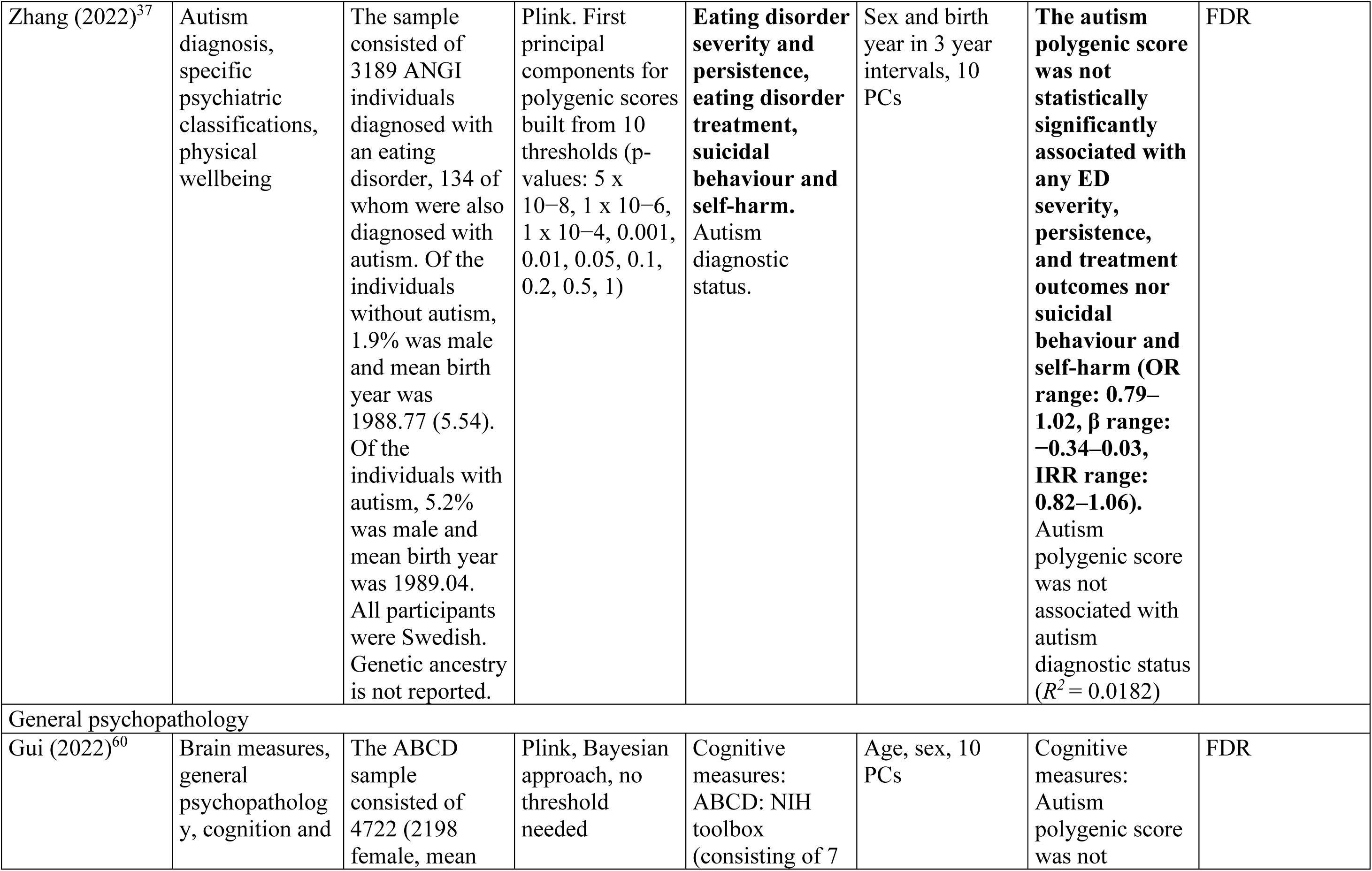
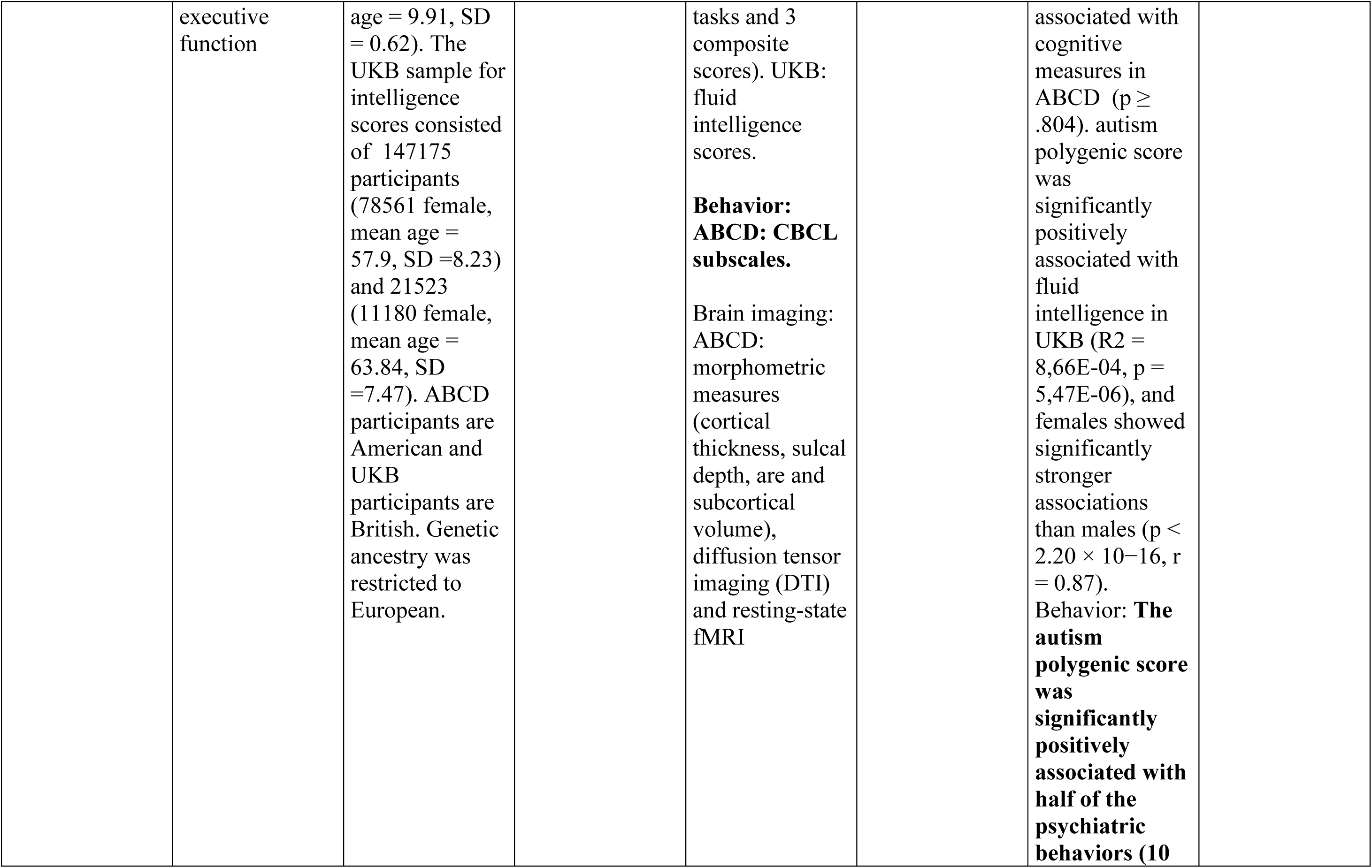
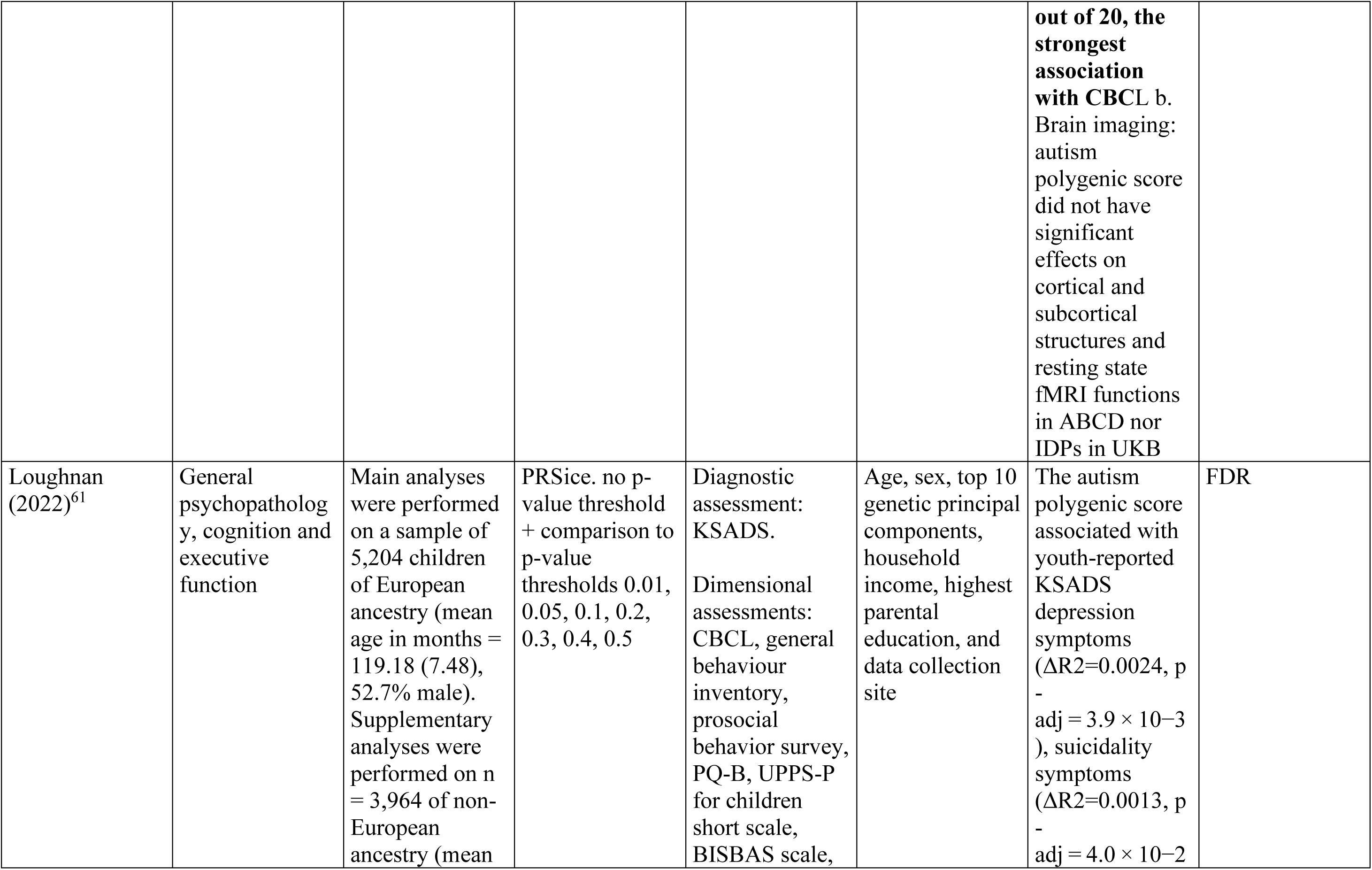
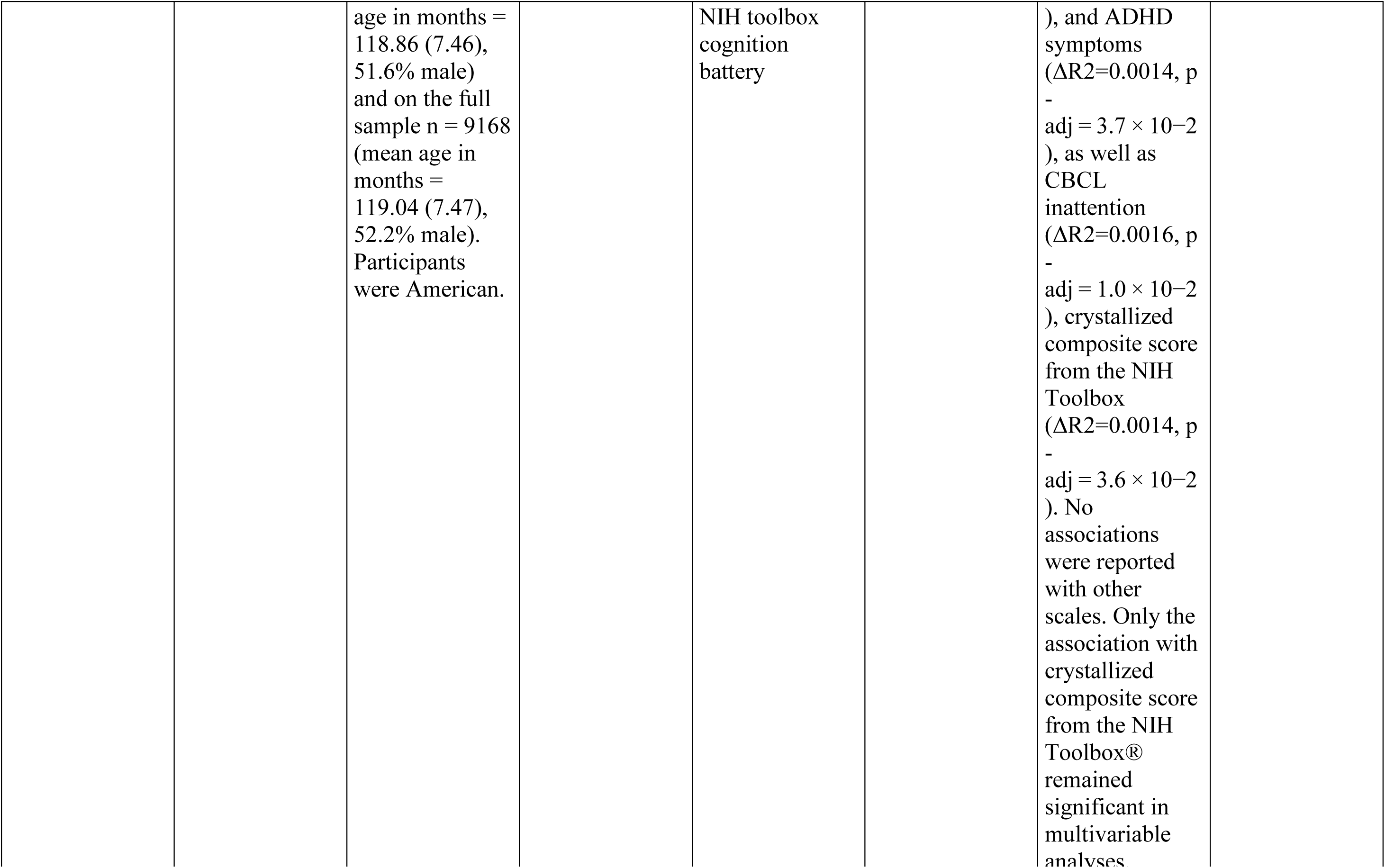
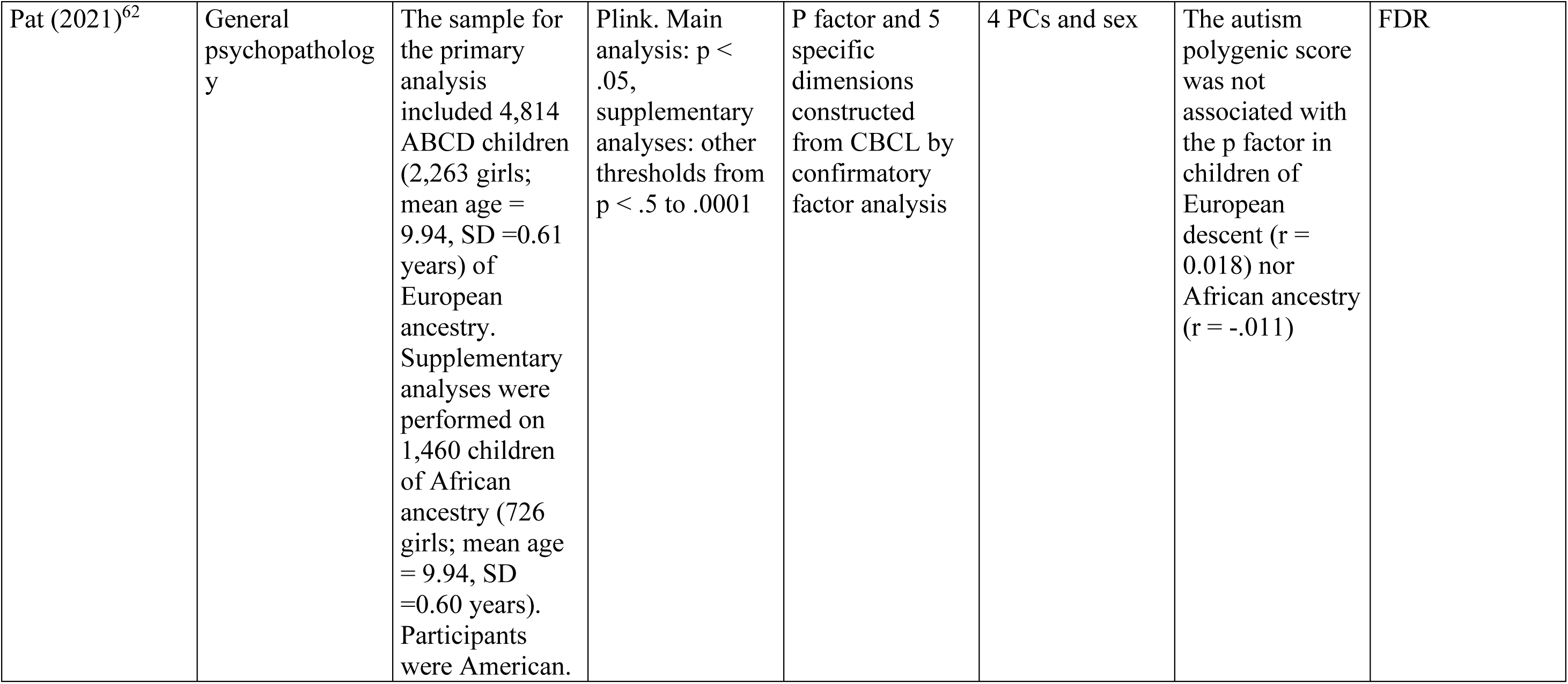
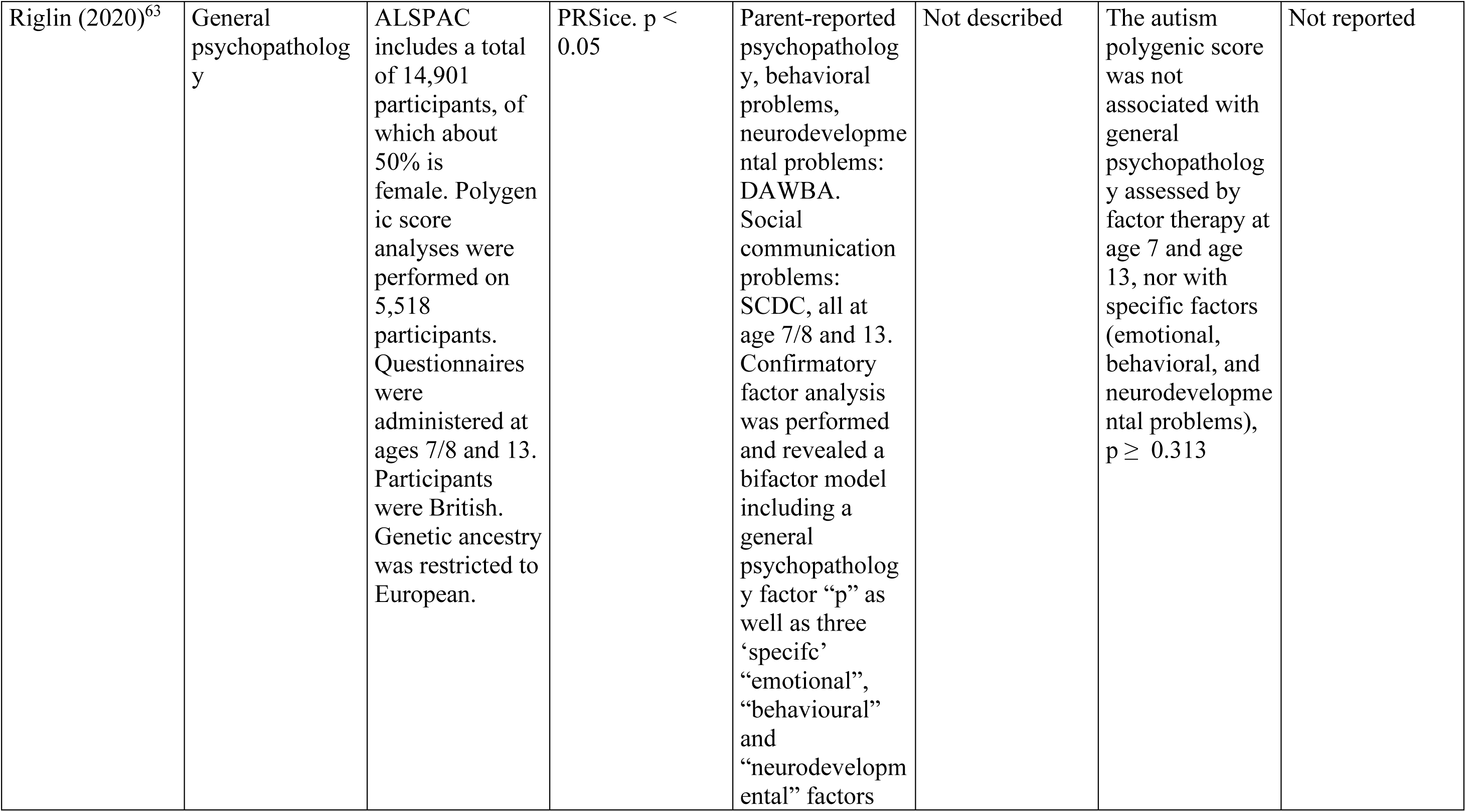
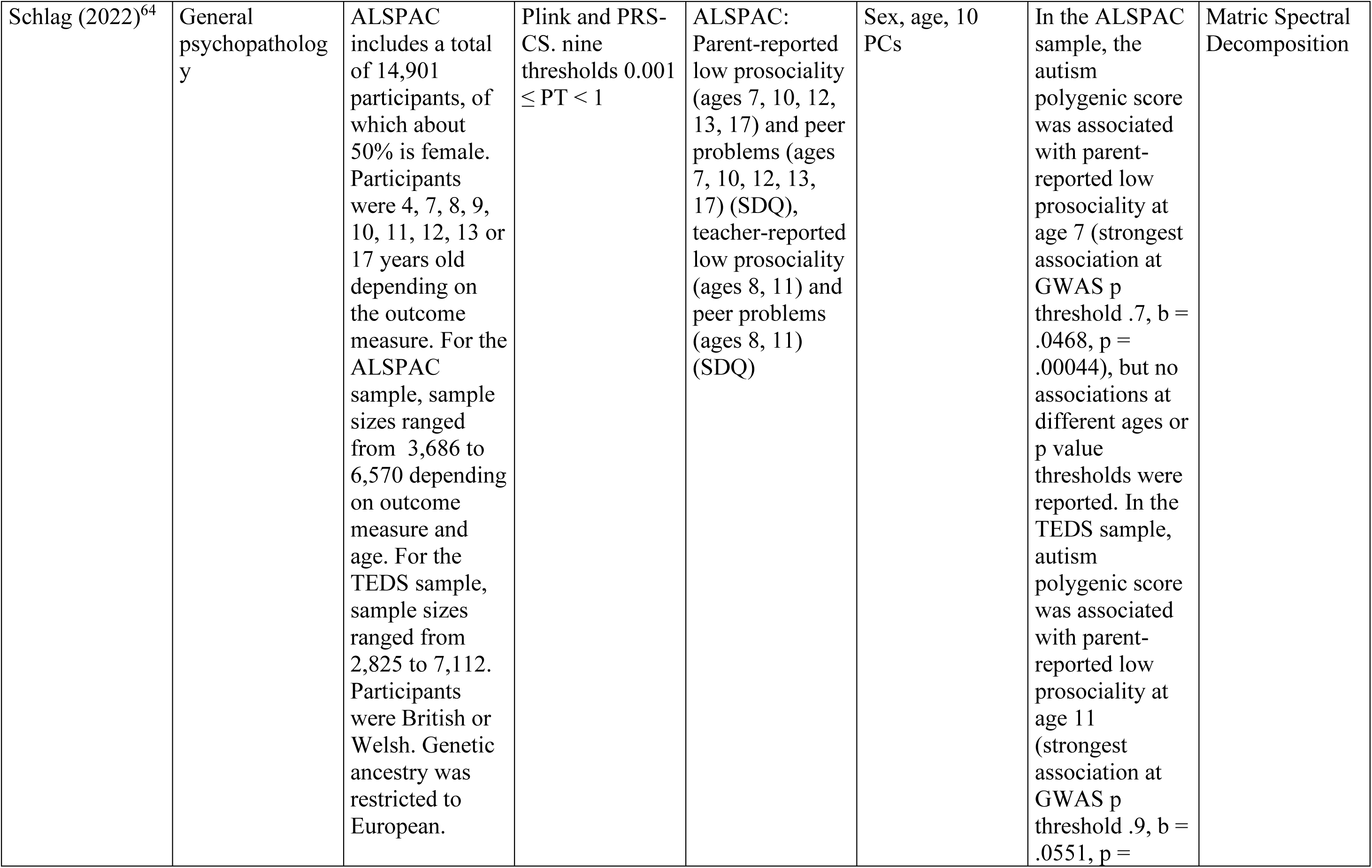
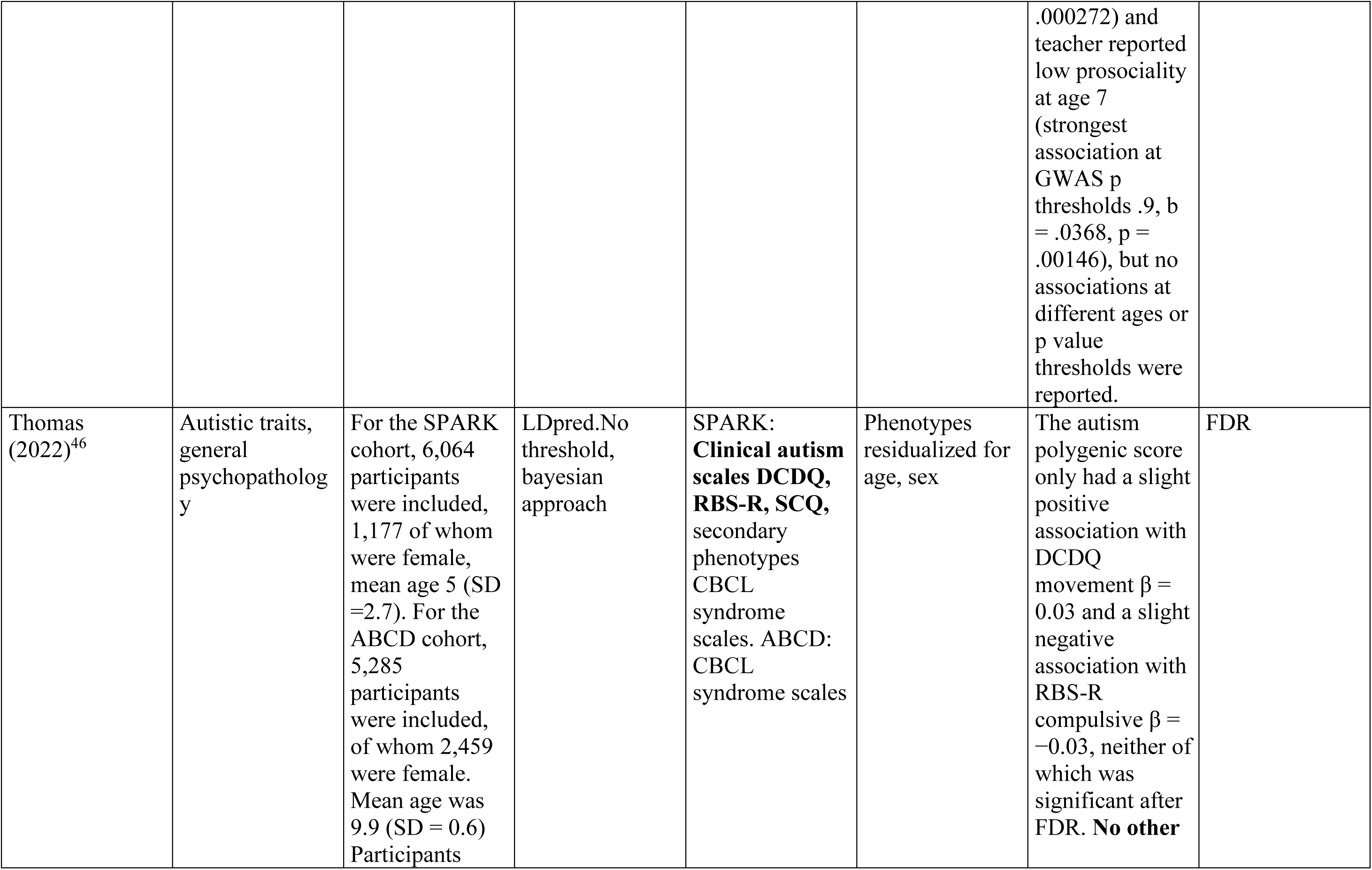
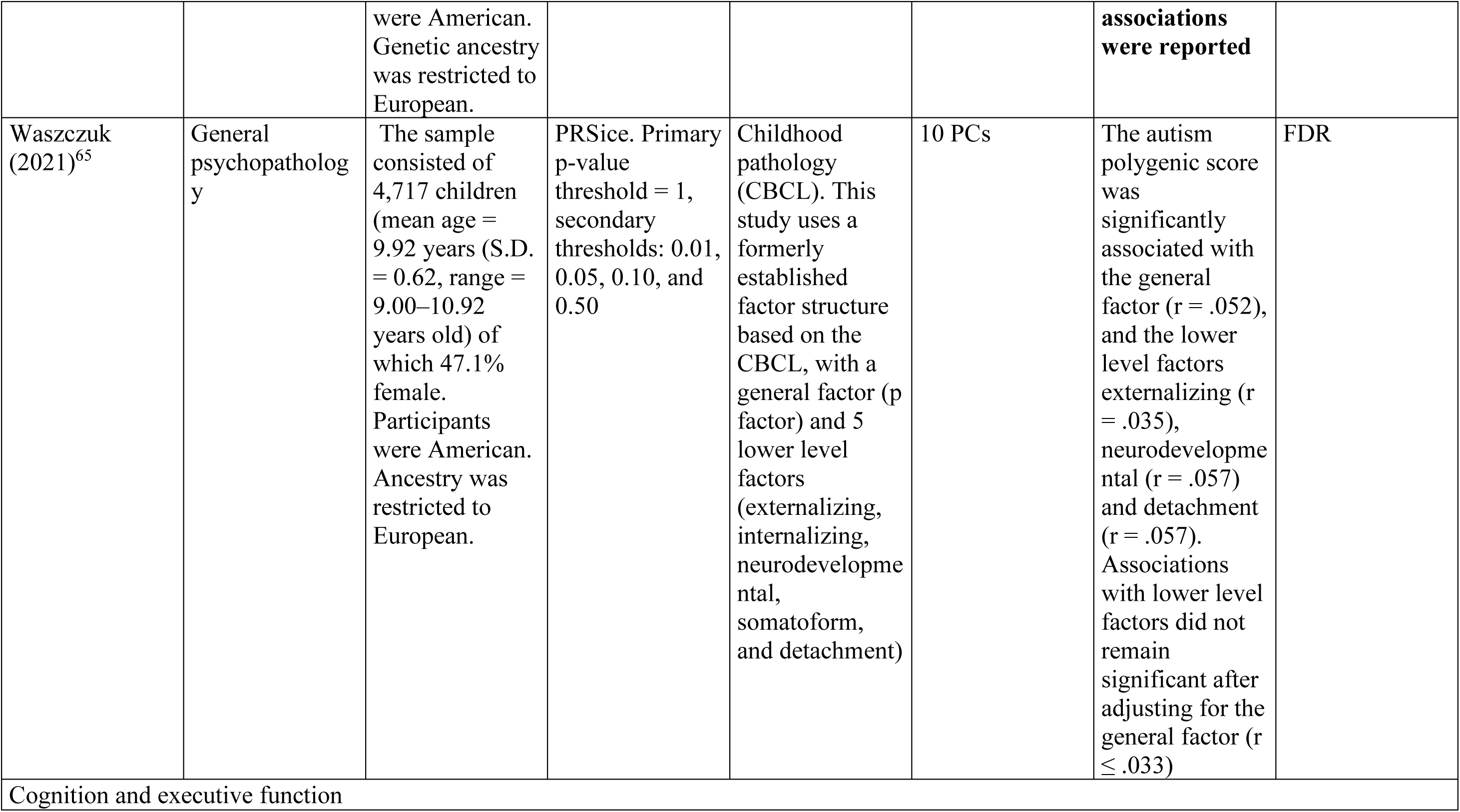
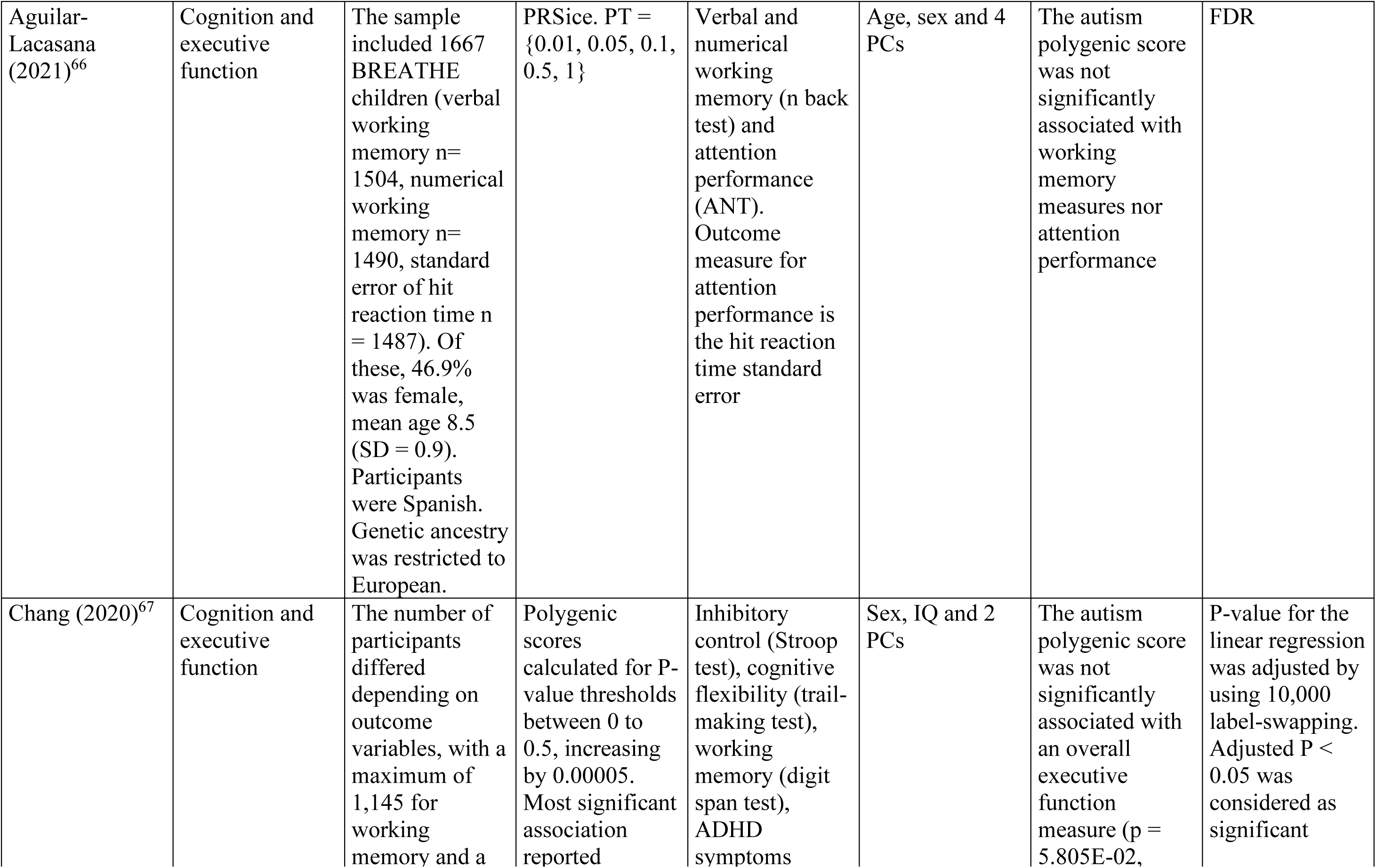
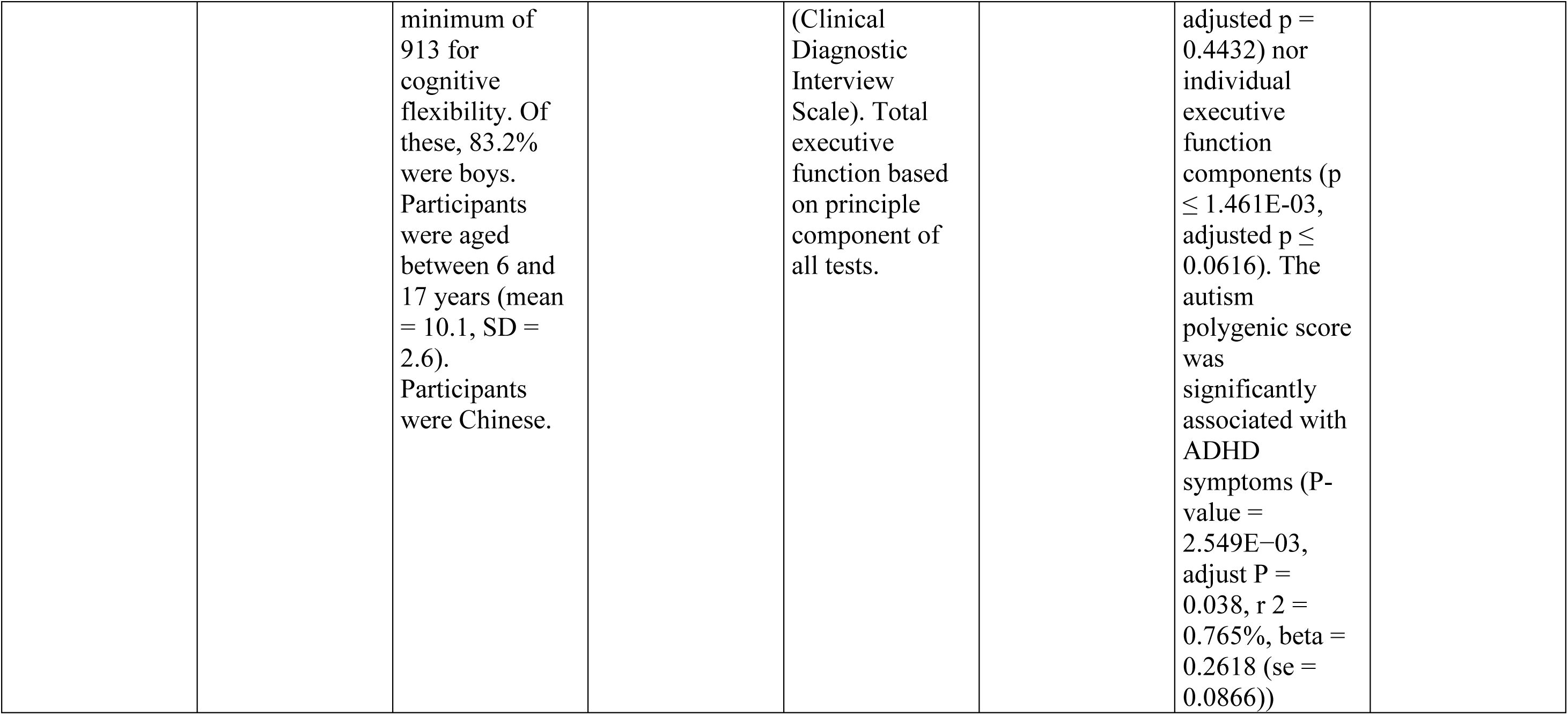
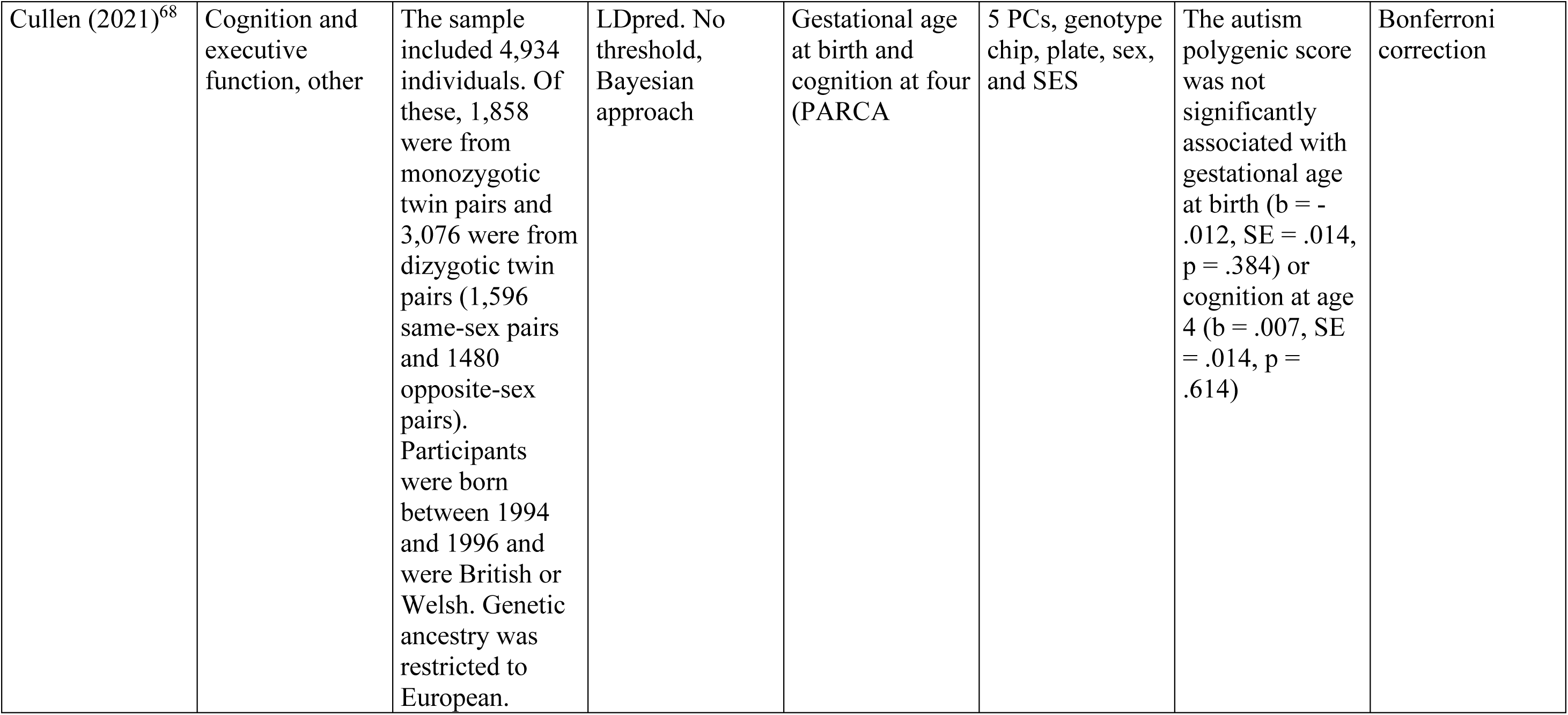
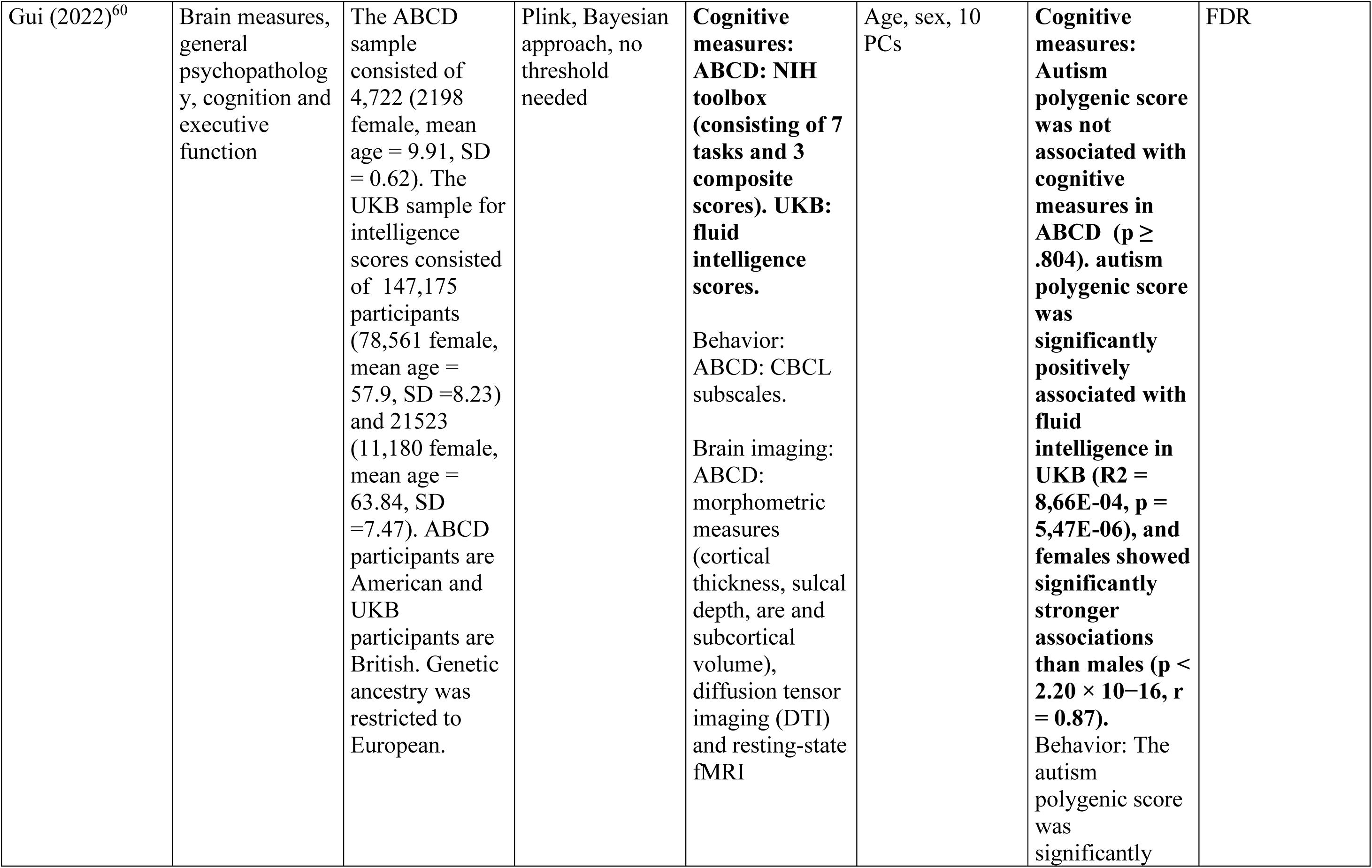
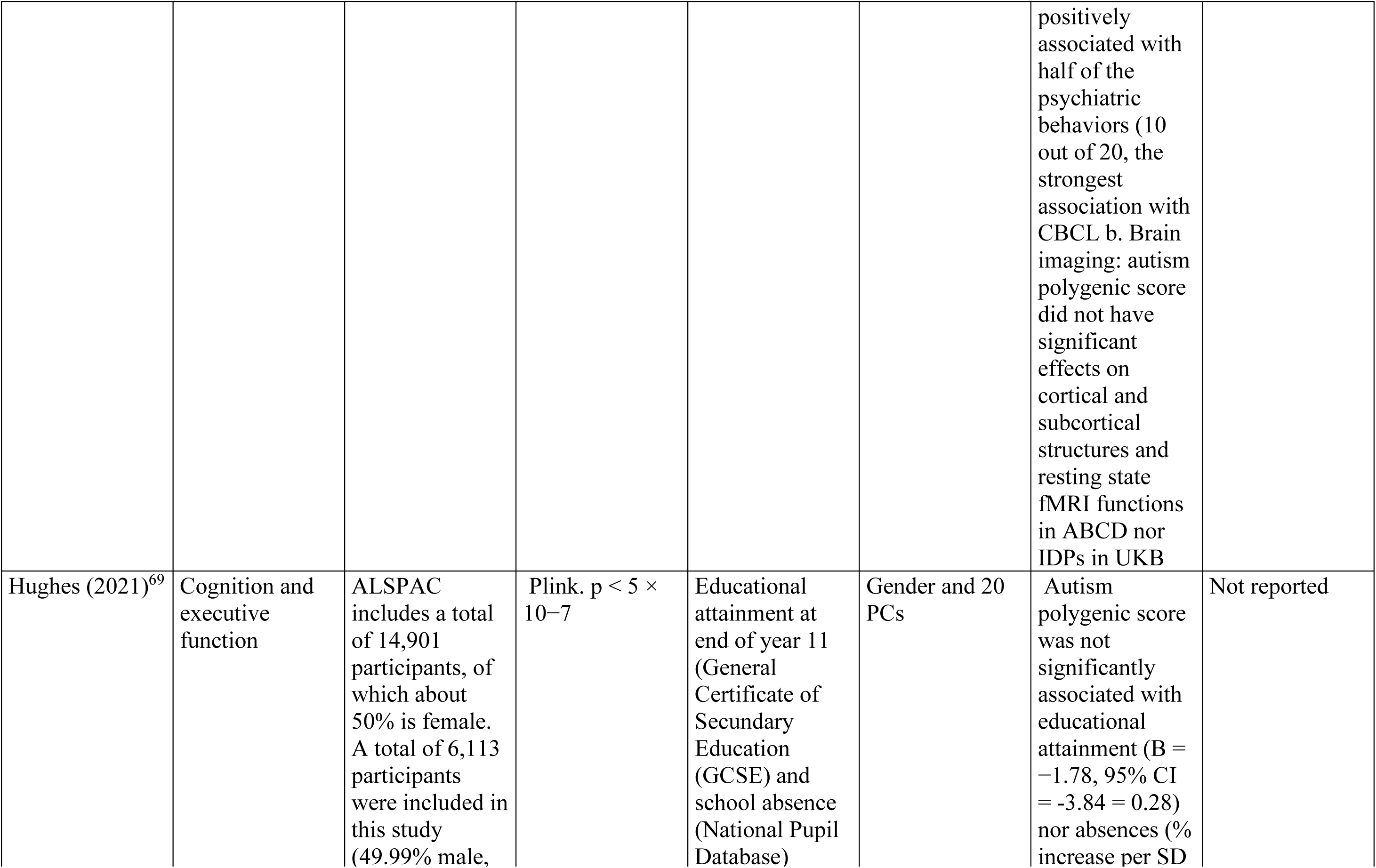
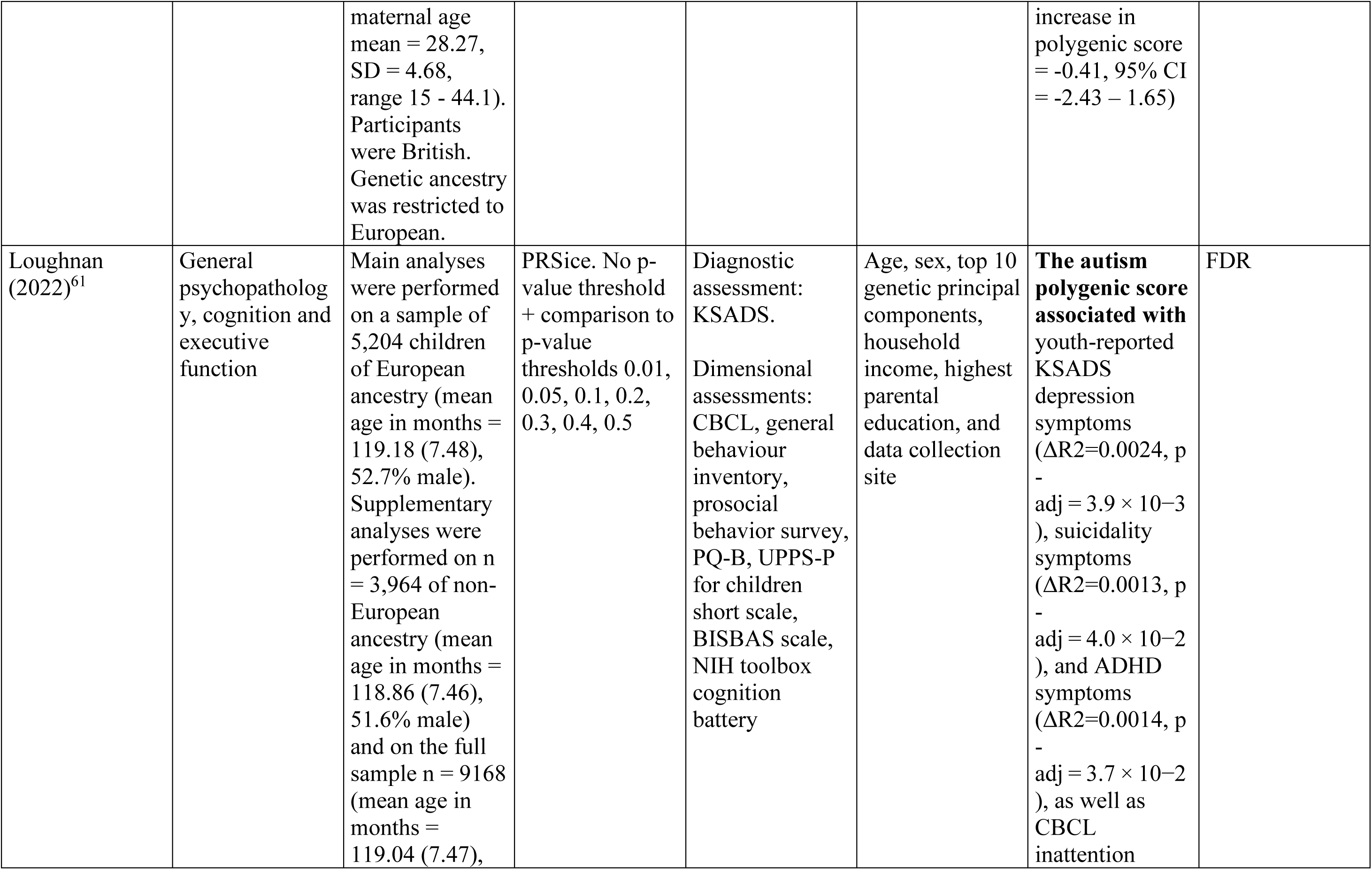
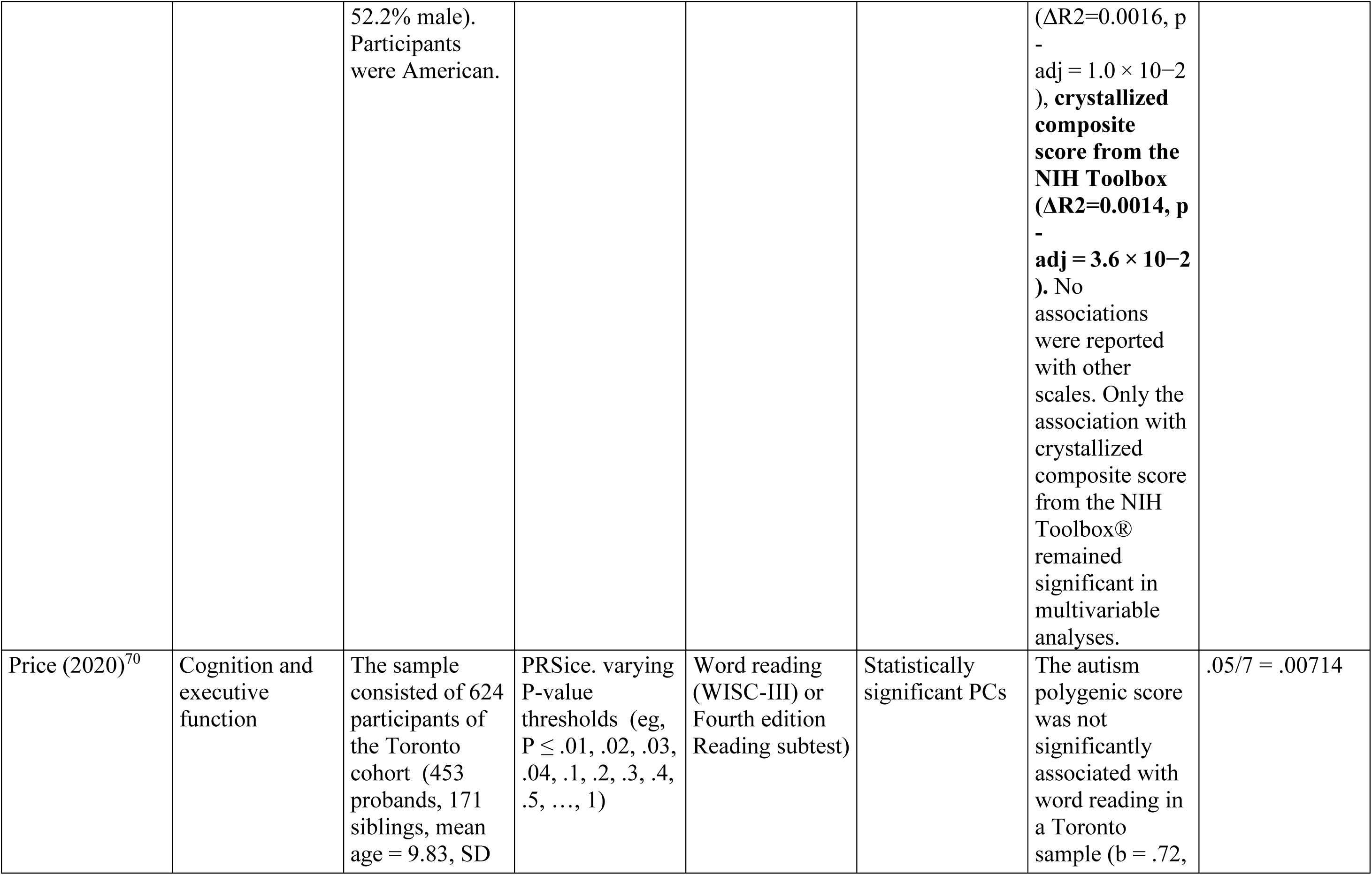
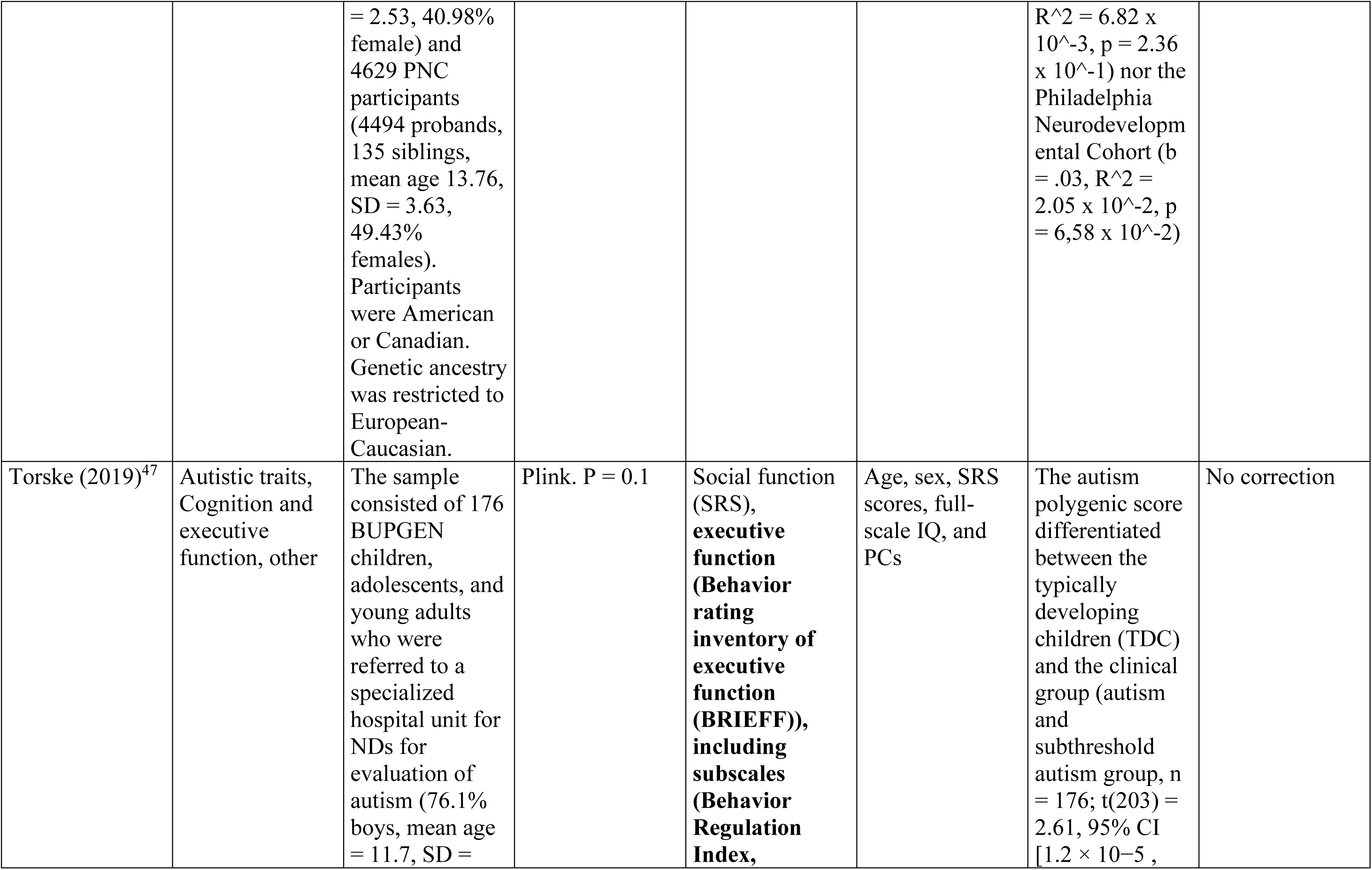
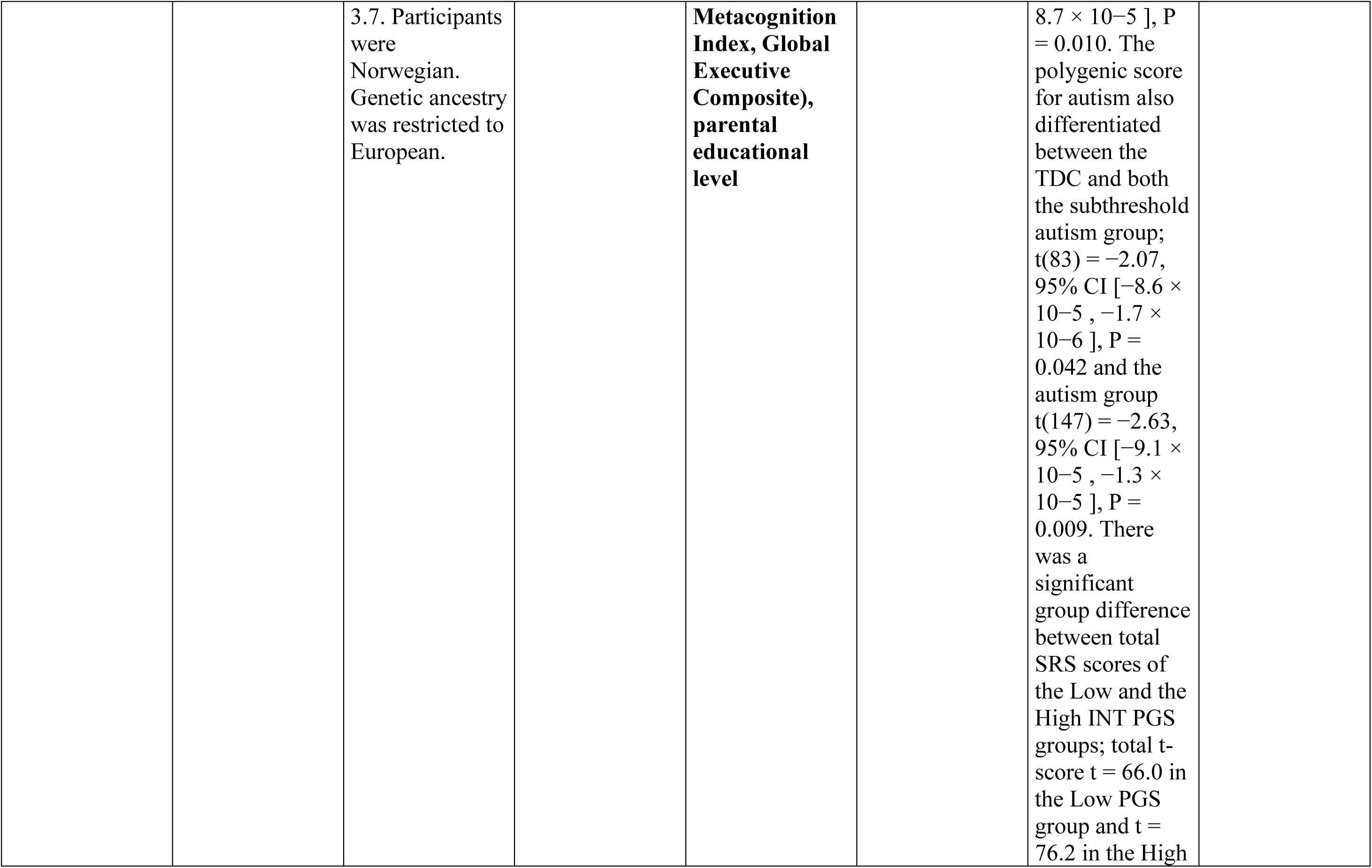
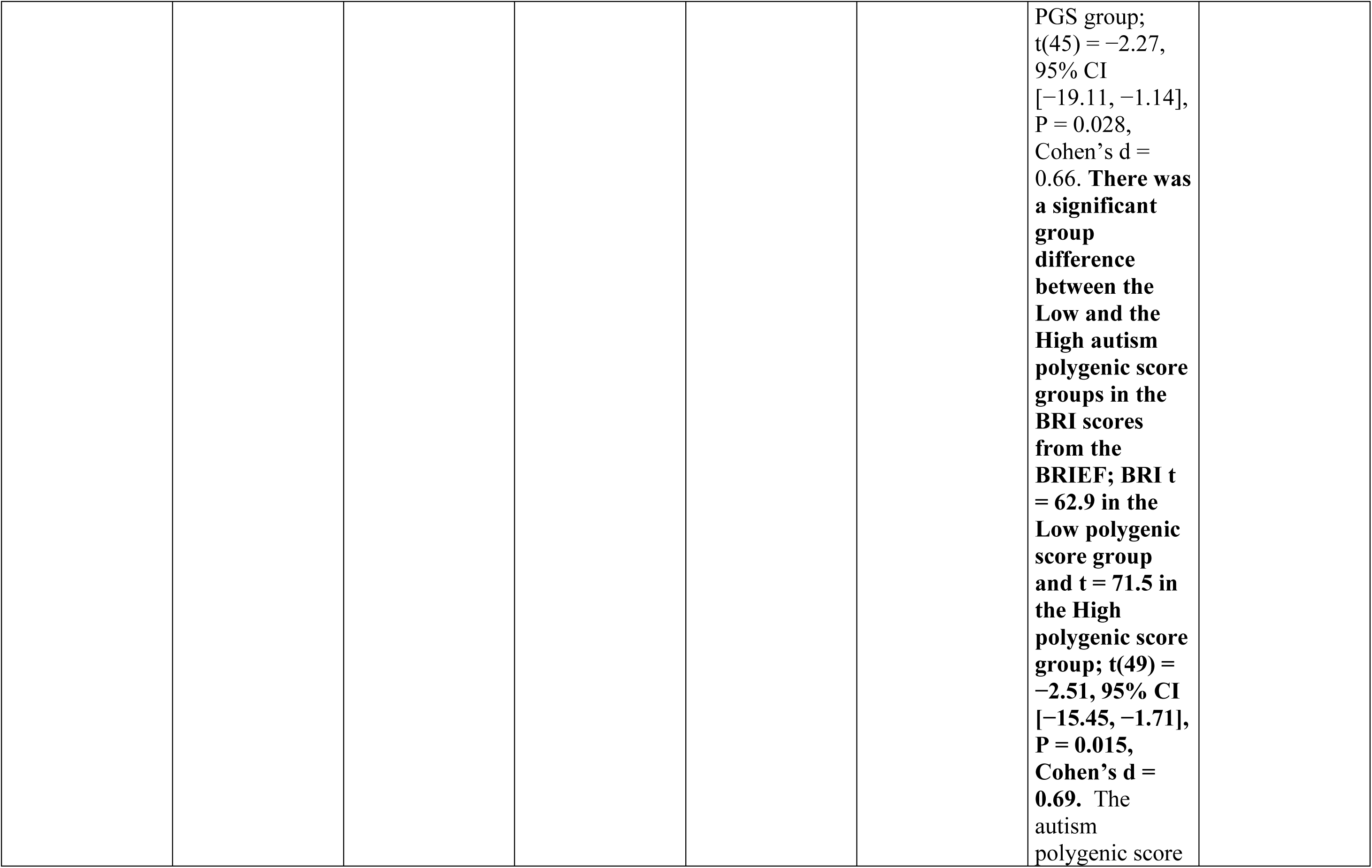
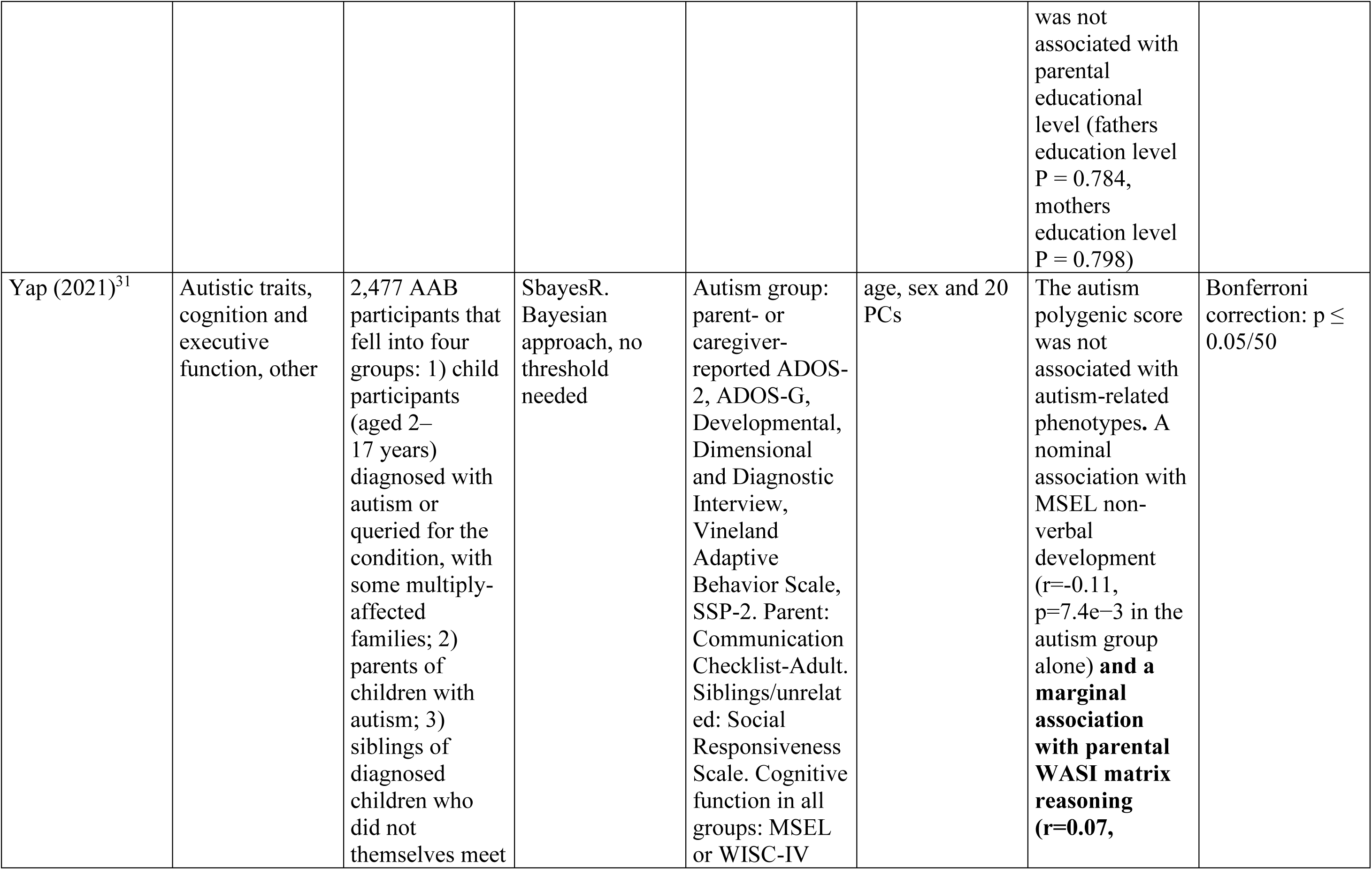
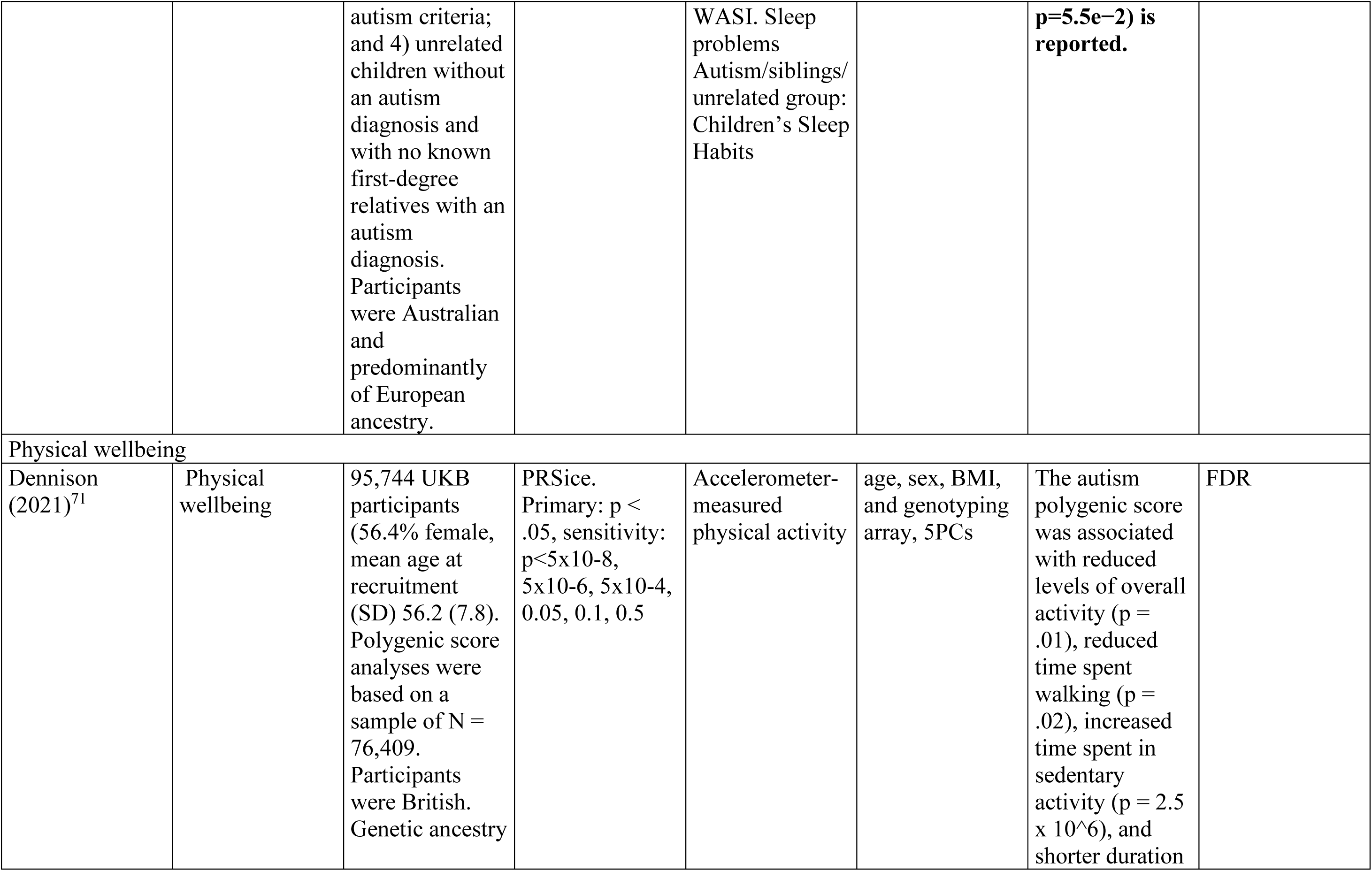
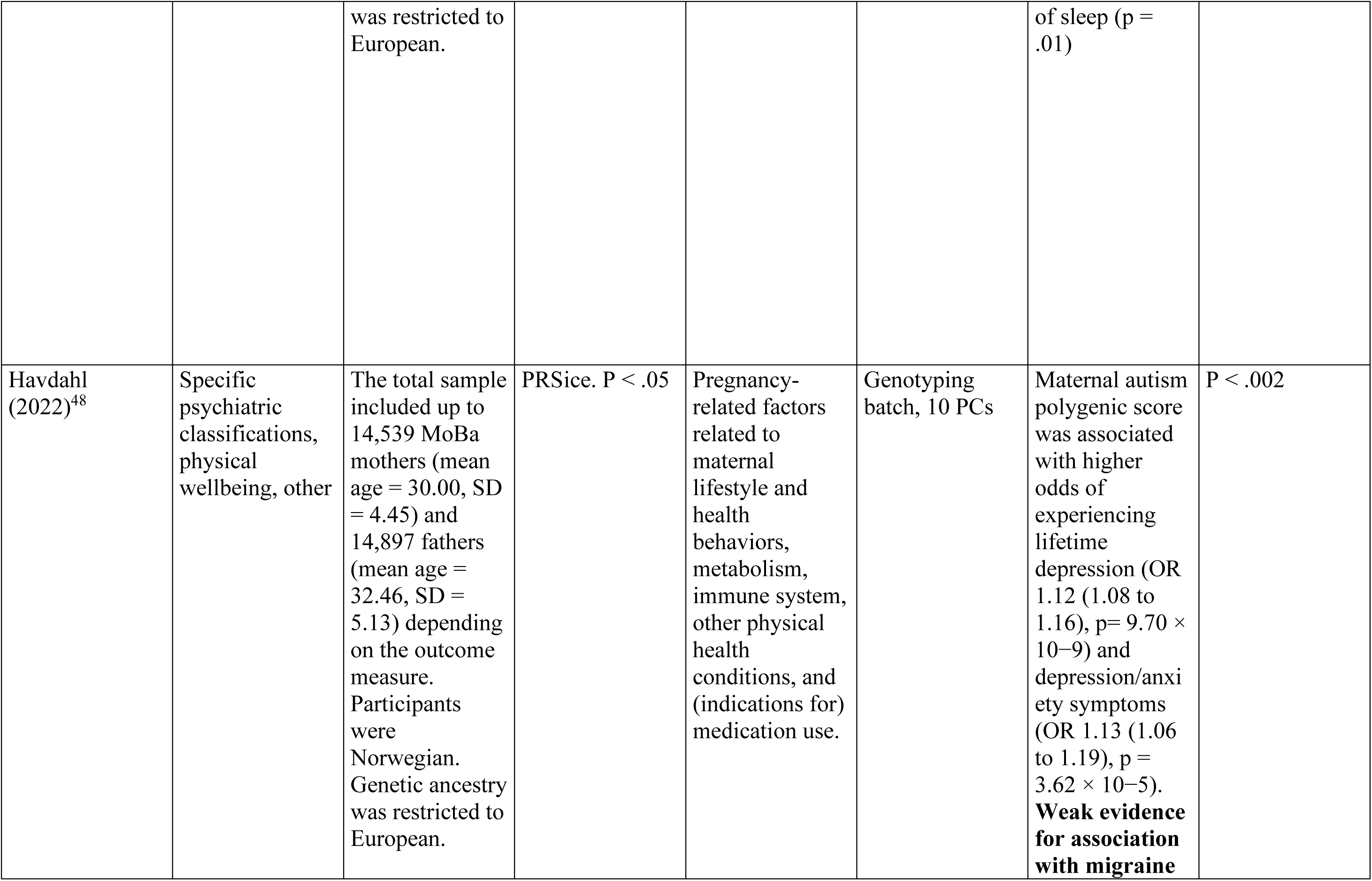
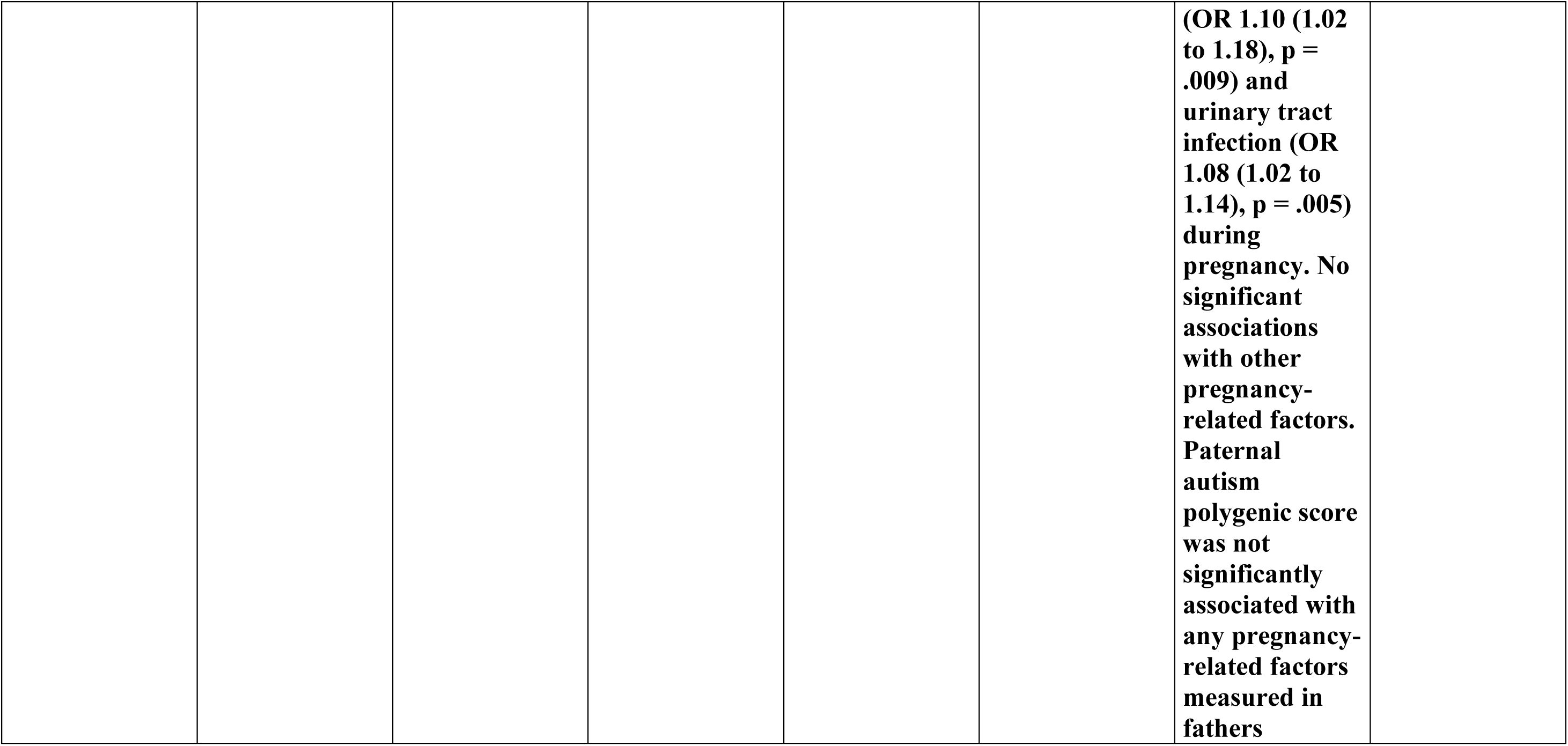
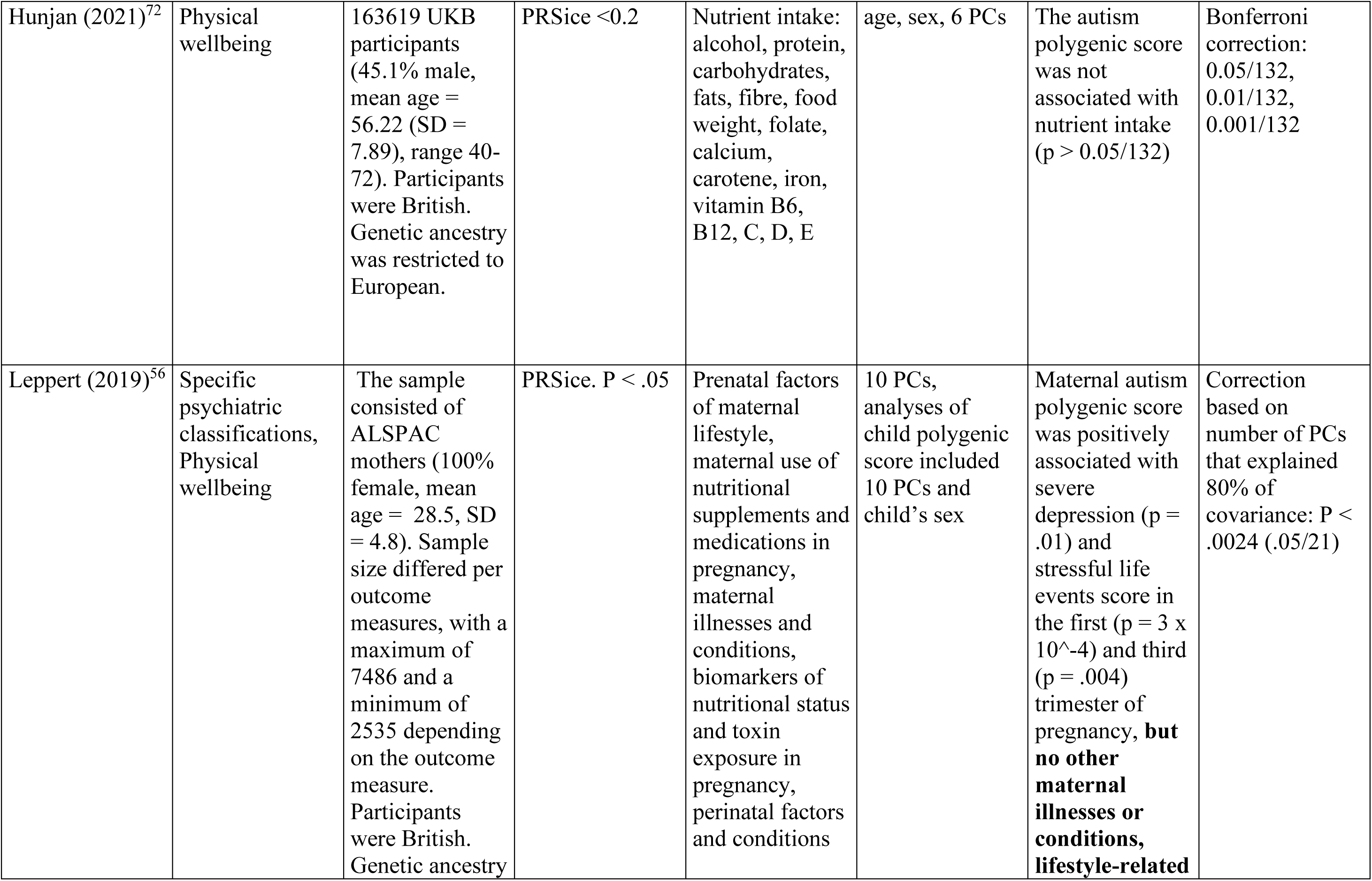
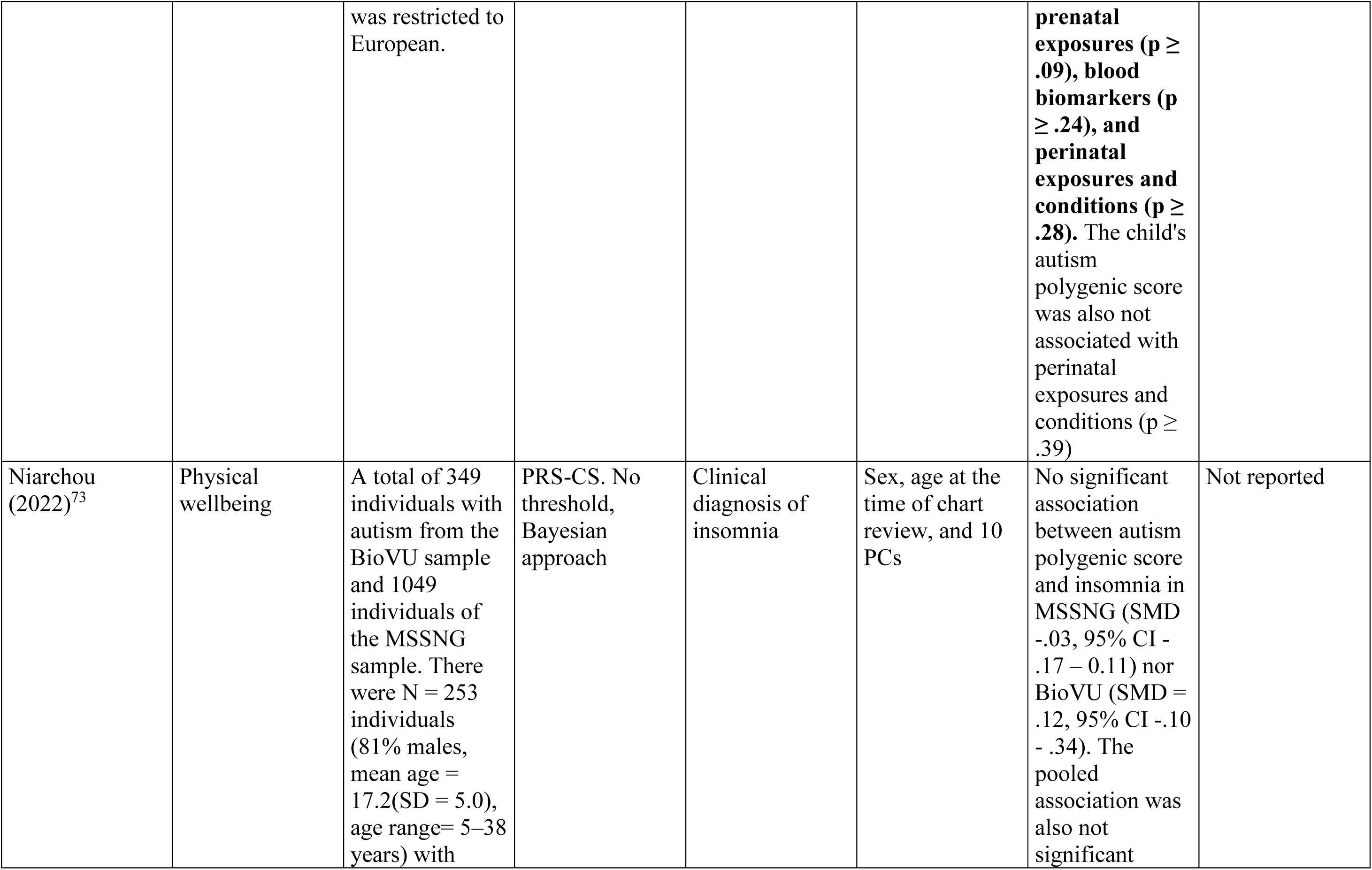
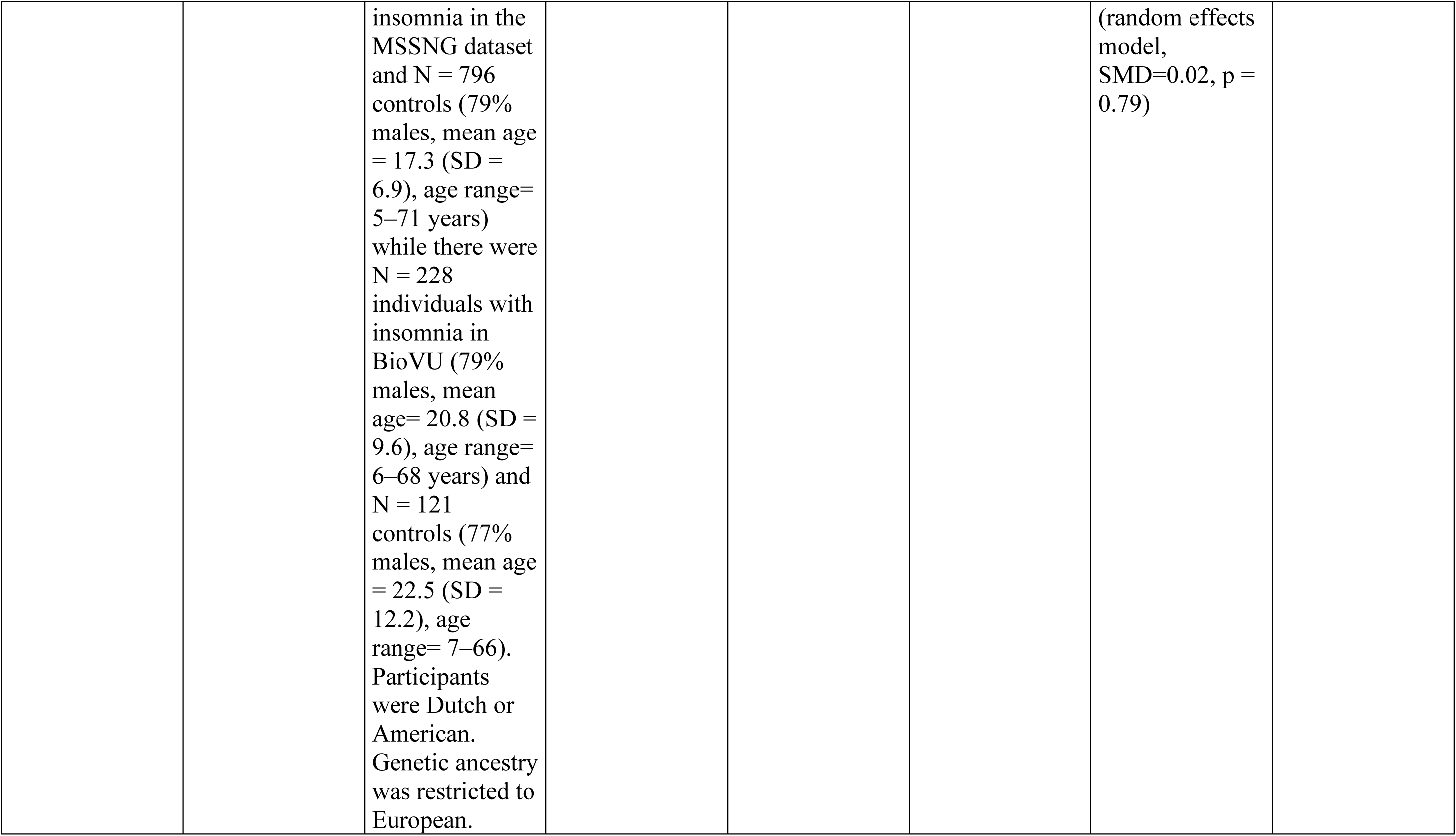
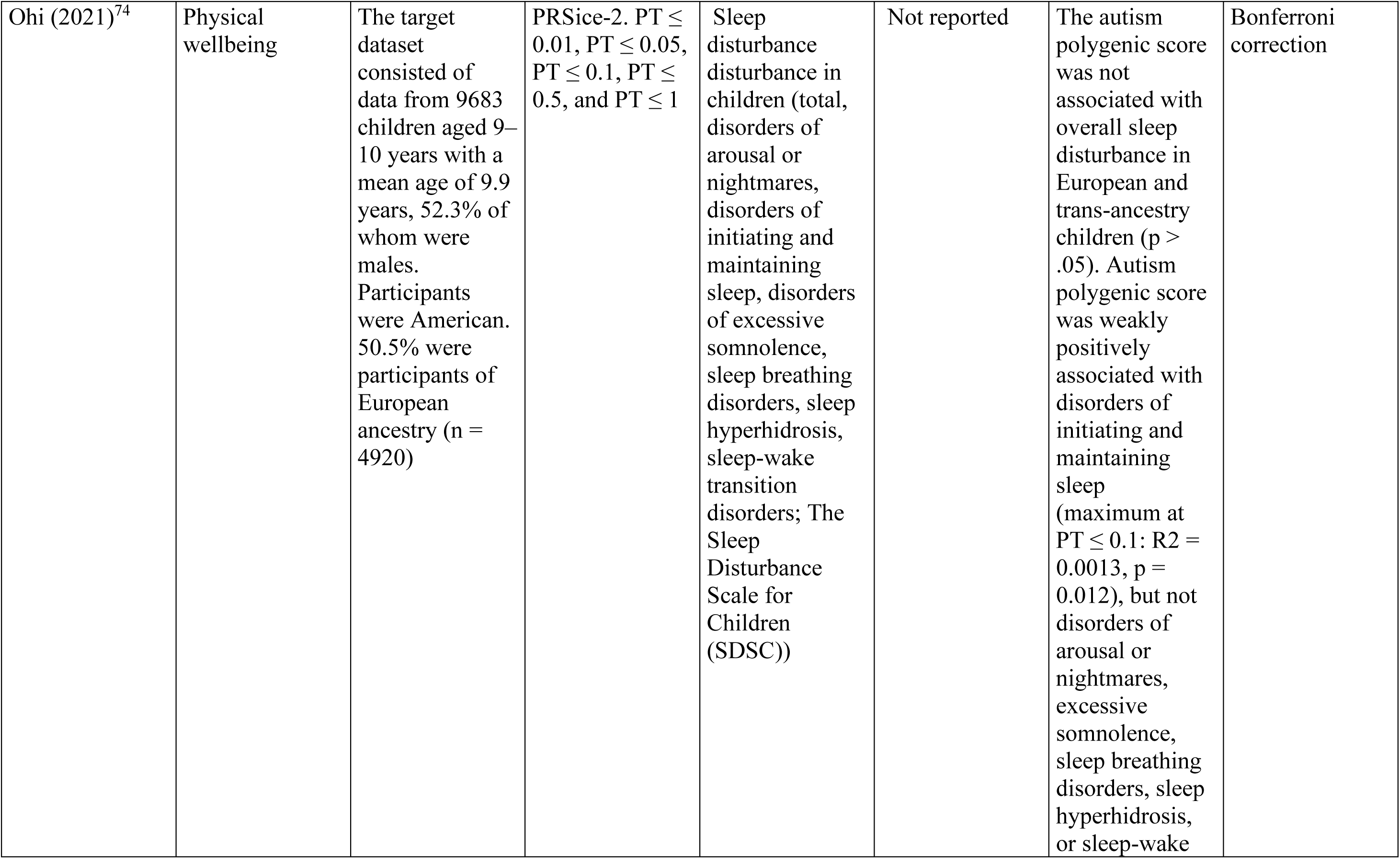
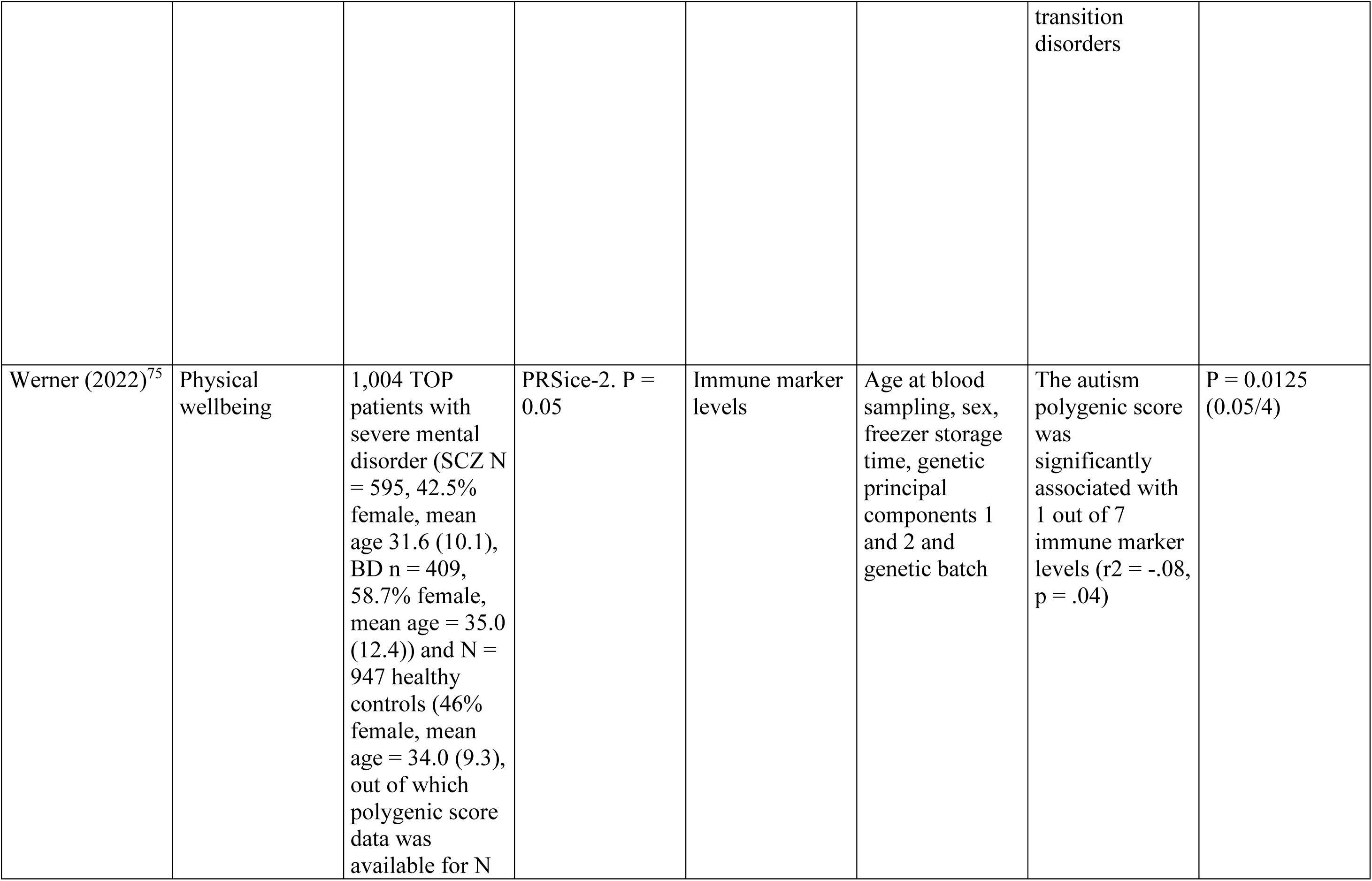
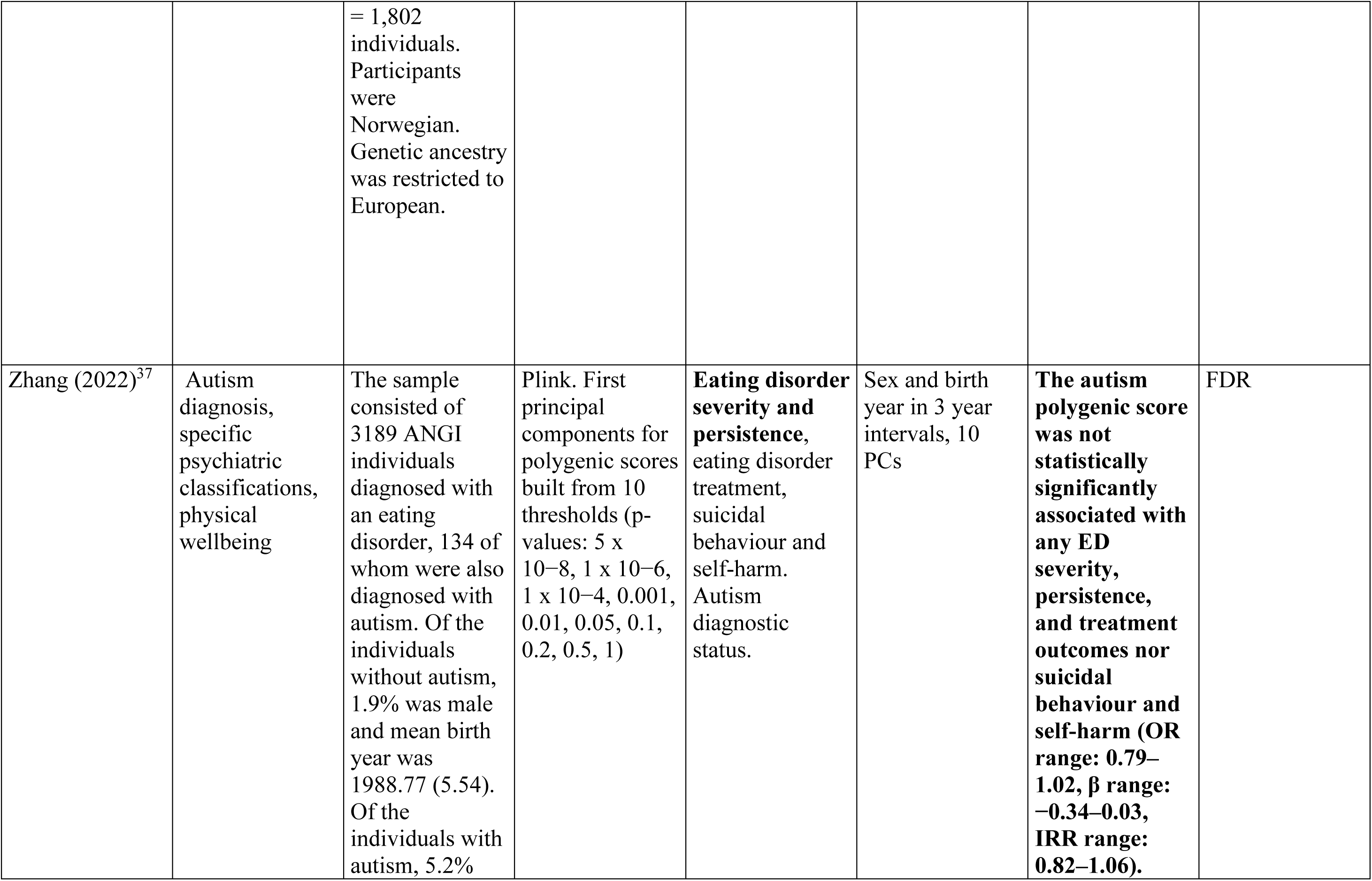
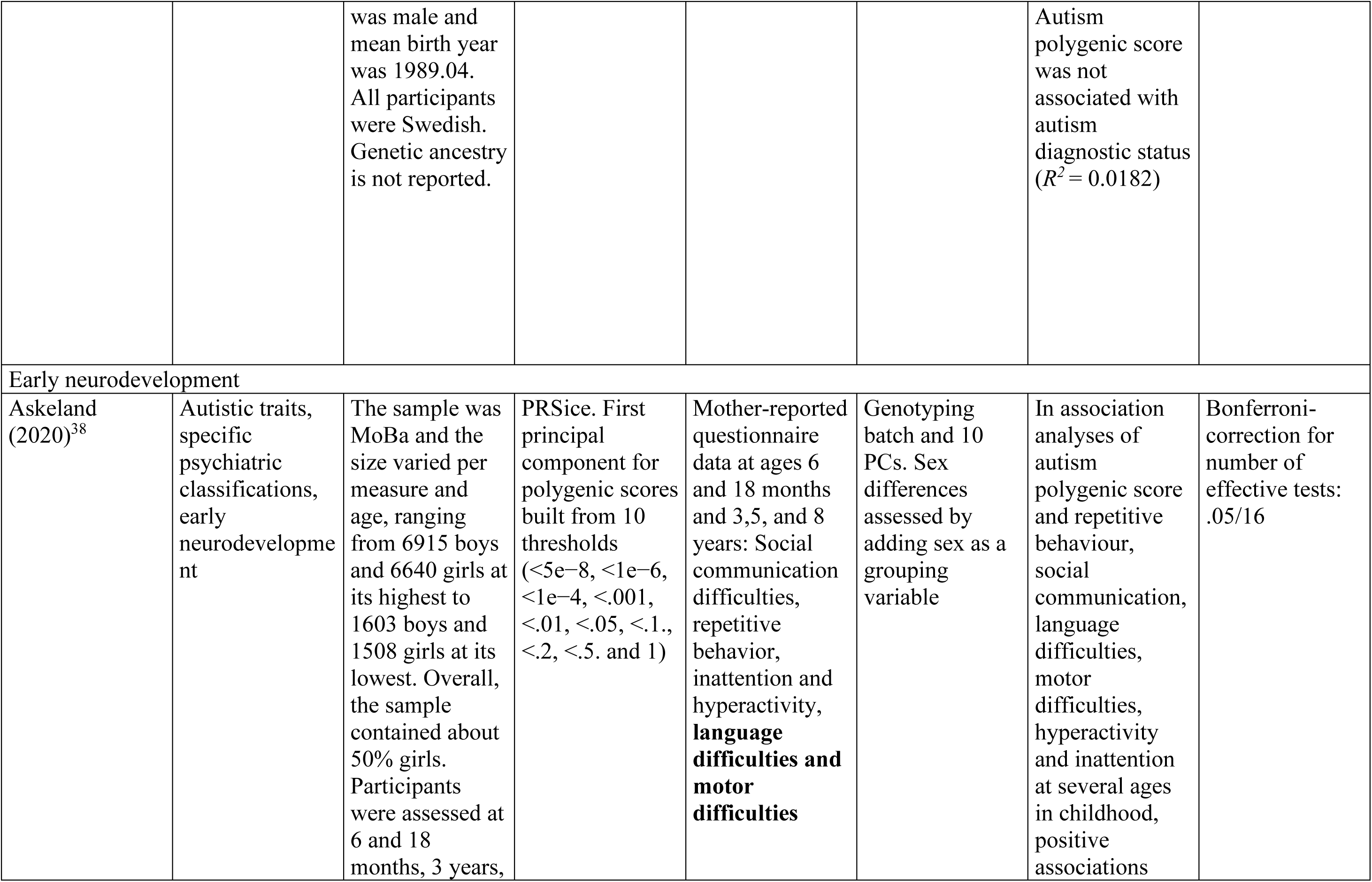
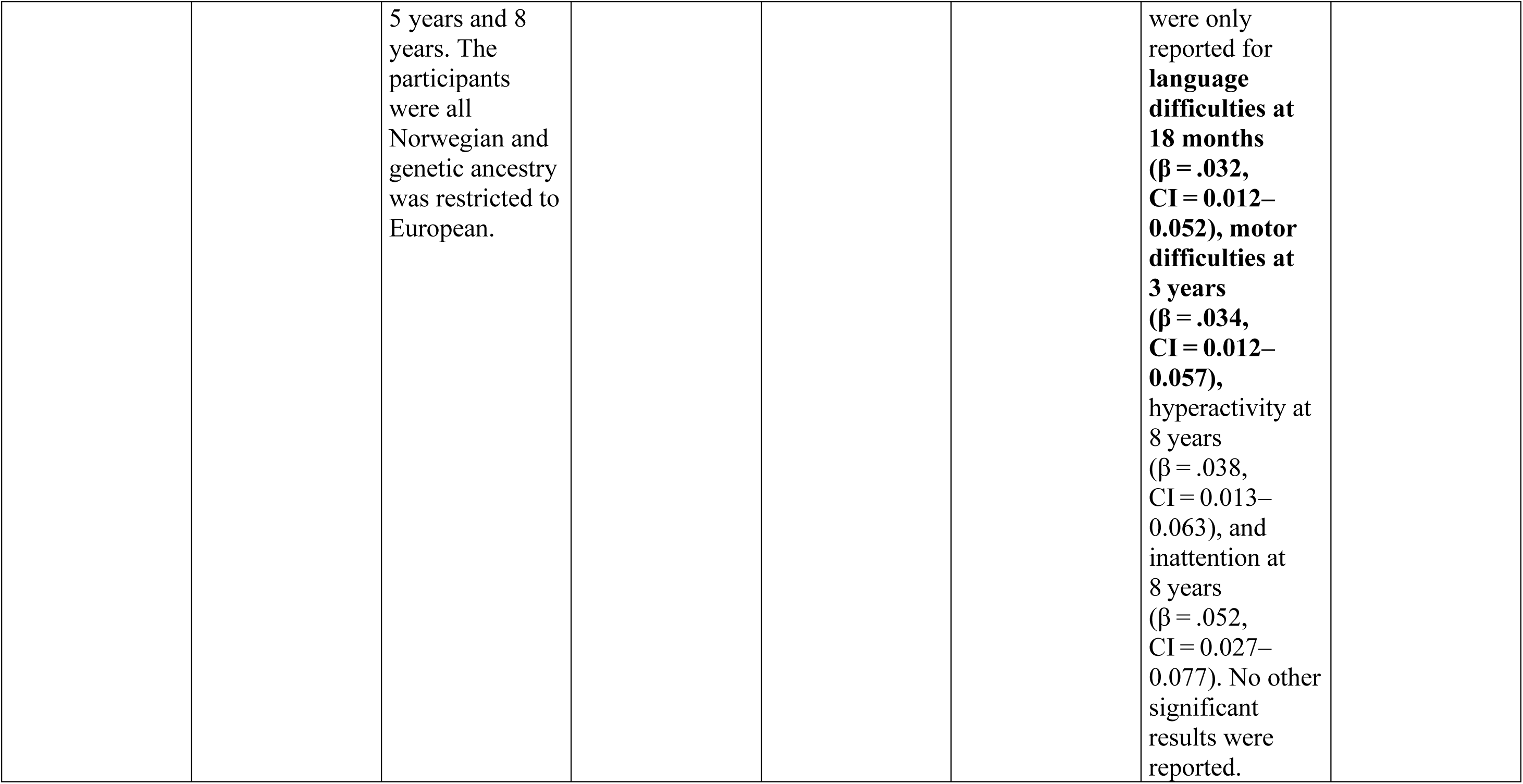
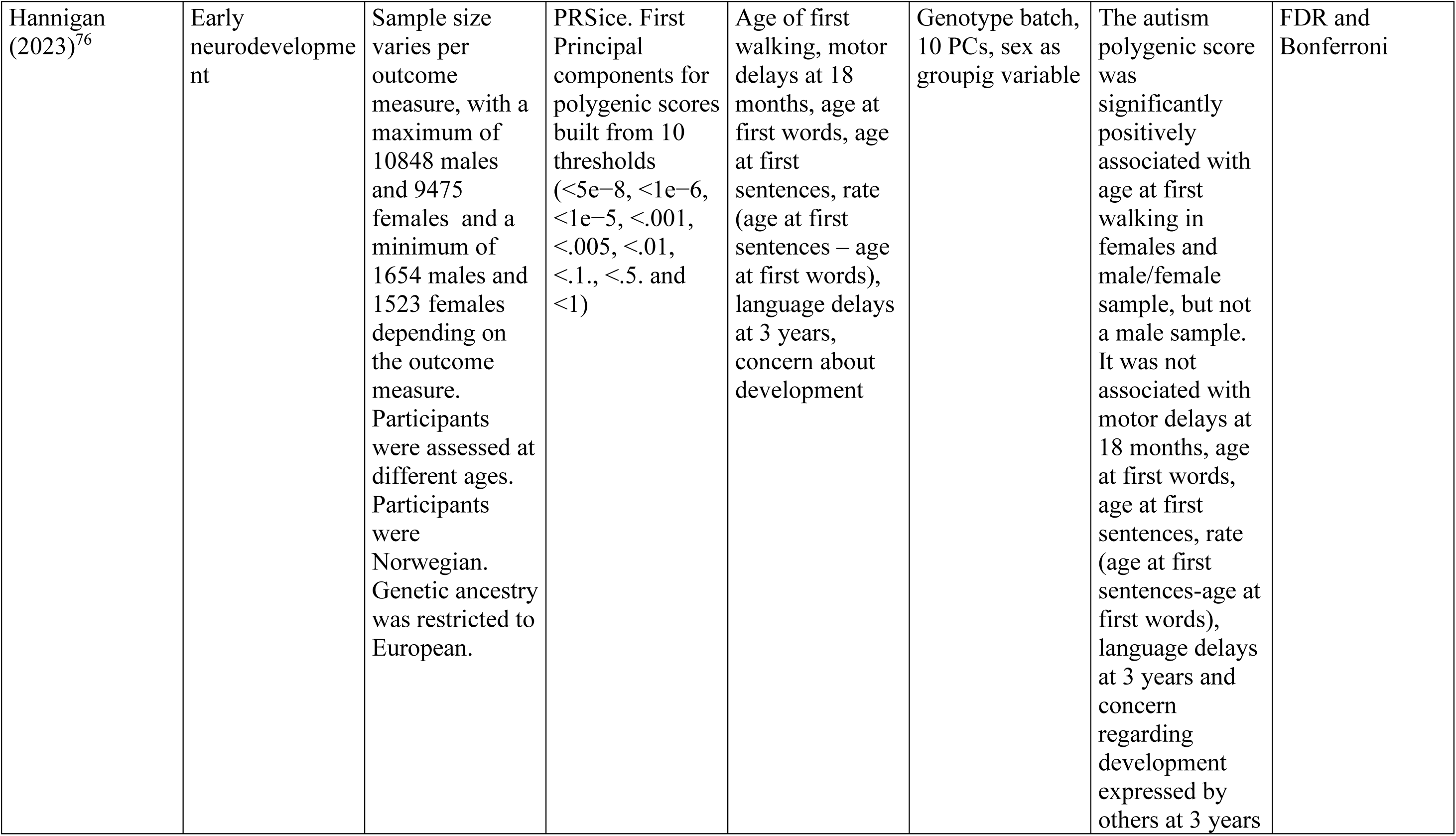
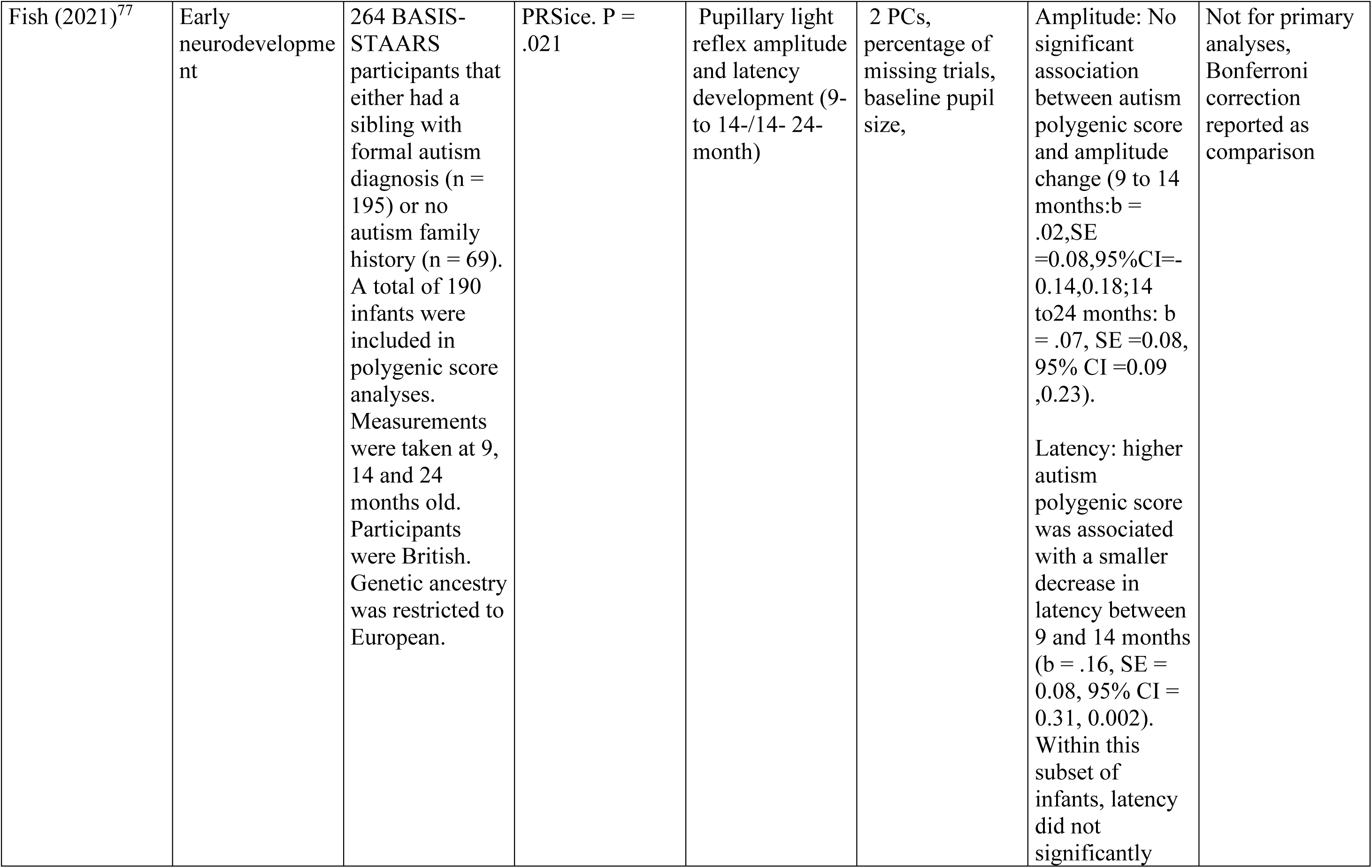
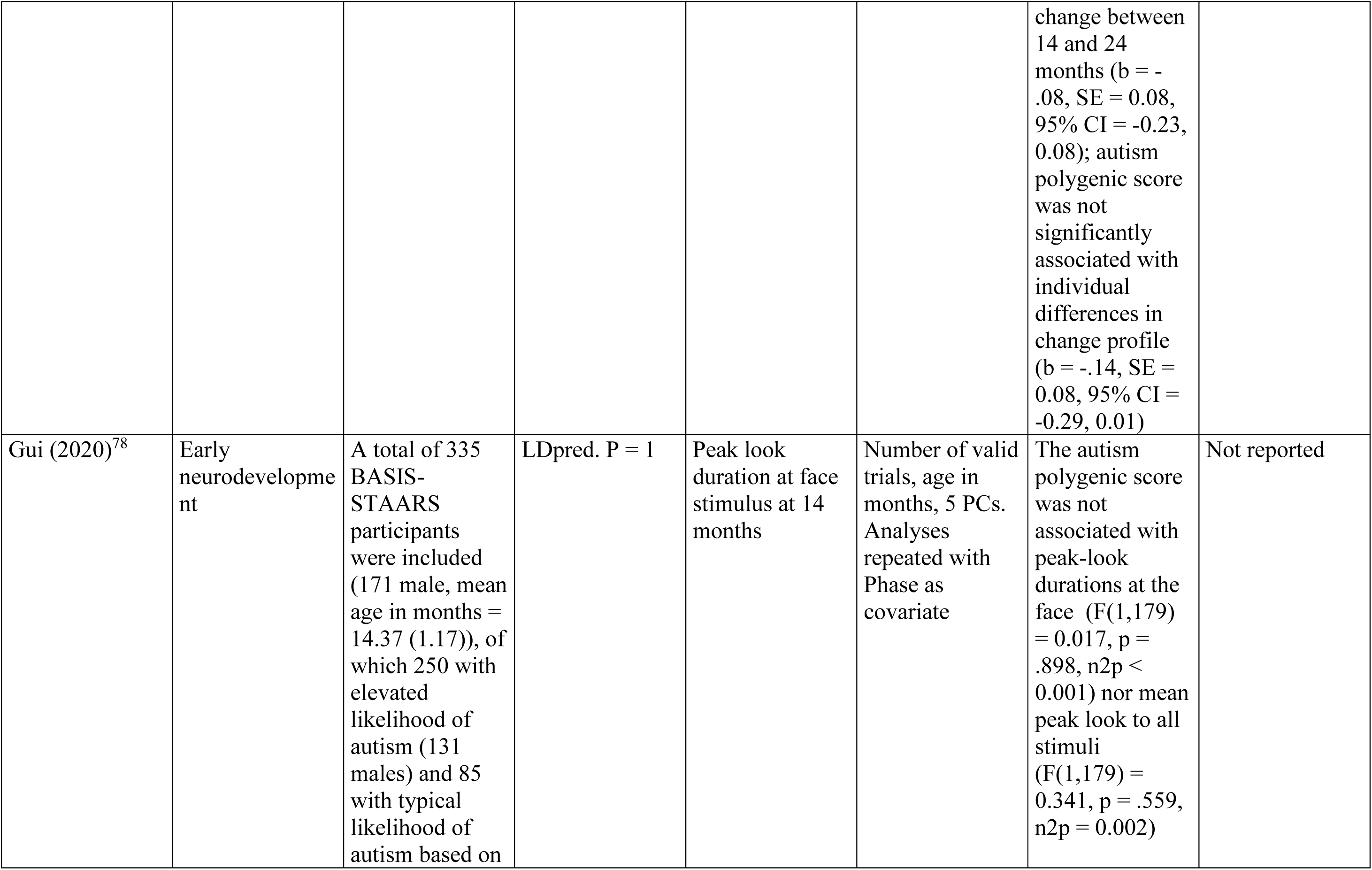
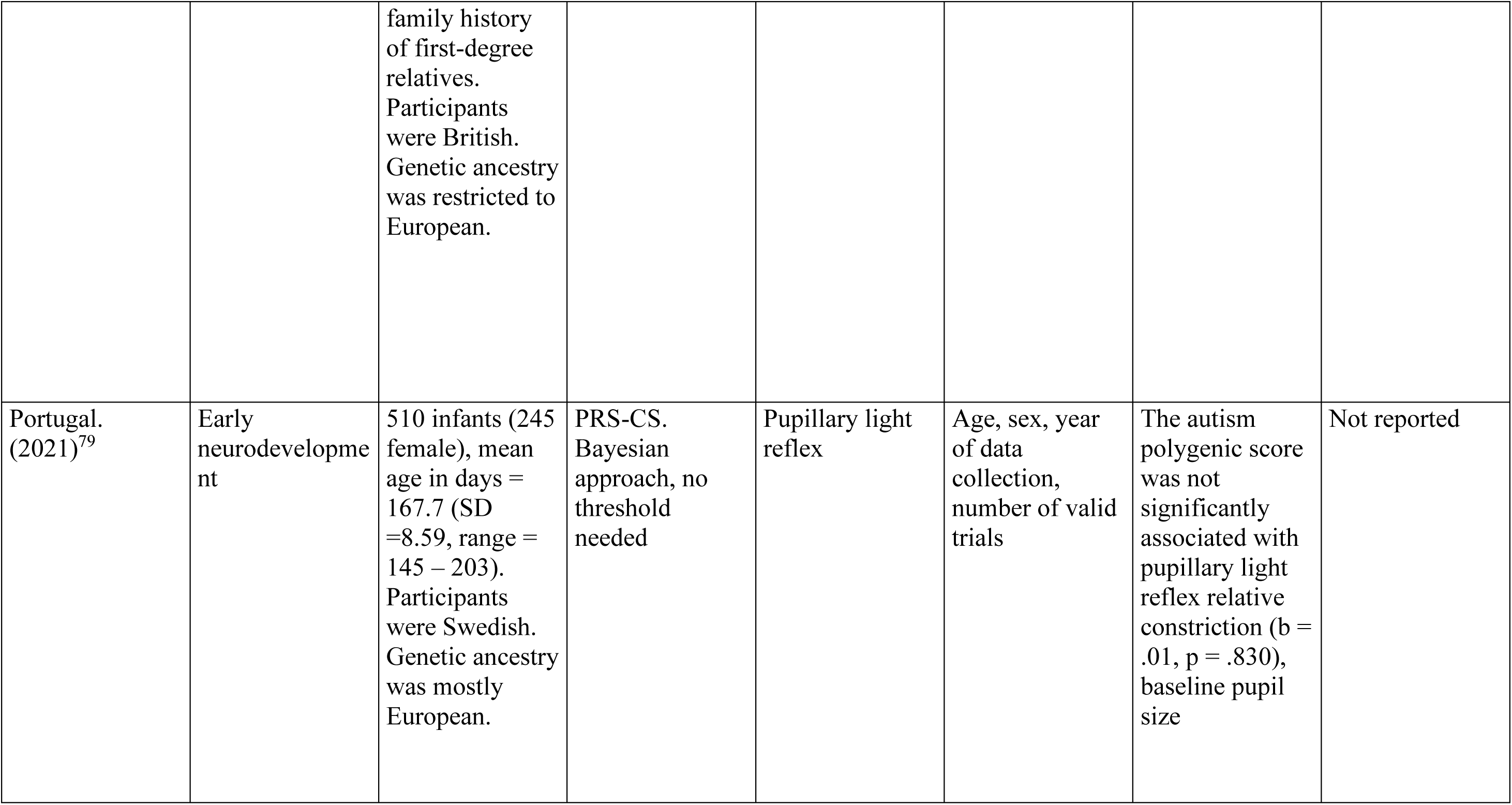
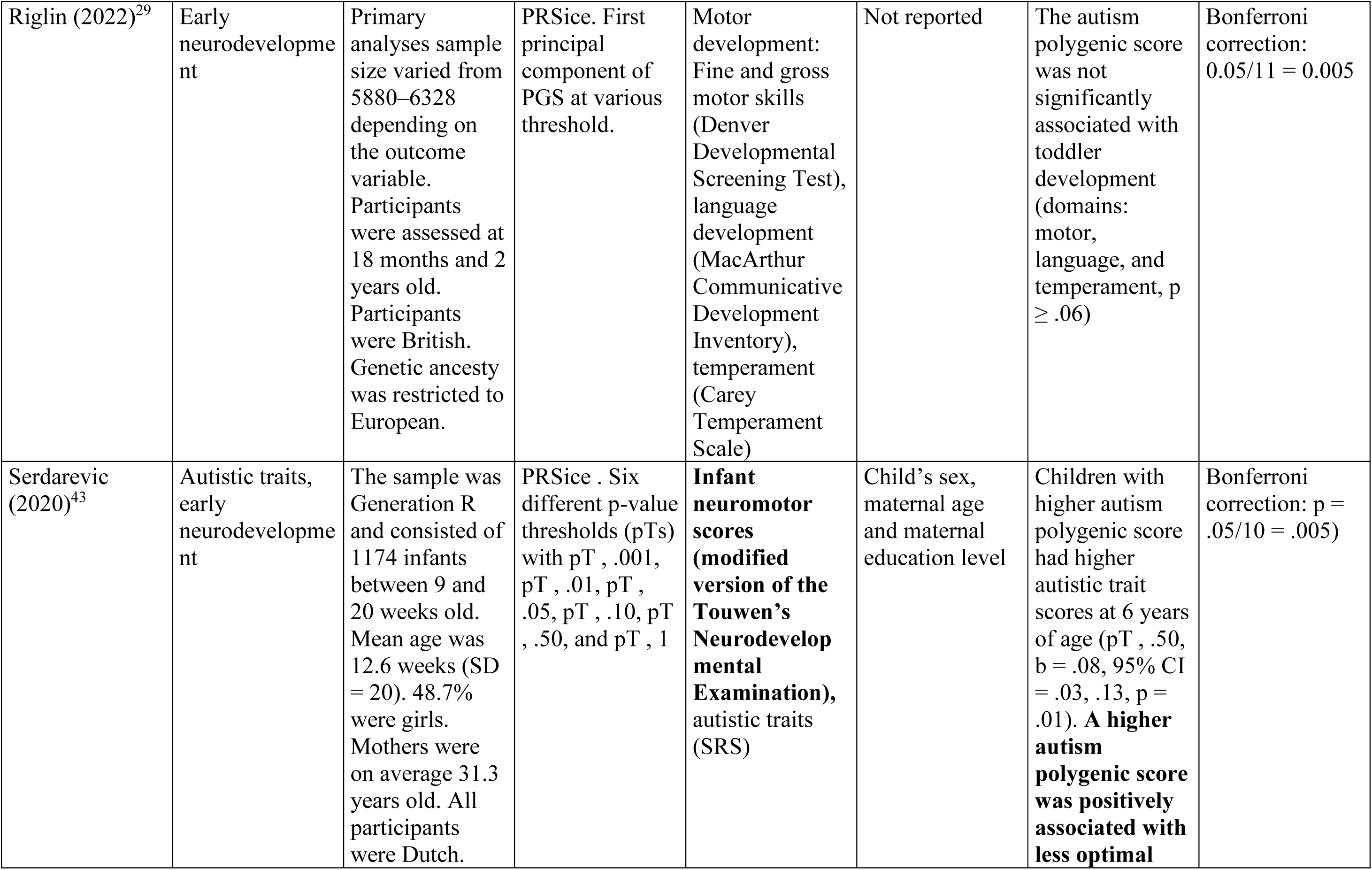
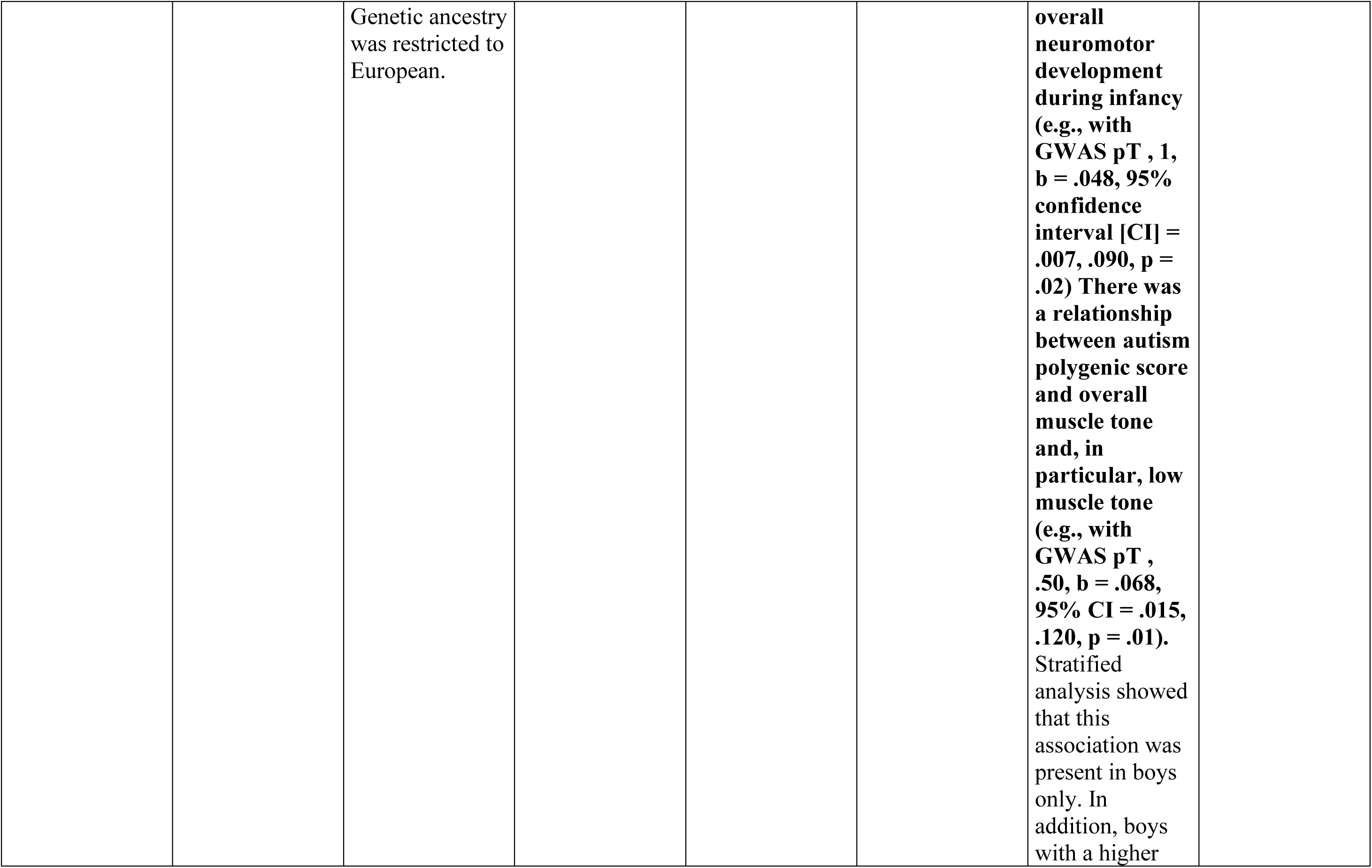
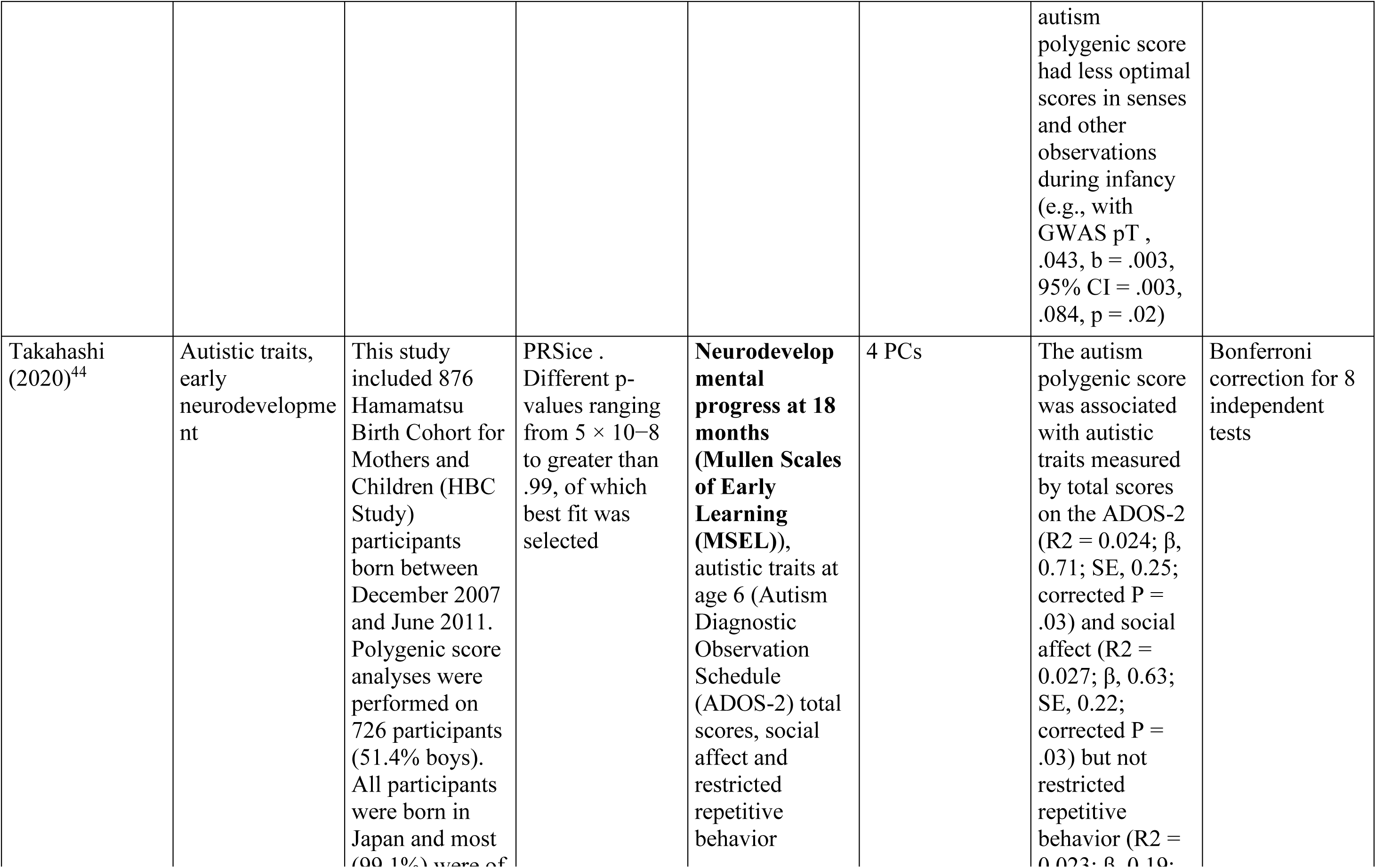
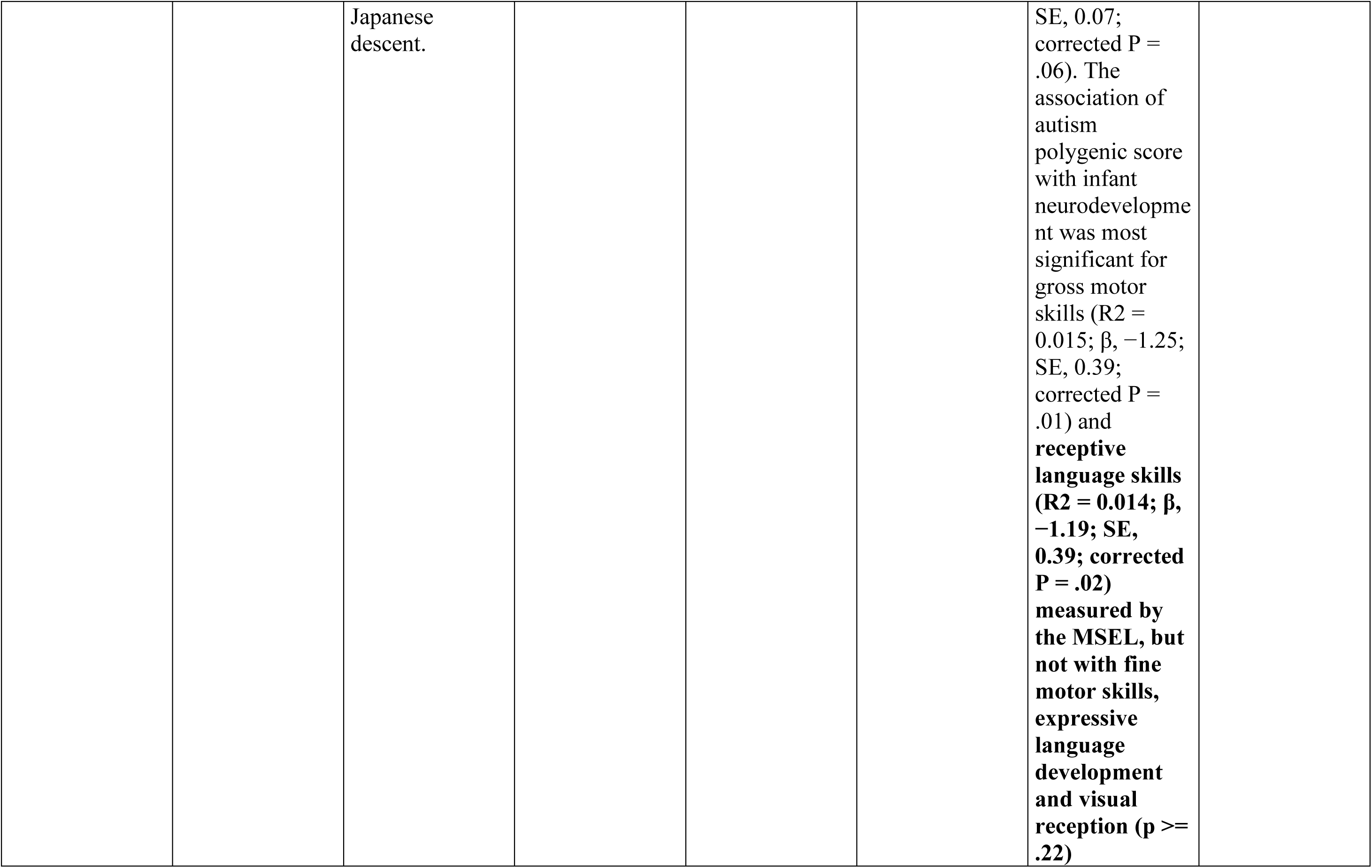
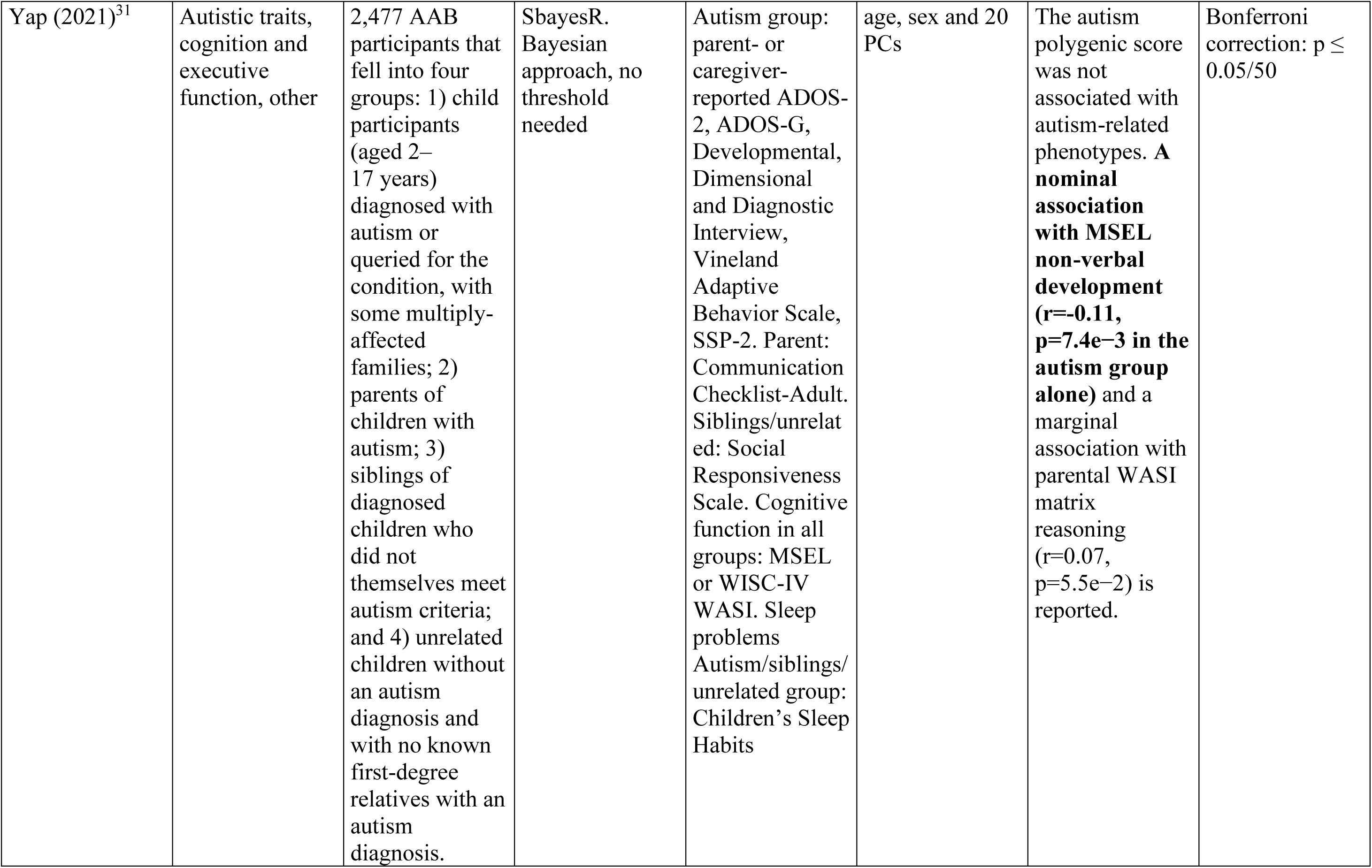
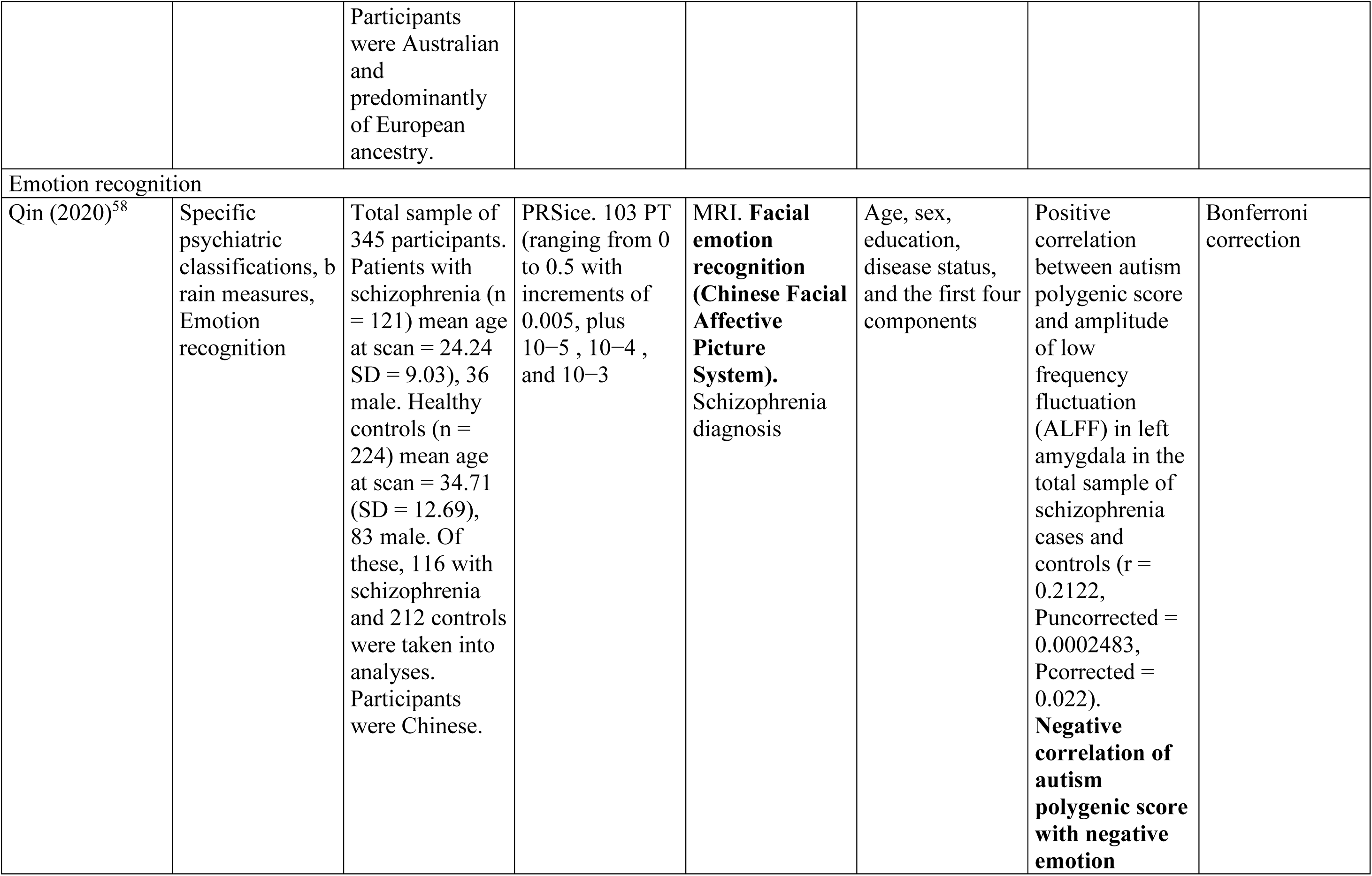
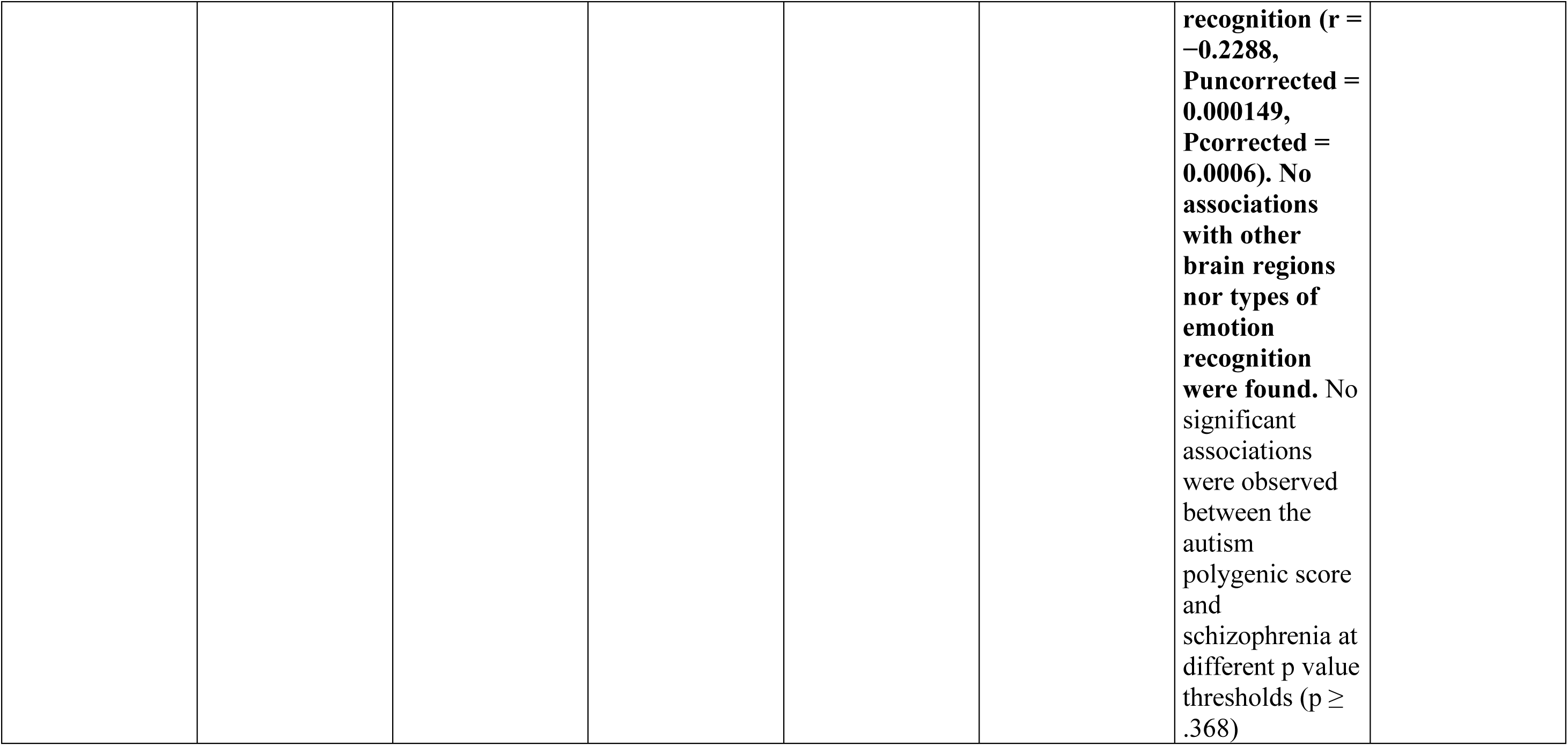
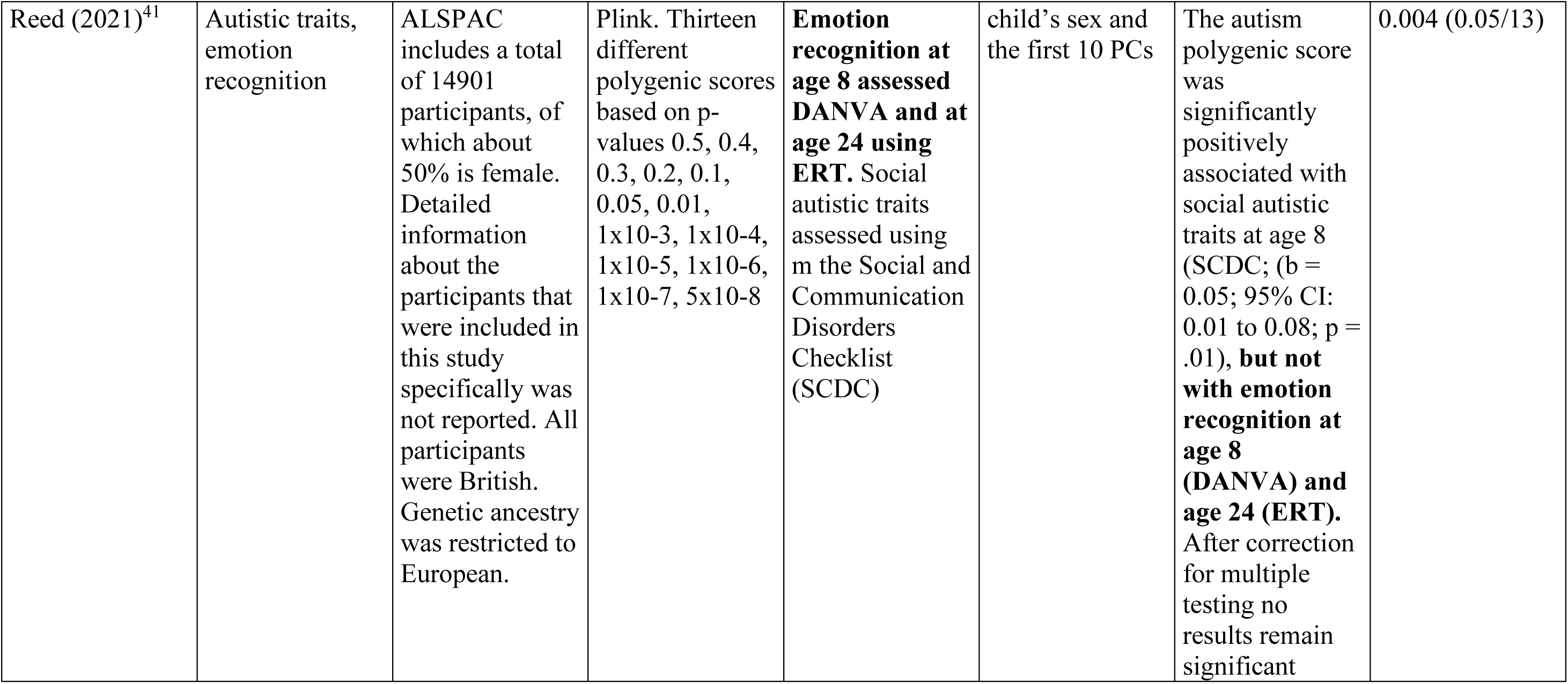
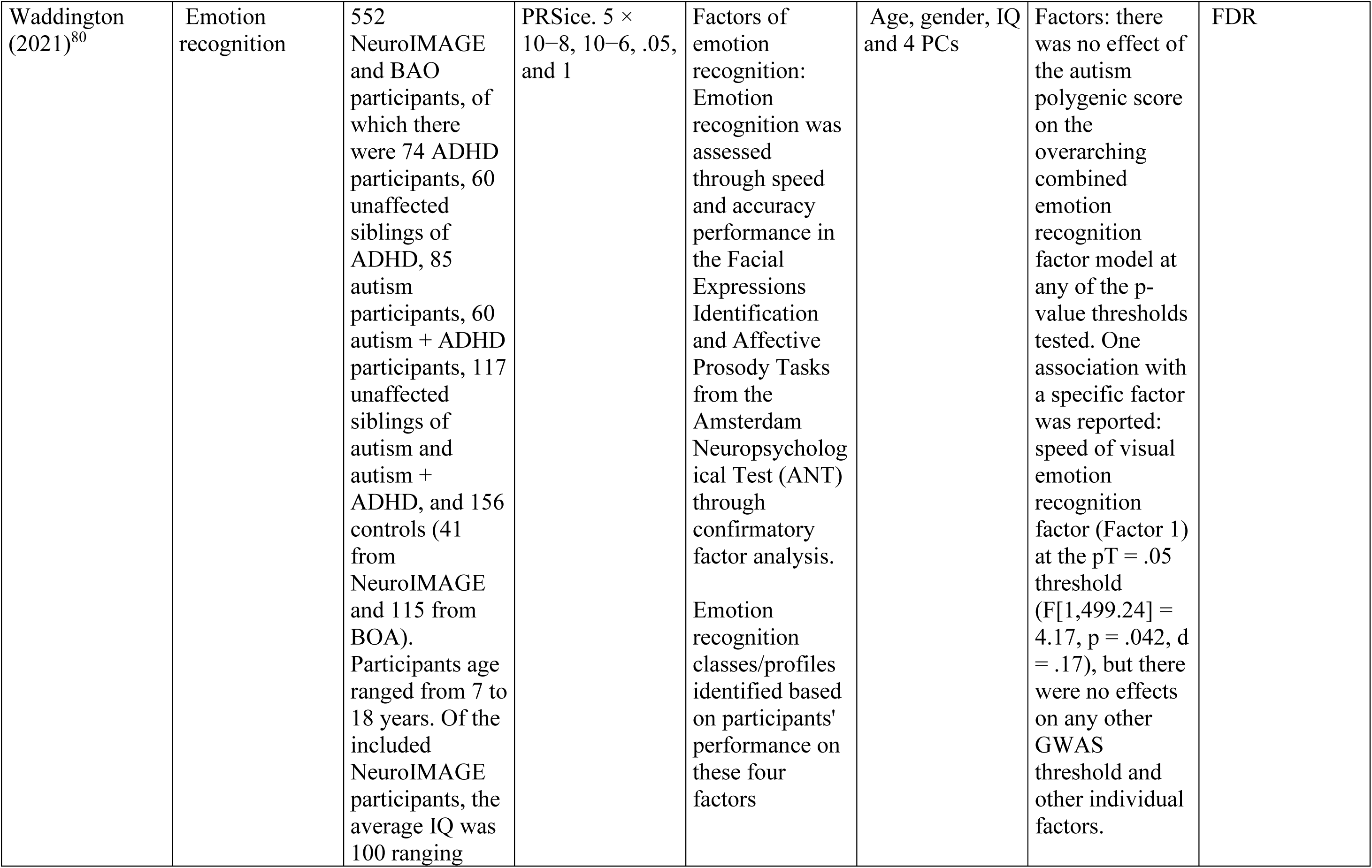
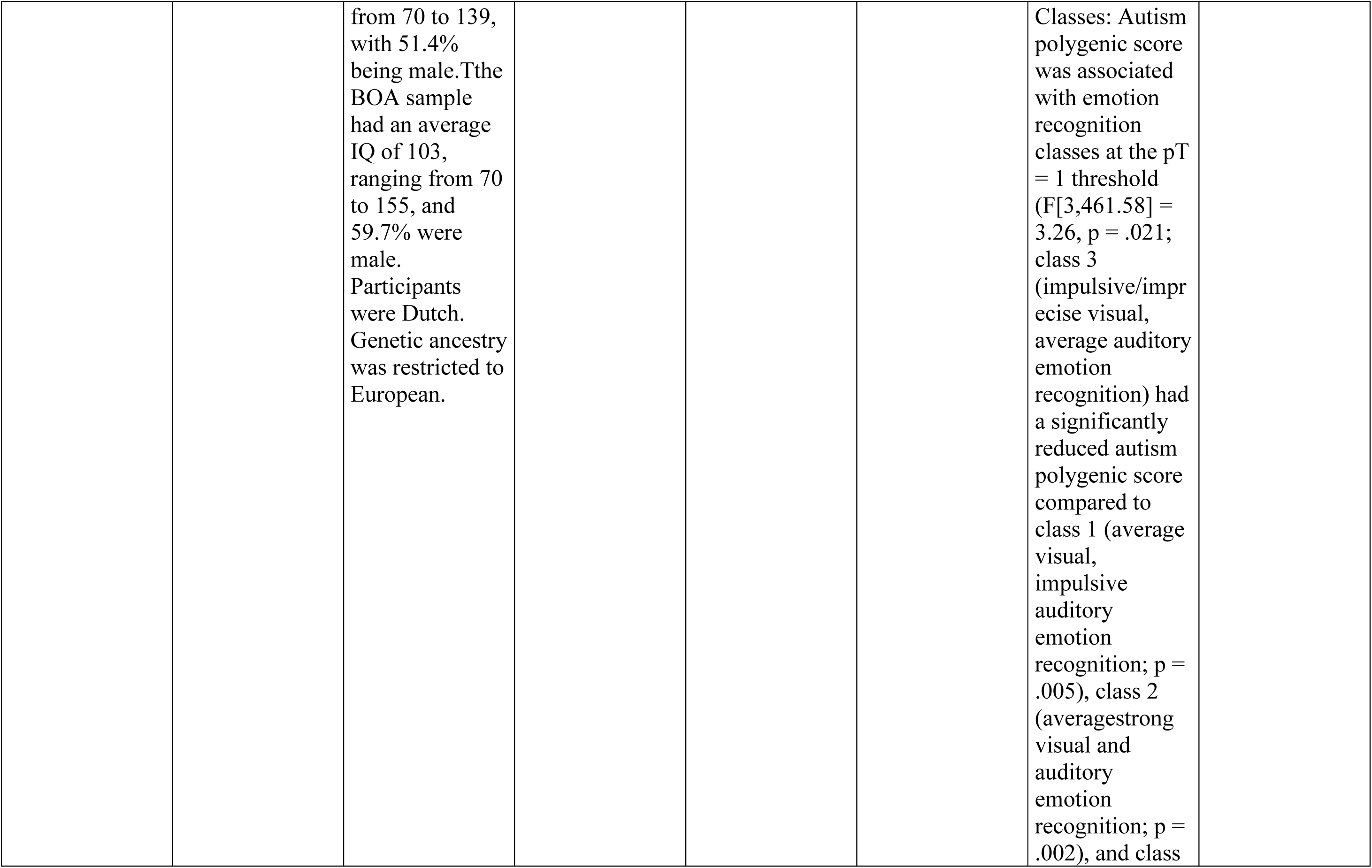
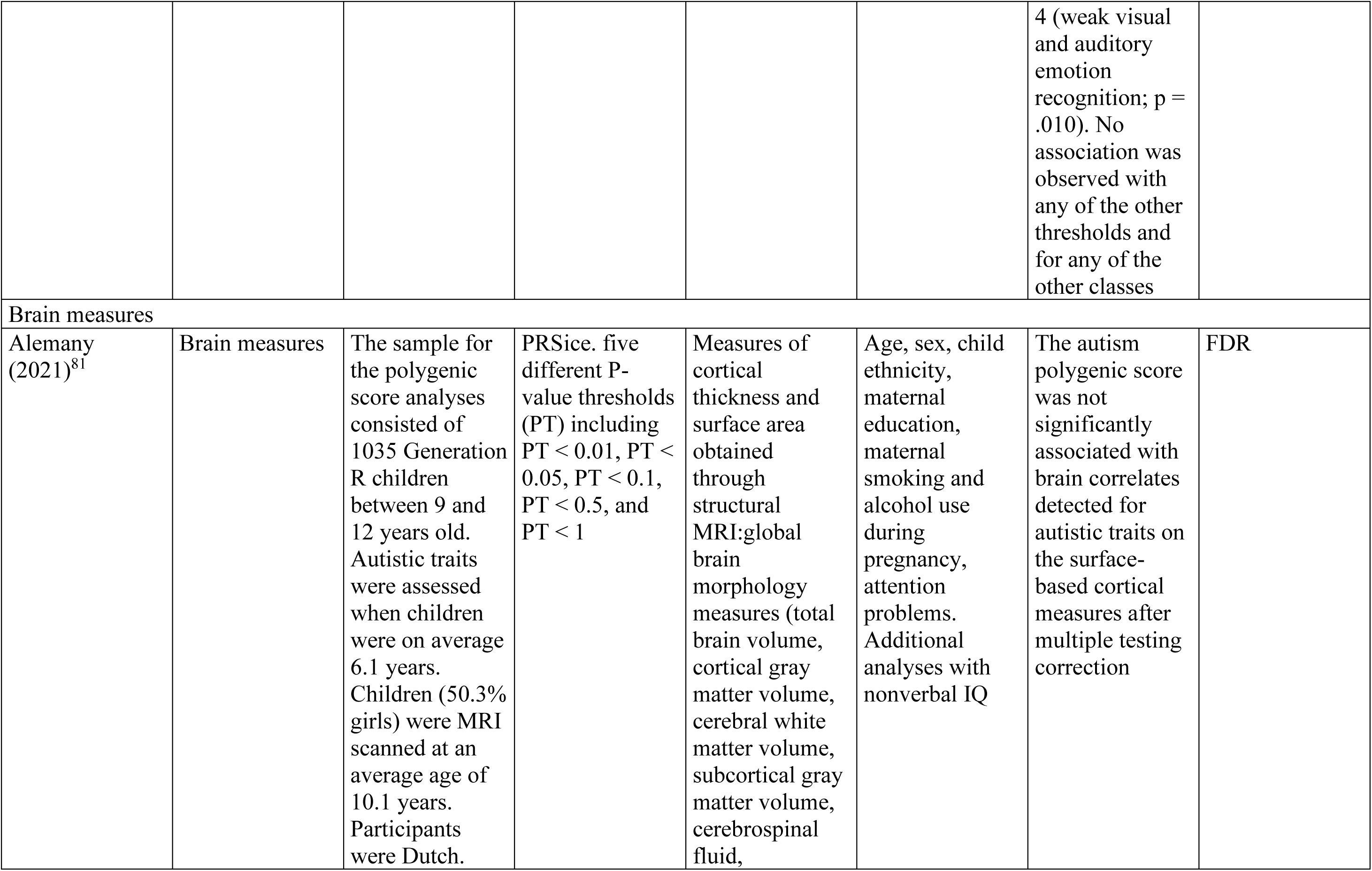
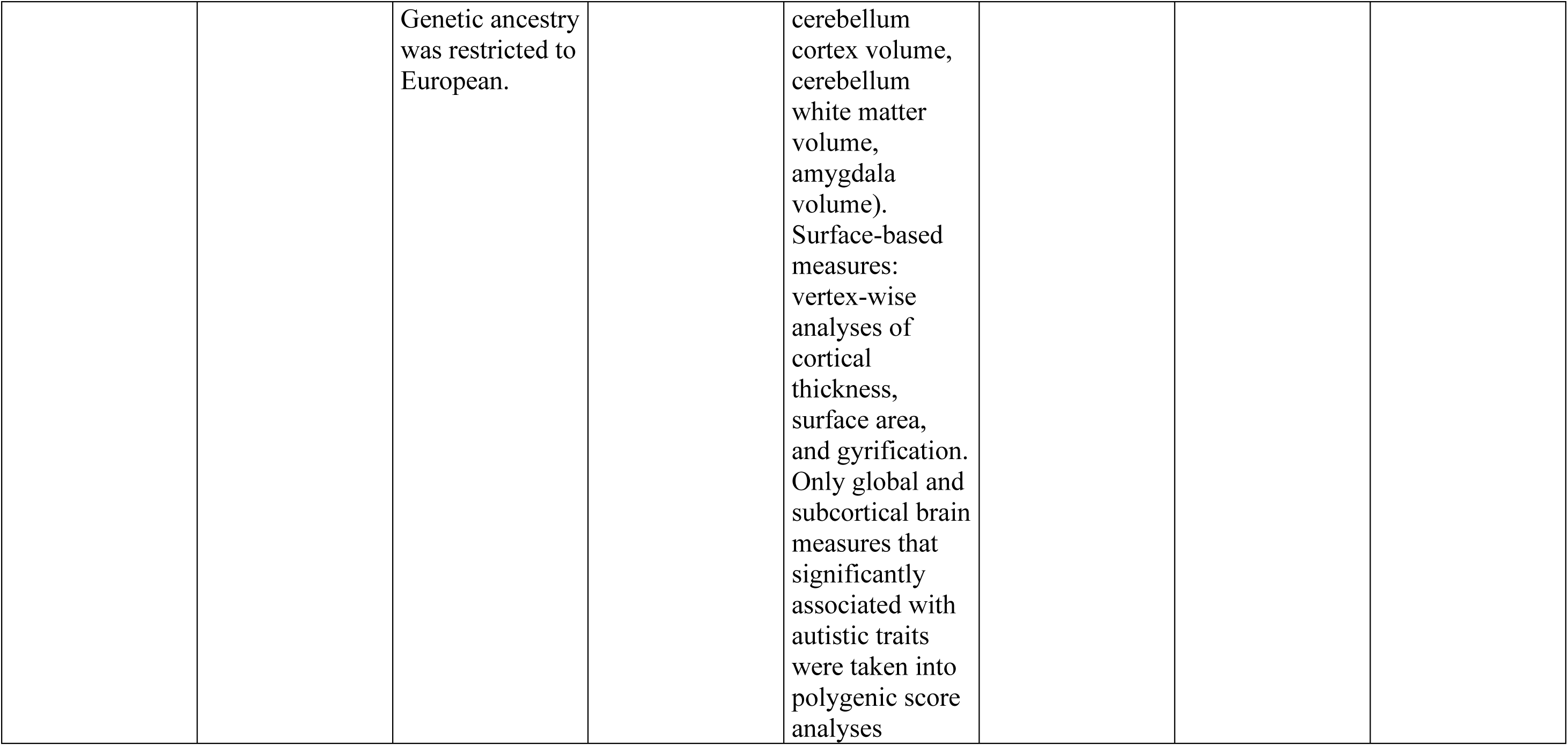
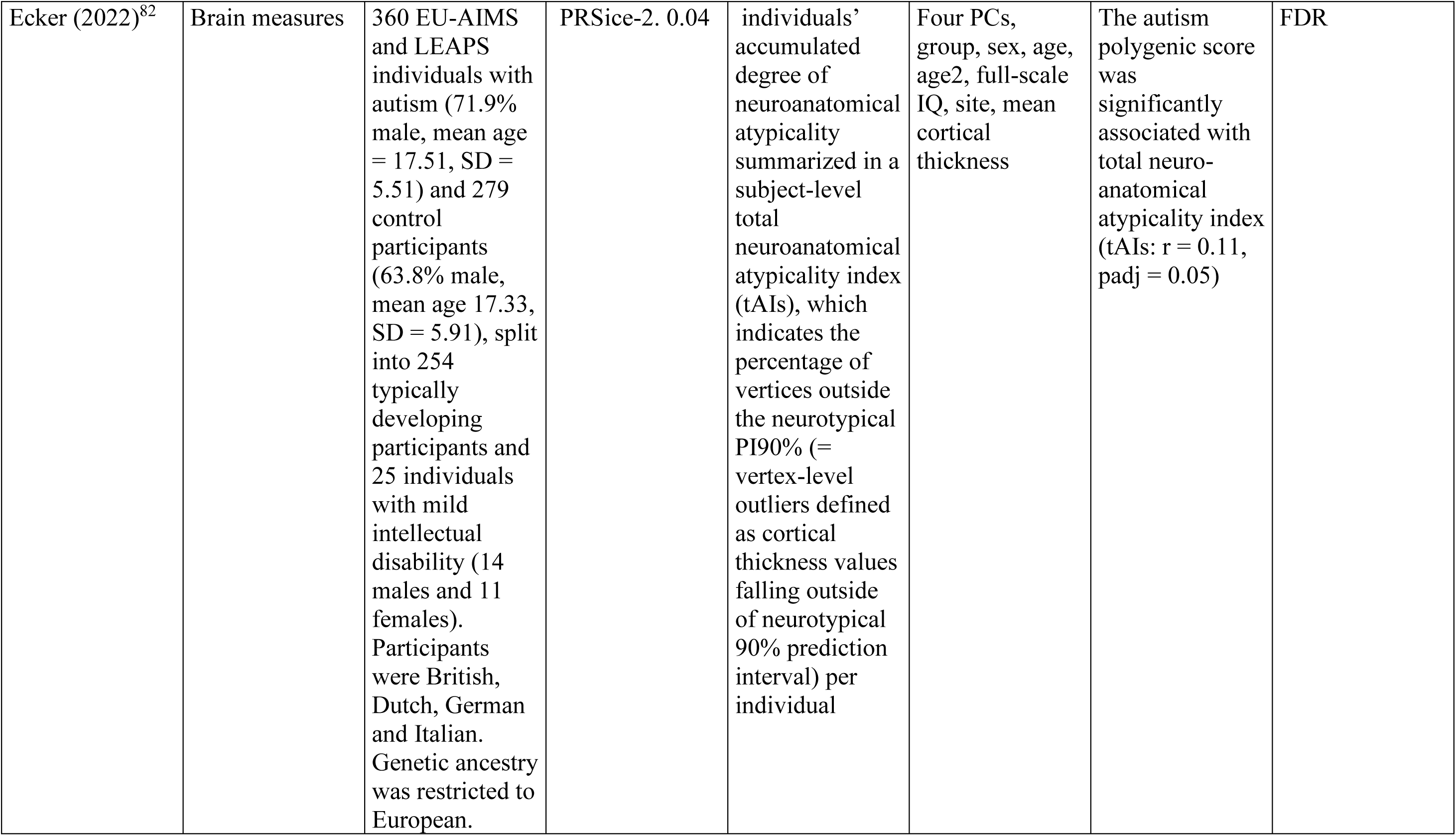
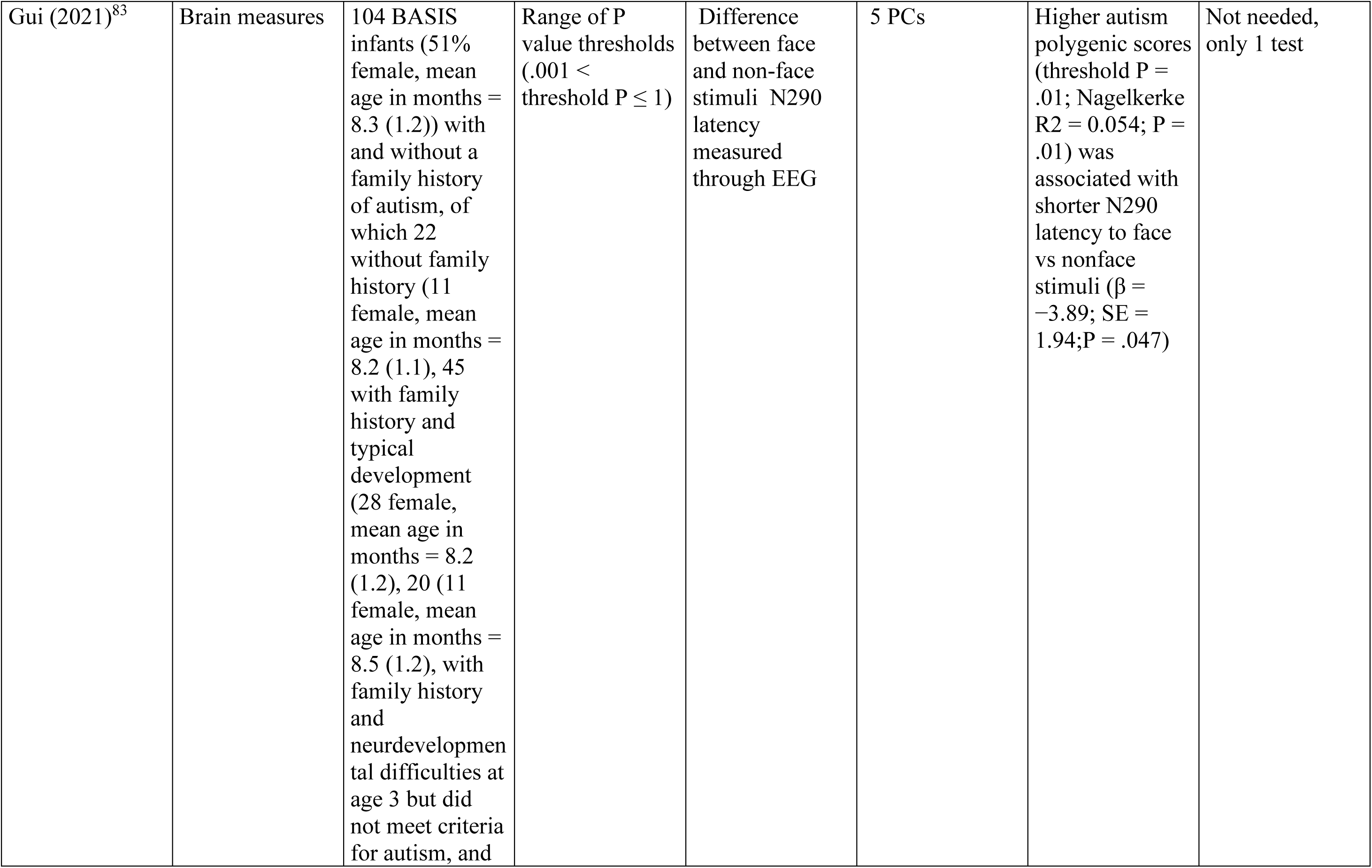
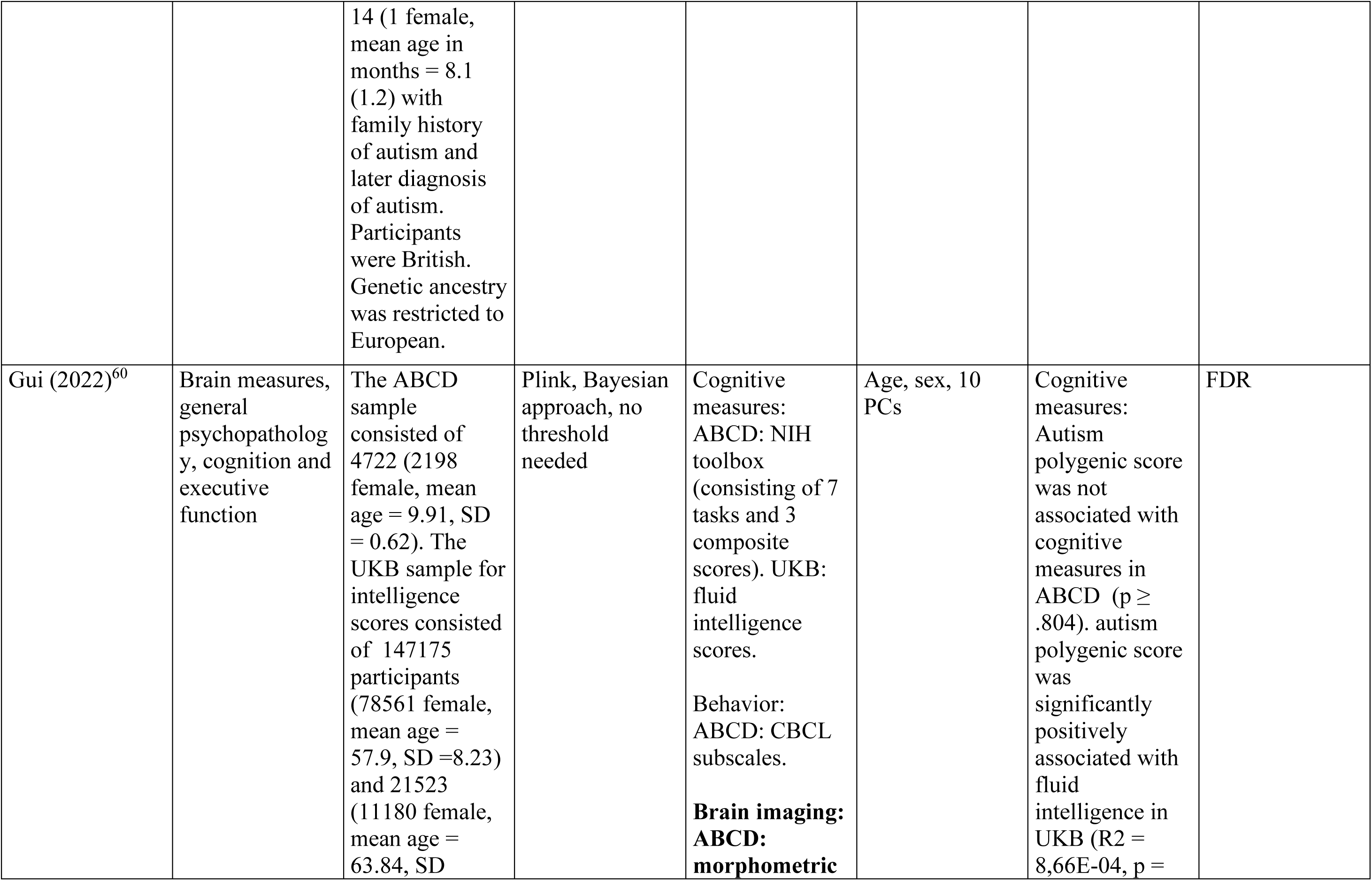
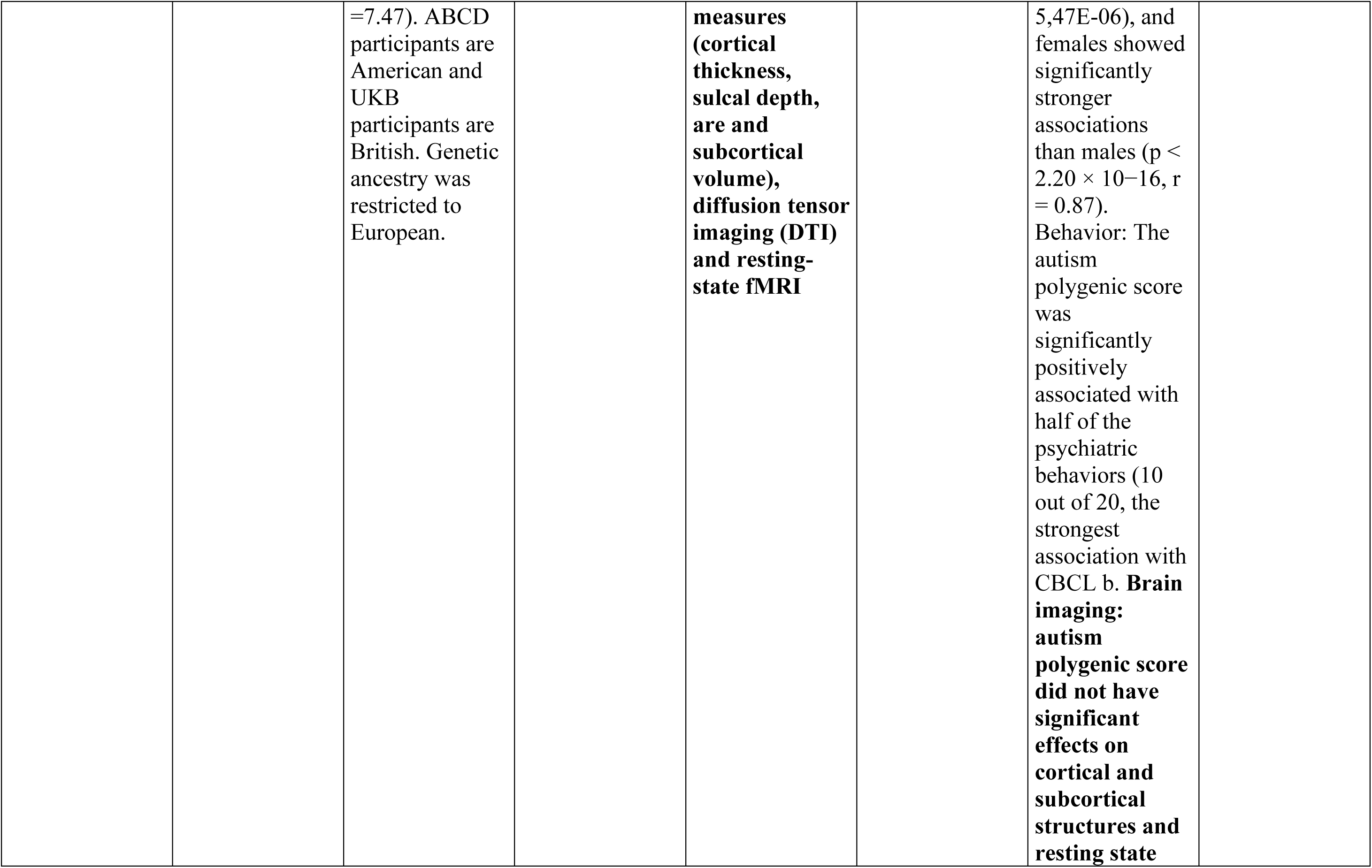
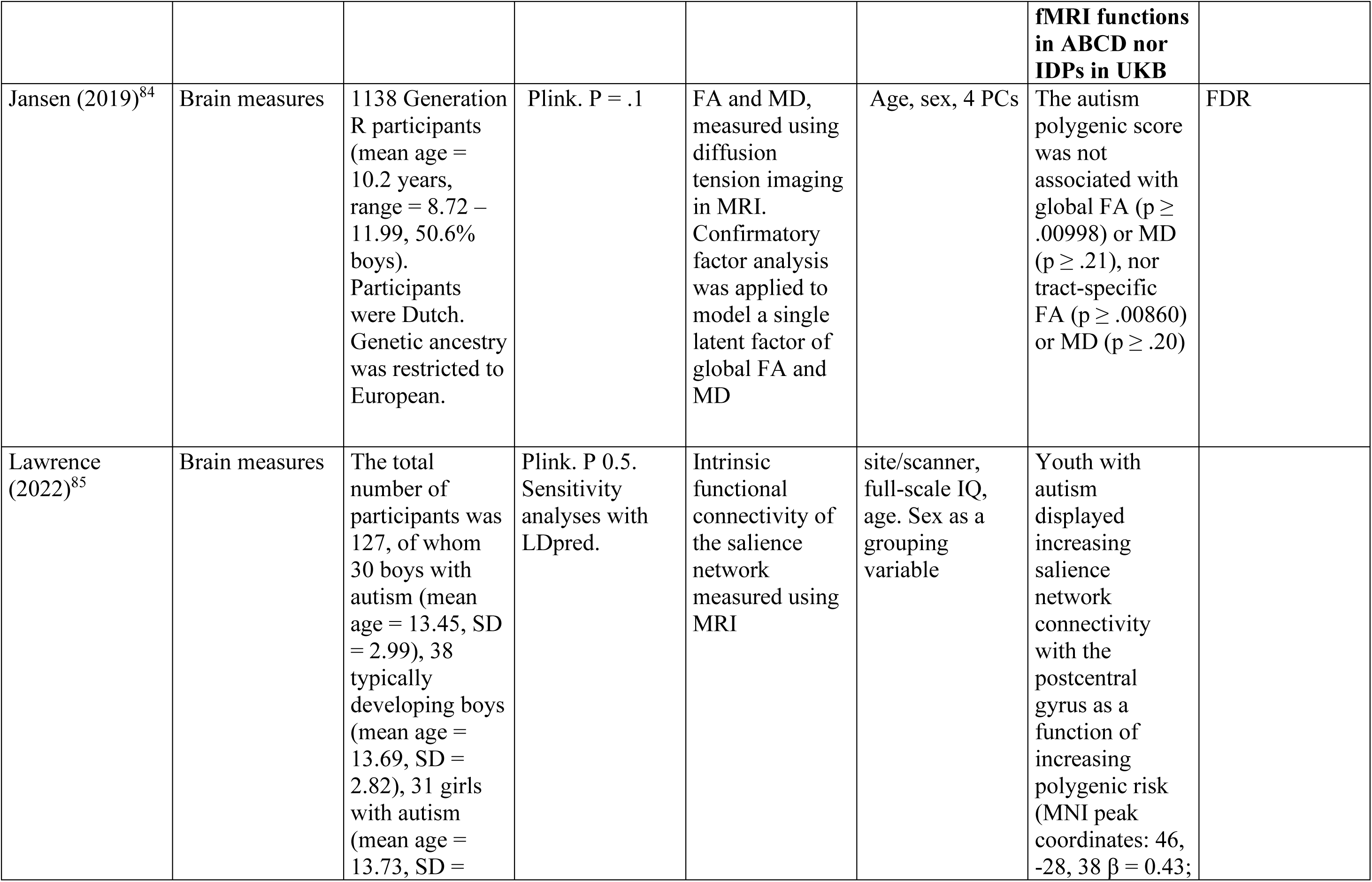
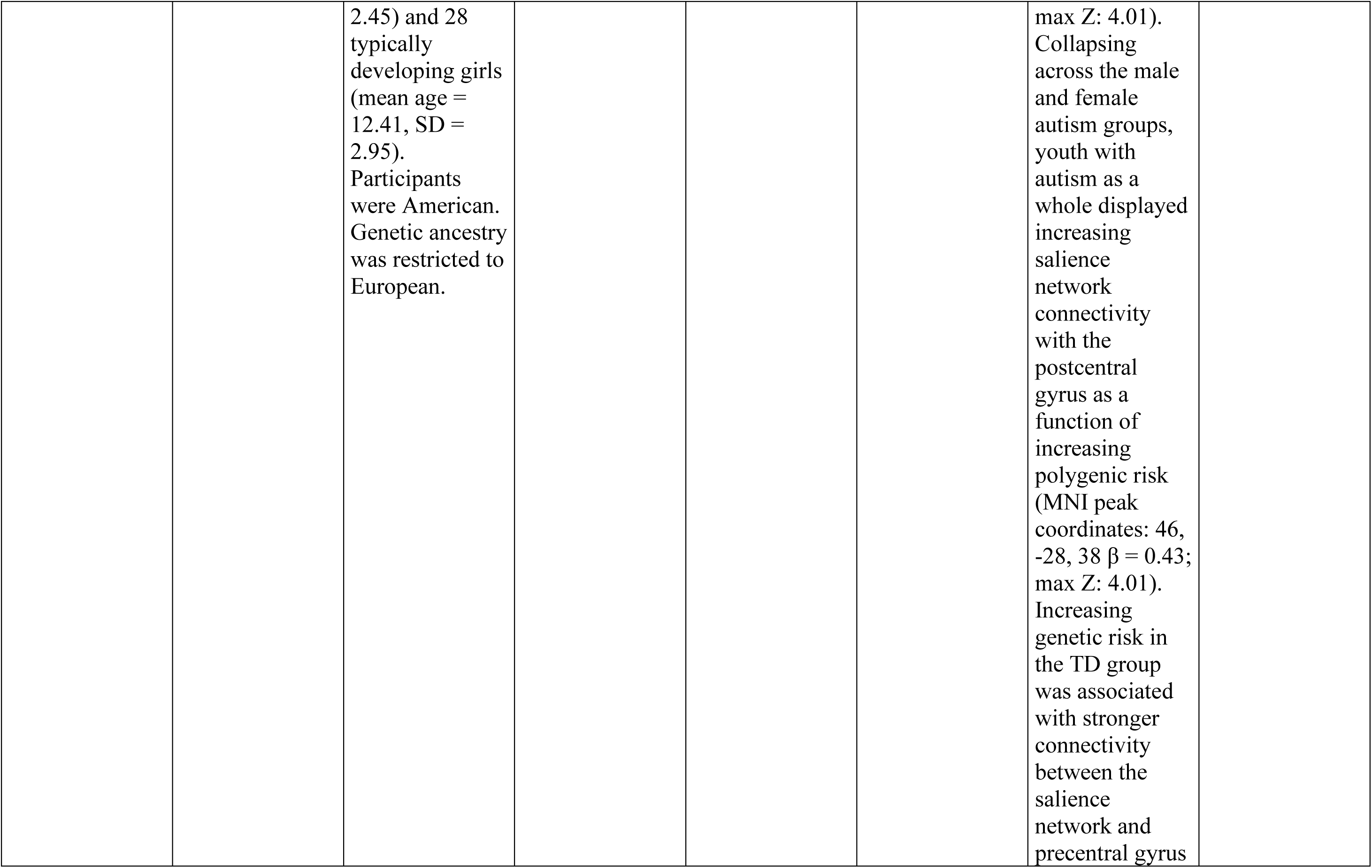
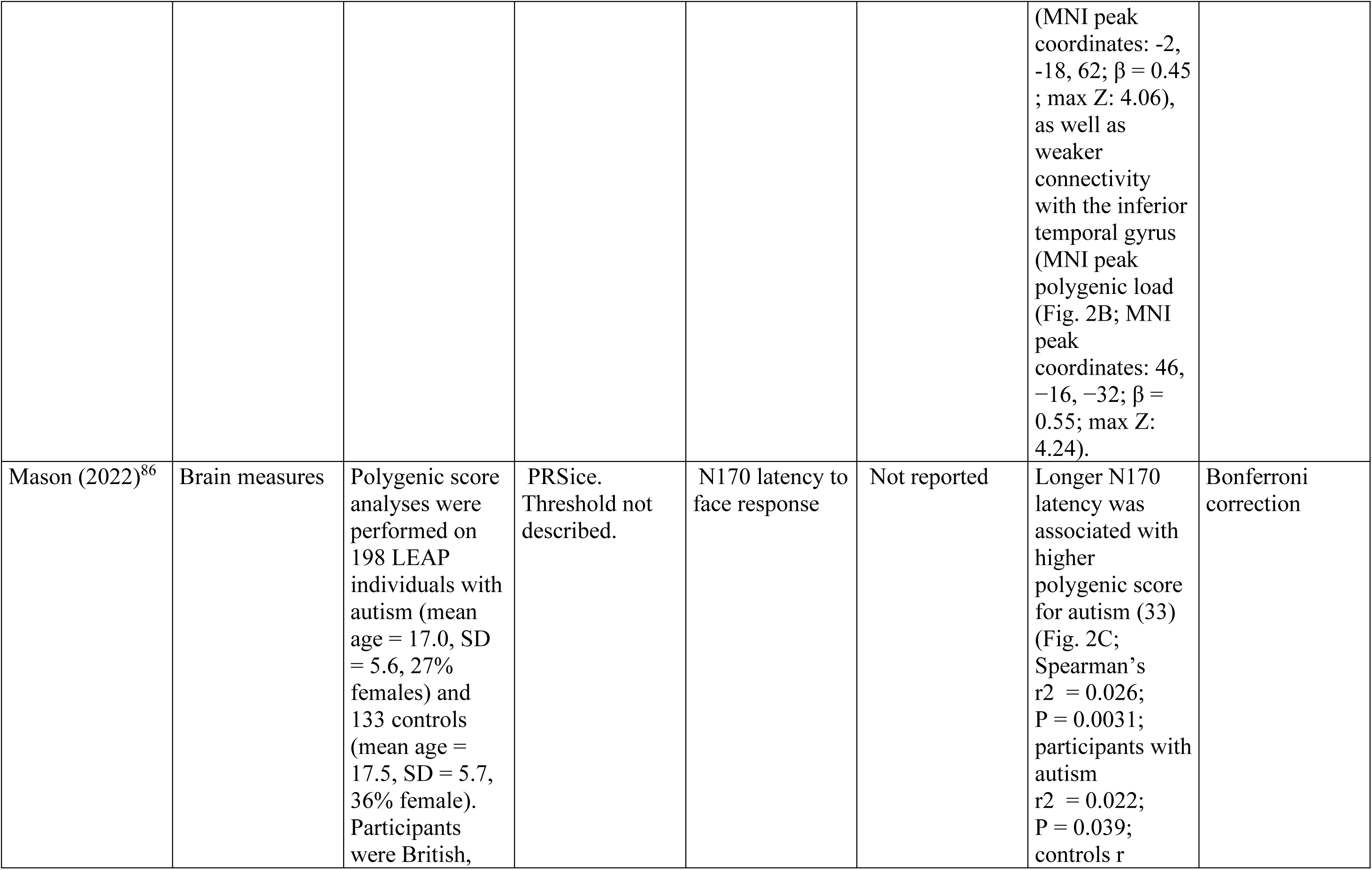
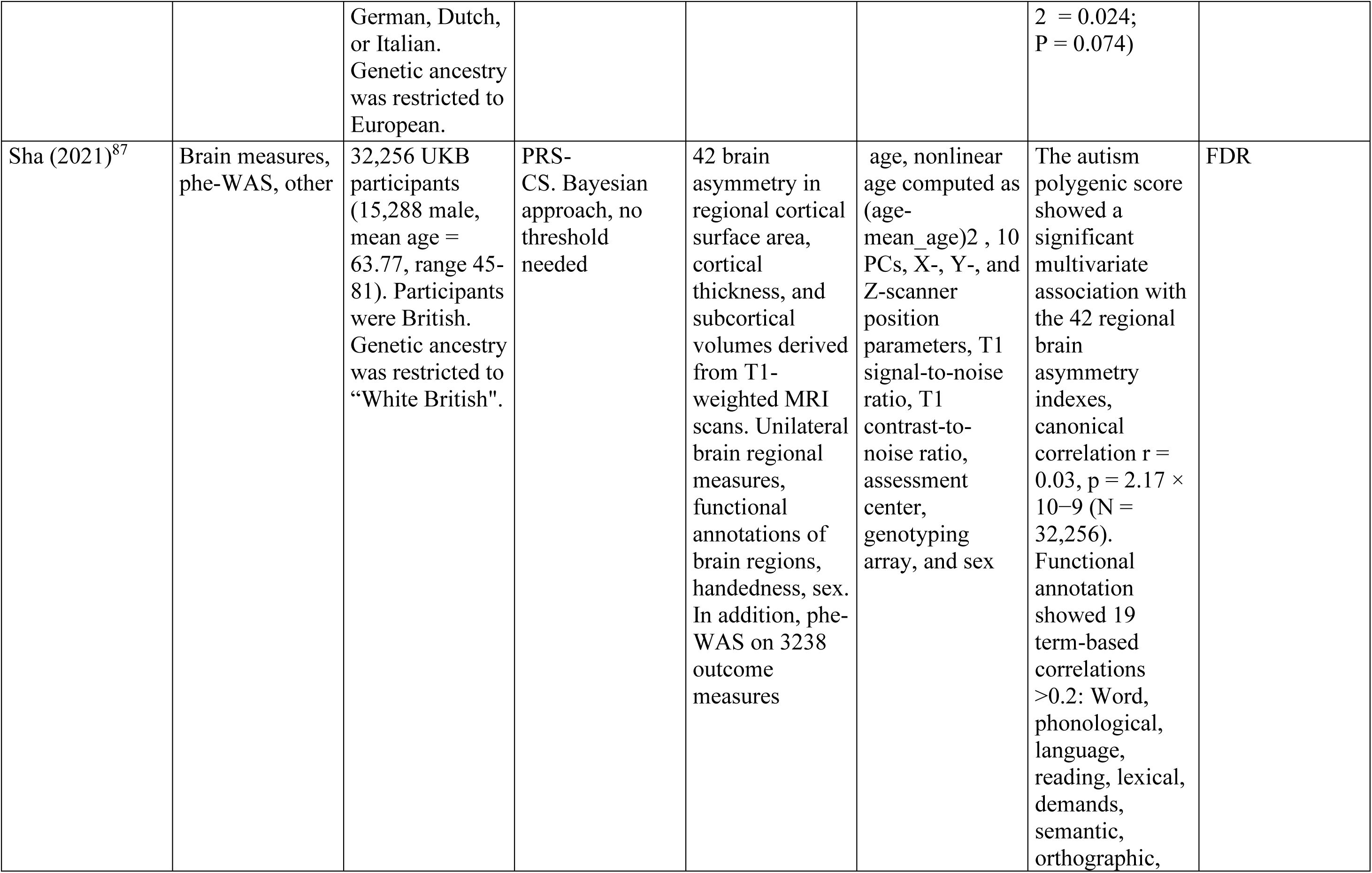
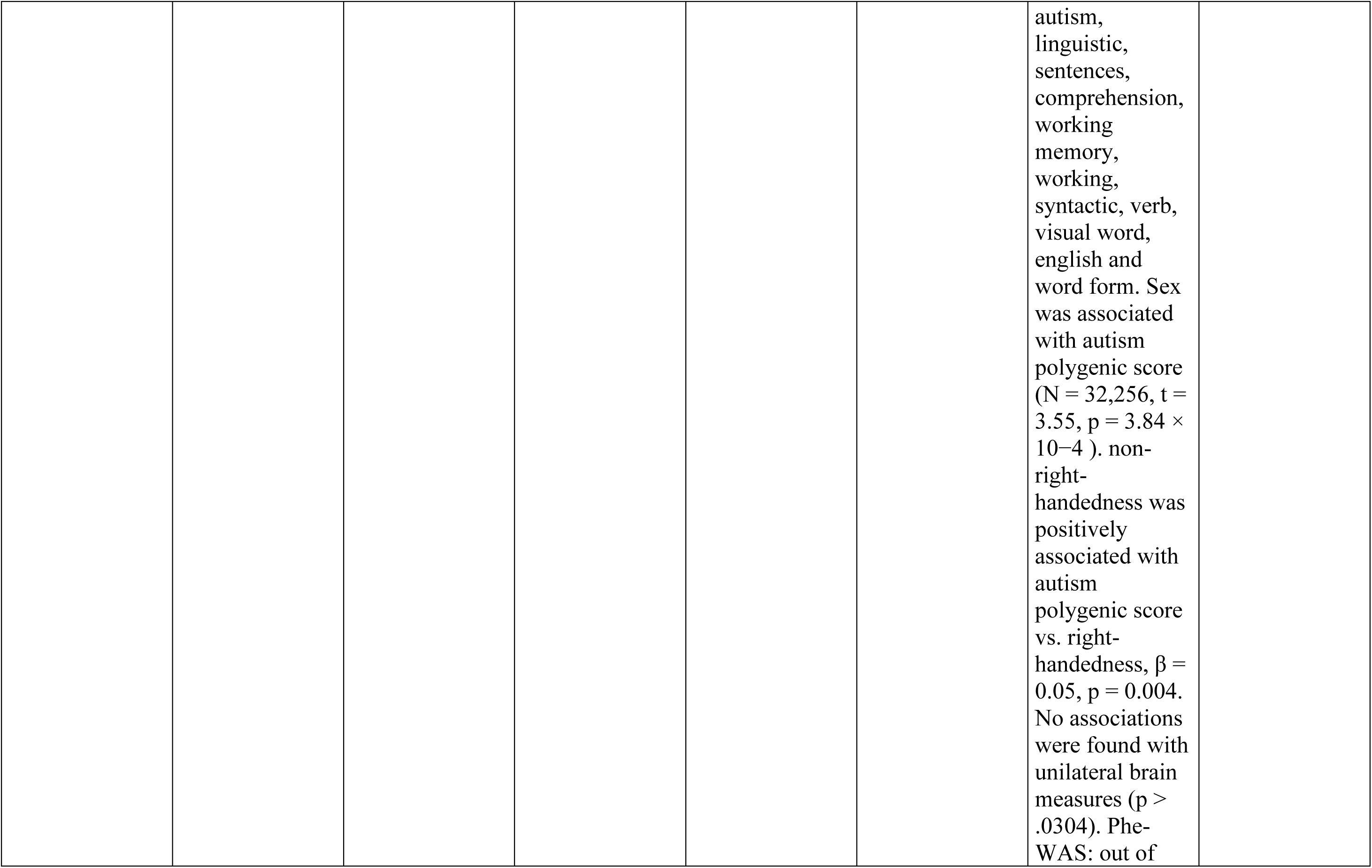
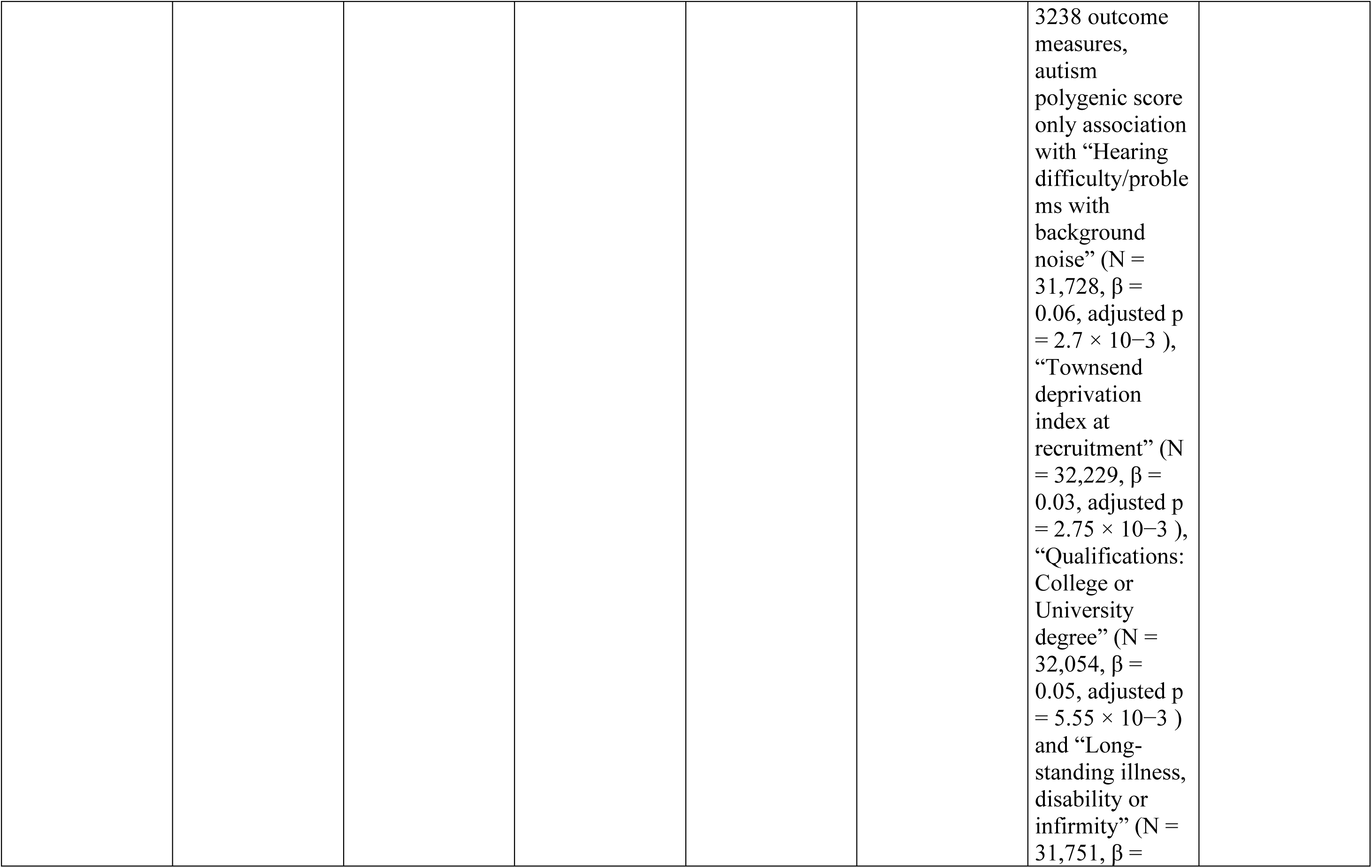
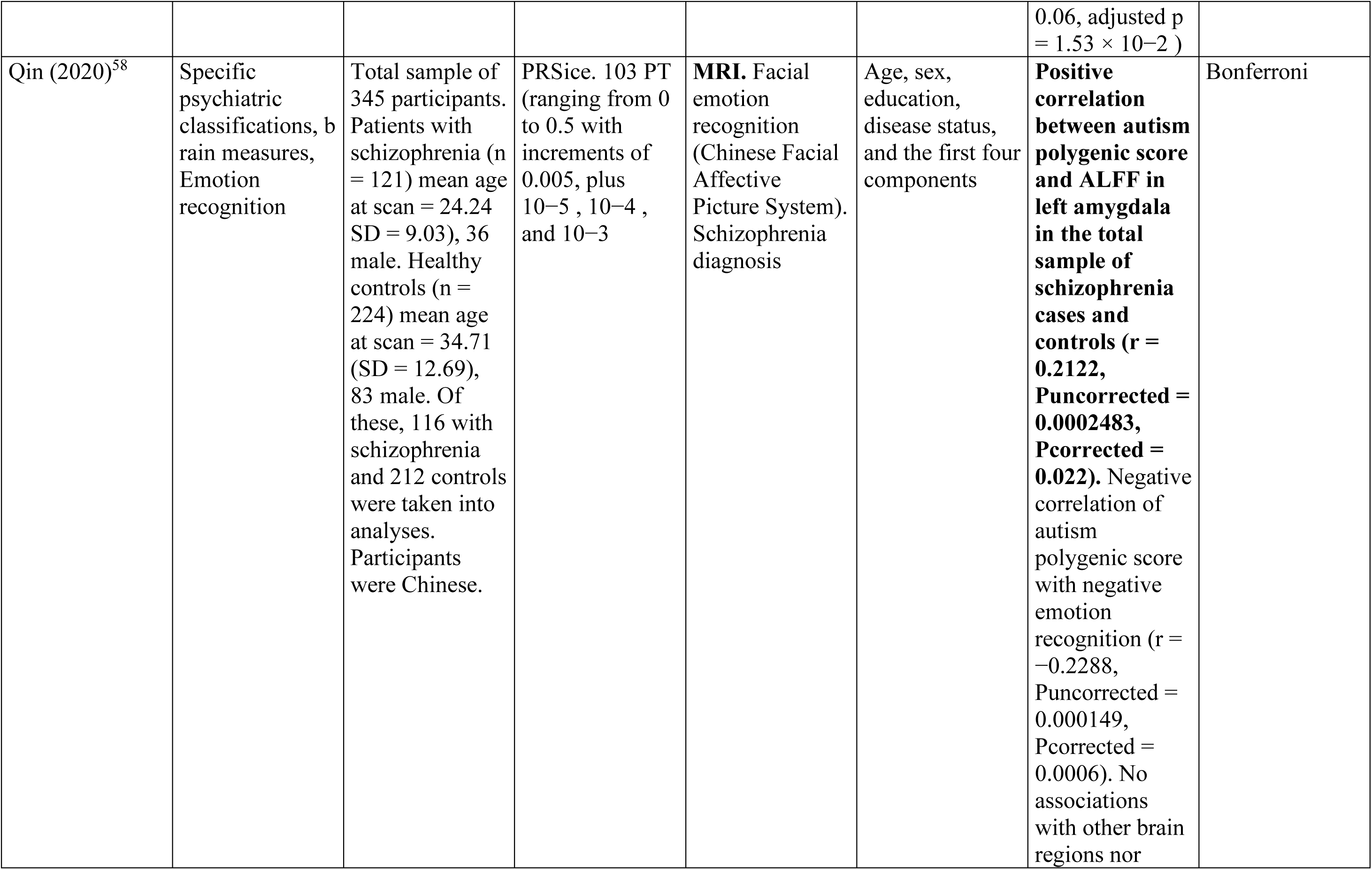
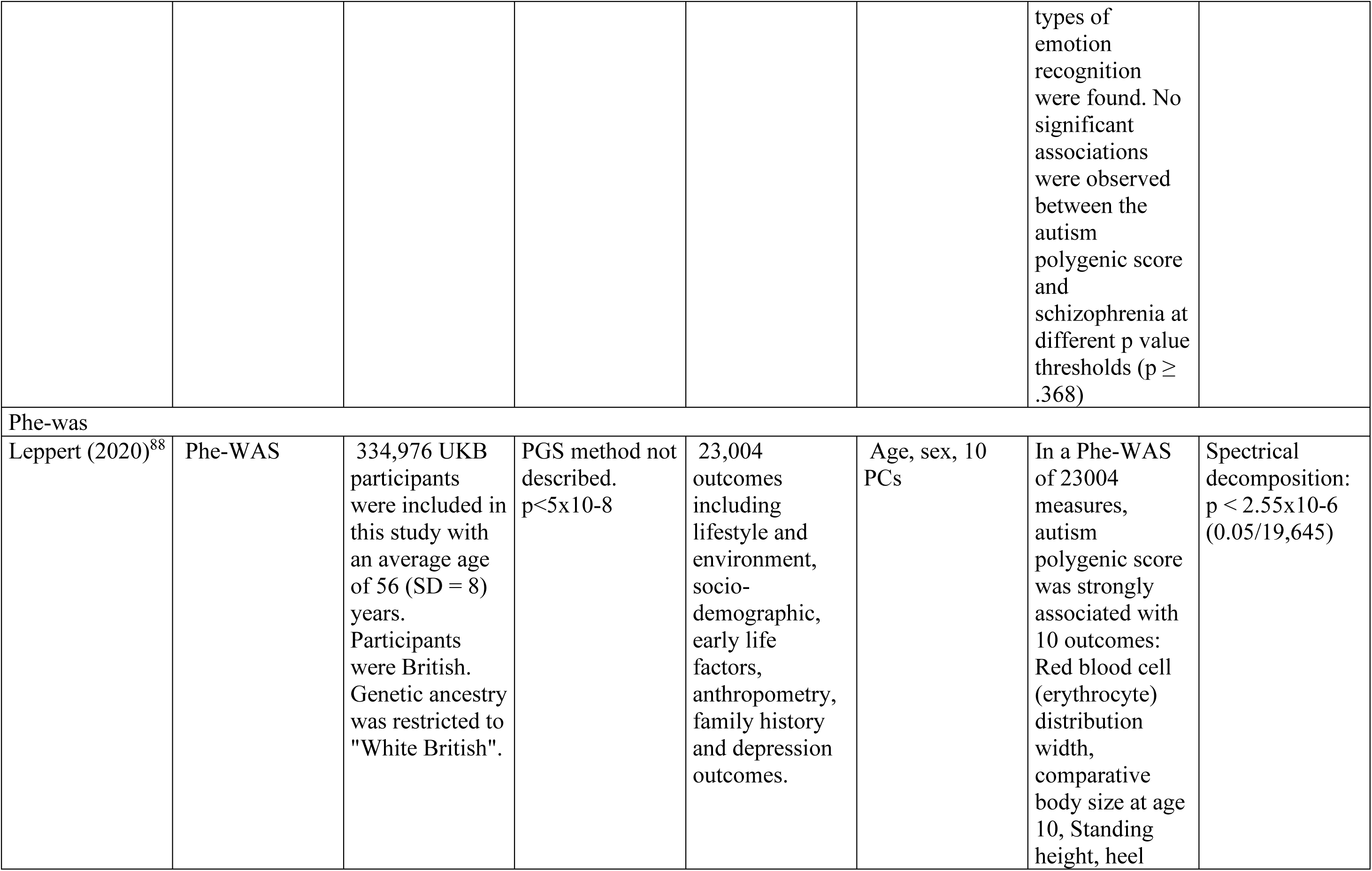
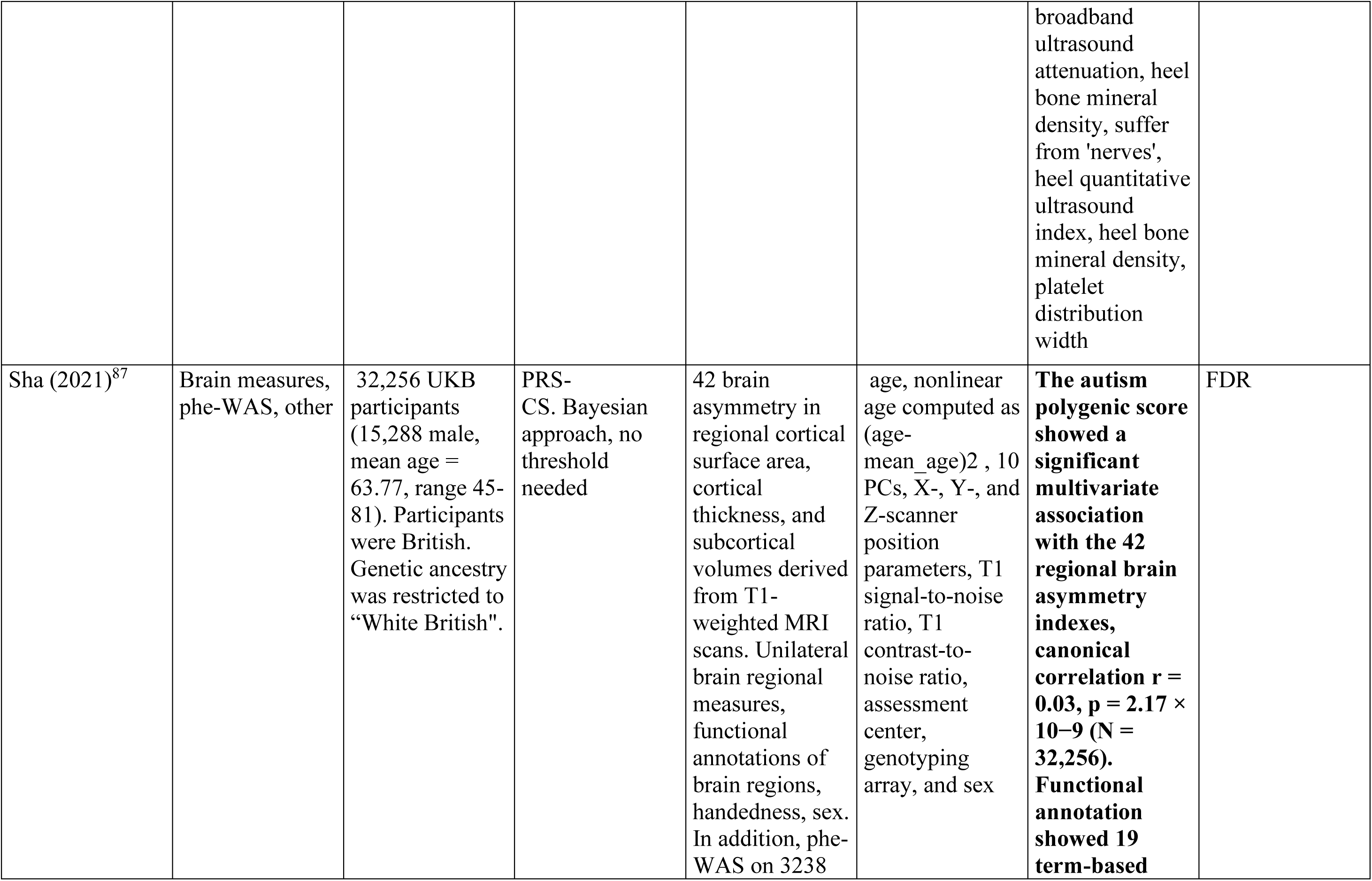
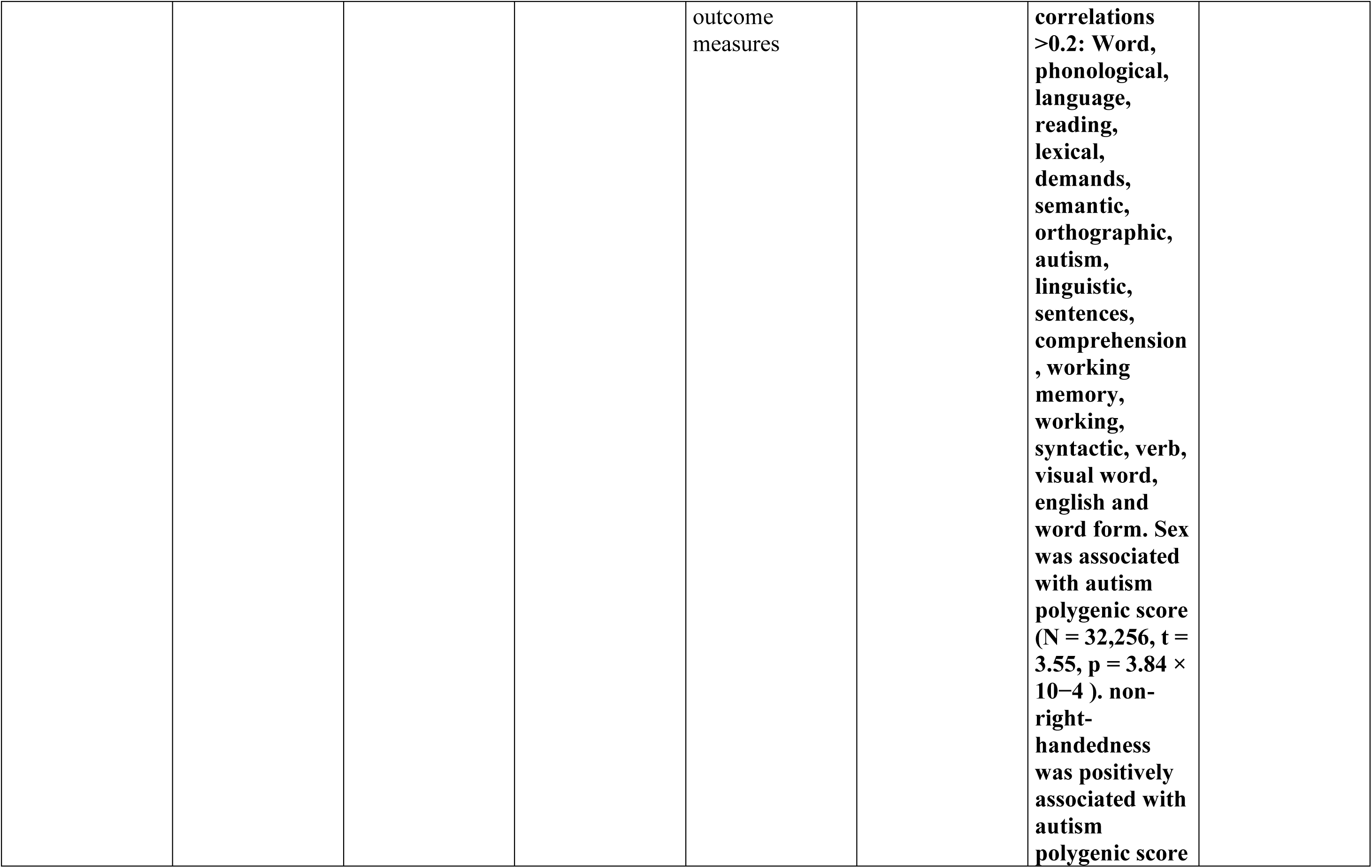
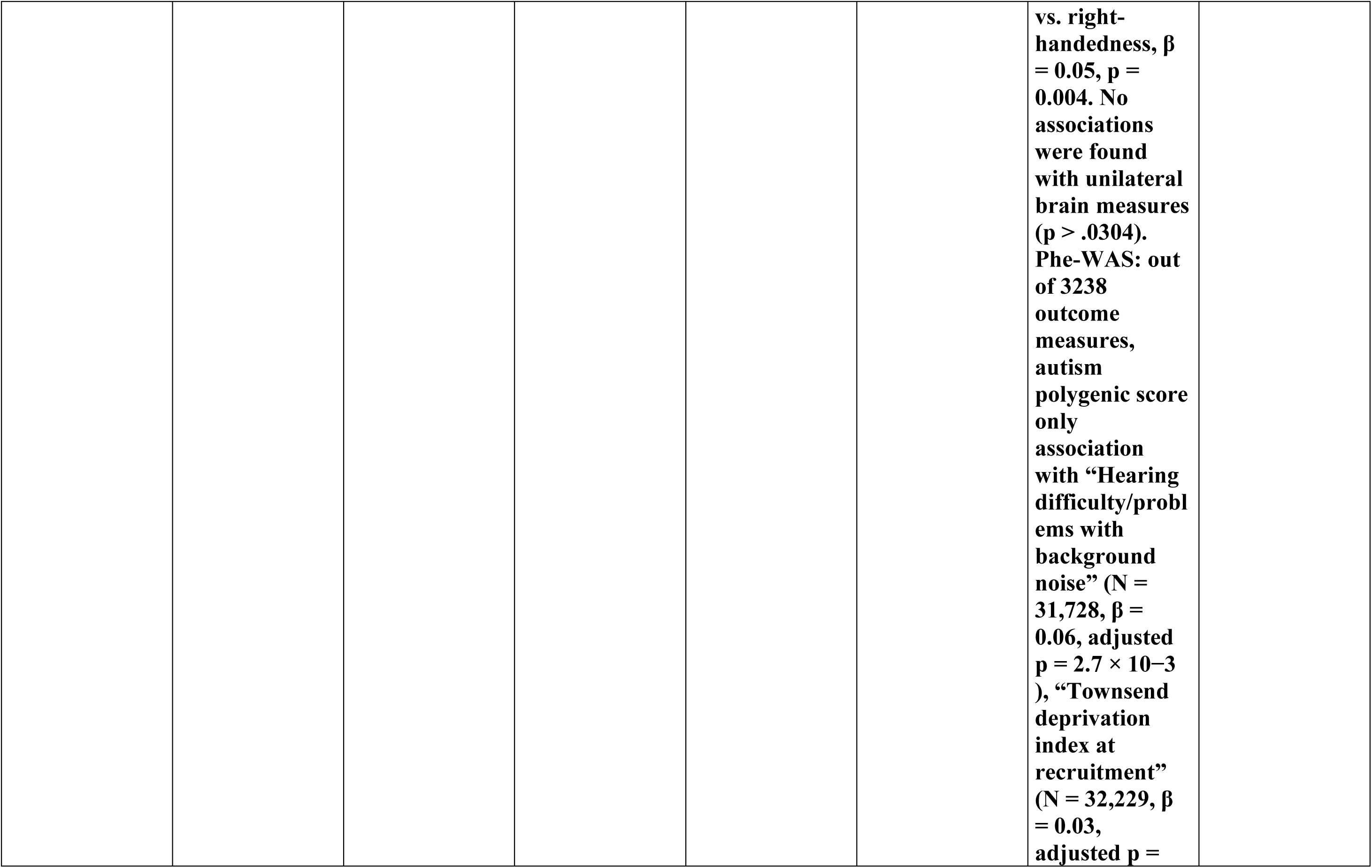
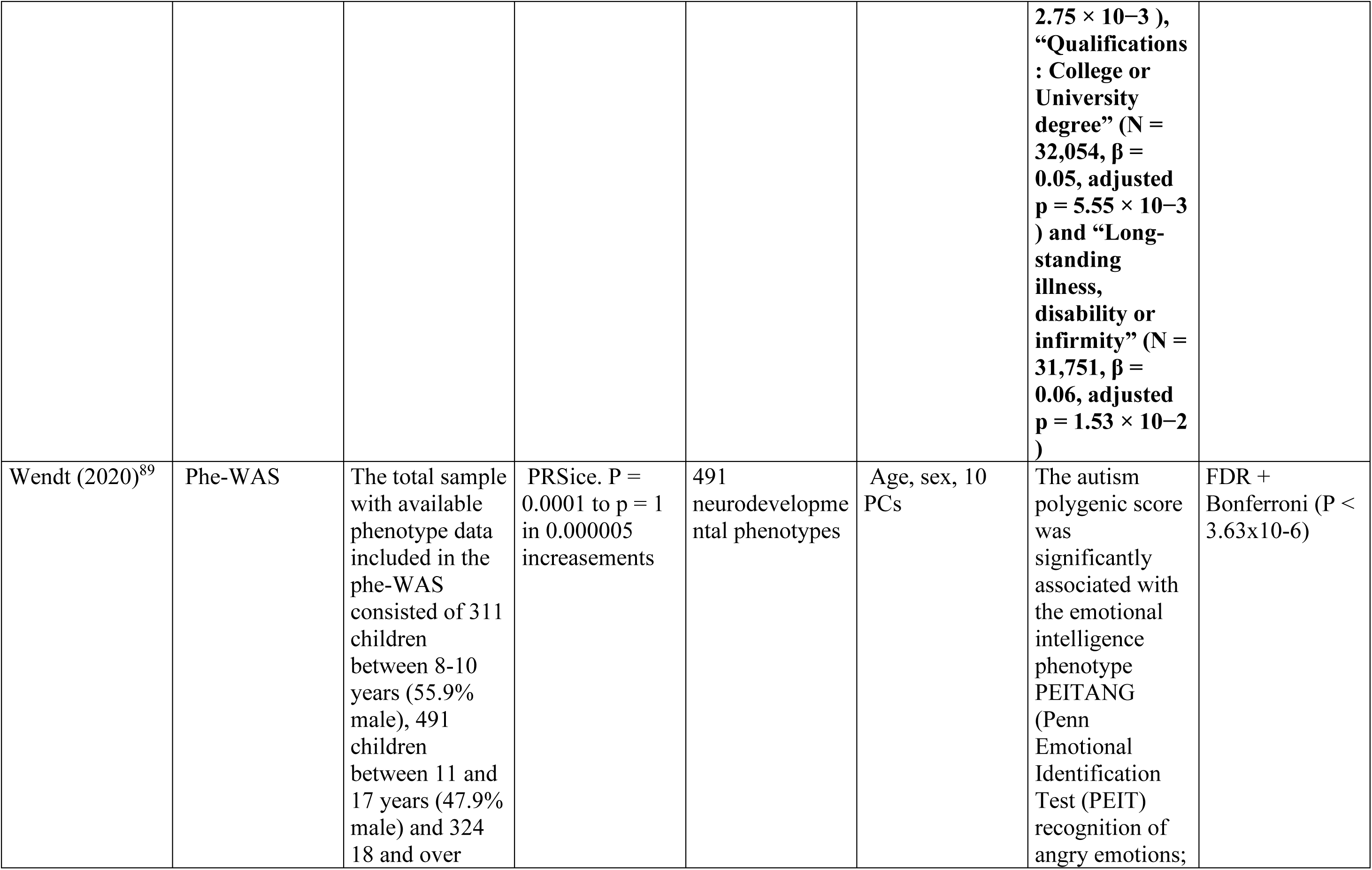
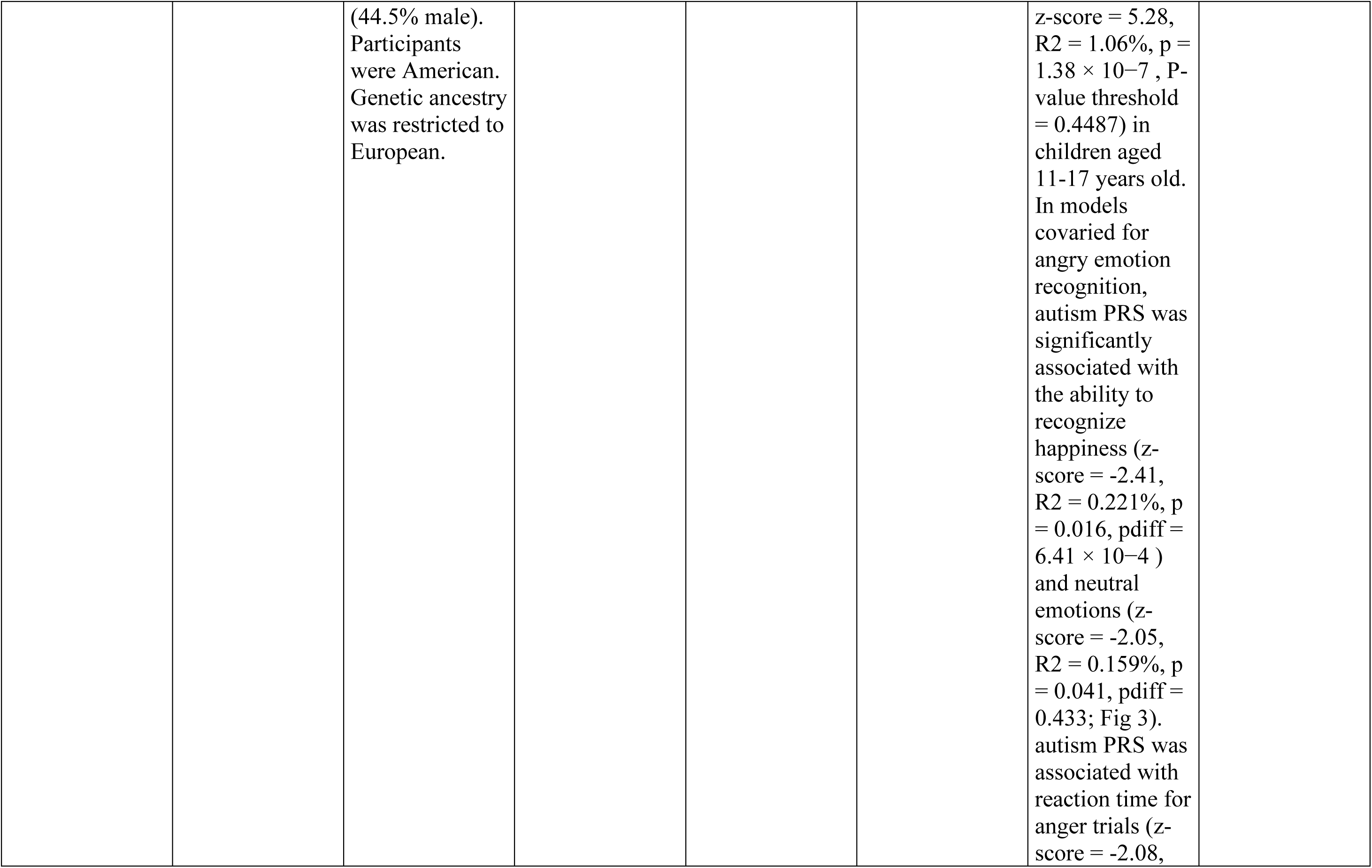
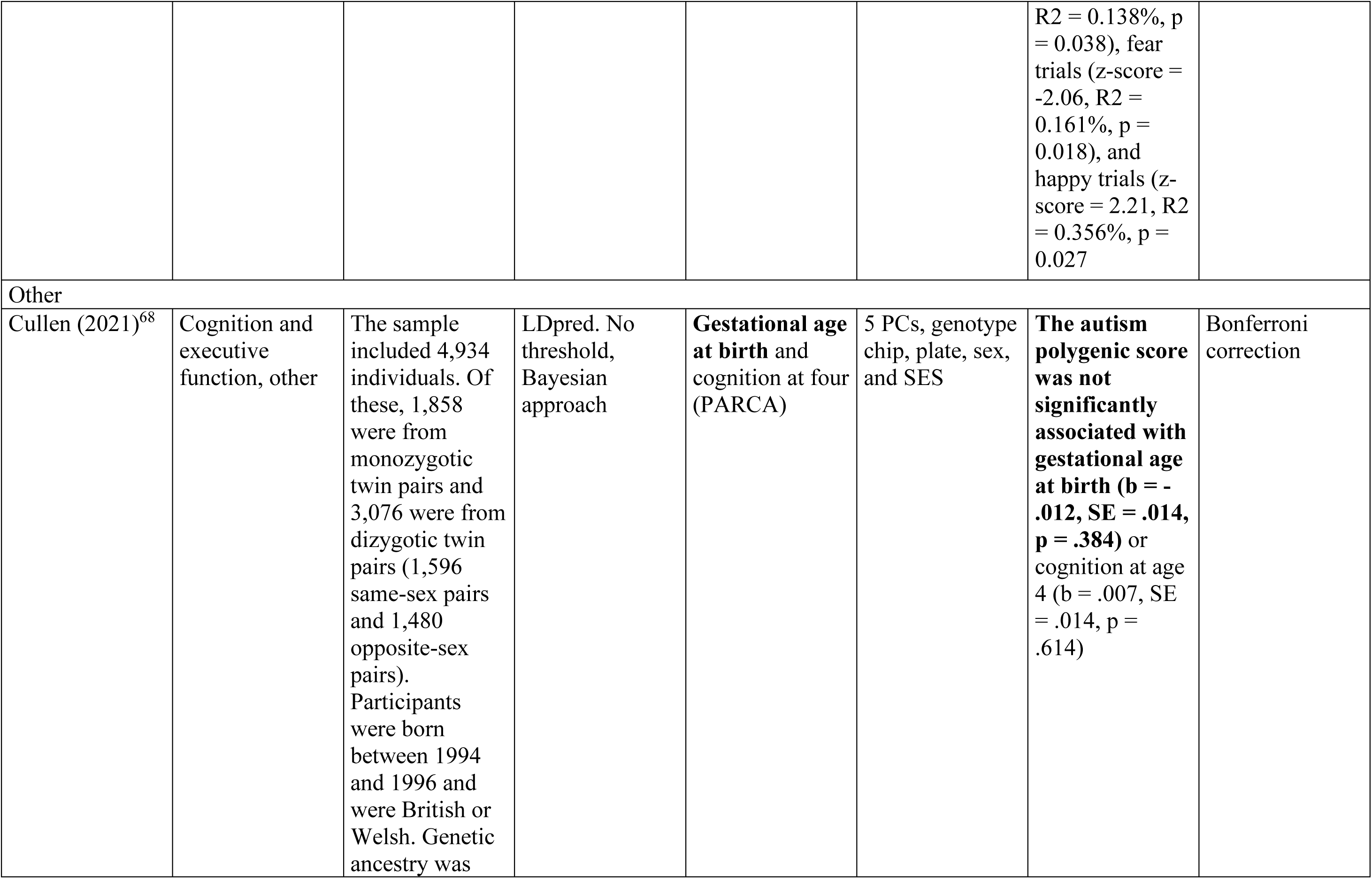
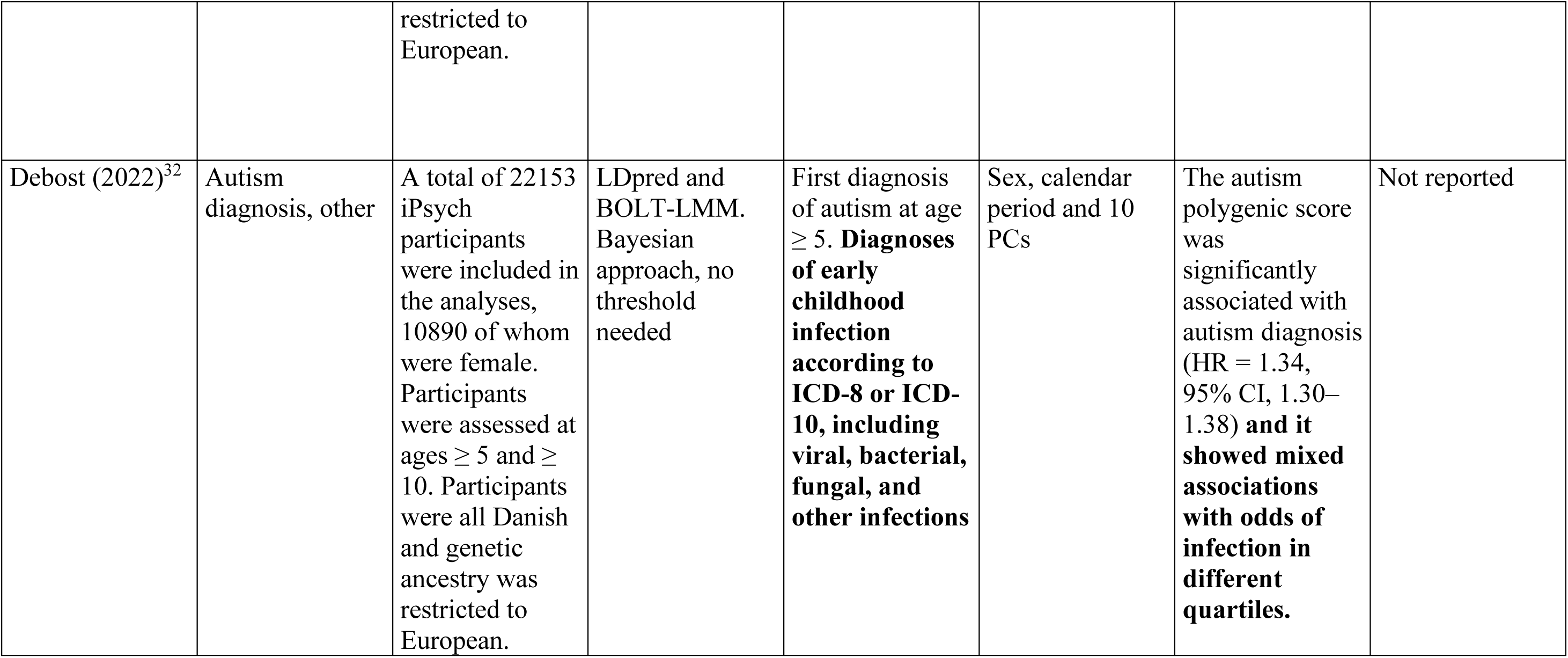
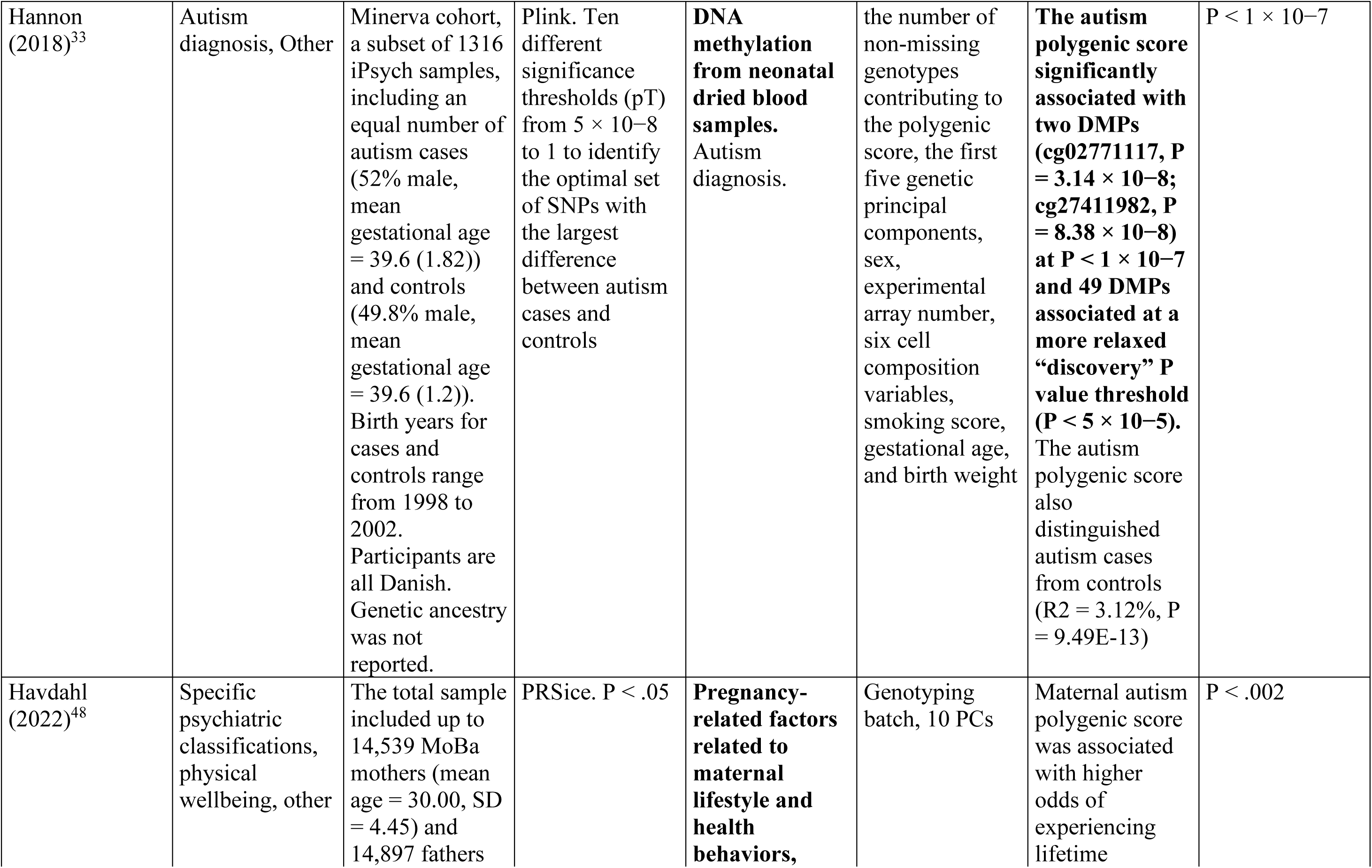
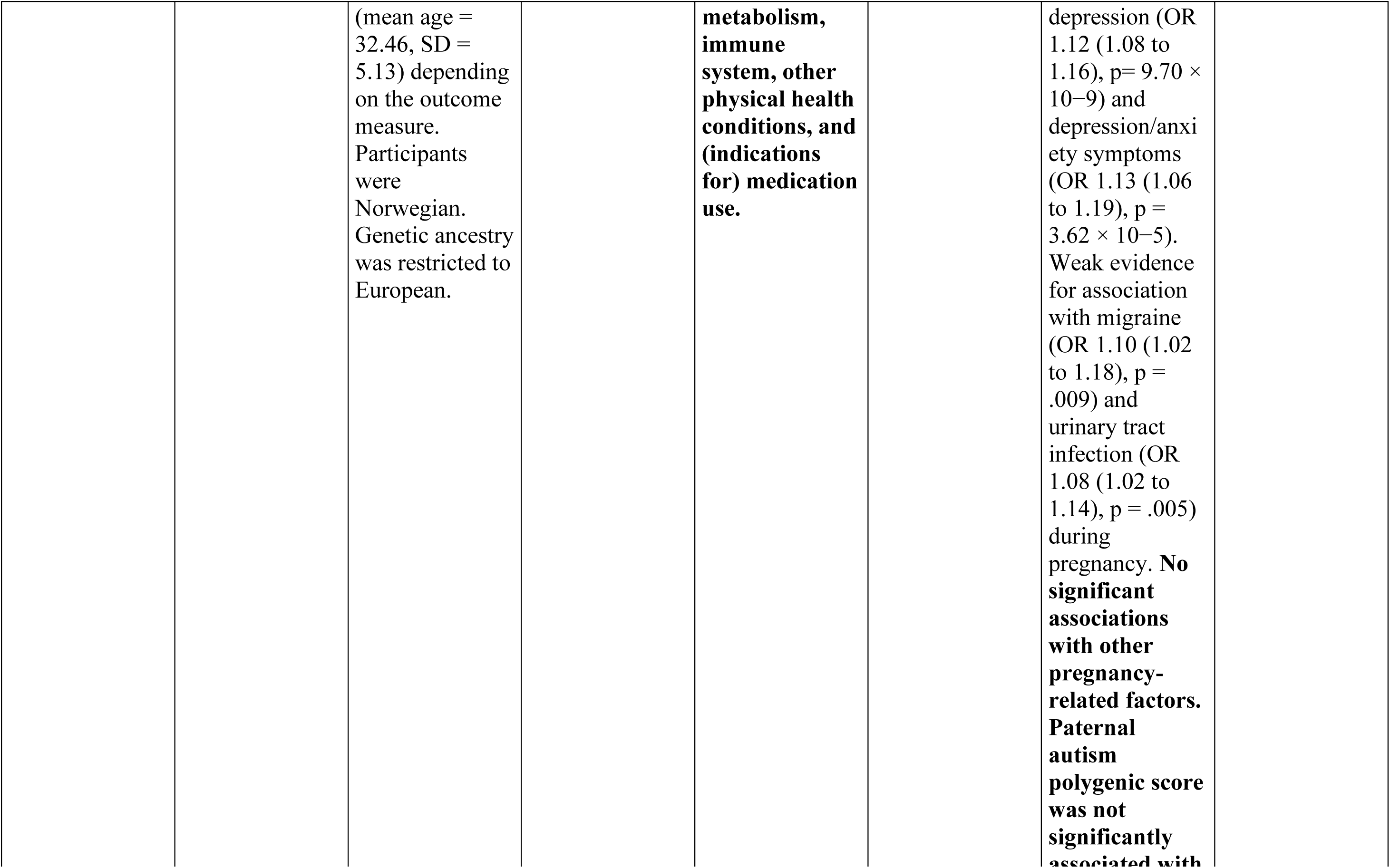
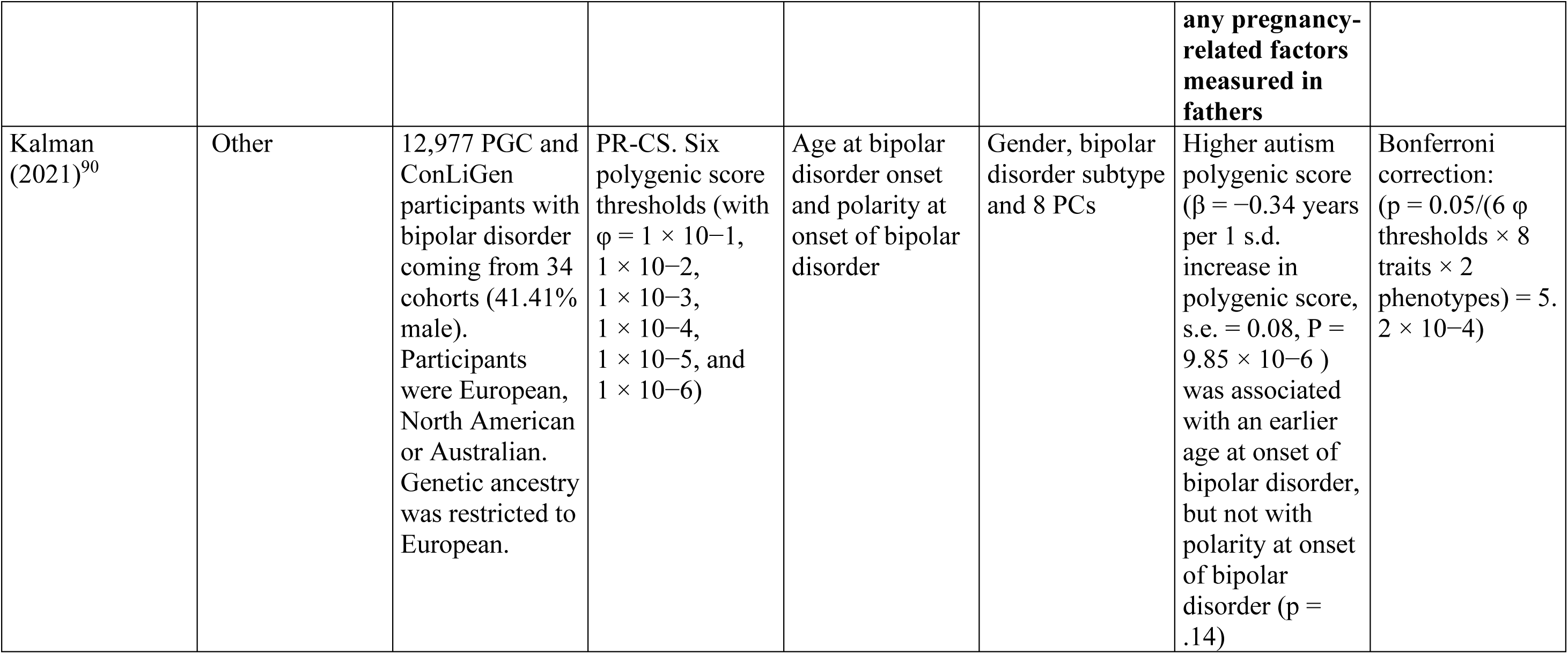
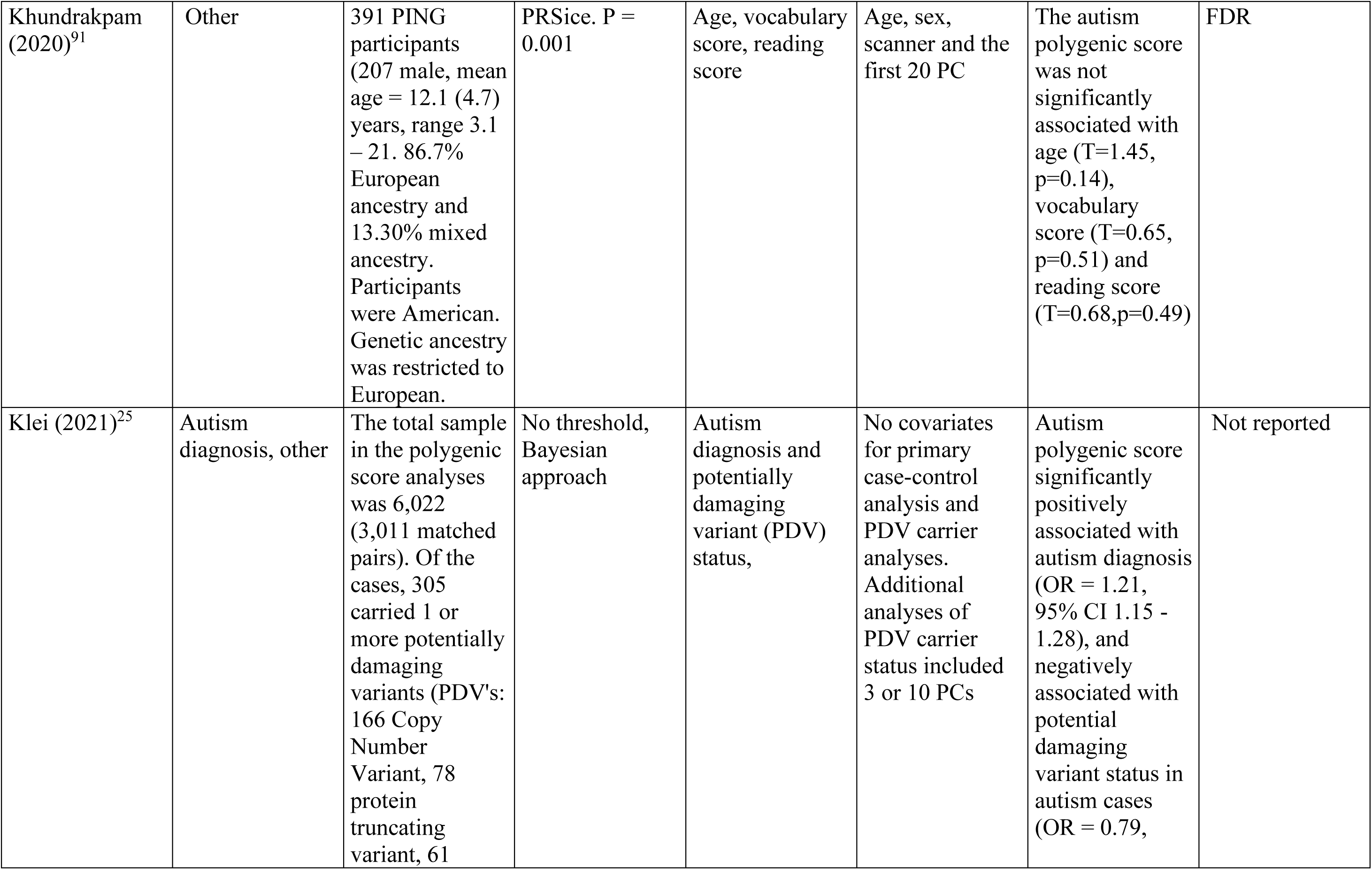
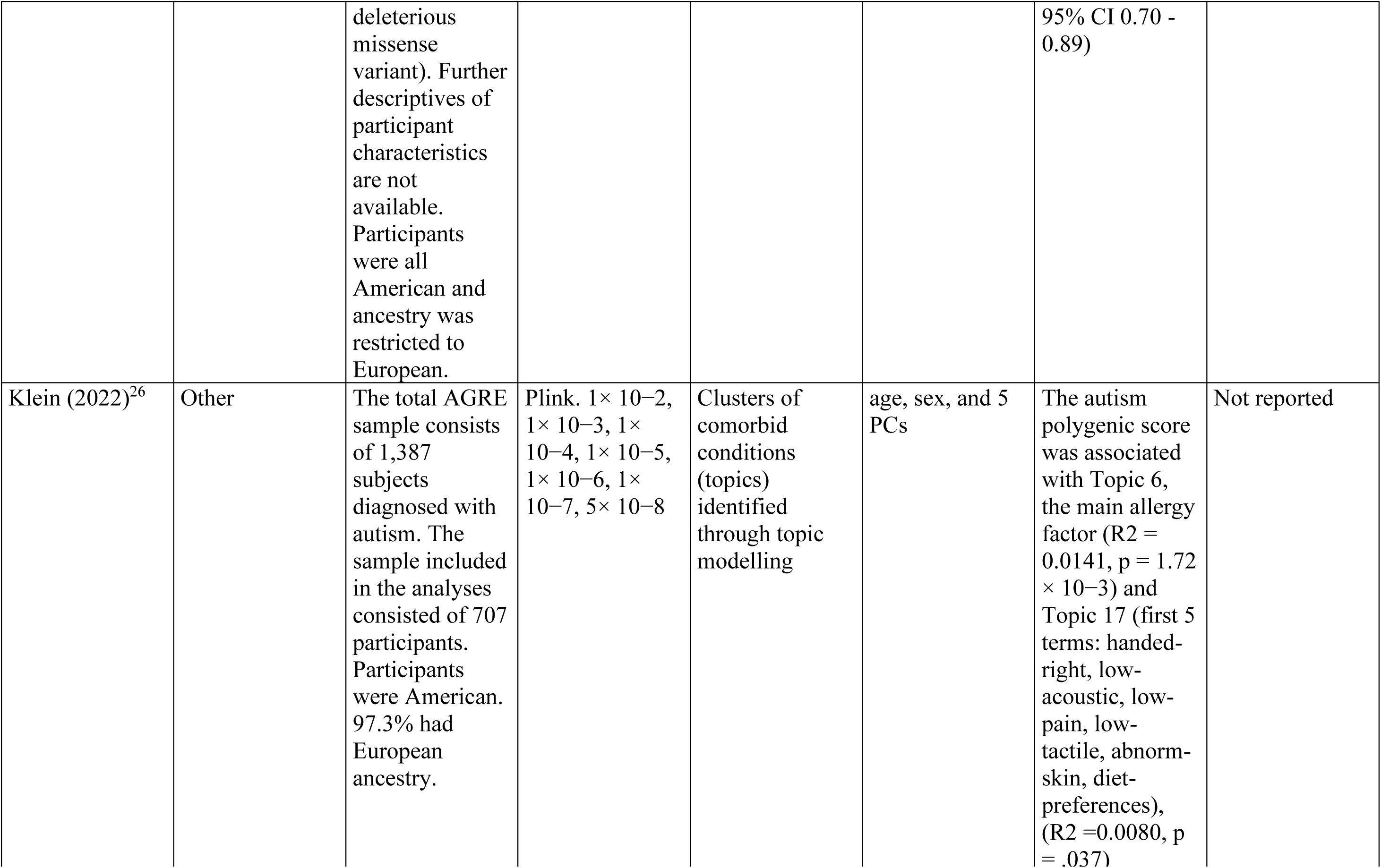
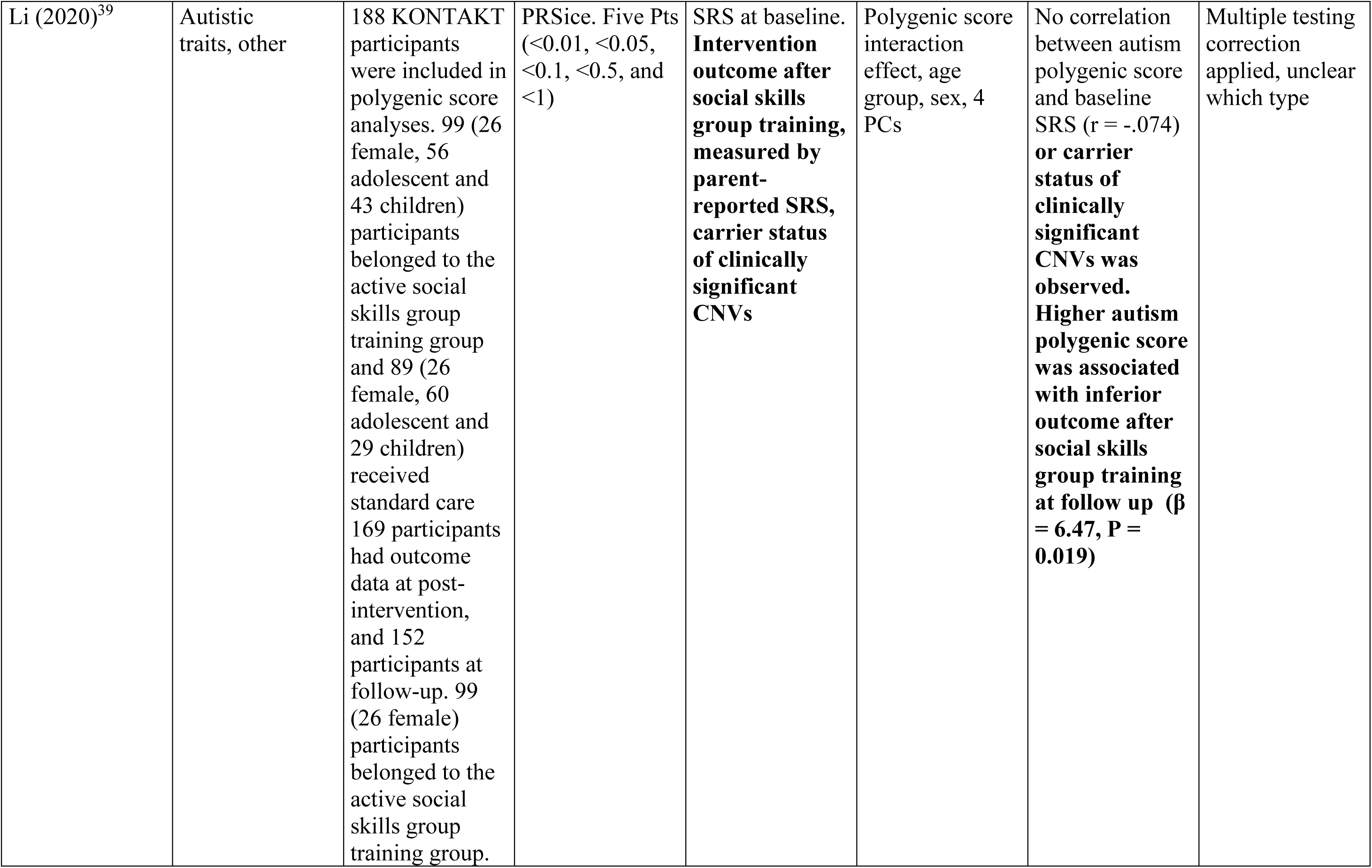
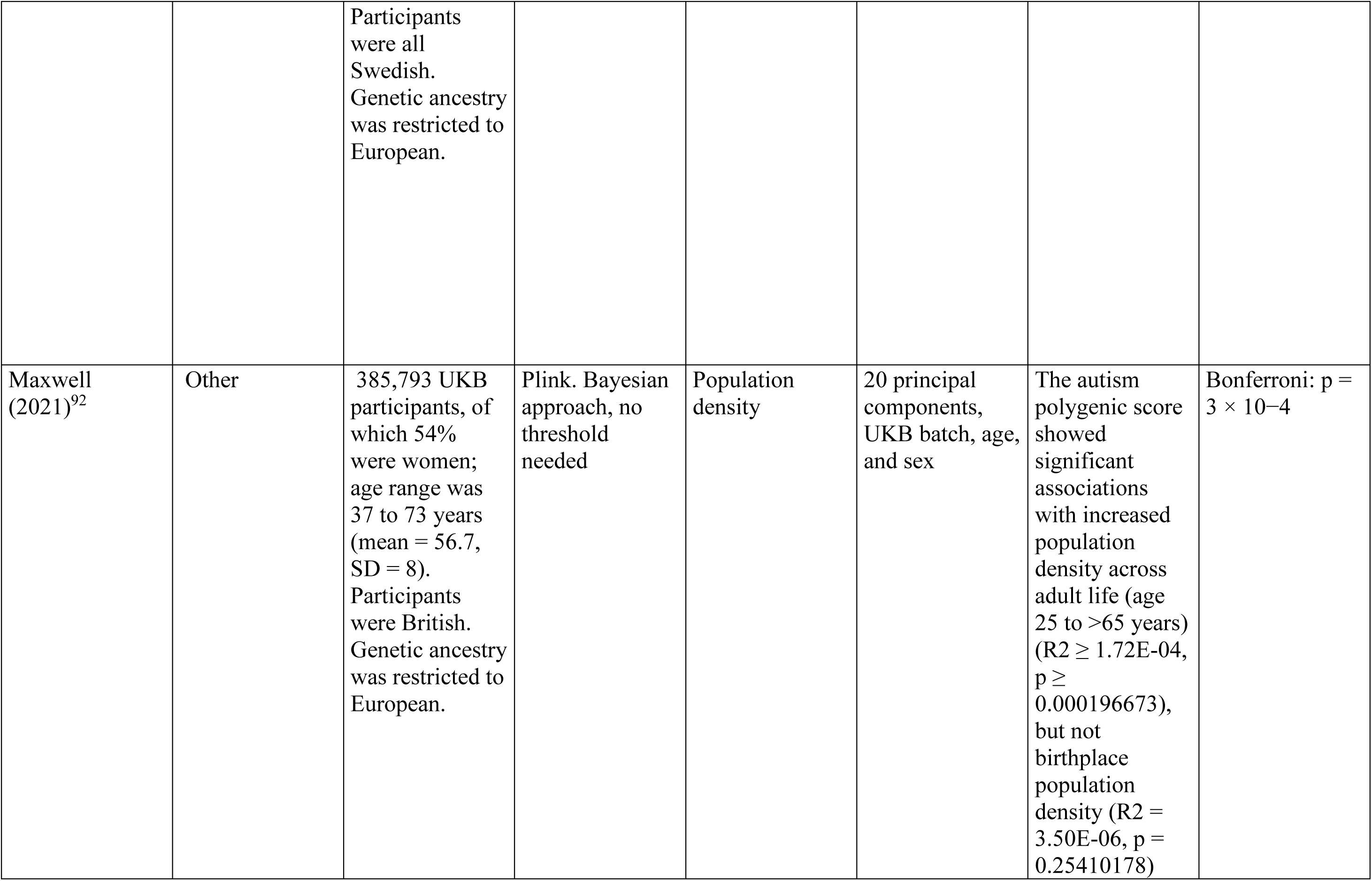
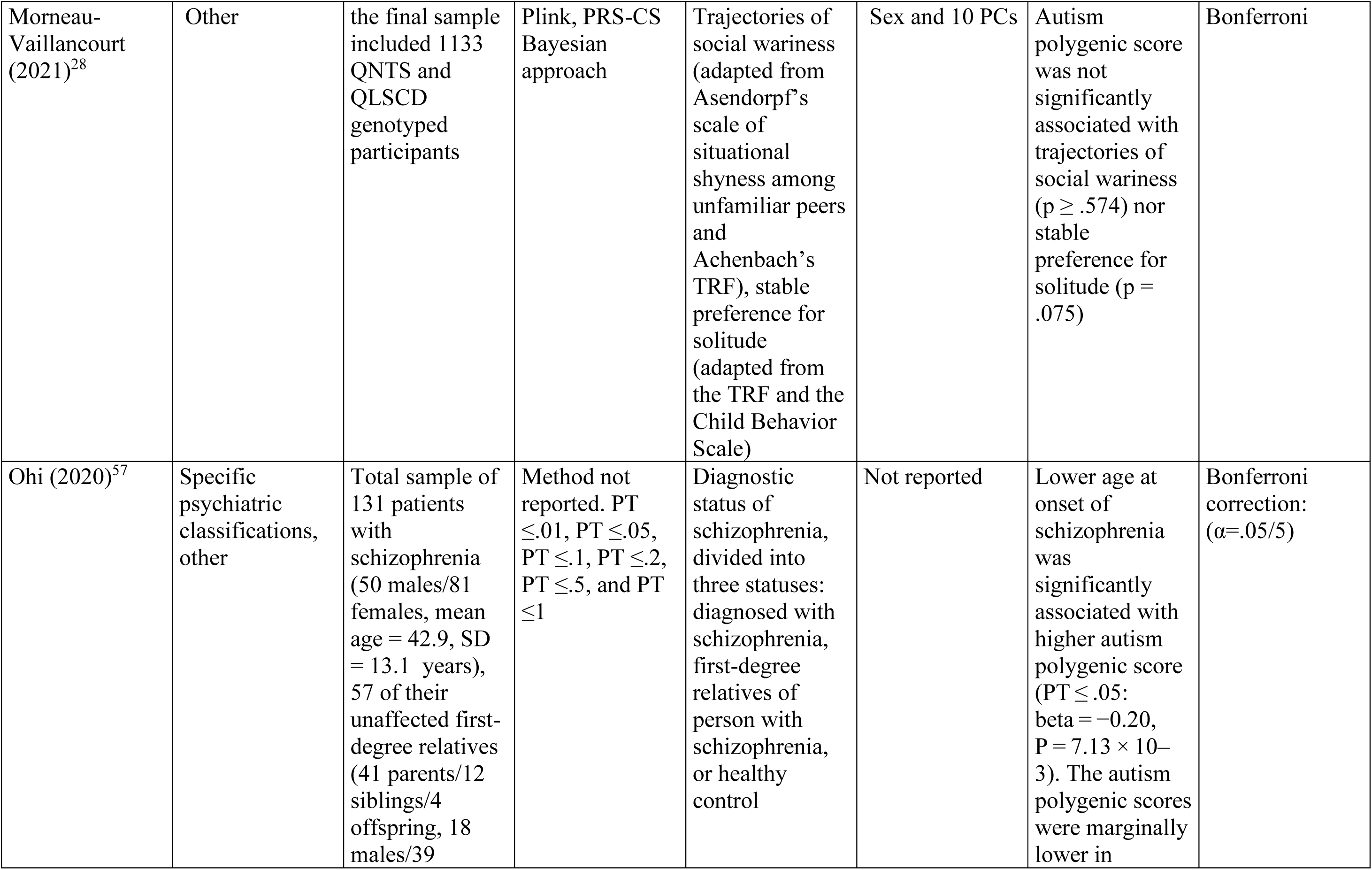
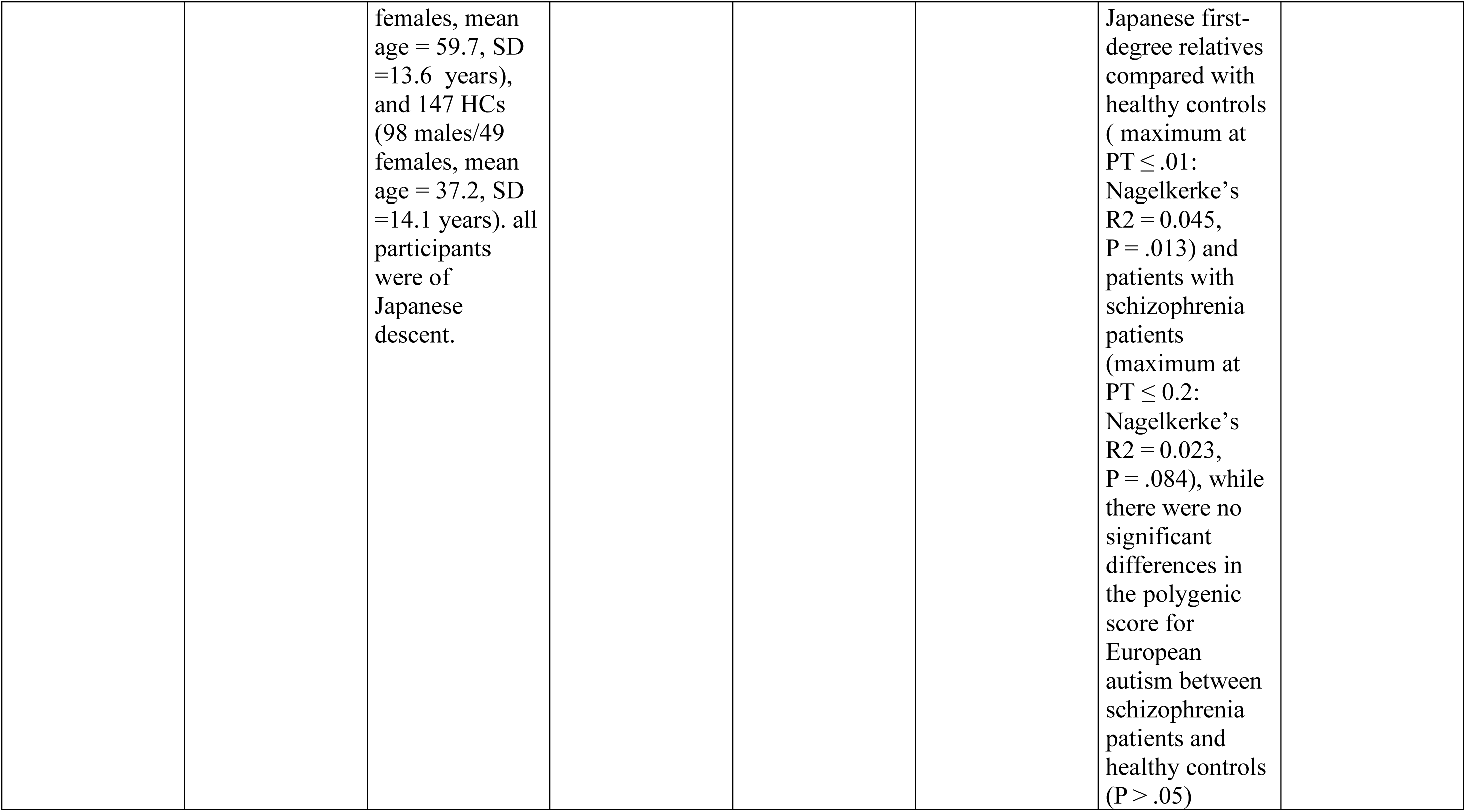
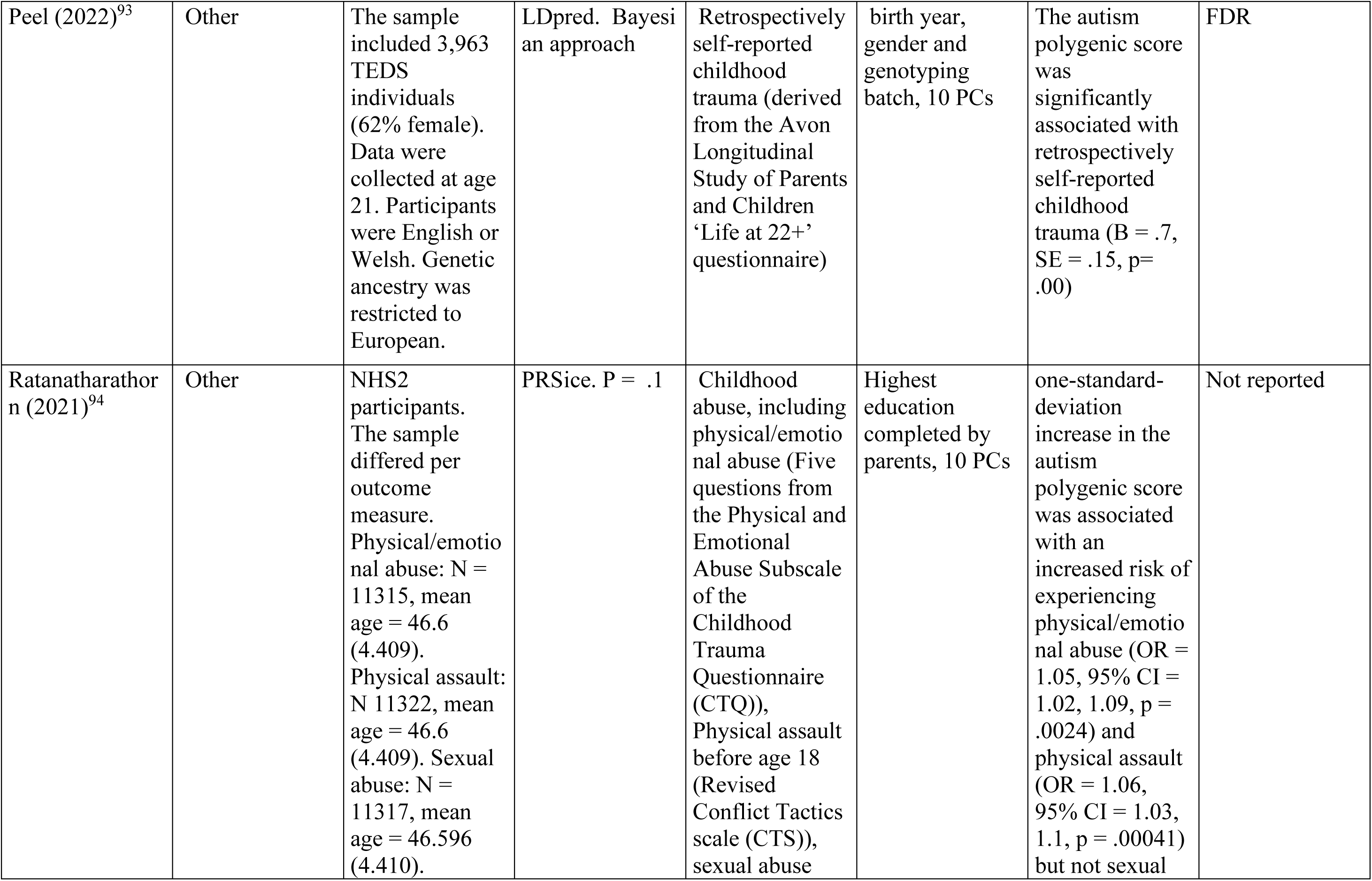
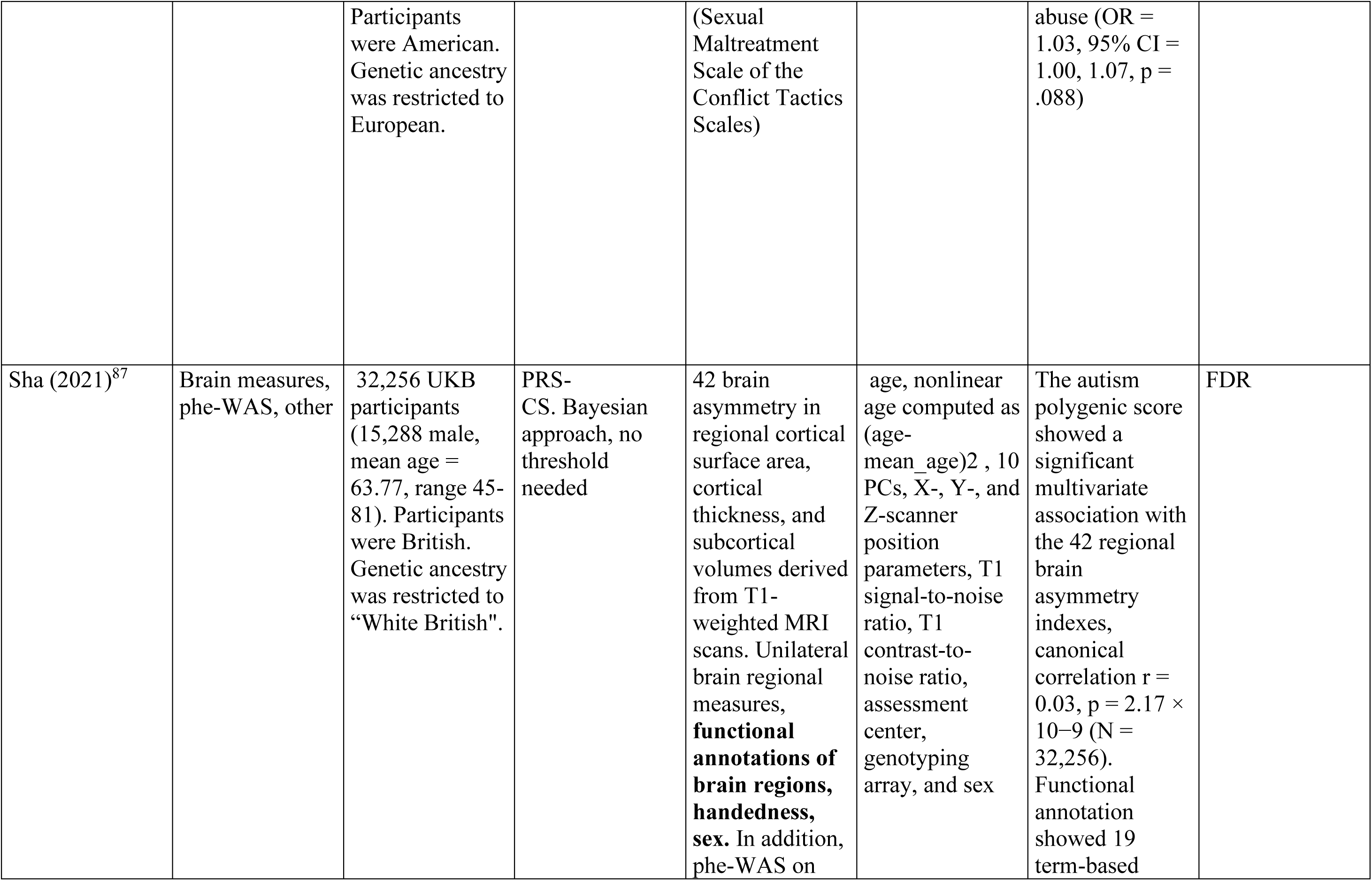
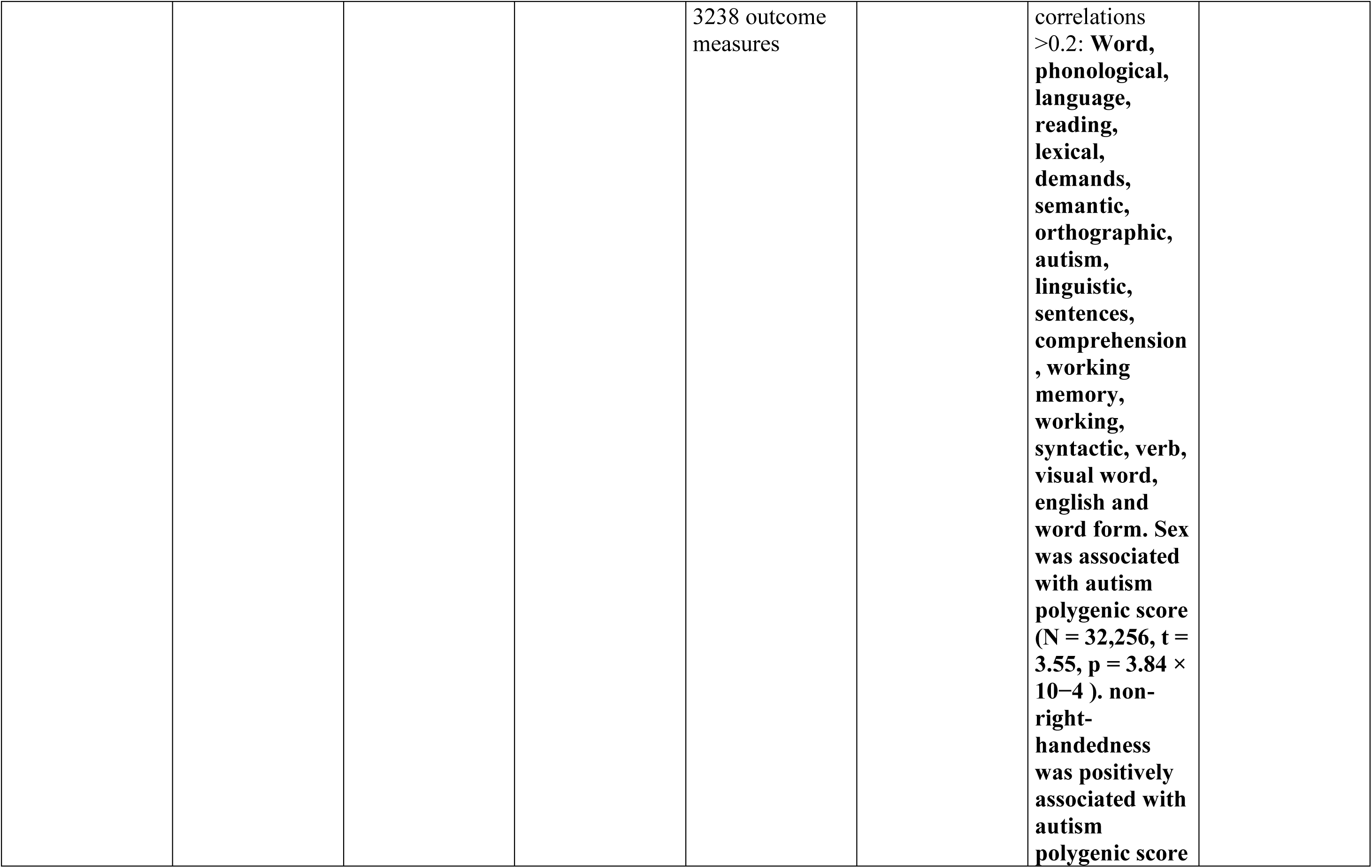
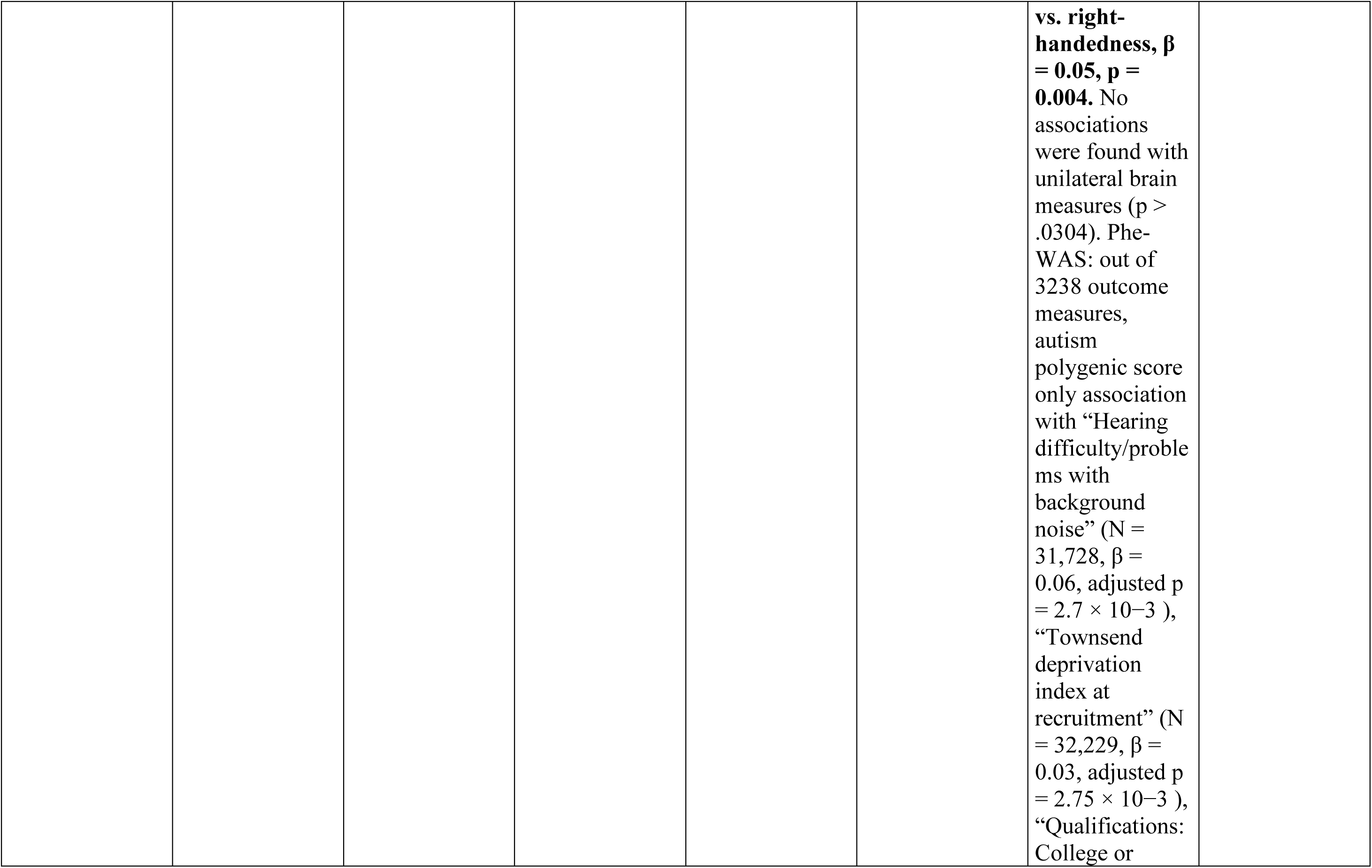
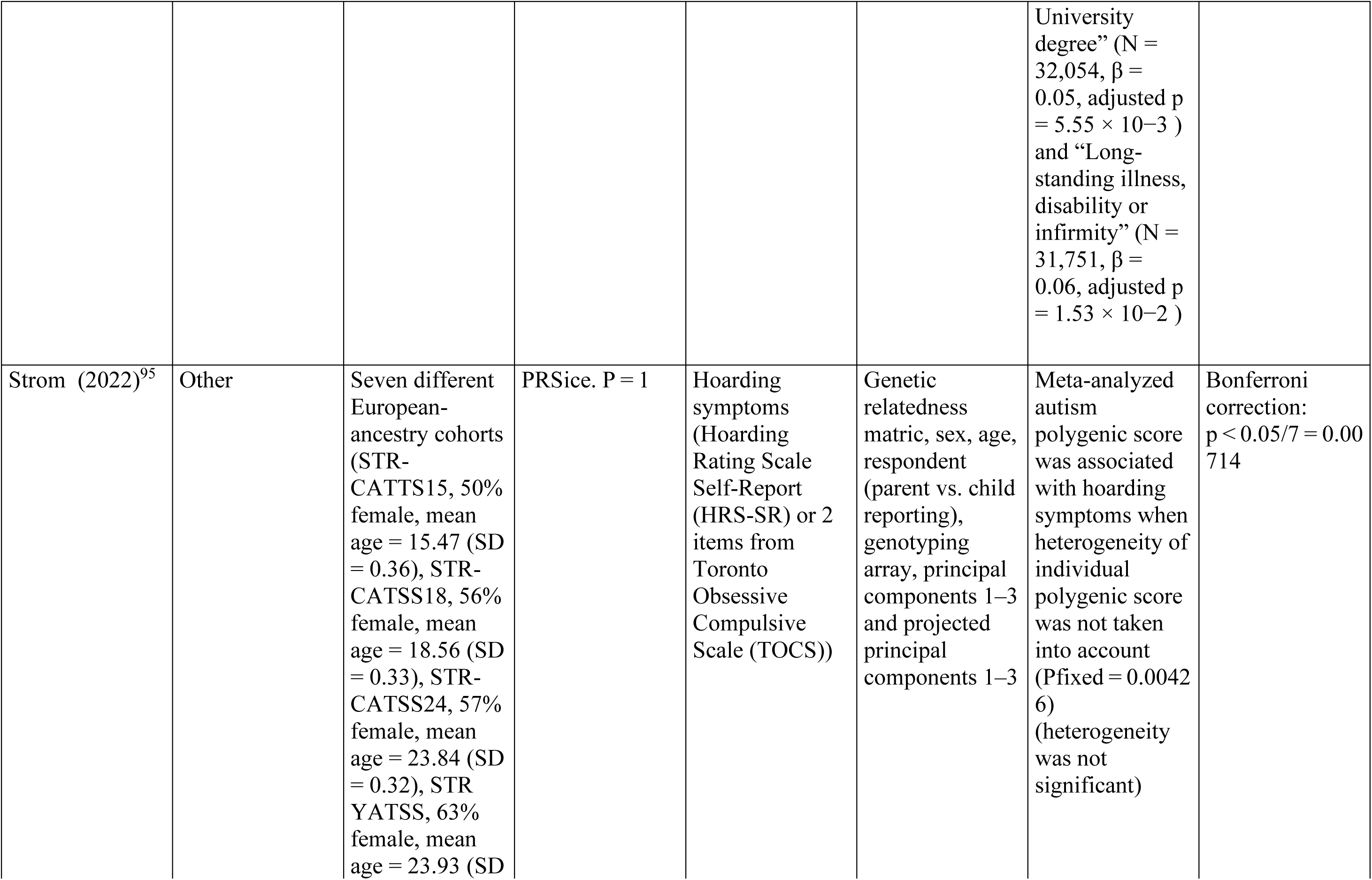
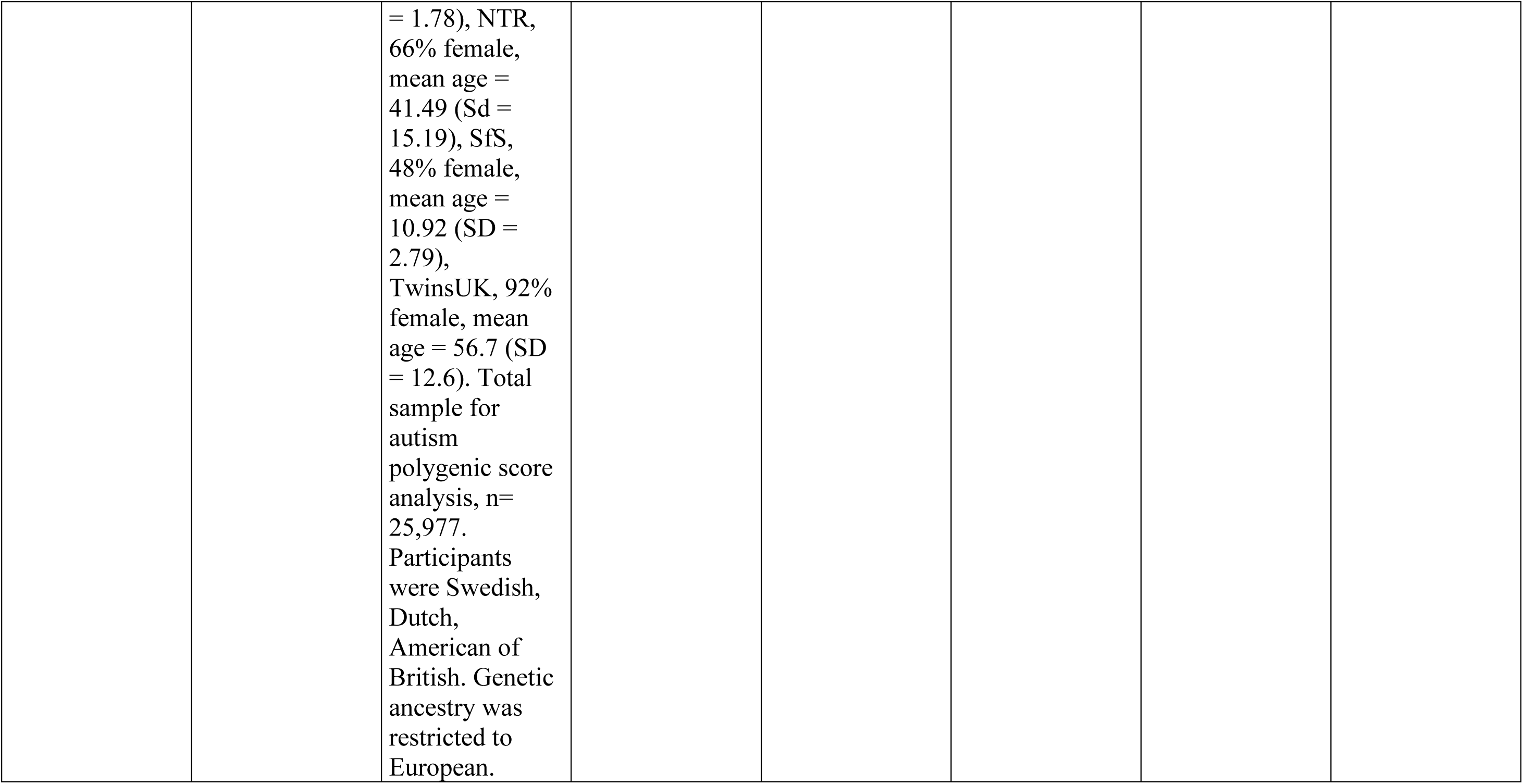
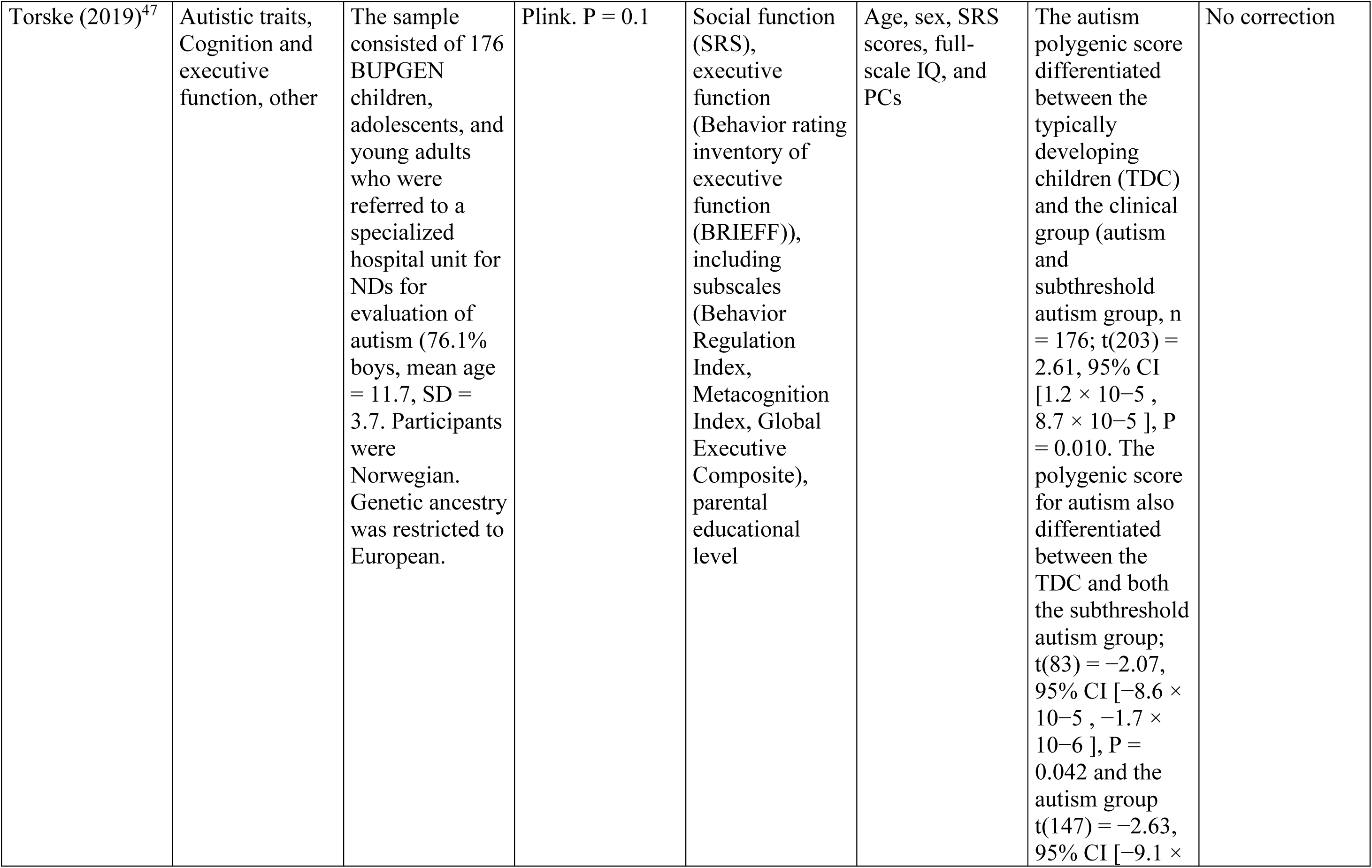
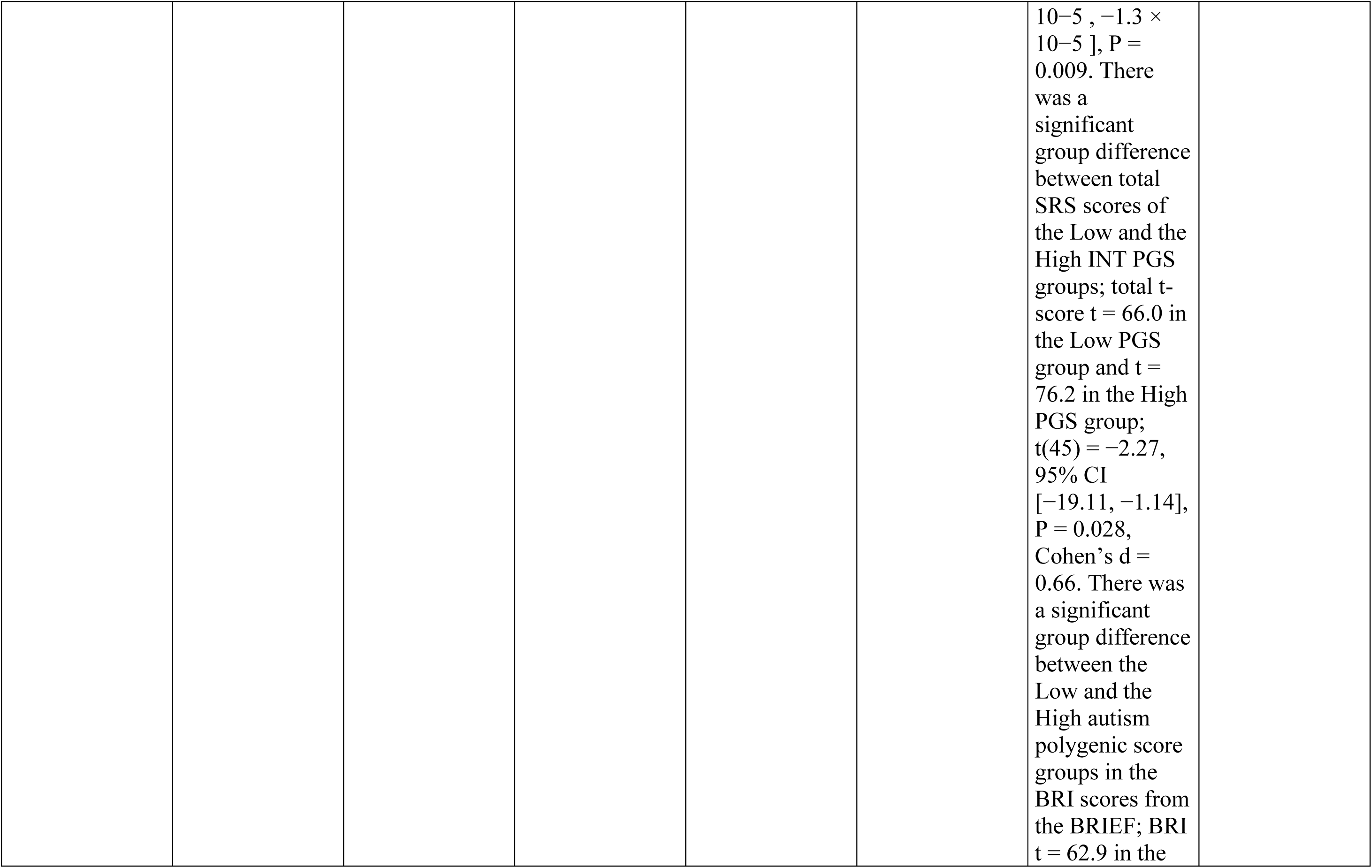
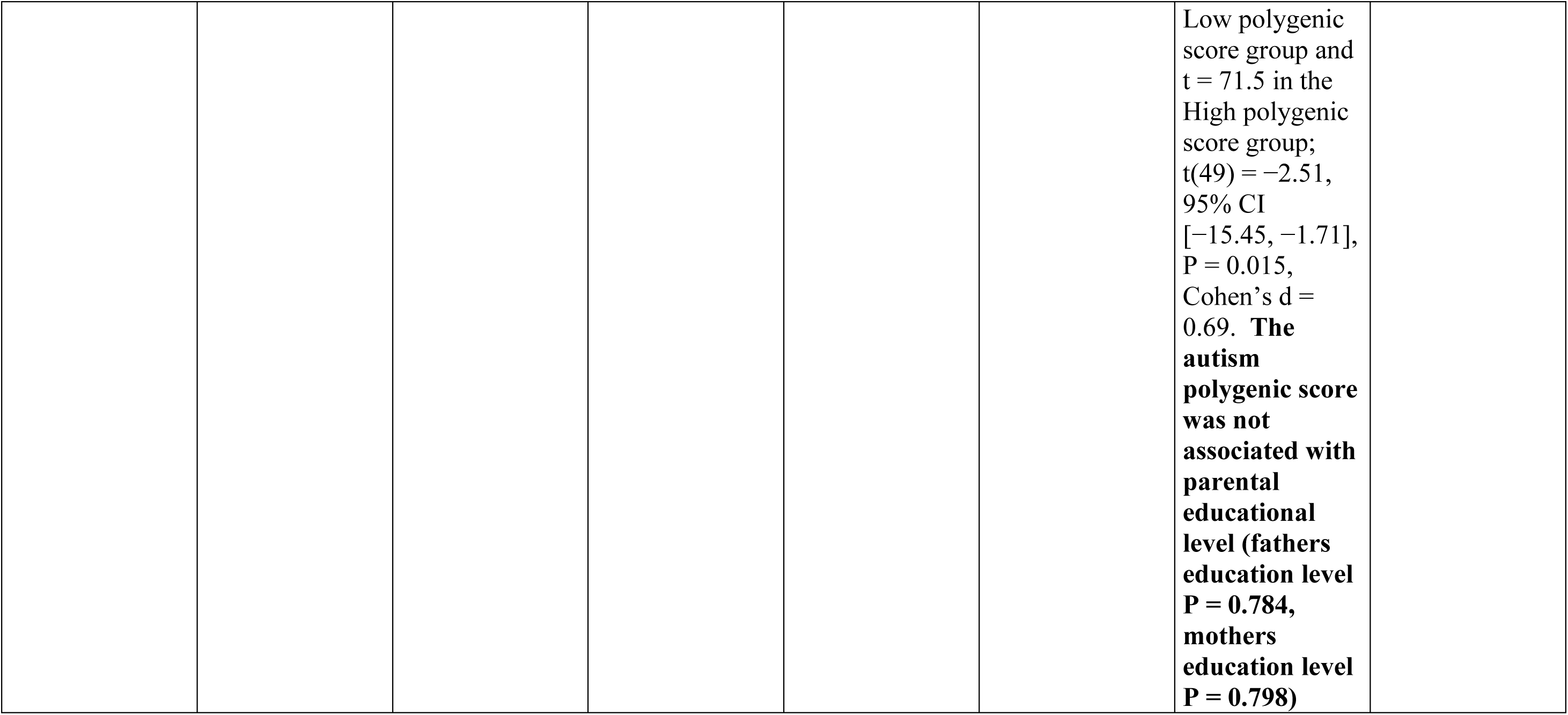
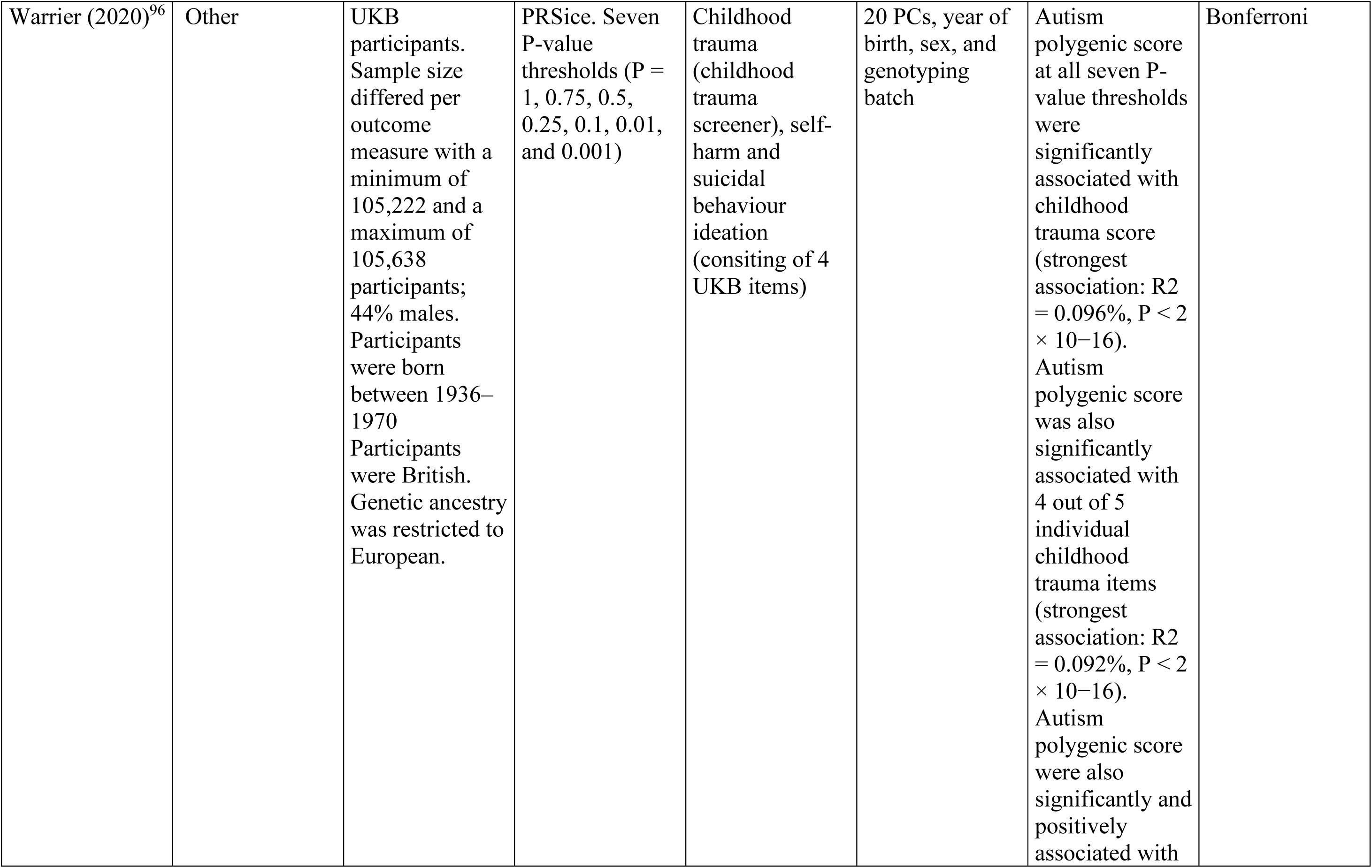
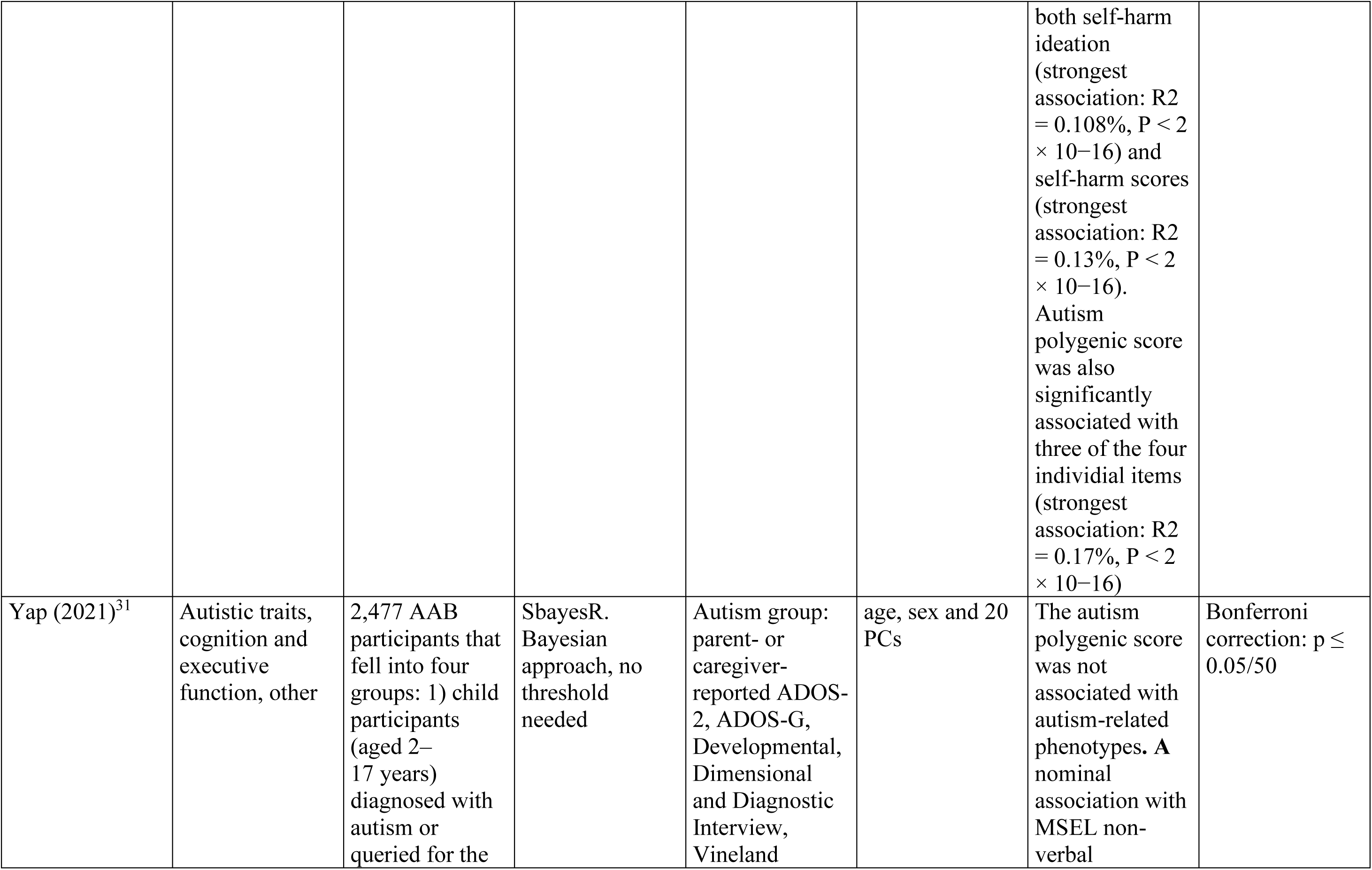
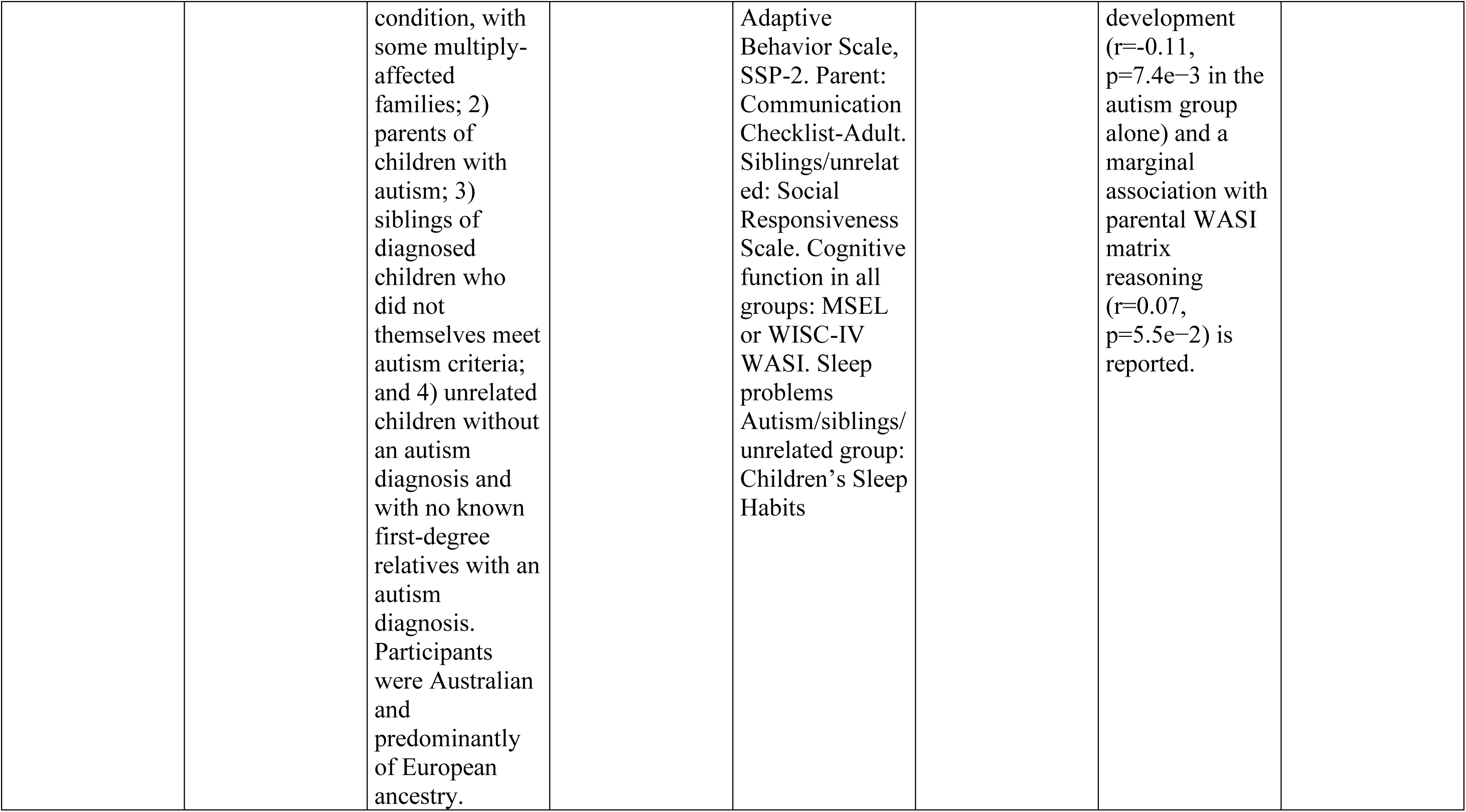
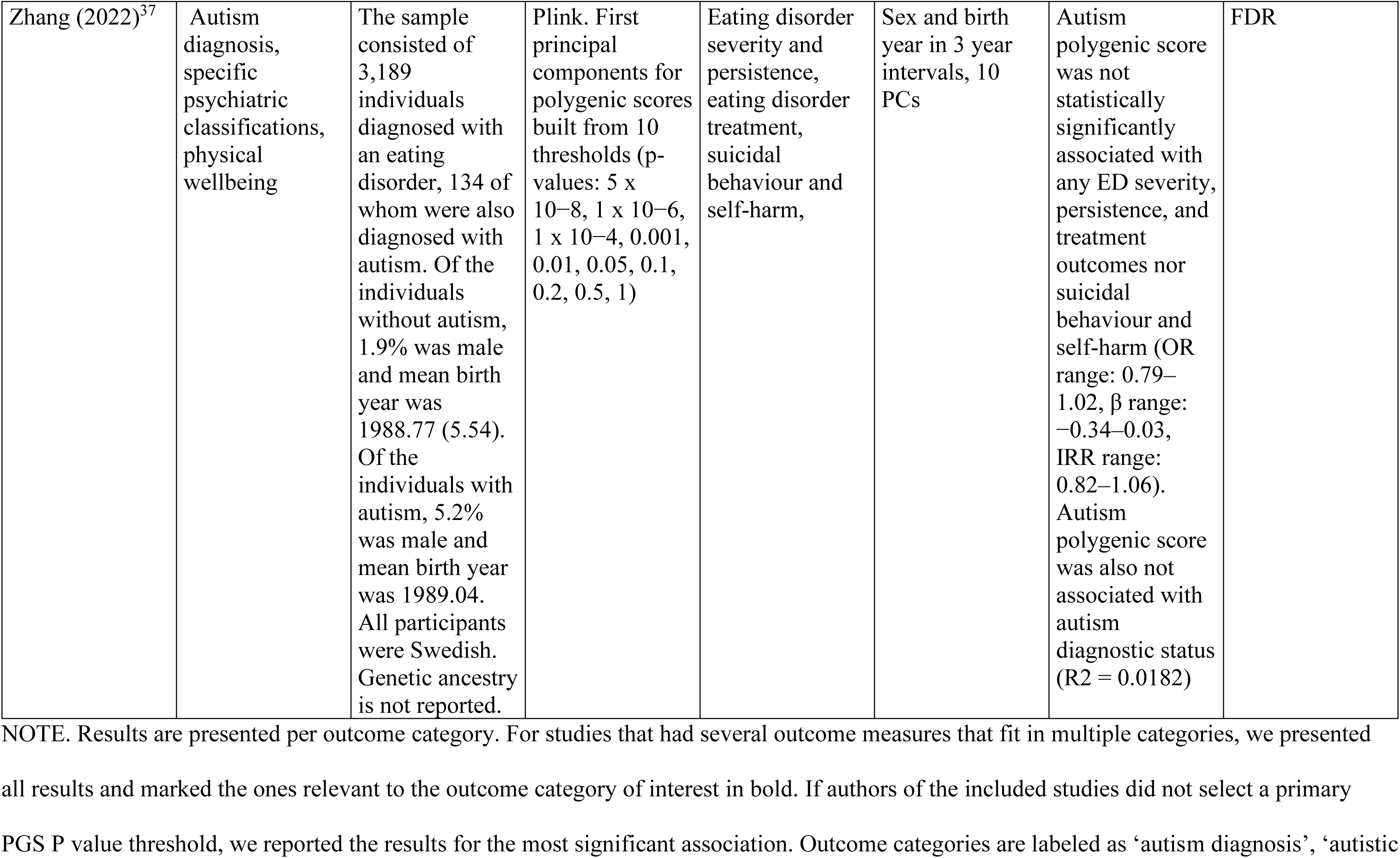
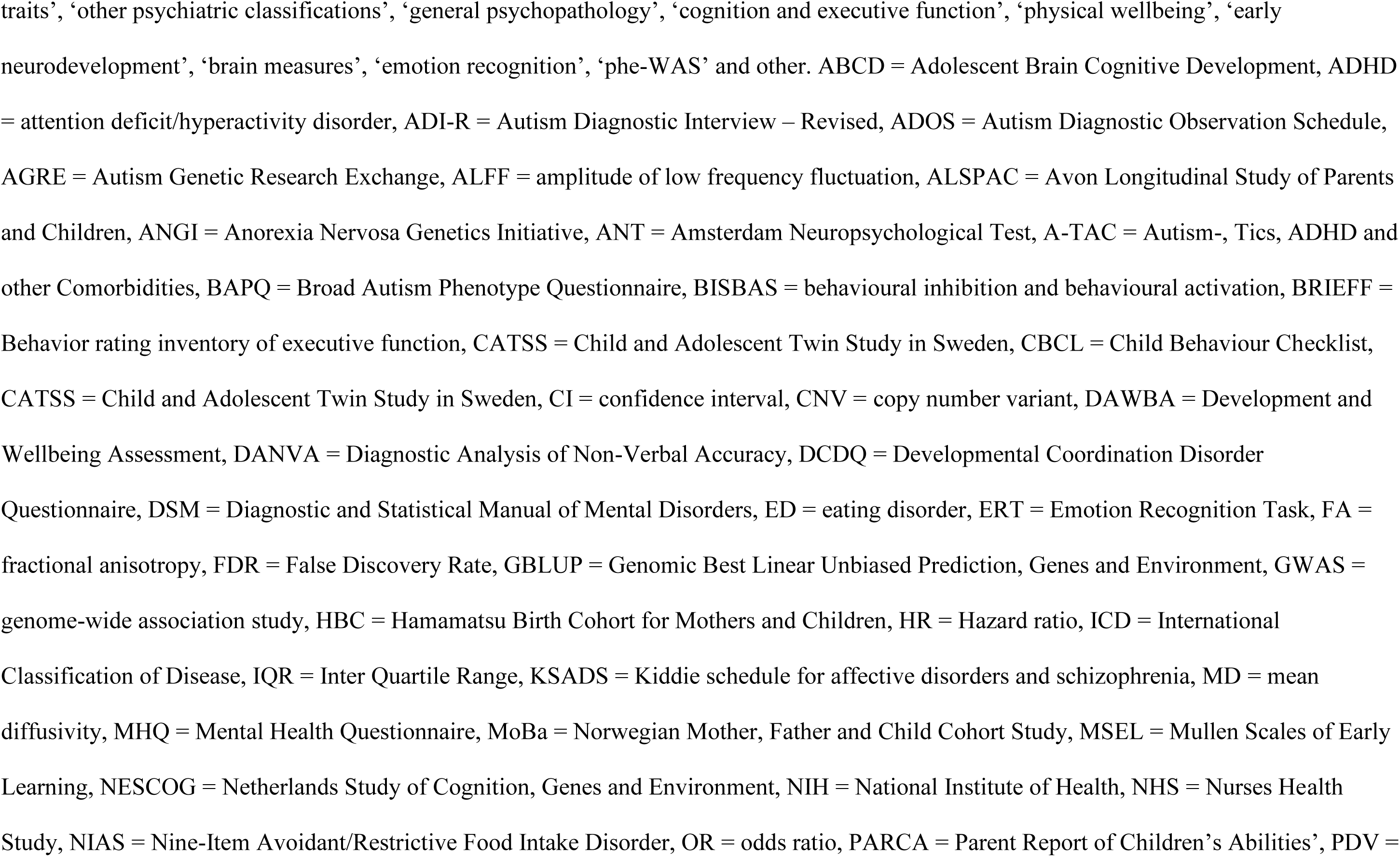
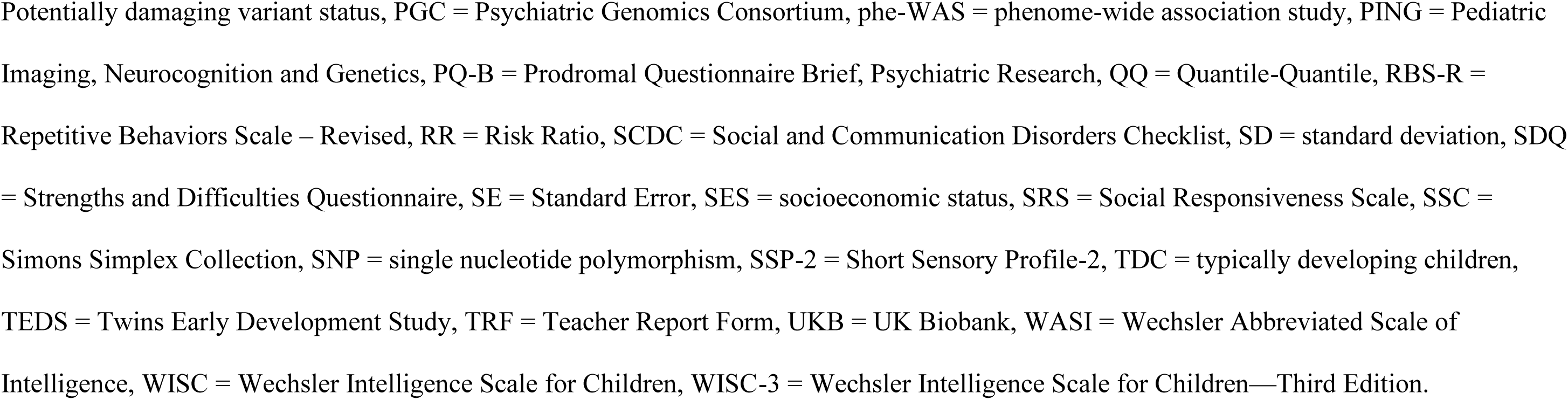
Characteristics and Results of Included Studies on the Association between the Autism Polygenic Score and Outcome Measures.

### Category construction

Outcome categories were constructed through consultation and careful consideration between authors MdW, MM, TJP, SB, AR and AA. They were loosely based on codes from the International Classification of Diseases (ICD) and the World Health Organization International Classification of Functioning, Disability and Health (ICF) and resembled Ronald et al.’s (2021)^18^ category structure, although there were some dissimilarities (e.g., no addiction category, but a category for emotion recognition). This resulted in 11 categories, reflecting the literature that was identified: Autism diagnostic status, autistic traits, other specific psychiatric classifications, general psychopathology, brain measures, cognition and executive function, emotion recognition, early neurodevelopment, physical wellbeing, phenome-wide association studies (phe-WAS) and other. Note that some studies included outcome variables that fit into multiple categories.

### Quality assessment

For 60 studies, 0 biases were detected. Nine studies had one bias, one had two, and one had three biases. We concluded that the quality of included studies is overall good and biases were not clustered in specific outcome categories. The quality assessments are presented in sTable 3, available online.

### Systematic review and meta-analysis

A total of 760 outcome variables were included in the meta-analysis. Rosenthal’s failsafe N ranged from 102 for emotion recognition to 39,921 for autism diagnosis (sTable 4, available online), meaning that 102 to 39,921 null findings would be needed to negate significant findings. Funnel plot tests for asymmetry per outcome category based on standard error were not significant (*p >* .088) except for brain measures (*p* = .006) and autistic traits (*p* < .001), indicating a potential publication bias. Histograms and boxplots of effect sizes, and funnel plots of standard errors are presented in Supplementary Figures 1 – 3, available online. Detailed descriptions of sample characteristics, methods of calculating polygenic scores, GWAS p-value threshold, outcome measures, covariates, results, and multiple testing corrections per study and per outcome category are provided in Table 1. Meta-analytic pooled correlation coefficients ranged from .013 for early neurodevelopment to .162 for autism diagnosis. The number of studies in each category ranged from 3 (phe-was) to 20 (other), with a minimum of 2 and a maximum of 11 independent cohorts per category. A summary of the meta-analysis results for all outcome categories is presented in Table 2 and Figure 2.

**Table 2.** Summary of Multi-level Meta-Analysis Results per Outcome Category

| Outcome category | *n* studies | *n* independent cohorts | *n* estimates | Pooled *r* [95% CI] | *r* range | *P* | *I*<sup>2</sup> |  |  |
| --- | --- | --- | --- | --- | --- | --- | --- | --- | --- |
|  |  |  |  |  |  |  | Total | Between-cohort | Within-cohort |
| Autism diagnosis | 9 | 4 | 10 | .162<br>[.066 – .258] | .030 – .346 | .004 | 99.6% | 28.4% | 71.2% |
| Autistic traits | 11 | 9 | 44 | .042<br>[.004 – .081] | –.074 – .313 | .031 | 94.8% | 87.3% | 7.5% |
| Specific psychiatric classifications | 17 | 12 | 81 | .044<br>[.016 – .073] | –.185 – .213 | .003 | 99.2% | 33.3% | 65.9% |
| General psychopathology | 7 | 4 | 190 | .035<br>[.026 – .044] | –.143 – .292 | <.0001 | 91.5% | 1.0% | 90.5% |
| Cognition and executive function | 9 | 9 | 39 | .042<br>[.008 – .075] | –.338 – .170 | .017 | 94.1% | 88.4% | 5.7% |
| Physical wellbeing | 8 | 8 | 181 | .016<br>[.006 – .026] | –.198 – .166 | .002 | 98.3% | 1.5% | 96.8% |
| Early neurodevelopment | 8 | 7 | 43 | .013<br>[–.038 – .063] | –.110 – .112 | .612 | 94.2% | 93.3% | 0.9% |
| Emotion recognition | 3 | 3 | 5 | .018<br>[ -.034 – .381] | –.229 – .400 | .900 | 99.0% | 36.6% | 62.3% |
| Brain measures | 9 | 9 | 217 | .160<br>[ .031 – .288] | –.364 – .57 | .015 | 99.9% | 80.5% | 19.4% |
| Phe-WAS | 3 | 2 | 26733 | NA | NA | NA | NA | NA | NA |
| Other | 20 | 28 |  | NA | NA | NA | NA | NA | NA |

*NOTE.* *n* studies = number of studies, *n* ind. coh = number of independent cohorts the included studies are based on, *n* est = number of effect size estimates included

**Figure 2.**
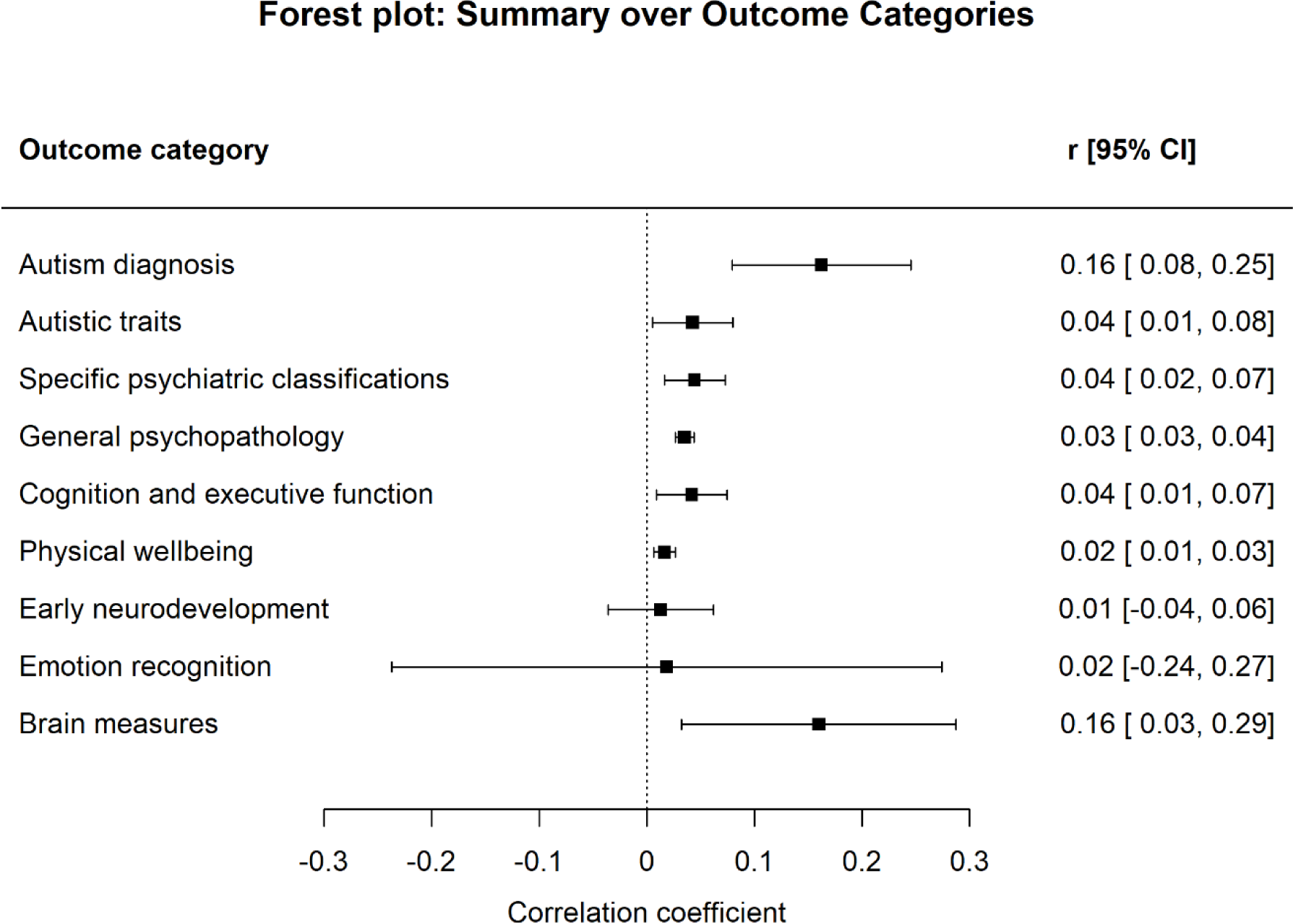
Summary of Multilevel Meta-Analysis over Outcome Categories

#### Autism diagnosis

##### Systematic review

Nine studies assessed the association between the autism polygenic score and autism diagnosis. Of these, seven reported a significant positive association^10,25,27,30,32,33,36^. Two studies, based on smaller samples than the ones that reported significant associations, reported no significant association^37,50^. In addition, the autism polygenic score acted in a dose-dependent manner, where people in higher percentiles on average were more likely to be diagnosed with autism^10,36^.

##### Meta-analysis

Standardized correlation coefficients ranged from 0.030 to 0.346. One outcome was omitted from this range, as it differentiated whether the autism polygenic score distinguished autism diagnosis versus ADHD diagnosis^27^. The overall meta-analyzed effect size was *r* = .162 (95% CI .066 – .258), indicating a small positive association between the autism polygenic score and autism diagnosis. Total *I*^2^ was 99.6%, of which 28.4% between-study and 71.2% within-study. All meta-analysis results for the category of autism diagnosis, including secondary results are presented in Supplementary Figure 4.

#### Autistic traits

##### Systematic review

Eleven studies assessed the association between the autism polygenic score and autistic traits, either in the form of a total score or specific traits such as repetitive behavior, language and social communication. Four of these assessed total scores for autistic traits^40,42,44,45^, one of which was performed in a non-European sample. In children, the autism polygenic score was associated with overall autistic traits in a Japanese^44^ and Swedish sample^45^, but not in a United States (US) sample^40^. In adults, the autism polygenic score was not associated with overall autistic traits^40,42^.

Social behavior was the most commonly studied specific autistic trait, with seven studies assessing the association Four did not find a significant association^31,38,39,45^, and three did^41,43,44^, one of which was a Japanese sample. The autism polygenic score did not associate with restricted and repetitive behavior^38,46^, nor language and communication measures at various ages in childhood^38,45,46^, except for language difficulties at the age of 18 months^38^. In addition, the autism polygenic score was not associated with broad autistic traits as measured by the Broad Autism Phenotype Questionnaire (BAPQ) in children with autism or their parents, except for two significant positive associations with the pragmatic and total scale of the BAPQ in mothers of autistic children^40^.

##### Meta-analysis

One study on autistic traits was omitted from the meta-analysis effect sizes because effect sizes were not reported^40^. Correlation coefficients ranged from −.074 to .313. The overall meta-analyzed correlation coefficient was *r* = .04 (95% CI .004 – .081). Total *I*^2^ was 94.8%, of which 87.3% between-study and 7.5% within-study. Meta-analysis results for autistic traits are presented in Supplementary Figure 5.

##### Specific psychiatric classifications

Eighteen studies assessed the relationship between the autism polygenic score and specific psychiatric classifications (other than autism). Included studies assessed a range of classifications or traits : ADHD (n = 2)^27,34^, ADHD traits (n= 2)^38,42^, depression (n = 2)^48,56^, addiction (n = 1)^49^, suicidal ideation and/or suicide attempt (n = 3)^51,59,97^, eating disorders (n= 2)^37,53^, psychotic spectrum (n= 3)^54,55,57^ and nocturnal enuresis (n = 1)^52^.

One study showed a significant association with ADHD diagnosis^27^, whereas another did not^34^. The autism polygenic score was significantly associated with ADHD symptoms in an east Asian sample^67^, but not in two studies European ancestry samples^38,42^, except for one positive association with inattention and hyperactivity at the age of 8 years. The autism polygenic score did not associate with schizophrenia diagnosis in two East Asian samples^57,58^, and in a European ancestry sample it positively associated with some, but not all specific domains of the psychotic spectrum (e.g. auditory and visual hallucinations)^55^. Two studies found consistent positive associations with lifetime depression^48,56^ and one with depression or anxiety symptoms specifically during pregnancy^48^. No associations were found between the autism polygenic score and disordered eating behavior in two studies^37,53^, except for increased parent-reported Avoidant/restrictive food intake disorder (ARFID;. One study reported no associations between the autism polygenic score and cannabis use disorder^49^, except for an unexpected negative association in people diagnosed with DSM classifications other than schizophrenia. One study reported no association with nocturnal enuresis^52^. Lastly, one study reported that the autism polygenic score was not associated with the case-control status of any psychiatric classification^50^.

One study found that the autism polygenic score distinguished people with autism from those with ADHD^27^.

##### Meta-analysis

Correlation coefficients ranged from −.185 to .213 The overall meta-analyzed correlation coefficient was *r* = .044 (95% CI .016 – .073). Total *I*^2^ was 99.2%, of which 33.9% between-study and 65.4% within-study. Meta-analysis results for specific psychiatric classifications are presented in sFigure 6, and secondary results in sTable 5 and sFigure 7.

### Other outcome categories

Associations between the autism polygenic score and other outcome categories were overall small (median *r* = .03) and inconsistent. Detailed descriptions of these associations are provided in Table 1, Figure 1 and the Supplementary Results and Supplementary Results, available online. Forest plots are presented in sFigures 8 – 13, available online.

### Heterogeneity

We found considerable heterogeneity across all outcome categories (*I*^2^ ≥ 91.5%). To provide a more complete picture of the heterogeneity in the included studies, and because *I*^2^ is not a particularly suitable statistic when assessing very large sample sizes, all meta-analyses forest plots also report Q statistics^98^.

## Discussion

Overall, our results showed that the autism polygenic score was most strongly and most consistently associated with autism diagnosis in independent cohorts. A higher polygenic score translates into an increased likelihood of being diagnosed with autism^10,36^. The association of the autism polygenic score with autistic traits was significant but smaller. Associations with outcomes other than autism diagnosis and autistic traits were inconsistent and meta-analytic effect sizes were generally small. Here we discuss our results, specifically zooming in on autism diagnosis, autistic traits and other psychiatric classifications because these categories contained the most consistent results. We discuss implications and provide recommendations for future directions.

The lack of consistent associations with outcomes other than autism diagnosis and autistic traits is in contrast with a vast body of literature supporting phenotypic associations between autism and neurodevelopmental, psychiatric, and physical conditions and other traits, as well as genetic associations found in twin studies. They are also in contrast with studies that used alternative genetic methodologies such as genome-wide restricted maximum likelihood (GREML), genome-wide complex trait analysis (GCTA), and linkage disequilibrium score (LDSC) regression. A difference between polygenic scores and these methods is that the latter captures the total influence of measured SNPs whereas polygenic scores do not, potentially resulting in less power to detect genetic associations (explanations of these methods are described in Martin et al.^12^). Our results also highlight disparities between the autism polygenic score and the ADHD polygenic score, with the latter showing stronger (e.g., OR between 1.22% and 1.76% for ADHD diagnosis) and more reliable, consistent associations. This is despite comparable heritability estimates and GWAS sample sizes for ADHD and autism, and also both being a neurodevelopmental condition^18^.

The overarching category of specific psychiatric classifications comprised numerous classifications and had an overall significant but small association with the autism polygenic score. However, results varied markedly for different classifications. Our secondary analyses to dissecting the overarching category revealed a significant association with the psychotic spectrum, but not with ADHD. These findings were in contrast with some past studies in this area^10,99–102^.

The inconsistency of our results and their lack of alignment with previous findings indicates that the current autism polygenic score may lack the statistical power to adequately capture shared polygenic effects. In line with this notion, two included studies found that the autism polygenic score acts largely independent from psychiatric family history^36,61^, despite both factors being expected to reflect a genetic predisposition to autism. This finding extends beyond autism to multiple psychiatric classifications^103,104^, indicating that current psychiatric polygenic scores may overall insufficiently capture genetic propensity towards psychiatric traits. As the predictive power of polygenic scores depends heavily on the statistical power of the discovery GWAS, we underscore the importance of ongoing efforts to increase GWAS sample sizes.

Our study is not without limitations. First, by focusing on univariate associations and excluding genetic measures and methods other than polygenic scores, we may have overlooked important insights into autism genetics. We refer the reader to Akingbuwa et al. (2022)^99^ for results that are based on other methods than univariate associations^99^. Second, we were not able to include all the associations in our meta-analyses because some studies did not report all effect sizes in text. Similarly, we did not include all outcome measures assessed through phenome-wide association studies in our meta-analysis, as this was infeasible due to the number of studies. Lastly, we point out the considerable heterogeneity in our included studies, as is reflected in the high *I*^2^ statistics. A high *I*^2^ value could be due to low within-sample estimation error, which is likely to occur with large sample sizes such as the ones included in our study^105^. However, we acknowledge that this heterogeneity may also be due to the construction of our outcome variable categories. However, even in the categories that are largely similar in their outcomes (such as autism diagnosis), heterogeneity is high. This heterogeneity remained substantial even when dissecting outcome categories into more homogeneous subcategories. This heterogeneity may have added noise to our estimates. Future studies should aim to reduce such heterogeneity to enhance our understanding of autism’s polygenic nature.

A specific source of heterogeneity may be the substantial variation in polygenic score calculation, both in terms of software and the selection of the GWAS *p*-value thresholds. While the majority of studies apply methods that select the polygenic score that yields the strongest associations, others use a pre-defined threshold or employ principal component analyses to construct a polygenic score from multiple thresholds. Similarly, a diverse set of polygenic score calculation programs was applied. Several recent reports address this issue and advocate for standardized guidelines and reporting practices^106,107^, which might improve the comparability of polygenic score findings.

Despite the marked diversity in methods between studies, we observed limited diversity in the participant characteristics. The majority of studies were based on people of European ancestry, resulting in our current understanding of the autism polygenic score being almost solely based on people of European ancestry. This observation is concerning, especially considering the increasing autism prevalence rates in people from non-European backgrounds^108^. Similarly, few included studies addressed understudied subgroups of autism, such as those with intellectual disability, or people with autism that are nonverbal. Both scientifically, and ethically, it would be of great value to radically improve inclusive research practices^109^.

We propose that GWASs, and subsequently polygenic scores, based on individual autistic traits instead of the overarching autism diagnosis might enhance our understanding of autism genetics^110^. Early twin studies revealed that the three domains of autistic traits (social, communication, and restricted/repetitive behavior) only modestly correlate genetically, a finding that suggests largely independent genetic effects may affect different autistic traits^111,112^. Yet molecular genetics have not consistently pursued this research direction^96^. Although some molecular genetic studies were performed on specific autistic traits, large-scale approaches are still lacking^46,113^.

Despite its limitations, the current autism polygenic score comprises part of a bigger picture, and should ideally, as suggested by several recent papers, be integrated with other genetic factors such as common and rare variants, psychiatric family history, and sex, in a comprehensive approach to studying autism^25,36,61,114^. For example, Grove et al. (2019)^10^ found that a combined polygenic score of autism and correlated traits, such as major depressive disorder and schizophrenia, improved the prediction of autism diagnostic status. Rare genetic variants are not captured by typical polygenic scores, but have been suggested as factors of high influence on autism diagnostic status^115^. In line, Antaki et al. (2022)^114^ showed that a genetic score containing a combination of common and rare genetic factors significantly increased statistical power over separate common and rare variant scores. Schendel et al. (2022)^36^ argue that psychiatric family history and polygenic scores only minimally correlate and should therefore be considered separate, mostly uncorrelated, measures of autism’s genetic etiology. Lastly, although we do not find significant sex differences in the association of the autism polygenic score with outcome variables, there is building evidence for a different genetic makeup for men and women with autism. Autistic women on average have a higher number of genetic mutations^116^, higher mean polygenic load^87,114^ and more rare copy number variants (CNV) load^117^. However, a potentially male-biased clinical view on the manifestation of autism might explain diagnostic differences, and subsequently observed genetic differences^118^

In conclusion, we show that the autism polygenic score as a standalone variable associates consistently with autism diagnostic status and dimensional autistic traits, but is unable to capture the spectrum’s complex phenotypical and etiological differences and its genetic overlap with other traits and conditions. We propose considering the current autism polygenic score as a complementary measure in research, with improvements needed for a more robust understanding of autism’s polygenic underpinnings.

## Data Availability

All data extracted for this study are available upon reasonable request to the authors.

## Acknowledgements

We thank our EGAL (Environmental and Genetic Influences on Autism across the Lifespan) panel for their valuable input.

## Footnotes

1* We use a combination of identity-first and person-first language to accommodate to everyone’s preferences^5,6^.

## References

1. Bottema-Beutel K, Kapp SK, Lester JN, Sasson NJ, Hand BN. Avoiding Ableist Language: Suggestions for Autism Researchers. Autism Adulthood. 2021;3(1):18–29. doi:10.1089/aut.2020.0014

2. Zeidan J, Fombonne E, Scorah J, et al. Global prevalence of autism: A systematic review update. Autism Res. 2022;15(5):778–790. doi:10.1002/aur.2696

3. Tick B, Bolton P, Happé F, Rutter M, Rijsdijk F. Heritability of autism spectrum disorders: a meta-analysis of twin studies. J Child Psychol Psychiatry. 2016;57(5):585–595. doi:10.1111/jcpp.12499

4. Martini MI, Kuja-Halkola R, Butwicka A, et al. Sex Differences in Mental Health Problems and Psychiatric Hospitalization in Autistic Young Adults. JAMA Psychiatry. 2022;79(12):1188–1198. doi:10.1001/jamapsychiatry.2022.3475

5. Buijsman R, Begeer S, Scheeren AM. ‘Autistic person’ or ‘person with autism’? Person-first language preference in Dutch adults with autism and parents. Autism. 2023;27(3):788–795. doi:10.1177/13623613221117914

6. Taboas A, Doepke K, Zimmerman C. Preferences for identity-first versus person-first language in a US sample of autism stakeholders. Autism. 2023;27(2):565–570. doi:10.1177/13623613221130845

7. Bougeard C, Picarel-Blanchot F, Schmid R, Campbell R, Buitelaar J. Prevalence of Autism Spectrum Disorder and Co-morbidities in Children and Adolescents: A Systematic Literature Review. Front Psychiatry. 2021;12. Accessed September 18, 2023. https://www.frontiersin.org/articles/10.3389/fpsyt.2021.744709

8. Gidziela A, Ahmadzadeh YI, Michelini G, et al. A meta-analysis of genetic effects associated with neurodevelopmental disorders and co-occurring conditions. Nat Hum Behav. 2023;7(4):642–656. doi:10.1038/s41562-023-01530-y

9. Tick B, Colvert E, McEwen F, et al. Autism Spectrum Disorders and Other Mental Health Problems: Exploring Etiological Overlaps and Phenotypic Causal Associations. J Am Acad Child Adolesc Psychiatry. 2016;55(2):106–113.e4. doi:10.1016/j.jaac.2015.11.013

10. Grove J, Ripke S, Als TD, et al. Identification of common genetic risk variants for autism spectrum disorder. Nat Genet 2019 513. 2019;51(3):431–444. doi:10.1038/s41588-019-0344-8

11. Willoughby EA, Polderman TJC, Boutwell BB. Behavioural genetics methods. Nat Rev Methods Primer. 2023;3(1):1–16. doi:10.1038/s43586-022-00191-x

12. Martin AR, Daly MJ, Robinson EB, Hyman SE, Neale BM. Predicting Polygenic Risk of Psychiatric Disorders. Biol Psychiatry. 2019;86(2):97–109. doi:10.1016/j.biopsych.2018.12.015

13. Wray NR, Lin T, Austin J, et al. From Basic Science to Clinical Application of Polygenic Risk Scores: A Primer. JAMA Psychiatry. 2021;78(1):101–109. doi:10.1001/jamapsychiatry.2020.3049

14. Polderman TJC, Benyamin B, de Leeuw CA, et al. Meta-analysis of the heritability of human traits based on fifty years of twin studies. Nat Genet. 2015;47(7):702–709. doi:10.1038/ng.3285

15. Du Rietz E, Coleman J, Glanville K, Choi SW, O’Reilly PF, Kuntsi J. Association of Polygenic Risk for Attention-Deficit/Hyperactivity Disorder With Co-occurring Traits and Disorders. Biol Psychiatry Cogn Neurosci Neuroimaging. 2018;3(7):635–643. doi:10.1016/j.bpsc.2017.11.013

16. Green A, Baroud E, DiSalvo M, Faraone SV, Biederman J. Examining the impact of ADHD polygenic risk scores on ADHD and associated outcomes: A systematic review and meta-analysis. J Psychiatr Res. 2022;155:49–67. doi:10.1016/j.jpsychires.2022.07.032

17. Mistry S, Harrison JR, Smith DJ, Escott-Price V, Zammit S. The use of polygenic risk scores to identify phenotypes associated with genetic risk of schizophrenia: Systematic review. Schizophr Res. 2018;197:2–8. doi:10.1016/j.schres.2017.10.037

18. Ronald A, de Bode N, Polderman TJC. Systematic Review: How the Attention-Deficit/Hyperactivity Disorder Polygenic Risk Score Adds to Our Understanding of ADHD and Associated Traits. J Am Acad Child Adolesc Psychiatry. 2021;60(10):1234–1277. doi:10.1016/j.jaac.2021.01.019

19. Hayden JA, Côté P, Bombardier C. Evaluation of the Quality of Prognosis Studies in Systematic Reviews. Ann Intern Med. 2006;144(6):427–437. doi:10.7326/0003-4819-144-6-200603210-00010

20. Hayden JA, van der Windt DA, Cartwright JL, Côté P, Bombardier C. Assessing Bias in Studies of Prognostic Factors. Ann Intern Med. 2013;158(4):280–286. doi:10.7326/0003-4819-158-4-201302190-00009

21. Viechtbauer W. Conducting Meta-Analyses in R with the metafor Package. J Stat Softw. 2010;36:1–48. doi:10.18637/jss.v036.i03

22. Assink M, Wibbelink CJM. Fitting three-level meta-analytic models in R: A step-by-step tutorial. Quant Methods Psychol. 2016;12(3):154–174. doi:10.20982/tqmp.12.3.p154

23. Harrer M, Cuijpers P, Furukawa T, Ebert D. Doing Meta-Analysis with R: A Hands-on Guide. Chapman and Hall/CRC; 2021. Accessed December 20, 2023. https://www.taylorfrancis.com/books/mono/10.1201/9781003107347/meta-analysis-mathias-harrer-pim-cuijpers-toshi-furukawa-david-ebert

24. Rosenthal R. The file drawer problem and tolerance for null results. Psychol Bull. 1979;86(3):638–641. doi:10.1037/0033-2909.86.3.638

25. Klei L, McClain LL, Mahjani B, et al. How rare and common risk variation jointly affect liability for autism spectrum disorder. Mol Autism. 2021;12(1):1–13. doi:10.1186/S13229-021-00466-2/FIGURES/5

26. Klein L, D’Urso S, Eapen V, Hwang LD, Lin PI. Exploring polygenic contributors to subgroups of comorbid conditions in autism spectrum disorder. Sci Rep. 2022;12(1):3416. doi:10.1038/s41598-022-07399-7

27. Mattheisen M, Grove J, Als TD, et al. Identification of shared and differentiating genetic architecture for autism spectrum disorder, attention-deficit hyperactivity disorder and case subgroups. Nat Genet. 2022;54(10):1470–1478. doi:10.1038/s41588-022-01171-3

28. Morneau-Vaillancourt G, Andlauer TFM, Ouellet-Morin I, et al. Polygenic scores differentially predict developmental trajectories of subtypes of social withdrawal in childhood. J Child Psychol Psychiatry. 2021;62(11):1320–1329. doi:10.1111/jcpp.13459

29. Riglin L, Tobarra-Sanchez E, Stergiakouli E, et al. Early manifestations of genetic liability for ADHD, autism and schizophrenia at ages 18 and 24 months. JCPP Adv. 2022;2(3):e12093. doi:10.1002/jcv2.12093

30. Trost B, Thiruvahindrapuram B, Chan AJS, et al. Genomic architecture of autism from comprehensive whole-genome sequence annotation. Cell. 2022;185(23):4409–4427.e18. doi:10.1016/j.cell.2022.10.009

31. Yap CX, Alvares GA, Henders AK, et al. Analysis of common genetic variation and rare CNVs in the Australian Autism Biobank. Mol Autism. 2021;12(1):1–17. doi:10.1186/S13229-020-00407-5/FIGURES/3

32. Debost JCPG, Thorsteinsson E, Trabjerg B, et al. Genetic and psychosocial influence on the association between early childhood infections and later psychiatric disorders. Acta Psychiatr Scand. 2022;146(5):406–419. doi:10.1111/acps.13491

33. Hannon E, Schendel D, Ladd-Acosta C, et al. Elevated polygenic burden for autism is associated with differential DNA methylation at birth. Genome Med. 2018;10(1):19. doi:10.1186/s13073-018-0527-4

34. Jansen AG, Dieleman GC, Jansen PR, Verhulst FC, Posthuma D, Polderman TJC. Psychiatric Polygenic Risk Scores as Predictor for Attention Deficit/Hyperactivity Disorder and Autism Spectrum Disorder in a Clinical Child and Adolescent Sample. Behav Genet. 2020;50(4):203–212. doi:10.1007/S10519-019-09965-8/FIGURES/1

35. Demontis D, Walters RK, Martin J, et al. Discovery of the first genome-wide significant risk loci for attention deficit/hyperactivity disorder. Nat Genet. 2019;51(1):63–75. doi:10.1038/s41588-018-0269-7

36. Schendel D, Munk Laursen T, Albiñana C, et al. Evaluating the interrelations between the autism polygenic score and psychiatric family history in risk for autism. Autism Res. 2022;15(1):171–182. doi:10.1002/AUR.2629

37. Zhang R, Birgegård A, Fundín B, et al. Association of autism diagnosis and polygenic scores with eating disorder severity. Eur Eat Disord Rev. 2022;30(5):442–458. doi:10.1002/erv.2941

38. Askeland RB, Hannigan LJ, Ask H, et al. Early manifestations of genetic risk for neurodevelopmental disorders. J Child Psychol Psychiatry. 2021;63(7):810–819. doi:10.1111/JCPP.13528

39. Li D, Choque-Olsson N, Jiao H, et al. The influence of common polygenic risk and gene sets on social skills group training response in autism spectrum disorder. Npj Genomic Med 2020 51. 2020;5(1):1–8. doi:10.1038/s41525-020-00152-x

40. Nayar K, Sealock JM, Maltman N, et al. Elevated Polygenic Burden for Autism Spectrum Disorder Is Associated With the Broad Autism Phenotype in Mothers of Individuals With Autism Spectrum Disorder. Biol Psychiatry. 2021;89(5):476–485. doi:10.1016/J.BIOPSYCH.2020.08.029

41. Reed ZE, Mahedy L, Jackson A, et al. Examining the bidirectional association between emotion recognition and social autistic traits using observational and genetic analyses. J Child Psychol Psychiatry. 2021;62(11):1330–1338. doi:10.1111/JCPP.13395

42. Riglin L, Leppert B, Langley K, et al. Investigating attention-deficit hyperactivity disorder and autism spectrum disorder traits in the general population: What happens in adult life? J Child Psychol Psychiatry. 2021;62(4):449–457. doi:10.1111/JCPP.13297

43. Serdarevic F, Tiemeier H, Jansen PR, et al. Polygenic Risk Scores for Developmental Disorders, Neuromotor Functioning During Infancy, and Autistic Traits in Childhood. Biol Psychiatry. 2020;87(2):132–138. doi:10.1016/J.BIOPSYCH.2019.06.006

44. Takahashi N, Harada T, Nishimura T, et al. Association of Genetic Risks With Autism Spectrum Disorder and Early Neurodevelopmental Delays Among Children Without Intellectual Disability. JAMA Netw Open. 2020;3(2):1921644. doi:10.1001/jamanetworkopen.2019.21644

45. Taylor MJ, Martin J, Lu Y, et al. Association of Genetic Risk Factors for Psychiatric Disorders and Traits of These Disorders in a Swedish Population Twin Sample. JAMA Psychiatry. 2019;76(3):280–289. doi:10.1001/JAMAPSYCHIATRY.2018.3652

46. Thomas TR, Koomar T, Casten LG, Tener AJ, Bahl E, Michaelson JJ. Clinical autism subscales have common genetic liabilities that are heritable, pleiotropic, and generalizable to the general population. Transl Psychiatry. 2022;12(1):247. doi:10.1038/s41398-022-01982-2

47. Torske T, Nærland T, Bettella F, et al. Autism spectrum disorder polygenic scores are associated with every day executive function in children admitted for clinical assessment. Autism Res Off J Int Soc Autism Res. 2020;13(2):207–220. doi:10.1002/AUR.2207

48. Havdahl A, Wootton RE, Leppert B, et al. Associations Between Pregnancy-Related Predisposing Factors for Offspring Neurodevelopmental Conditions and Parental Genetic Liability to Attention-Deficit/Hyperactivity Disorder, Autism, and Schizophrenia: The Norwegian Mother, Father and Child Cohort Study (MoBa). JAMA Psychiatry. 2022;79(8):799. doi:10.1001/jamapsychiatry.2022.1728

49. Hjorthøj C, Uddin MJ, Wimberley T, et al. No evidence of associations between genetic liability for schizophrenia and development of cannabis use disorder. Psychol Med. 2021;51(3):479–484. doi:10.1017/S0033291719003362

50. Jansen AG, Jansen PR, Savage JE, et al. The predictive capacity of psychiatric and psychological polygenic risk scores for distinguishing cases in a child and adolescent psychiatric sample from controls. J Child Psychol Psychiatry. 2021;62(9):1079–1089. doi:10.1111/JCPP.13370

51. Joo YY, Moon SY, Wang HH, et al. Association of Genome-Wide Polygenic Scores for Multiple Psychiatric and Common Traits in Preadolescent Youths at Risk of Suicide. JAMA Netw Open. 2022;5(2):e2148585. doi:10.1001/jamanetworkopen.2021.48585

52. Jørgensen CS, Horsdal HT, Rajagopal VM, et al. Identification of genetic loci associated with nocturnal enuresis: a genome-wide association study. Lancet Child Adolesc Health. 2021;5(3):201–209. doi:10.1016/S2352-4642(20)30350-3

53. Koomar T, Thomas TR, Pottschmidt NR, Lutter M, Michaelson JJ. Estimating the Prevalence and Genetic Risk Mechanisms of ARFID in a Large Autism Cohort. Front Psychiatry. 2021;12:849. doi:10.3389/FPSYT.2021.668297/BIBTEX

54. Legge SE, Jones HJ, Kendall KM, et al. Association of Genetic Liability to Psychotic Experiences With Neuropsychotic Disorders and Traits. JAMA Psychiatry. 2019;76(12):1256–1265. doi:10.1001/JAMAPSYCHIATRY.2019.2508

55. Legge SE, Cardno AG, Allardyce J, et al. Associations Between Schizophrenia Polygenic Liability, Symptom Dimensions, and Cognitive Ability in Schizophrenia. JAMA Psychiatry. 2021;78(10):1143–1151. doi:10.1001/JAMAPSYCHIATRY.2021.1961

56. Leppert B, Havdahl A, Riglin L, et al. Association of Maternal Neurodevelopmental Risk Alleles With Early-Life Exposures. JAMA Psychiatry. 2019;76(8):834–842. doi:10.1001/JAMAPSYCHIATRY.2019.0774

57. Ohi K, Nishizawa D, Shimada T, et al. Polygenetic Risk Scores for Major Psychiatric Disorders Among Schizophrenia Patients, Their First-Degree Relatives, and Healthy Participants. Int J Neuropsychopharmacol. 2020;23(3):157–164. doi:10.1093/IJNP/PYZ073

58. Qin Y, Kang J, Jiao Z, et al. Polygenic risk for autism spectrum disorder affects left amygdala activity and negative emotion in schizophrenia. Transl Psychiatry 2020 101. 2020;10(1):1-12. doi:10.1038/s41398-020-01001-2

59. Russell AE, Hemani G, Jones HJ, et al. An exploration of the genetic epidemiology of non-suicidal self-harm and suicide attempt. BMC Psychiatry. 2021;21(1):207. doi:10.1186/S12888-021-03216-Z/FIGURES/2

60. Gui Y, Zhou X, Wang Z, et al. Sex-specific genetic association between psychiatric disorders and cognition, behavior and brain imaging in children and adults. Transl Psychiatry. 2022;12(1):1–8. doi:10.1038/s41398-022-02041-6

61. Loughnan RJ, Palmer CE, Makowski C, et al. Unique prediction of developmental psychopathology from genetic and familial risk. J Child Psychol Psychiatry. 2022;63(12):1631–1643. doi:10.1111/jcpp.13649

62. Pat N, Riglin L, Anney R, et al. Motivation and Cognitive Abilities as Mediators Between Polygenic Scores and Psychopathology in Children. J Am Acad Child Adolesc Psychiatry. 2022;61(6):782–795.e3. doi:10.1016/j.jaac.2021.08.019

63. Riglin L, Thapar AK, Leppert B, et al. Using Genetics to Examine a General Liability to Childhood Psychopathology. Behav Genet. 2020;50(4):213–220. doi:10.1007/S10519-019-09985-4/TABLES/2

64. Schlag F, Allegrini AG, Buitelaar J, et al. Polygenic risk for mental disorder reveals distinct association profiles across social behaviour in the general population. Mol Psychiatry. 2022;27(3):1588–1598. doi:10.1038/s41380-021-01419-0

65. Waszczuk MA, Miao J, Docherty AR, et al. General v. specific vulnerabilities: polygenic risk scores and higher-order psychopathology dimensions in the Adolescent Brain Cognitive Development (ABCD) Study. Psychol Med. Published online 2021:1–10. doi:10.1017/S0033291721003639

66. Aguilar-Lacasaña S, Vilor-Tejedor N, Jansen PR, et al. Polygenic risk for ADHD and ASD and their relation with cognitive measures in school children. Psychol Med. 2022;52(7):1356–1364. doi:10.1017/S0033291720003189

67. Chang S, Yang L, Wang Y, Faraone SV. Shared polygenic risk for ADHD, executive dysfunction and other psychiatric disorders. Transl Psychiatry 2020 101. 2020;10(1):1-9. doi:10.1038/s41398-020-00872-9

68. Cullen H, Selzam S, Dimitrakopoulou K, Plomin R, Edwards AD. Greater genetic risk for adult psychiatric diseases increases vulnerability to adverse outcome after preterm birth. Sci Rep 2021 111. 2021;11(1):1–8. doi:10.1038/s41598-021-90045-5

69. Hughes A, Wade KH, Dickson M, et al. Common health conditions in childhood and adolescence, school absence, and educational attainment: Mendelian randomization study. Npj Sci Learn 2021 61. 2021;6(1):1-9. doi:10.1038/s41539-020-00080-6

70. Price KM, Wigg KG, Feng Y, et al. Genome-wide association study of word reading: Overlap with risk genes for neurodevelopmental disorders. Genes Brain Behav. 2020;19(6). doi:10.1111/GBB.12648

71. Dennison CA, Legge SE, Bracher-Smith M, et al. Association of genetic liability for psychiatric disorders with accelerometer-assessed physical activity in the UK Biobank. PLOS ONE. 2021;16(3):e0249189. doi:10.1371/JOURNAL.PONE.0249189

72. Hunjan AK, Hübel C, Lin Y, Eley TC, Breen G. Association between polygenic propensity for psychiatric disorders and nutrient intake. Commun Biol 2021 41. 2021;4(1):1–9. doi:10.1038/s42003-021-02469-4

73. Niarchou M, Singer EV, Straub P, Malow BA, Davis LK. Investigating the genetic pathways of insomnia in Autism Spectrum Disorder. Res Dev Disabil. 2022;128:104299. doi:10.1016/j.ridd.2022.104299

74. Ohi K, Ochi R, Noda Y, et al. Polygenic risk scores for major psychiatric and neurodevelopmental disorders contribute to sleep disturbance in childhood: Adolescent Brain Cognitive Development (ABCD) Study. Transl Psychiatry. 2021;11(1):187. doi:10.1038/s41398-021-01308-8

75. Werner MCF, Wirgenes KV, Shadrin A, et al. Immune marker levels in severe mental disorders: associations with polygenic risk scores of related mental phenotypes and psoriasis. Transl Psychiatry. 2022;12(1):38. doi:10.1038/s41398-022-01811-6

76. Hannigan LJ, Askeland RB, Ask H, et al. Developmental milestones in early childhood and genetic liability to neurodevelopmental disorders. Psychol Med. 2023;53(5):1750–1758. doi:10.1017/S0033291721003330

77. Fish LA, Nyström P, Gliga T, et al. Development of the pupillary light reflex from 9 to 24 months: association with common autism spectrum disorder (ASD) genetic liability and 3-year ASD diagnosis. J Child Psychol Psychiatry. 2021;62(11):1308–1319. doi:10.1111/jcpp.13518

78. Gui A, Mason L, Gliga T, et al. Look duration at the face as a developmental endophenotype: elucidating pathways to autism and ADHD. Dev Psychopathol. 2020;32(4):1303–1322. doi:10.1017/S0954579420000930

79. Portugal AM, Taylor MJ, Viktorsson C, et al. Pupil size and pupillary light reflex in early infancy: heritability and link to genetic liability to schizophrenia. J Child Psychol Psychiatry. 2022;63(9):1068–1077. doi:10.1111/jcpp.13564

80. Waddington F, Franke B, Hartman C, Buitelaar JK, Rommelse N, Mota NR. A polygenic risk score analysis of ASD and ADHD across emotion recognition subtypes ADHD, ASD, emotion recognition, polygenic risk score, subtyping. Am J Med Genet. 2021;186:401–411. doi:10.1002/ajmg.b.32818

81. Alemany S, Blok E, Jansen PR, Muetzel RL, White T. Brain morphology, autistic traits, and polygenic risk for autism: A population-based neuroimaging study. Autism Res. 2021;14(10):2085–2099. doi:10.1002/aur.2576

82. Ecker C, Pretzsch CM, Bletsch A, et al. Interindividual Differences in Cortical Thickness and Their Genomic Underpinnings in Autism Spectrum Disorder. Am J Psychiatry. 2022;179(3):242–254. doi:10.1176/appi.ajp.2021.20050630

83. Gui A, Meaburn EL, Tye C, Charman T, Johnson MH, Jones EJH. Association of Polygenic Liability for Autism With Face-Sensitive Cortical Responses From Infancy. JAMA Pediatr. 2021;175(9):968. doi:10.1001/jamapediatrics.2021.1338

84. Jansen PR, Muetzel RL, Polderman TJC, et al. Polygenic Scores for Neuropsychiatric Traits and White Matter Microstructure in the Pediatric Population. Biol Psychiatry Cogn Neurosci Neuroimaging. 2019;4(3):243–250. doi:10.1016/j.bpsc.2018.07.010

85. Lawrence KE, Hernandez LM, Fuster E, et al. Impact of autism genetic risk on brain connectivity: a mechanism for the female protective effect. Brain. 2022;145(1):378–387. doi:10.1093/brain/awab204

86. Mason L, Moessnang C, Chatham C, et al. Stratifying the autistic phenotype using electrophysiological indices of social perception. Sci Transl Med. 2022;14(658):eabf8987. doi:10.1126/scitranslmed.abf8987

87. Sha Z, Schijven D, Francks C. Patterns of brain asymmetry associated with polygenic risks for autism and schizophrenia implicate language and executive functions but not brain masculinization. Mol Psychiatry. 2021;26(12):7652–7660. doi:10.1038/s41380-021-01204-z

88. Leppert B, Millard LAC, Riglin L, et al. A cross-disorder PRS-pheWAS of 5 major psychiatric disorders in UK Biobank. Zhu X, ed. PLOS Genet. 2020;16(5):e1008185. doi:10.1371/journal.pgen.1008185

89. Wendt FR, Carvalho CM, Pathak GA, Gelernter J, Polimanti R. Polygenic risk for autism spectrum disorder associates with anger recognition in a neurodevelopment-focused phenome-wide scan of unaffected youths from a population-based cohort. Williams SM, ed. PLOS Genet. 2020;16(9):e1009036. doi:10.1371/journal.pgen.1009036

90. Kalman JL, Loohuis LMO, Vreeker A, et al. Characterisation of age and polarity at onset in bipolar disorder. Br J Psychiatry. 2021;219(6):659–669. doi:10.1192/BJP.2021.102

91. Khundrakpam B, Vainik U, Gong J, et al. Neural correlates of polygenic risk score for autism spectrum disorders in general population. Brain Commun. 2020;2(2):fcaa092. doi:10.1093/braincomms/fcaa092

92. Maxwell J, Coleman J, Breen G, psychiatry EVJ, 2021 undefined. Association between genetic risk for psychiatric disorders and the probability of living in urban settings. *jamanetwork.com*. Accessed January 4, 2023. https://jamanetwork.com/journals/jamapsychiatry/article-abstract/2785027?casa_token=1R2Ny3LXULsAAAAA:JYE0YLpJAFn5twgh5CPVA4rYANrax0NF3NQk6ji2tGoymlKGCjNVpfSnnfDgyHKNVZkKDa2h2ps

93. Peel AJ, Purves KL, Baldwin JR, et al. Genetic and early environmental predictors of adulthood self-reports of trauma. Br J Psychiatry. 2022;221(4):613–620. doi:10.1192/bjp.2021.207

94. Ratanatharathorn A, Koenen KC, Chibnik LB, Weisskopf MG, Rich-Edwards JW, Roberts AL. Polygenic risk for autism, attention-deficit hyperactivity disorder, schizophrenia, major depressive disorder, and neuroticism is associated with the experience of childhood abuse. Mol Psychiatry. 2021;26(5):1696–1705. doi:10.1038/s41380-020-00996-w

95. Strom NI, Smit DJA, Silzer T, et al. Meta-analysis of genome-wide association studies of hoarding symptoms in 27,651 individuals. Transl Psychiatry. 2022;12(1):1–8. doi:10.1038/s41398-022-02248-7

96. Warrier V, Toro R, Won H, et al. Social and non-social autism symptoms and trait domains are genetically dissociable. Commun Biol. 2019;2(1):1–13. doi:10.1038/s42003-019-0558-4

97. Warrier V, Baron-Cohen S. Childhood trauma, life-time self-harm, and suicidal behaviour and ideation are associated with polygenic scores for autism. Mol Psychiatry. 2021;26(5):1670–1684. doi:10.1038/s41380-019-0550-x

98. Cochran WG. Some Methods for Strengthening the Common χ2 Tests. Biometrics. 1954;10(4):417–451. doi:10.2307/3001616

99. Akingbuwa WA, Hammerschlag AR, Bartels M, Middeldorp CM. Systematic Review: Molecular Studies of Common Genetic Variation in Child and Adolescent Psychiatric Disorders. J Am Acad Child Adolesc Psychiatry. 2022;61(2):227–242. doi:10.1016/j.jaac.2021.03.020

100. Lee PH, Anttila V, Won H, et al. Genomic Relationships, Novel Loci, and Pleiotropic Mechanisms across Eight Psychiatric Disorders. Cell. 2019;179(7):1469–1482.e11. doi:10.1016/j.cell.2019.11.020

101. Ronald A, Happé F, Plomin R. A twin study investigating the genetic and environmental aetiologies of parent, teacher and child ratings of autistic-like traits and their overlap. Eur Child Adolesc Psychiatry. 2008;17(8):473–483. doi:10.1007/S00787-008-0689-5/FIGURES/2

102. Taylor MJ, Robinson EB, Happé F, Bolton P, Freeman D, Ronald A. A longitudinal twin study of the association between childhood autistic traits and psychotic experiences in adolescence. Mol Autism. 2015;6(1):44. doi:10.1186/s13229-015-0037-9

103. Agerbo E, Sullivan PF, Vilhjálmsson BJ, et al. Polygenic Risk Score, Parental Socioeconomic Status, Family History of Psychiatric Disorders, and the Risk for Schizophrenia: A Danish Population-Based Study and Meta-analysis. JAMA Psychiatry. 2015;72(7):635–641. doi:10.1001/jamapsychiatry.2015.0346

104. Agerbo E, Trabjerg BB, Børglum AD, et al. Risk of Early-Onset Depression Associated With Polygenic Liability, Parental Psychiatric History, and Socioeconomic Status. JAMA Psychiatry. 2021;78(4):387–397. doi:10.1001/jamapsychiatry.2020.4172

105. Higgins JPT, Thompson SG. Quantifying heterogeneity in a meta-analysis. Stat Med. 2002;21(11):1539–1558. doi:10.1002/sim.1186

106. Pain O, Glanville KP, Hagenaars SP, et al. Evaluation of polygenic prediction methodology within a reference-standardized framework. PLOS Genet. 2021;17(5):e1009021. doi:10.1371/journal.pgen.1009021

107. Wand H, Lambert SA, Tamburro C, et al. Improving reporting standards for polygenic scores in risk prediction studies. Nature. 2021;591(7849):211-219. doi:10.1038/s41586-021-03243-6

108. Liu BM, Paskov K, Kent J, et al. Racial and Ethnic Disparities in Geographic Access to Autism Resources Across the US. JAMA Netw Open. 2023;6(1):e2251182. doi:10.1001/jamanetworkopen.2022.51182

109. Martin AR, Kanai M, Kamatani Y, Okada Y, Neale BM, Daly MJ. Clinical use of current polygenic risk scores may exacerbate health disparities. Nat Genet. 2019;51(4):584–591. doi:10.1038/s41588-019-0379-x

110. Arenella M, Cadby G, De Witte W, et al. Potential role for immune-related genes in autism spectrum disorders: Evidence from genome-wide association meta-analysis of autistic traits. Autism. 2022;26(2):361–372. doi:10.1177/13623613211019547

111. Happé F, Ronald A. The ‘Fractionable Autism Triad’: A Review of Evidence from Behavioural, Genetic, Cognitive and Neural Research. Neuropsychol Rev. 2008;18(4):287–304. doi:10.1007/s11065-008-9076-8

112. Ronald A, Happé F, Bolton P, et al. Genetic heterogeneity between the three components of the autism spectrum: a twin study. J Am Acad Child Adolesc Psychiatry. 2006;45(6):691–699. doi:10.1097/01.chi.0000215325.13058.9d

113. Taylor SC, Steeman S, Gehringer BN, et al. Heritability of quantitative autism spectrum traits in adults: A family-based study. Autism Res. 2021;14(8):1543–1553. doi:10.1002/aur.2571

114. Antaki D, Guevara J, Maihofer AX, et al. A phenotypic spectrum of autism is attributable to the combined effects of rare variants, polygenic risk and sex. Nat Genet. 2022;54(9):1284–1292. doi:10.1038/s41588-022-01064-5

115. Buxbaum JD. Multiple rare variants in the etiology of autism spectrum disorders. Dialogues Clin Neurosci. 2009;11(1):35–43. doi:10.31887/DCNS.2009.11.1/jdbuxbaum

116. Jacquemont S, Coe BP, Hersch M, et al. A Higher Mutational Burden in Females Supports a “Female Protective Model” in Neurodevelopmental Disorders. Am J Hum Genet. 2014;94(3):415–425. doi:10.1016/j.ajhg.2014.02.001

117. Sanders SJ, He X, Willsey AJ, et al. Insights into Autism Spectrum Disorder Genomic Architecture and Biology from 71 Risk Loci. Neuron. 2015;87(6):1215–1233. doi:10.1016/j.neuron.2015.09.016

118. Kreiser NL, White SW. ASD in Females: Are We Overstating the Gender Difference in Diagnosis? Clin Child Fam Psychol Rev. 2014;17(1):67–84. doi:10.1007/s10567-013-0148-9

