## Supplemental Materials for "A Systematic Review and Meta-Analysis: Research Using the Autism Polygenic Score"

**Supplementary methods**

Quality assessment criteria.

| Criteria |
|---|

*1. Study participation; Study sample adequately represents the population of interest*

(A) Description of the key characteristics of the study population (distribution by age, gender and ancestry/ethnicity)

(B) The sampling frame and recruitment are described, including characteristics of the place of recruitment or authors clearly reference where this information can be found

(C) Inclusion and exclusion criteria are described or authors clearly reference where this information can be found

(D) Information about participation at baseline and potential attrition (for genetic data) are described or authors clearly reference where this information can be found

*2. Predictor measurement; autism polygenic score is adequately measured*

(E) Description of genetic data collection (e.g., blood, saliva) and genotyping (array) is provided, and target sample was not part of GWAS

(F) Genetic data were subject to adequate quality control (minor allele frequency, missing rate, relatedness participants, sex mismatch, and genotype quality), an up to date imputation method and an established reference panel was used

(G) The autism polygenic score is adequately calculated (e.g., pruning/clumping of SNPs), and the p-value threshold for calculating the autism polygenic score is reported.

3. Outcome measurement; Outcome of interest is measured in a similar way for all participants

(H) A clear definition of the outcome measures is provided

(I) Several indications are provided for the validity and reliability of the outcome measure, or a reference is provided.

(J) The method and setting of outcome measurement is the same for all study participants

4. Confounding measurement; Important potential confounders are appropriately accounted for

(K) Age, gender and Socio Economic Status are accounted for in the analysis

(L) Population stratification and potential batch effects are accounted for in the analysis

(M) In case of clinical samples, treatment and comorbidity are accounted for in the analyses

*5. Analysis and data presentation; Statistical analysis is appropriate*

(N) Sufficient presentation of the data to assess the adequacy of the analytic strategy

(O) The number of participants in the target sample supports sufficient statistical power (N > 400)

(P) The selected statistical model is adequate for the design of the study

(Q) There is no evidence of selective reporting of results, and proper correction for multiple testing was applied.

---

#### Supplementary results

Abbreviations:

ANT: Attention Network Test

BISBAS: Behavioral Inhibition and Behavioral Activation

BRIEF: Behavior Rating Inventory of Executive Function

CBCL: Child Behaviour Checklist

DAWBA: Development and Wellbeing Assessment

GBI: General Behavior Inventory

GSCE: General Certificate of Secondary Education

KSADS: Kiddie Schedule for Affective Disorders and Schizophrenia

NIH: National Institute of Health

OCD: Obsessive Compulsive Disorder

PARCA: Parent Report of Children's Abilities

SCDC: Social and Communication Disorders Checklist

SDQ: Strengths and Difficulties Questionnaire

UPPS-P: Impulsive Behavior Scale

WASI: Wechsler Abbreviated Scale of Intelligence

WISC: Wechsler Intelligence Scale for Children

WRAT: Wide Range Achievement Test

#### Primary results

##### General psychopathology

*Systematic review.* Seven studies assessed the association between the autism polygenic score and general psychopathology[1–7]. This category included studies that used general mental health questionnaires to assess general psychopathology, either by using the p-factor (the p factor reflects a general propensity towards psychiatric diagnoses) or by analysing total scores and subscale scores from these questionnaires.

*Overall liability for mental health issues.* Two studies reported a non-significant association with the Child Behaviour Checklist (CBCL[8]) total scale or the KSADS total scale[1,2], and one reported a significant association[7]. Two studies did not report an association with p-factor constructed from the CBCL[3] or a p-factor constructed from the DAWBA and SCDC [4].

*Questionnaire (sub)scales.* Included studies used the CBCL[8], the GBI[9], a three-item prosocial behaviour survey, the UPPS-P[10], the BISBAS[11], KSADS[12] and the SDQ[13].

For the CBCL, patterns of significant associations appear to be random: Gui et al. (2022)[1] and Thomas et al. (2022)[6] do not report significant associations with any CBCL subscales. Loughnan et al. (2022) only report an association with the caregiver reported inattention subscale of the CBCL, whereas Waszczuk et al. (2021)[7] find several associations with factors composed from the CBCL; externalizing, neurodevelopmental and detachment.

For the KSADS, Loughnan et al. (2022)[2] report an association with the self-reported depression symptoms in a mixed-ancestry sample and caregiver reported OCD symptoms in a non-European sample, self-reported depression, and suicidality symptoms, and caregiver reported ADHD. No association with other subscales was reported.

For the SDQ, In the ALSPAC sample, Schlag et al. (2022)[5] reported only one association with parent reported low prosociality in 7 year-olds, but not with any other parent or teacher reported low prosociality or peer problems. In the TEDS sample however, they do find several significant associations (low prosociality at age 7 and 11, peer problems at age 7, 9, 12 and PR peer problems at age 4, 7, 11).

*Meta-analysis.* Standardized beta coefficients ranged from -.143 to .292. The overall meta-analysed correlation coefficient was $r$ = .035 (95% CI .03 - .04). Total $I^2$ was 91.5%, of which 1.0% between-study and 90.5% within-study. Meta-analysis results for general psychopathology, are presented in Supplementary Figure 8.

Cognition and executive function

*Systematic review.* Nine studies assessed the association between the autism polygenic score and cognition and executive function[1,2,14–20]. Included studies used the NIH toolbox cognition battery [21], WISC, WASI, WRAT3, WRAT4, BRIEF, educational attainment (GSCE), PARCA, Stroop test, Trail-making test, digit span test, *n* back test, and ANT. Only 4 out of 40 measures were significant (10%). Three were a positive association with crystallized memory in samples of different ancestries[2]. However, another study did not support this association in an overlapping sample[1]. The other association was a negative association with behaviour regulation measured using BRIEF[19]

*Meta-analysis.* Standardized correlation coefficients ranged from -.338 to .170. The overall meta-analyzed correlation coefficient was $r$ = .042 (95% CI -.008 - .075). Total $I^2$ was 94.1%, of which 88.4% between-study and 5.7% within-study. Meta-analysis results for cognition and executive function, are presented in Supplementary Figure 9.

Physical wellbeing

*Systematic review.* Eight studies assessed the association between the autism polygenic score and physical wellbeing[22–29]. Physical wellbeing included phenotypes such as activity levels, general health and health before and during pregnancy, nutrient intake, smoking and alcohol consumption, sleep problems, BMI and immune marker levels. Only 5 out of 181 (2,8%) outcome measures were significantly associated with the autism polygenic score. This included a negative association with overall activity levels[22] and the level of immune marker sIL-2R[28].

*Meta-analysis.* Standardized correlation coefficients ranged from -.198 to .166. The overall meta-analyzed effect size was $r$ = .016 (95% CI .006 - .026). Total $I^2$ was 98.3%, of which 1.5% between-study and 96.8% within-study. Meta-analysis results for physical wellbeing, are presented in Supplementary Figure 10.

Early neurodevelopment

*Systematic review.* The category of early neurodevelopment included nine studies on eye tracking measures (n = 3) and other neurodevelopmental traits such as motor development and temperament[20,30–37].. Nine out of 43 tested associations were reported as significant; motor difficulties at age 3[30], age at first walking[33], overall neuromotor development and overall muscle tone and low and high muscle tone[36], gross motor skills and receptive language development[37].

*Meta-analysis.* Standardized correlation coefficients ranged from -.110 to .112. The overall meta-analyzed correlation coefficient was $r$ = .013 (95% CI -.038 – .066). Total $I^2$ was 94.2%, of which 93.3% between-study and 0.9% within-study. Meta-analysis results for early neurodevelopment are presented in Supplementary Figure 11.

Emotion recognition

*Systematic review.* Three studies assessed the association between the autism polygenic score and emotion recognition. Reed et al. (2020)[38] do not report a significant association with emotion recognition in healthy participants. Qin et al. (2020)[39], on the other hand, do report a negative association of autism polygenic score with recognition of negative emotions and total emotion recognition, but not positive or neutral emotions. Waddington et al. (2021)[40] studied how autism polygenic score relates to speed and accuracy of visual and auditory emotion recognition, and only find that it associates with faster visual emotion recognition.

*Meta-analysis.* Standardized beta coefficients ranged from -.229 to .400. The overall meta-analyzed correlation coefficient was $r$ = .18 (95% CI -.034 - .381).Total $I^2$ was 99.0%, of which 36.6% between-study and 62.3% within-study. Meta-analysis results for emotion recognition are presented in Supplementary Figure 12.

Brain measures

*Systematic review:* Nine studies assessed the association between the autism polygenic score and brain measures. This category included MRI and EEG measures. The autism polygenic score was significantly positively associated with an accumulated measure of neuroanatomical atypicality for cortical thickness[41], shorter N290 latency to face vs nonface stimuli[42], increased salience network connectivity with the postcentral gyrus in autistic and typically developing youth (significant sex differences were observed[43]), functional annotations related to language, executive functions and autism[44], longer N170 latency to face response[45], and higher amplitude of low frequency fluctuation in the left amygdala in schizophrenia cases and controls[39]. The autism polygenic score was not associated with cortical measures of autism-related brain regions, including surface area, thickness, and subcortical volume and gyrification measurements[44,46], nor with resting-state fMRI or diffusion tensor imaging (DTI) measurements[1], and neither with global and tract-specific fractional anisotropy and mean diffusivity[47].

*Meta-analysis:* One study was excluded from the meta-analysis since effect sizes were not reported (Gui et al., 2022). Standardized regression coefficients ranged from -.364 to .57. The overall meta-analyzed effect size was $r$ = .160 (95% CI .031 − .288). Total $I^2$ was 99.9% of which 80.5% between-study and 19.4% within-study. Meta-analysis results for brain measures are presented in Supplementary Figure 13.

Phe-WAS

*Systematic review.* Three studies applied a phenome-wide association approach[44,48,49], where a large number of outcomes is tested for its association with the autism polygenic score in a similar way genetic variants are tested for an association with an outcome in GWAS. From the 3238 tested variables by Sha et al. (2021)[44], four showed a significant association with autism polygenic score (hearing difficulty/problem with background noise, Townsend deprivation index at recruitment, Qualifications: College or University degree and Long-standing illness, disability or infirmity). In the same sample of residents of the UK but using a much larger number of outcomes (23004), Leppert et al. (2020)[48] find 10 significant associations, most of which were related to physical health (blood measures, body size, lung function), and some related to mental health (nervousness) and socio-demography. Wendt et al. (2020)[49] applied phe-was in a US based sample and specifically focused on 491 neurodevelopmental outcomes, and found a significant association with recognition of angry faces and nominal associations with other emotion recognition phenotypes.

*Meta-analysis.* No meta-analysis was performed considering the outcome variables in this category were too extensive.

Other

*Systematic review.* In the 21 studies with uncategorized outcome measures, autism polygenic score was negatively associated with having potentially damaging (rare) genetic variants[50], age at onset of bipolar disorder[51] and age at onset of schizophrenia (in a Japanese sample[52]), and was positively associated with female sex[44], non-righthandedness[44], paternal age[23], comorbid conditions related to allergies[53], higher population density in adult life and moving from rural areas to cities[54], increased Deoxyribonucleic acid (DNA) methylation[55], childhood trauma[56,57], experiencing physical/emotional abuse and physical assault[58]. No associations were reported with age[59], age at first parental concern and age of autism diagnosis[20], polarity at onset of bipolar disorder[51], sleep disturbance in children[27], gestational age[16], vocabulary[59], reading[59], parental educational level[19], several outcomes related to eating disorders (e.g. age of diagnosis, lowest BMI during adulthood, ever been in inpatient or outpatient care; Zhang et al., 2022), use of depression or anxiety medication[23], maternal age[23], response to social skills group training[60], trajectories of social wariness and preference for solitude[61], adulthood trauma[56], sexual abuse[58].

Some studies reported inconsistent associations. Autism polygenic score was negatively associated with the odds of childhood infections in people in the third quartile of the polygenic score, but there was no association in the first, second and fourth quartile[62]. The autism polygenic score was positively associated with hoarding symptoms in some, but not all samples included in Strom et al. (2022)[63], and the meta-analyzed result was only significant when heterogeneity between samples was not accounted for.

*Meta-analysis.* No meta-analysis was performed considering the outcome variables in this category were too diverse.

**Secondary results**

Systematic review: Sex differences

Sex differences were assessed in 14 of the 72 included studies. Overall, there is little evidence for sex differences in associations of autism polygenic score with outcomes[14,35,36,44,54,54,64]. Yet some differences were reported; the autism polygenic score had a stronger association in boys for repetitive behavior, social communication[30] and age at first walking[33]. The autism polygenic score had a stronger association in women for childhood trauma[57], sameness [6], and psychiatric behavior [1]. Some studies assessed the influences of autism polygenic score in mothers and fathers[23,65], but they reported no differences in the associations.

Meta-analysis: Population differences in polygenic score association with autism diagnosis

Based on recent work that pointed out how polygenic score accuracy may vary not only between genetic ancestries, but within ancestries too[66], we performed secondary analyses assessing whether the polygenic score association differed between Europe and US-based samples. For EU-based samples, the association between autism polygenic score and autism diagnosis was $r$ = .20 (95% CI .12 – .28) whereas the association for US-based samples was $r$ = .12 (95% .02 – .21). A test for subgroup differences revealed no significant difference between these populations, $Q$ = 2.77 df = 2, $p$ = .25. Meta-analysis results for autism diagnosis, including subgroup analyses, are presented in Figure 2.

Meta-analysis: Analyses for Subclassifications within Specific Psychiatric Classifications

Our secondary results show that the autism polygenic score only significantly associates with the subclassification ‘psychotic spectrum’, but not ADHD, eating disorders, or self-harm and suicide ideation, see sFigure 7.

#### Supplementary Tables

sTable 1. PRISMA checklist.

| Section and Topic | Item # | Checklist item | Location where item is reported |
|---|---|---|---|
| **TITLE** | | | |
| Title | 1 | Identify the report as a systematic review. | Title page |
| **ABSTRACT** | | | |
| Abstract | 2 | See the PRISMA 2020 for Abstracts checklist. | Main manuscript, P1 |
| **INTRODUCTION** | | | |
| Rationale | 3 | Describe the rationale for the review in the context of existing knowledge. | Main manuscript, P2-P3 |
| Objectives | 4 | Provide an explicit statement of the objective(s) or question(s) the review addresses. | Main manuscript, P3 |
| **METHODS** | | | |
| Eligibility criteria | 5 | Specify the inclusion and exclusion criteria for the review and how studies were grouped for the syntheses. | Main manuscript, P4 |
| Information sources | 6 | Specify all databases, registers, websites, organisations, reference lists and other sources searched or consulted to identify studies. Specify the date when each source was last searched or consulted. | Main manuscript, P4 |
| Search | 7 | Present the full search strategies for all databases, registers and websites, including any filters and limits used. | Supplemental |

| Section and Topic | Item # | Checklist item | Location where item is reported |
|---|---|---|---|
| strategy | | | material, sTable 2 |
| Selection process | 8 | Specify the methods used to decide whether a study met the inclusion criteria of the review, including how many reviewers screened each record and each report retrieved, whether they worked independently, and if applicable, details of automation tools used in the process. | Main manuscript, P4 |
| Data collection process | 9 | Specify the methods used to collect data from reports, including how many reviewers collected data from each report, whether they worked independently, any processes for obtaining or confirming data from study investigators, and if applicable, details of automation tools used in the process. | Main manuscript, P4 |
| Data items | 10a | List and define all outcomes for which data were sought. Specify whether all results that were compatible with each outcome domain in each study were sought (e.g. for all measures, time points, analyses), and if not, the methods used to decide which results to collect. | Main manuscript, P4 |
| | 10b | List and define all other variables for which data were sought (e.g. participant and intervention characteristics, funding sources). Describe any assumptions made about any missing or unclear information. | Main manuscript, P4 |
| Study risk of bias assessment | 11 | Specify the methods used to assess risk of bias in the included studies, including details of the tool(s) used, how many reviewers assessed each study and whether they worked independently, and if applicable, details of automation tools used in the process. | Main manuscript, P5 |
| Effect measures | 12 | Specify for each outcome the effect measure(s) (e.g. risk ratio, mean difference) used in the synthesis or presentation of results. | Main manuscript, Table 1 |
| Synthesis methods | 13a | Describe the processes used to decide which studies were eligible for each synthesis (e.g. tabulating the study intervention characteristics and comparing against the planned groups for each synthesis (item #5)). | Main manuscript, P4 |
| | 13b | Describe any methods required to prepare the data for presentation or synthesis, such as handling of missing summary statistics, or data conversions. | Main manuscript, P6 |

| Section and Topic | Item # | Checklist item | Location where item is reported |
|---|---|---|---|
| | 13c | Describe any methods used to tabulate or visually display results of individual studies and syntheses. | Main manuscript, P6 |
| | 13d | Describe any methods used to synthesize results and provide a rationale for the choice(s). If meta-analysis was performed, describe the model(s), method(s) to identify the presence and extent of statistical heterogeneity, and software package(s) used. | Main manuscript, P6 |
| | 13e | Describe any methods used to explore possible causes of heterogeneity among study results (e.g. subgroup analysis, meta-regression). | Main manuscript, P6 |
| | 13f | Describe any sensitivity analyses conducted to assess robustness of the synthesized results. | Main manuscript, P7 |
| Reporting bias assessment | 14 | Describe any methods used to assess risk of bias due to missing results in a synthesis (arising from reporting biases). | Main manuscript, P6 |
| Certainty assessment | 15 | Describe any methods used to assess certainty (or confidence) in the body of evidence for an outcome. | - |
| **RESULTS** | | | |
| Study selection | 16a | Describe the results of the search and selection process, from the number of records identified in the search to the number of studies included in the review, ideally using a flow diagram. | Main manuscript, Figure 1 |
| | 16b | Cite studies that might appear to meet the inclusion criteria, but which were excluded, and explain why they were excluded. | Main manuscript, P5 |
| Study characteristics | 17 | Cite each included study and present its characteristics. | Main manuscript, |

| Section and Topic | Item # | Checklist item | Location where item is reported |
|---|---|---|---|
| | | | Table 1 |
| Risk of bias in studies | 18 | Present assessments of risk of bias for each included study. | Supplemental Material, sTable 3 |
| Results of individual studies | 19 | For all outcomes, present, for each study: (a) summary statistics for each group (where appropriate) and (b) an effect estimate and its precision (e.g. confidence/credible interval), ideally using structured tables or plots. | Main manuscript, Table 1 |
| Results of syntheses | 20a | For each synthesis, briefly summarise the characteristics and risk of bias among contributing studies. | Main manuscript, Table 2 and Supplemental Material, sTable 3 |
| | 20b | Present results of all statistical syntheses conducted. If meta-analysis was done, present for each the summary estimate and its precision (e.g. confidence/credible interval) and measures of statistical heterogeneity. If comparing groups, describe the direction of the effect. | Main manuscript, P136-141 and Supplemental Material, Results |
| | 20c | Present results of all investigations of possible causes of heterogeneity among study results. | Main manuscript, Table 2 and Supplemental Material, results and sFigure 4 and |

| Section and Topic | Item # | Checklist item | Location where item is reported |
|---|---|---|---|
| | | | 7 |
| | 20d | Present results of all sensitivity analyses conducted to assess the robustness of the synthesized results. | - |
| Reporting biases | 21 | Present assessments of risk of bias due to missing results (arising from reporting biases) for each synthesis assessed. | Main manuscript, P136-137 and Supplemental Material, sTable 4 |
| Certainty of evidence | 22 | Present assessments of certainty (or confidence) in the body of evidence for each outcome assessed. | - |
| **DISCUSSION** | | | |
| Discussion | 23a | Provide a general interpretation of the results in the context of other evidence. | Main manuscript, P142-143 |
| | 23b | Discuss any limitations of the evidence included in the review. | Main manuscript, P143-144 |
| | 23c | Discuss any limitations of the review processes used. | Main manuscript, P143-144 |
| | 23d | Discuss implications of the results for practice, policy, and future research. | Main manuscript, P144-146 |
| **OTHER INFORMATION** | | | |
| Registration | 24a | Provide registration information for the review, including register name and registration number, or state that the | Main |

| Section and Topic | Item # | Checklist item | Location where item is reported |
|---|---|---|---|
| and protocol | | review was not registered. | manuscript, P4 |
| | 24b | Indicate where the review protocol can be accessed, or state that a protocol was not prepared. | Main manuscript, P4 |
| | 24c | Describe and explain any amendments to information provided at registration or in the protocol. | Main manuscript, P4 |
| Support | 25 | Describe sources of financial or non-financial support for the review, and the role of the funders or sponsors in the review. | Title Page |
| Competing interests | 26 | Declare any competing interests of review authors. | Title Page |
| Availability of data, code and other materials | 27 | Report which of the following are publicly available and where they can be found: template data collection forms; data extracted from included studies; data used for all analyses; analytic code; any other materials used in the review. | - |

sTable 2. Search Terms per Search Engine.

| | |
|---|---|
| PsychInfo<br>&<br>Medline | (AB asd OR AB autism OR AB autistic) AND (AB "polygenic score*" OR AB "polygenic risk score*" OR AB "genetic risk score*" OR AB "genetic score*") |

| | |
|---|---|
| PubMed | (("ASD"[Title/Abstract] OR "autism"[Title/Abstract] OR "autistic"[Title/Abstract]) AND (("polygenic score*"[Title/Abstract] OR "polygenic risk score*"[Title/Abstract] OR "genetic risk score*"[Title/Abstract]) OR "genetic score*"[Title/Abstract])) |
| Web of Science | TS=(asd OR autism OR autistic) AND TS=("polygenic score*" OR "polygenic risk score*" OR "genetic risk score*" OR "genetic score*") |
| Scopus | TITLE-ABS-KEY ( asd OR autism OR autistic ) AND TITLE-ABS-KEY ( "polygenic score*" OR "polygenic risk score*" OR "genetic risk score*" OR "genetic score*" ) |

sTable 3. Quality Assessment

| | Participants | | | | Predictor | | | Outcome | | | Analyses | | | Confounding | | | | NBias |
|---|---|---|---|---|---|---|---|---|---|---|---|---|---|---|---|---|---|---|
| Study/Criterium | A | B | C | D | E | F | G | H | I | J | K | L | M | N | O | P | Q | |
| **ASD diagnosis** | | | | | | | | | | | | | | | | | | |
| Debost et al. (2022) | + | + | + | + | + | - | + | +- | + | - | - | + | +- | +- | + | + | +- | 0 |
| Grove et al. (2019) | + | + | + | + | +- | + | + | + | + | - | - | + | - | + | + | + | + | 0 |
| Hannon et al. (2018) | +- | + | + | + | + | +- | + | + | + | + | +- | + | +- | + | + | + | + | 0 |
| Jansen et al. (2020) | +- | + | + | + | + | + | + | + | + | + | +- | + | +- | + | + | + | + | 0 |
| Klei et al. (2021) | - | - | - | - | +- | + | + | - | - | - | - | - | - | + | + | + | + | 3 |
| Mattheisen et al. (2022) | +- | + | + | + | - | + | + | +- | + | - | - | +- | +- | +- | + | + | +- | 0 |
| Schendel et al. (2022) | + | + | + | + | - | +- | + | + | + | + | +- | + | +- | + | + | + | + | 0 |
| Trost et al. (2022) | +- | - | - | - | + | +- | + | + | + | - | - | - | - | + | + | + | +- | 2 |
| Zhang et al. (2022) | +- | + | + | + | +- | - | + | + | + | +- | +- | + | +- | + | + | + | + | 0 |
| **Autistic traits** | | | | | | | | | | | | | | | | | | |
| Askeland et al. (2021) | + | + | +- | + | + | + | + | + | + | + | +- | + | NA | + | + | + | + | 0 |
| Li et al. (2020) | +- | + | + | + | + | + | + | +- | + | + | +- | + | +- | + | - | + | + | 0 |
| Nayar et al. (2021) | + | + | + | + | +- | + | +- | + | + | + | - | + | NA | + | + | + | + | 0 |
| Reed et al. (2021) | + | + | +- | + | + | + | + | + | + | + | - | + | NA | + | + | + | + | 0 |
| Riglin et al. (2021) | + | + | + | + | +- | + | + | + | + | + | - | - | NA | + | + | + | - | 0 |

| | | | | | | | | | | | | | | | | | | |
|---|---|---|---|---|---|---|---|---|---|---|---|---|---|---|---|---|---|---|
| Serdarevic et al. (2020) | + | + | + | + | + | + | + | + | + | + | +- | + | NA | + | + | + | + | 0 |
| Taylor et al. (2019) | +- | + | + | + | + | + | + | + | + | +- | +- | + | - | +- | + | + | +- | 0 |
| Takahashi et al. (2020) | + | + | + | + | +- | + | + | + | + | + | - | + | NA | + | + | + | +- | 0 |
| Thomas et al. (2022) | + | + | + | + | - | + | - | + | + | - | +- | + | - | + | + | + | + | 1 |
| Yap et al. (2021) | - | +- | + | + | + | + | +- | - | + | - | +- | + | - | - | + | + | +- | 1 |
| Torske et al. (2020) | + | + | + | + | + | + | + | + | + | + | +- | + | +- | + | - | + | - | 0 |
| **Specific psychiatric classifications** | | | | | | | | | | | | | | | | | | |
| Askeland et al. (2021) | + | + | +- | + | + | + | + | + | + | + | +- | + | NA | + | + | + | + | 0 |
| Chang et al. (2020) | + | + | + | - | +- | + | + | + | + | + | +- | +- | +- | + | + | + | + | 0 |
| Havdahl et al. (2022) | + | + | - | + | +- | + | + | + | - | + | - | + | NA | + | + | + | + | 0 |
| Hjorthøj et al. (2021) | - | + | + | + | +- | - | + | + | +- | - | +- | - | - | + | + | + | + | 0 |
| Jansen et al. (2020) | +- | + | + | + | + | + | + | + | + | + | +- | + | +- | + | + | + | + | 0 |
| Jansen et al. (2021) | + | + | + | + | + | + | + | + | + | +- | - | + | - | + | + | + | + | 0 |
| Joo et al. (2022) | + | + | + | + | + | + | + | + | + | + | + | + | NA | + | + | + | +- | 0 |
| Jørgensen et al. (2021) | +- | + | + | + | +- | + | + | + | + | - | - | + | +- | + | + | + | +- | 0 |
| Koomar et al. (2021) | + | + | +- | + | - | + | + | +- | + | + | - | + | - | + | + | + | +- | 0 |
| Legge et al. (2019) | + | + | + | + | +- | + | + | + | - | + | - | + | +- | + | + | + | +- | 0 |
| Legge et al. (2021) | + | + | - | - | +- | + | + | + | + | - | +- | + | - | + | + | + | + | 0 |
| Leppert et al. (2019) | +- | + | - | + | +- | +- | + | + | - | + | - | + | NA | + | + | + | + | 0 |
| Mattheisen et al. (2022) | - | + | + | + | - | + | + | +- | + | - | - | + | - | - | + | + | +- | 0 |
| Ohi et al. (2020) | + | + | + | + | + | + | +- | +- | + | +- | - | - | - | +- | - | + | + | 1 |

| | | | | | | | | | | | | | | | | | | |
|---|---|---|---|---|---|---|---|---|---|---|---|---|---|---|---|---|---|---|
| Qin et al. (2020) | +- | + | + | + | + | +- | + | + | + | +- | +- | + | - | +- | - | + | + | 0 |
| Riglin et al. (2021) | + | + | + | + | +- | + | + | + | + | + | - | - | NA | + | + | + | - | 0 |
| Russell et al. (2021) | - | + | - | + | +- | + | + | + | - | + | - | - | NA | + | + | + | +- | 1 |
| Zhang et al. (2022) | +- | + | + | + | +- | - | + | + | + | +- | +- | + | +- | + | + | + | + | 0 |
| **General psychopathology** | | | | | | | | | | | | | | | | | | |
| Loughnan et al. (2022) | + | +- | - | + | + | + | + | +- | + | + | + | + | NA | + | + | + | + | 0 |
| Y. Gui et al. (2022) | + | + | +- | + | - | + | - | + | +- | + | +- | + | NA | +- | + | +- | + | 1 |
| Pat et al. (2022) | + | + | + | + | + | + | + | + | + | + | - | + | NA | + | + | + | + | 0 |
| Riglin et al. (2020) | + | + | + | + | +- | + | + | + | + | + | - | - | NA | + | + | +- | +- | 1 |
| Schlag et al. (2022) | + | +- | + | +- | +- | + | + | + | + | - | +- | + | NA | + | + | + | + | 0 |
| Thomas et al. (2022) | + | + | + | + | - | + | - | + | + | - | +- | + | - | + | + | + | + | 1 |
| Waszczuk et al. (2021) | + | + | + | + | + | + | + | + | + | + | - | + | NA | +- | + | + | + | 0 |
| **Cognition and executive functioning** | | | | | | | | | | | | | | | | | | |
| Aguilar-Lacasaña et al. (2022) | + | + | + | + | + | + | + | + | + | + | +- | + | NA | + | + | + | + | 0 |
| Chang et al. (2020) | + | + | + | - | +- | + | + | + | + | + | +- | +- | +- | + | + | + | + | 0 |
| Cullen et al. (2021) | + | + | + | + | + | + | +- | + | + | + | +- | + | NA | + | + | + | + | 0 |
| Y. Gui et al. (2022) | + | + | +- | + | - | + | - | + | +- | + | +- | + | NA | +- | + | +- | + | 1 |
| Hughes et al. (2021) | + | + | + | + | +- | + | + | + | + | + | +- | + | NA | + | + | + | +- | 0 |
| Loughnan et al. (2022) | + | +- | - | + | + | + | + | +- | + | + | + | + | NA | + | + | + | + | 0 |
| Price et al. (2020) | + | + | + | + | + | + | + | + | + | - | - | + | NA | + | + | + | +- | 0 |

| | | | | | | | | | | | | | | | | | | |
|---|---|---|---|---|---|---|---|---|---|---|---|---|---|---|---|---|---|---|
| Torske et al. (2020) | + | + | + | + | + | + | + | + | + | + | +- | + | +- | + | - | + | - | 0 |
| Yap et al. (2021) | - | +- | + | + | + | + | +- | - | + | - | +- | + | - | - | + | + | +- | 1 |
| **Early neurodevelopment** | | | | | | | | | | | | | | | | | | |
| Askeland et al. (2021) | + | + | +- | + | + | + | + | + | + | + | +- | + | NA | + | + | + | + | 0 |
| Fish et al. (2021) | +- | - | - | + | + | + | + | + | + | +- | - | + | - | + | - | + | + | 0 |
| A. Gui et al. (2020) | + | + | + | + | + | + | + | + | + | + | - | +- | NA | + | - | + | +- | 0 |
| Hannigan et al. (2023) | + | + | + | + | + | + | + | + | +- | + | +- | + | NA | + | + | + | + | 0 |
| Portugal et al. (2022) | + | + | + | + | +- | +- | - | + | + | + | +- | + | NA | + | + | + | + | 0 |
| Riglin et al. (2022) | +- | + | + | + | +- | + | + | + | + | + | +- | + | NA | +- | + | + | + | 0 |
| Serdarevic et al. (2020) | + | + | + | + | + | + | + | + | + | + | +- | + | NA | + | + | + | + | 0 |
| Takahashi et al. (2020) | + | + | + | + | +- | + | + | + | + | + | - | + | NA | + | + | + | +- | 0 |
| Yap et al. (2021) | - | +- | + | + | + | + | +- | - | + | - | +- | + | - | - | + | + | +- | 1 |
| **Physical wellbeing** | | | | | | | | | | | | | | | | | | |
| Dennison et al. (2021) | + | + | + | + | +- | + | +- | + | + | + | +- | + | + | + | + | + | + | 0 |
| Havdahl et al. (2022) | + | + | - | + | +- | + | +- | + | - | + | - | + | NA | + | + | + | + | 0 |
| Hunjan et al. (2021) | + | + | + | + | - | +- | + | + | - | + | + | + | NA | + | + | + | + | 0 |
| Leppert et al. (2019) | +- | + | - | + | +- | +- | + | + | - | + | - | + | NA | + | + | + | + | 0 |
| Niarchou et al. (2022) | + | + | + | + | +- | + | +- | + | +- | - | +- | + | - | +- | +- | + | + | 0 |
| Ohi et al. (2021) | + | + | - | +- | + | +- | + | + | + | + | - | +- | NA | + | + | + | + | 0 |
| Werner et al. (2022) | + | + | + | +- | + | + | + | + | + | + | +- | + | - | + | +- | + | + | 0 |
| Zhang et al. (2022) | +- | + | + | + | +- | - | + | + | + | +- | +- | + | +- | + | + | + | + | 0 |

| | | | | | | | | | | | | | | | | | | |
|---|---|---|---|---|---|---|---|---|---|---|---|---|---|---|---|---|---|---|
| **Emotion recognition** | | | | | | | | | | | | | | | | | | |
| Qin et al. (2020) | +- | + | + | + | + | +- | + | + | + | +- | +- | + | - | +- | - | + | + | 0 |
| Reed et al. (2021) | + | + | +- | + | + | + | + | + | + | + | - | + | NA | + | + | + | + | 0 |
| Waddington et al. (2021) | + | + | + | + | + | + | + | + | + | +- | +- | + | - | + | - | + | + | 0 |
| **Phe-WAS** | | | | | | | | | | | | | | | | | | |
| Leppert et al. (2020) | + | + | + | + | + | +- | + | +- | - | + | +- | + | NA | + | + | + | + | 0 |
| Sha et al. (2021) | + | + | + | +- | - | +- | + | + | + | +- | +- | + | NA | +- | + | + | + | 0 |
| Wendt et al. (2020) | + | + | + | - | - | + | + | - | - | - | +- | + | NA | + | + | + | +- | 1 |
| **Brain measures** | | | | | | | | | | | | | | | | | | |
| Alemany et al. (2021) | + | + | + | + | + | +- | + | + | + | + | +- | - | NA | + | + | + | + | 0 |
| Ecker et al. (2022) | + | + | + | + | + | + | +- | + | +- | + | - | +- | - | +- | + | +- | + | 0 |
| Gui et al. (2021) | + | +- | +- | + | + | + | + | + | + | + | - | +- | - | + | - | +- | +- | 0 |
| Gui et al. (2022) | + | + | +- | + | - | + | - | + | +- | + | +- | + | NA | +- | + | +- | + | 1 |
| P. R. Jansen et al. (2019) | + | + | + | + | + | + | +- | + | + | + | +- | + | NA | + | + | + | + | 0 |
| Khundrakpam et al. (2020) | + | + | + | + | + | + | + | + | + | +- | +- | + | NA | +- | - | + | + | 0 |
| Lawrence et al. (2022) | + | +- | + | - | + | + | +- | + | + | +- | +- | + | - | - | - | + | - | 1 |
| Mason et al. (2022) | + | + | + | + | +- | +- | + | + | + | +- | - | - | - | + | - | + | - | 1 |
| Qin et al. (2020) | +- | + | + | + | + | +- | + | + | + | +- | +- | + | - | +- | - | + | + | 0 |
| Sha et al. (2021) | + | + | + | +- | - | +- | + | + | + | +- | +- | + | NA | + | + | + | + | 0 |
| **Other** | | | | | | | | | | | | | | | | | | |
| Cullen et al. (2021) | + | + | + | + | + | + | +- | + | + | + | +- | + | NA | + | + | + | + | 0 |

| | | | | | | | | | | | | | | | | | | |
|---|---|---|---|---|---|---|---|---|---|---|---|---|---|---|---|---|---|---|
| Debost et al. (2022) | + | + | + | + | + | - | + | +- | + | - | - | + | +- | +- | + | + | +- | 0 |
| Hannon et al. (2018) | +- | + | + | + | + | +- | + | + | + | + | +- | + | +- | + | + | + | + | 0 |
| Havdahl et al. (2022) | + | + | - | + | +- | + | +- | + | - | + | - | + | NA | + | + | + | + | 0 |
| Kalman et al. (2021) | +- | + | + | - | +- | + | + | + | +- | - | - | + | +- | + | + | + | + | 0 |
| Klei et al. (2021) | - | - | - | - | +- | + | + | - | - | - | - | - | - | + | + | + | + | 3 |
| Klein et al. (2022) | - | + | + | + | +- | + | + | - | +- | +- | +- | + | +- | +- | + | + | +- | 0 |
| Li et al. (2020) | +- | + | + | + | + | + | + | +- | + | + | +- | + | +- | + | - | + | + | 0 |
| Maxwell et al. (2021) | + | + | +- | + | + | + | +- | + | + | + | +- | + | NA | + | + | + | + | 0 |
| Morneau-Vaillancourt et al. (2021) | +- | + | +- | + | + | + | +- | + | +- | - | - | + | NA | + | + | + | + | 0 |
| Ohi et al. (2020) | + | + | + | + | + | + | +- | +- | + | +- | - | - | - | +- | - | + | + | 1 |
| Ohi et al. (2021) | + | + | - | +- | + | +- | + | + | + | + | - | +- | NA | + | + | + | + | 0 |
| Peel et al. (2022) | + | + | + | + | + | + | + | + | +- | + | +- | + | NA | + | + | + | + | 0 |
| Strom et al. (2022) | + | + | +- | + | +- | + | +- | + | + | - | +- | +- | NA | + | + | + | + | 0 |
| Torske et al. (2020) | + | + | + | + | + | + | + | + | + | + | +- | + | +- | + | - | + | - | 0 |
| Ratanatharathorn et al. (2021) | + | + | + | + | + | +- | + | + | +- | + | +- | + | NA | + | + | + | +- | 0 |
| Sha et al. (2021) | + | + | + | +- | - | +- | + | + | + | +- | +- | + | NA | +- | + | + | + | 0 |
| Warrier & Baron-Cohen (2021) | + | + | + | + | +- | +- | + | + | + | + | +- | + | +- | + | +- | + | + | 0 |
| Yap et al. (2021) | - | +- | + | + | + | + | +- | - | + | - | +- | + | - | - | + | + | +- | 1 |

| | | | | | | | | | | | | | | | | | | |
|---|---|---|---|---|---|---|---|---|---|---|---|---|---|---|---|---|---|---|
| Zhang et al. (2022) | +- | + | + | + | +- | - | + | + | + | +- | +- | + | +- | + | + | + | + | 0 |

Note. Studies highlighted in gray have been presented earlier in the table due to them being in multiple outcome categories. A bias is detected when > 50% of the criteria within one domain are scored -. Criterium M was not taken included when counting biases.

sTable 4. Rosenthal's fail-safe N per outcome category

| **Outcome category** | Rosenthal's fail-safe N |
|---|---|
| Autism diagnosis | 39921 |
| Autistic traits | 1544 |
| Other specific psychiatric classifications | 95187 |
| General psychopathology | 90341 |
| Cognition and executive function | 1805 |
| Physical wellbeing | 30482 |
| Early neurodevelopment | 329 |
| Emotion recognition | 102 |
| Brain measures | 10733 |

sTable S5. Multi-level Meta-Analysis Results for Subcategories Specific Psychiatric Classifications – Secondary Results

| | | | | | | | $I^2$ | | |
|---|---|---|---|---|---|---|---|---|---|
| Subcategory | *n* studies | *n* ind. Coh. | *n* est. | $r_{Pooled}$ [95% CI] | *r* range | *P* | Total | Between-cohort | Within-cohort |
| ADHD | 5 | 5 | 13 | .049 [−.002 − .100] | −.03 − .110 | .057 | 96.3% | 84.5% | 11.8% |
| Psychotic | 4 | 4 | 18 | .072 [ .025 − .119] | −.031 − .213 | .005 | 98.9% | 72.7% | 26.2% |
| Eating disorder | 2 | 2 | 11 | .005 [−.015 − .024] | −.038 − .058 | .625 | 58.6% | 0.0% | 58.6% |
| Self-harm | 3 | 3 | 18 | .029 [ -.076 − .135] | −.185 − .17 | .564 | 99.7% | 71.6% | 28.1% |
| Other | 7 | 5 | 18 | .063 [ .001 − .125] | −.117 − .199 | .047 | 99.2% | 24.7% | 74.5% |

*NOTE*. *n* studies = number of studies, *n* ind. Coh = number of independent cohorts the included studies are based on, *n* est = number of effect size estimates included

**Supplementary figures**

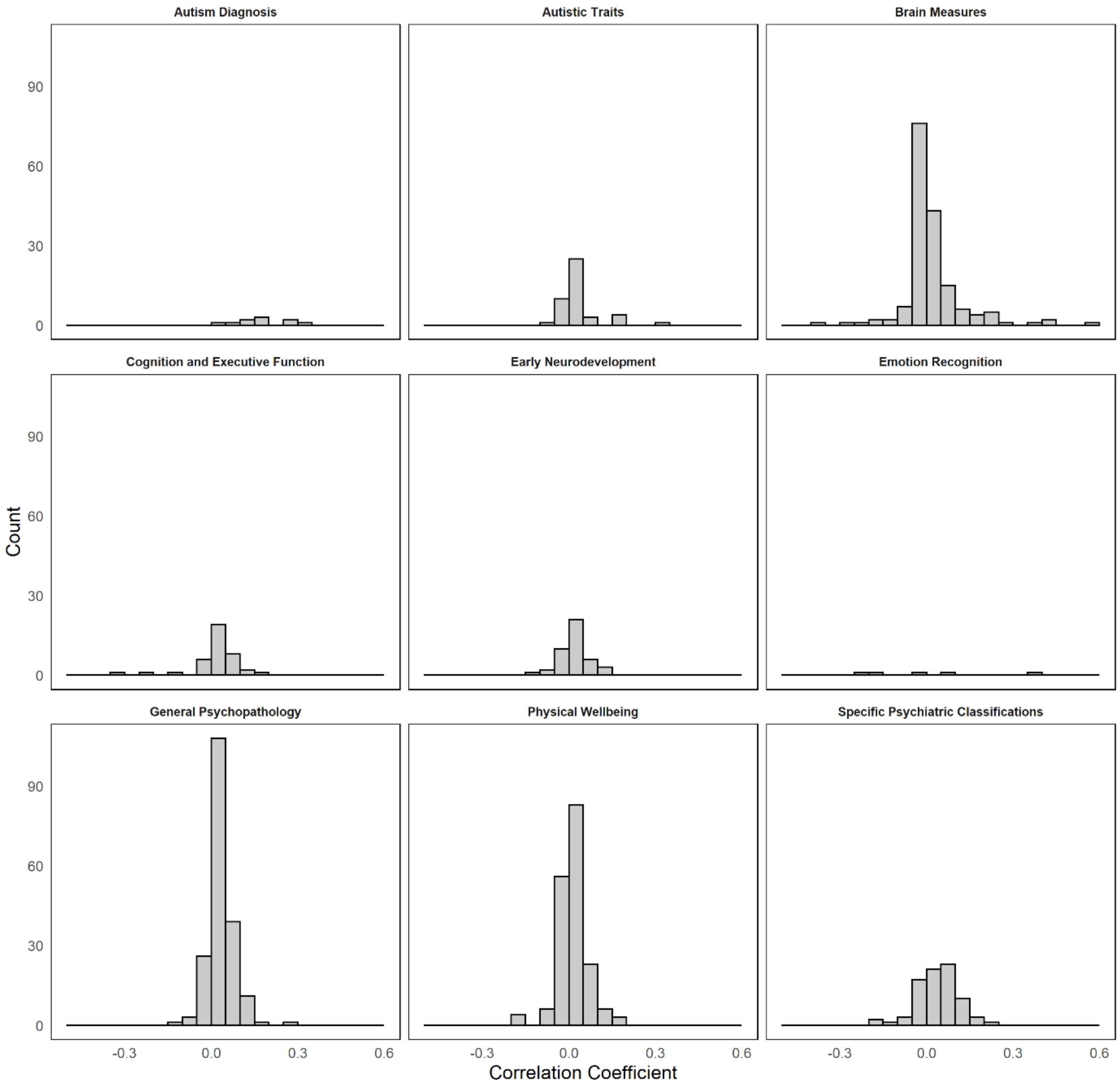

sFigure 1. Histograms of Effect Sizes (r) per Outcome Category

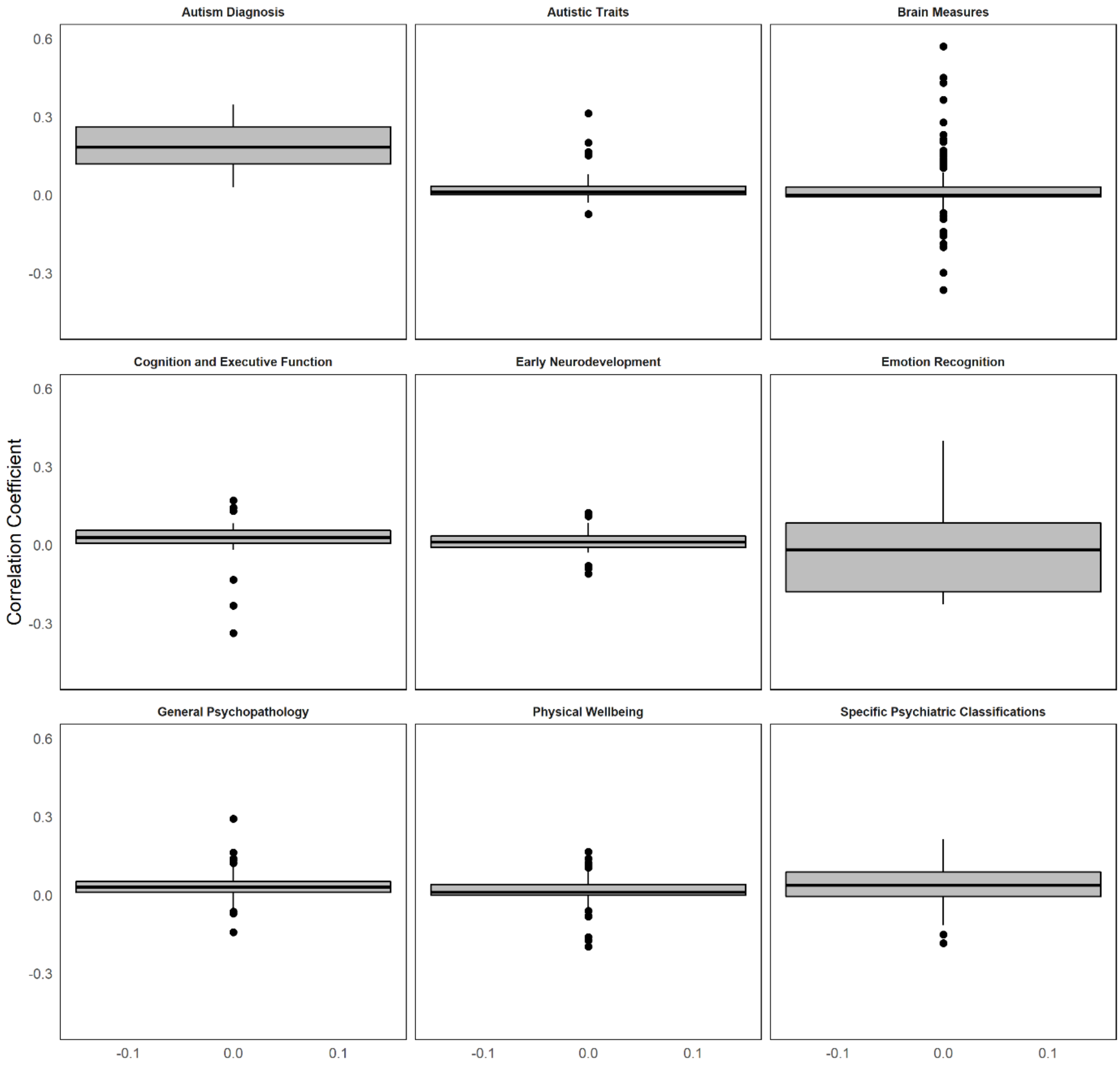

sFigure 2. Boxplots of Effect Sizes (r) per Outcome Category

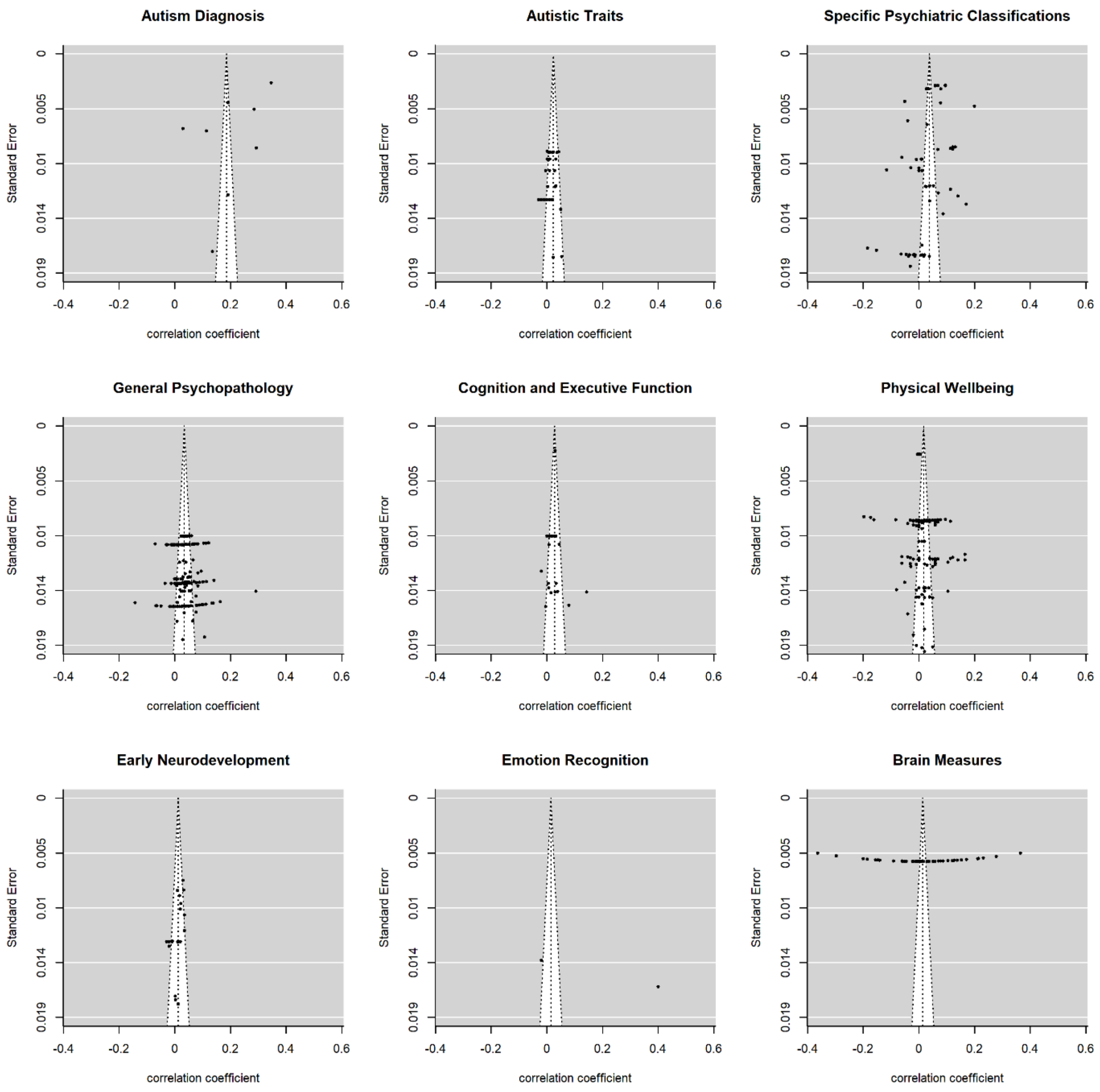

sFigure 3. Funnel Plots of Standard Errors per Outcome Category

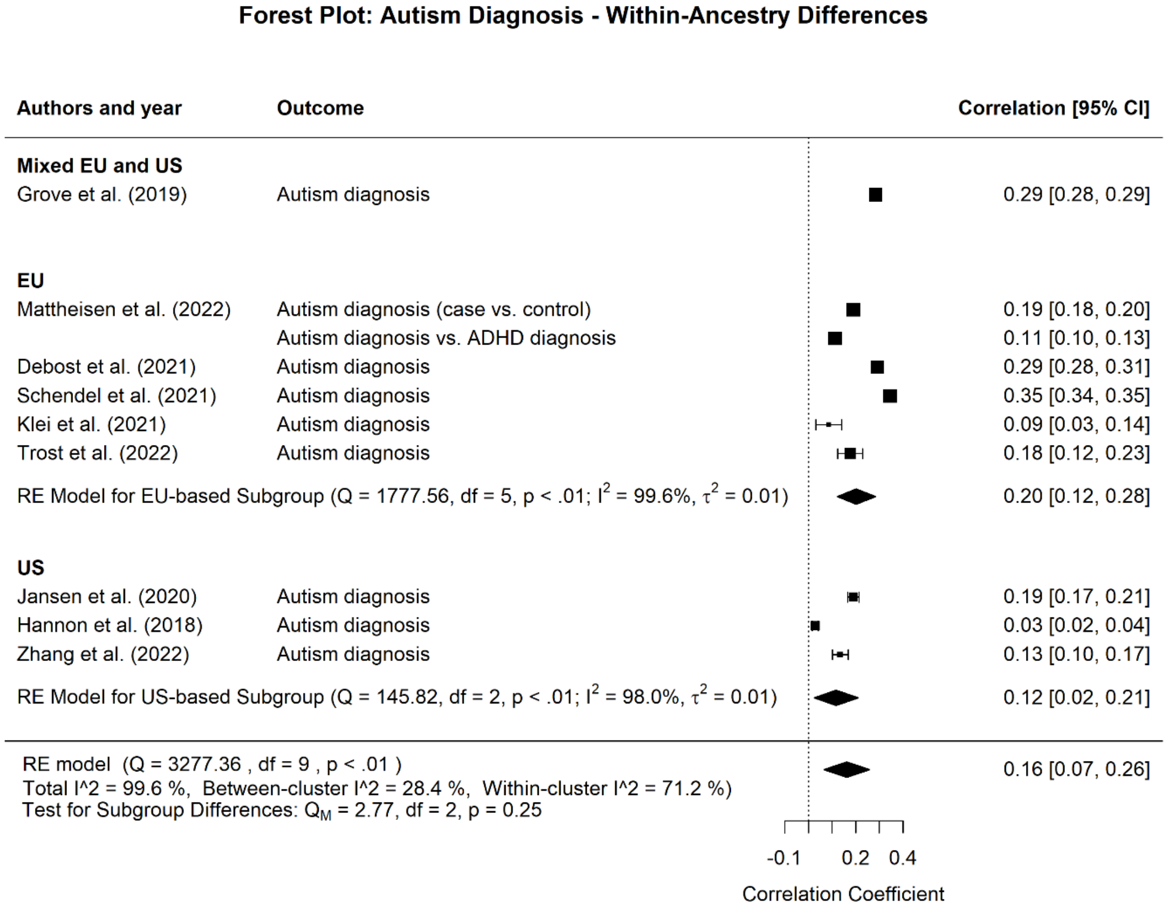

sFigure 4. Multi-level Meta-Analysis Results on the Association between Autism Polygenic Score and Autism Diagnosis, including a Test for Population Differences.

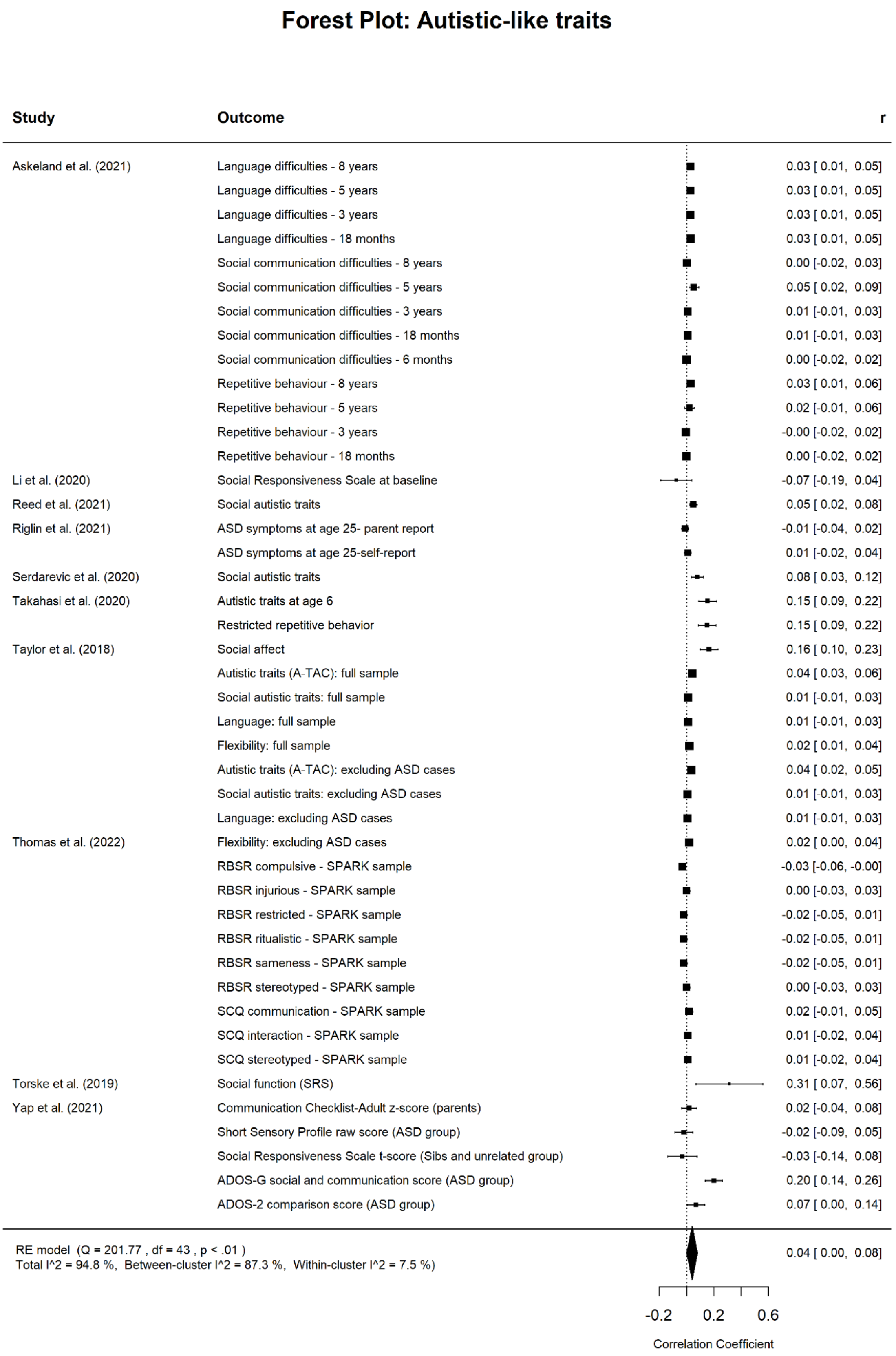

sFigure 5. Multi-level Meta-Analysis Results on the Association between Autism Polygenic Score and Autistic-like Traits.

### Forest Plot: Specific Psychiatric Classifications

| Study | Outcome | r [95% CI] |
|---|---|---|
| Askeland et al.(2020) | Inattention - 8 years | 0.05 [ 0.03, 0.07] |
| | Inattention - 5 years | 0.02 [-0.00, 0.04] |
| | Inattention - 3 years | -0.00 [-0.02, 0.02] |
| | Inattention - 18 months | 0.01 [-0.01, 0.03] |
| | Hyperactivity - 8 years | 0.04 [ 0.02, 0.06] |
| | Hyperactivity - 5 years | 0.03 [ 0.00, 0.05] |
| | Hyperactivity - 3 years | 0.01 [-0.01, 0.03] |
| | Hyperactivity 18 months | -0.01 [-0.03, 0.01] |
| Chang et al. (2020) | ADHD symptoms | 0.09 [ 0.03, 0.14] |
| Havdahl et al.(2022) | Lifetime depression (mother) | 0.11 [ 0.10, 0.13] |
| | Lifetime depression (father) | 0.07 [ 0.05, 0.08] |
| | Depression/anxiety symptoms | 0.12 [ 0.11, 0.14] |
| | Depression/anxiety symptoms - during pregnancy | 0.12 [ 0.11, 0.14] |
| | Depression/anxiety symptoms - trimester 1 | 0.13 [ 0.11, 0.15] |
| | Depression/anxiety symptoms - trimester 2-3 | 0.12 [ 0.11, 0.14] |
| Hjortoj et al.(2019) | Cannabis use disorder - Other disorders | -0.05 [-0.06, -0.04] |
| | Cannabis use disorder - Schizophrenia patients | 0.01 [-0.02, 0.04] |
| | Cannabis use disorder - controls | -0.04 [-0.05, -0.03] |
| Jansen et al. (2020) | Either ADHD or ASD diagnosis | 0.06 [ 0.02, 0.11] |
| | ADHD diagnosis (ASD not permitted as co-diagnosis) | 0.11 [ 0.05, 0.17] |
| Jansen et al. (2021) | Case-control status for any DSM diagnosis | -0.03 [-0.07, 0.01] |
| Joo et al. (2022) | Active suicidal ideation - European cohort | 0.17 [ 0.14, 0.20] |
| | Overall suicidal ideation - multi-ethnic cohort | 0.07 [ 0.05, 0.09] |
| | Active suicidal ideation - multi-ethnic cohort | 0.14 [ 0.12, 0.16] |
| | Overall suicidal thoughts and behaviors - multi-ethnic cohort | 0.07 [ 0.05, 0.09] |
| Jorgenson et al. (2021) | Diagnosis of nocturnal eneresis | 0.03 [ 0.02, 0.04] |
| Koomar et al. (2021) | Picky factor - proband | 0.04 [ 0.00, 0.07] |
| | Fear factor - proband | -0.04 [-0.07, -0.00] |
| | Appetite factor - proband | -0.02 [-0.05, 0.02] |
| | ARFID score - proband | 0.02 [-0.02, 0.05] |
| | Picky factor - parent | 0.03 [-0.01, 0.07] |
| | Fear factor - parent | -0.01 [-0.05, 0.04] |
| | Appetite factor - parent | -0.02 [-0.06, 0.02] |
| | ARFID score - parent | 0.06 [ 0.02, 0.10] |
| Legge et al. (2021) | Current cognitive ability (schizophrenia) | 0.03 [-0.05, 0.10] |
| | Disorganized symptoms (schizophrenia) | 0.06 [-0.02, 0.13] |
| | Negative symptoms of motivation and pleasure (schizophrenia) | -0.02 [-0.10, 0.05] |
| | Negative symptoms of diminished expressivity (schizophrenia) | -0.03 [-0.11, 0.04] |
| | Positive symptoms (schizophrenia) | 0.09 [ 0.02, 0.16] |
| Legge et al. (2019) | Delusions of persecution | 0.10 [ 0.09, 0.10] |
| | Delusions of reference | 0.10 [ 0.09, 0.10] |
| | Auditory hallucinations | 0.10 [ 0.09, 0.10] |
| | Visual hallucinations | 0.06 [ 0.05, 0.06] |
| | Multiple psychotic experiences vs, controls | 0.10 [ 0.09, 0.10] |
| | Non-distressing psychotic experience vs, controls | 0.06 [ 0.05, 0.06] |
| | Distressing vs non-distressing psychotic experience | 0.04 [ 0.01, 0.06] |
| | Distressing vs, controls | 0.10 [ 0.09, 0.10] |
| | Any psychotic experience vs, controls | 0.07 [ 0.06, 0.07] |

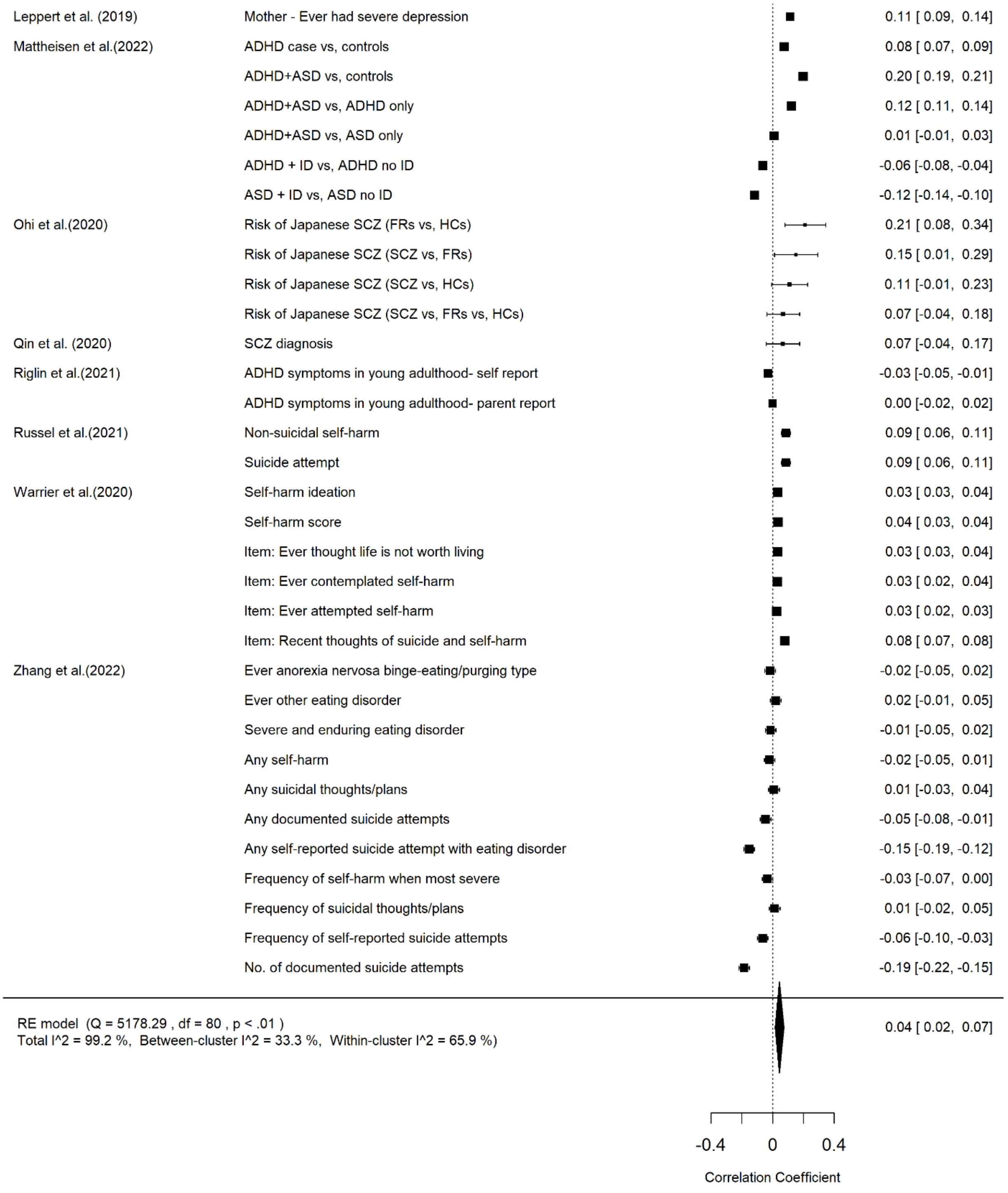

sFigure 6. Multi-level Meta-Analysis Results on the Association between Autism Polygenic Score and Specific Psychiatric Classifications.

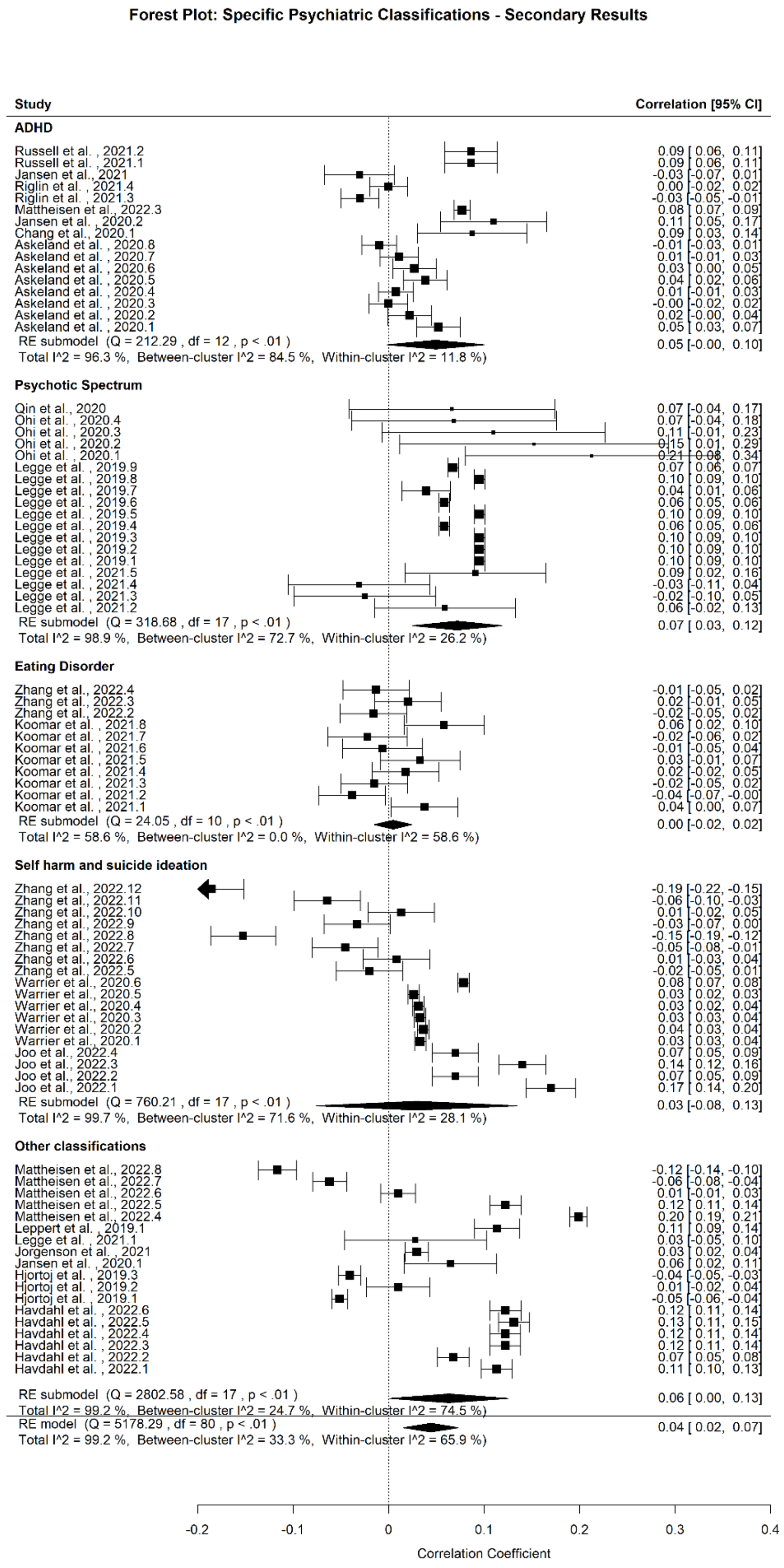

sFigure 7. Multi-level Meta-Analysis Results on the Association between Autism Polygenic Score and Subclassifications of Specific Psychiatric Classifications - Secondary results.

### Forest Plot: General Psychopathology

| Study | Outcome | r [95% CI] |
|---|---|---|
| Gui et al. (2022) | cbcl_scr_syn_anxdep_r | 0.02 [ 0.01, 0.04] |
| | cbcl_scr_syn_withdep_r | 0.05 [ 0.03, 0.07] |
| | cbcl_scr_syn_somatic_r | 0.02 [ 0.00, 0.04] |
| | cbcl_scr_syn_social_r | 0.03 [ 0.01, 0.05] |
| | cbcl_scr_syn_thought_r | 0.03 [ 0.01, 0.05] |
| | cbcl_scr_syn_attention_r | 0.05 [ 0.03, 0.07] |
| | cbcl_scr_syn_rulebreak_r | 0.02 [ 0.01, 0.04] |
| | cbcl_scr_syn_aggressive_r | 0.03 [ 0.01, 0.05] |
| | cbcl_scr_syn_internal_r | 0.04 [ 0.02, 0.06] |
| | cbcl_scr_syn_external_r | 0.03 [ 0.01, 0.05] |
| | cbcl_scr_syn_totprob_r | 0.04 [ 0.02, 0.06] |
| | cbcl_scr_dsm5_depress_r | 0.04 [ 0.02, 0.06] |
| | cbcl_scr_dsm5_anxdisord_r | 0.02 [ 0.00, 0.04] |
| | cbcl_scr_dsm5_somaticpr_r | 0.02 [ 0.00, 0.04] |
| | cbcl_scr_dsm5_adhd_r | 0.05 [ 0.03, 0.06] |
| | cbcl_scr_dsm5_opposit_r | 0.03 [ 0.02, 0.05] |
| | cbcl_scr_dsm5_conduct_r | 0.03 [ 0.01, 0.05] |
| | cbcl_scr_07_sct_r | 0.06 [ 0.04, 0.08] |
| | cbcl_scr_07_ocd_r | 0.03 [ 0.01, 0.05] |
| | cbcl_scr_07_stress_r | 0.03 [ 0.02, 0.05] |
| Loughnan et al.(2022) | Prosociality (youth) - mixed ancestry | 0.03 [ 0.01, 0.05] |
| | BISBAS Drive (youth) - mixed ancestry | 0.01 [-0.01, 0.03] |
| | BISBAS Fun Seeking (youth) - mixed ancestry | -0.03 [-0.05, -0.01] |
| | BISBAS Reward Responsiveness (youth) - mixed ancestry | 0.01 [-0.01, 0.03] |
| | BISBAS Inhibition (youth) - mixed ancestry | 0.02 [-0.00, 0.04] |
| | UPPS Lack of Perseverance (youth) - mixed ancestry | -0.00 [-0.02, 0.02] |
| | UPPS Lack of Planning (youth) - mixed ancestry | 0.01 [-0.01, 0.03] |
| | UPPS Positive Urgency (youth) - mixed ancestry | -0.00 [-0.02, 0.02] |
| | UPPS Negative Urgency (youth) - mixed ancestry | 0.00 [-0.02, 0.02] |
| | UPPS Sensation Seeking (youth) - mixed ancestry | -0.01 [-0.03, 0.01] |
| | Prodromal Pyschosis Severity Score (youth) - mixed ancestry | 0.02 [ 0.00, 0.04] |
| | KSADS Symptoms Bipolar (youth) - mixed ancestry | 0.05 [ 0.03, 0.07] |
| | KSADS Symptoms Depression (youth) - mixed ancestry | 0.12 [ 0.10, 0.14] |
| | KSADS Symptoms Anxiety (youth) - mixed ancestry | 0.04 [ 0.02, 0.06] |
| | KSADS Symptoms Insomnia (youth) - mixed ancestry | 0.11 [ 0.09, 0.13] |
| | KSADS Symptoms Suicidality (youth) - mixed ancestry | 0.08 [ 0.06, 0.10] |
| | KSADS Total Symptoms (youth) - mixed ancestry | 0.07 [ 0.05, 0.09] |
| | KSADS Symptoms Bipolar (caregiver) - mixed ancestry | -0.01 [-0.03, 0.01] |
| | KSADS Symptoms Depression (caregiver) - mixed ancestry | 0.10 [ 0.08, 0.12] |
| | KSADS Symptoms Anxiety (caregiver) - mixed ancestry | 0.04 [ 0.02, 0.06] |
| | KSADS Symptoms OCD (caregiver) - mixed ancestry | 0.06 [ 0.04, 0.09] |
| | KSADS Symptoms Eating Disorder (caregiver) - mixed ancestry | 0.04 [ 0.02, 0.06] |

| | |
|---|---|
| KSADS Symptoms Eating Disorder (caregiver) - mixed ancestry | 0.04 [ 0.02, 0.06] |
| KSADS Symptoms ADHD (caregiver) - mixed ancestry | 0.08 [ 0.06, 0.10] |
| KSADS Symptoms Oppositional/Conduct (caregiver) - mixed ancestry | -0.00 [-0.02, 0.02] |
| KSADS Symptoms Developmental Disorders (caregiver) - mixed ancestry | 0.07 [ 0.05, 0.09] |
| KSADS Symptoms PTSD (caregiver) - mixed ancestry | -0.03 [-0.05, -0.01] |
| KSADS Symptoms Insomnia (caregiver) - mixed ancestry | -0.01 [-0.03, 0.01] |
| KSADS Symptoms Suicidality (caregiver) - mixed ancestry | -0.07 [-0.09, -0.05] |
| KSADS Total Symptoms (caregiver) - mixed ancestry | 0.03 [ 0.01, 0.05] |
| General Behavior Inventory - Mania (caregiver) - mixed ancestry | 0.01 [-0.01, 0.03] |
| CBCL Aggressive (caregiver) - mixed ancestry | -0.01 [-0.03, 0.01] |
| CBCL Anxious/Depressive (caregiver) - mixed ancestry | 0.01 [-0.01, 0.03] |
| CBCL Rule-breaking (caregiver) - mixed ancestry | -0.03 [-0.05, -0.01] |
| CBCL Inattention (caregiver) - mixed ancestry | 0.03 [ 0.01, 0.05] |
| CBCL Social Problems (caregiver) - mixed ancestry | -0.01 [-0.03, 0.01] |
| CBCL Thought Problems (caregiver) - mixed ancestry | 0.00 [-0.02, 0.02] |
| CBCL Somatic Complaints (caregiver) - mixed ancestry | -0.01 [-0.03, 0.01] |
| CBCL Withdrawn/Depressive (caregiver) - mixed ancestry | 0.06 [ 0.04, 0.08] |
| CBCL Total Problems (caregiver) - mixed ancestry | 0.01 [-0.01, 0.03] |
| Prosociality (youth) - non-European sample | 0.03 [-0.00, 0.06] |
| BISBAS Drive (youth) - non-European sample | -0.00 [-0.04, 0.03] |
| BISBAS Fun Seeking (youth) - non-European sample | -0.00 [-0.04, 0.03] |
| BISBAS Reward Responsiveness (youth) - non-European sample | -0.07 [-0.10, -0.04] |
| BISBAS Inhibition (youth) - non-European sample | 0.01 [-0.02, 0.04] |
| UPPS Lack of Perseverance (youth) - non-European sample | 0.00 [-0.03, 0.03] |
| UPPS Lack of Planning (youth) - non-European sample | 0.02 [-0.01, 0.05] |
| UPPS Positive Urgency (youth) - non-European sample | 0.02 [-0.01, 0.06] |
| UPPS Negative Urgency (youth) - non-European sample | -0.00 [-0.04, 0.03] |
| UPPS Sensation Seeking (youth) - non-European sample | -0.05 [-0.08, -0.02] |
| Prodromal Pyschosis Severity Score (youth) - non-European sample | 0.03 [-0.00, 0.06] |
| KSADS Symptoms Bipolar (youth) - non-European sample | 0.05 [ 0.02, 0.08] |
| KSADS Symptoms Depression (youth) - non-European sample | 0.06 [ 0.03, 0.09] |
| KSADS Symptoms Anxiety (youth) - non-European sample | 0.14 [ 0.11, 0.17] |
| KSADS Symptoms Insomnia (youth) - non-European sample | 0.11 [ 0.08, 0.14] |
| KSADS Symptoms Suicidality (youth) - non-European sample | 0.05 [ 0.02, 0.08] |
| KSADS Total Symptoms (youth) - non-European sample | 0.04 [ 0.01, 0.07] |
| KSADS Symptoms Bipolar (caregiver) - non-European sample | 0.02 [-0.01, 0.05] |
| KSADS Symptoms Depression (caregiver) - non-European sample | 0.16 [ 0.13, 0.19] |
| KSADS Symptoms Anxiety (caregiver) - non-European sample | 0.10 [ 0.07, 0.13] |
| KSADS Symptoms OCD (caregiver) - non-European sample | 0.29 [ 0.26, 0.32] |
| KSADS Symptoms Eating Disorder (caregiver) - non-European sample | 0.04 [ 0.01, 0.07] |
| KSADS Symptoms ADHD (caregiver) - non-European sample | 0.13 [ 0.10, 0.16] |
| KSADS Symptoms Oppositional/Conduct (caregiver) - non-European sample | -0.02 [-0.05, 0.01] |
| KSADS Symptoms Developmental Disorders (caregiver) - non-European sample | 0.05 [ 0.02, 0.08] |
| KSADS Symptoms PTSD (caregiver) - non-European sample | 0.04 [ 0.01, 0.07] |
| KSADS Symptoms Insomnia (caregiver) - non-European sample | 0.10 [ 0.07, 0.13] |
| KSADS Symptoms Suicidality (caregiver) - non-European sample | -0.14 [-0.17, -0.11] |
| KSADS Total Symptoms (caregiver) - non-European sample | 0.08 [ 0.05, 0.11] |
| General Behavior Inventory - Mania (caregiver) - non-European sample | 0.08 [ 0.05, 0.11] |
| CBCL Aggressive (caregiver) - non-European sample | -0.01 [-0.04, 0.02] |
| CBCL Anxious/Depressive (caregiver) - non-European sample | 0.04 [ 0.01, 0.07] |
| CBCL Rule-breaking (caregiver) - non-European sample | -0.06 [-0.10, -0.03] |
| CBCL Inattention (caregiver) - non-European sample | 0.02 [-0.01, 0.05] |
| CBCL Social Problems (caregiver) - non-European sample | 0.01 [-0.02, 0.04] |
| CBCL Thought Problems (caregiver) - non-European sample | 0.05 [ 0.02, 0.08] |
| CBCL Somatic Complaints (caregiver) - non-European sample | 0.00 [-0.03, 0.03] |

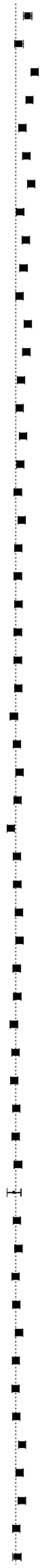

| | | |
|---|---|---|
| | CBCL Withdrawn/Depressive (caregiver) - non-European sample | 0.09 [ 0.06, 0.12] |
| | CBCL Total Problems (caregiver) - non-European sample | 0.03 [-0.01, 0.06] |
| | KSADS Symptoms Depression (youth) - European sample | 0.14 [ 0.11, 0.17] |
| | KSADS Total Symptoms (youth) - European sample | 0.10 [ 0.07, 0.13] |
| | CBCL Inattention (caregiver) - European sample | 0.05 [ 0.02, 0.08] |
| | KSADS Symptoms ADHD (caregiver) - European sample | 0.08 [ 0.05, 0.10] |
| | KSADS Symptoms Suicidality (youth) - European sample | 0.11 [ 0.09, 0.14] |
| | CBCL Total Problems (caregiver) - European sample | 0.03 [ 0.00, 0.06] |
| | KSADS Symptoms Bipolar (youth) - European sample | 0.08 [ 0.05, 0.10] |
| | Prodromal Pyschosis Severity Score (youth) - European sample | 0.06 [ 0.03, 0.09] |
| | Prosociality (youth) - European sample | 0.03 [-0.00, 0.05] |
| | KSADS Symptoms Insomnia (youth) - European sample | 0.09 [ 0.06, 0.12] |
| | KSADS Symptoms Depression (caregiver) - European sample | 0.08 [ 0.05, 0.11] |
| | CBCL Withdrawn/Depressive (caregiver) - European sample | 0.04 [ 0.01, 0.07] |
| | KSADS Total Symptoms (caregiver) - European sample | 0.03 [ 0.00, 0.06] |
| | KSADS Symptoms Developmental Disorders (caregiver) - European sample | 0.05 [ 0.03, 0.08] |
| | BISBAS Reward Responsiveness (youth) - European sample | 0.03 [ 0.01, 0.06] |
| | BISBAS Drive (youth) - European sample | 0.02 [-0.01, 0.04] |
| | KSADS Symptoms Oppositional/Conduct (caregiver) - European sample | 0.04 [ 0.02, 0.07] |
| | BISBAS Inhibition (youth) - European sample | 0.02 [-0.01, 0.05] |
| | UPPS Lack of Planning (youth) - European sample | 0.02 [-0.01, 0.04] |
| | CBCL Aggressive (caregiver) - European sample | 0.02 [-0.01, 0.05] |
| | UPPS Sensation Seeking (youth) - European sample | 0.02 [-0.01, 0.04] |
| | CBCL Thought Problems (caregiver) - European sample | 0.02 [-0.01, 0.04] |
| | CBCL Rule-breaking (caregiver) - European sample | 0.02 [-0.01, 0.05] |
| | BISBAS Fun Seeking (youth) - European sample | -0.01 [-0.04, 0.01] |
| | UPPS Lack of Perseverance (youth) - European sample | 0.01 [-0.02, 0.04] |
| | KSADS Symptoms Anxiety (caregiver) - European sample | 0.03 [-0.00, 0.05] |
| | CBCL Social Problems (caregiver) - European sample | 0.02 [-0.01, 0.04] |
| | KSADS Symptoms Insomnia (caregiver) - European sample | -0.03 [-0.06, -0.01] |
| | UPPS Negative Urgency (youth) - European sample | 0.01 [-0.02, 0.04] |
| | CBCL Anxious/Depressive (caregiver) - European sample | 0.01 [-0.02, 0.04] |
| | KSADS Symptoms Eating Disorder (caregiver) - European sample | 0.02 [-0.00, 0.05] |
| | KSADS Symptoms Suicidality (caregiver) - European sample | 0.03 [ 0.00, 0.05] |
| | CBCL Somatic Complaints (caregiver) - European sample | 0.01 [-0.02, 0.04] |
| | KSADS Symptoms Bipolar (caregiver) - European sample | 0.01 [-0.01, 0.04] |
| | KSADS Symptoms PTSD (caregiver) - European sample | -0.01 [-0.04, 0.01] |
| | General Behavior Inventory - Mania (caregiver) - European sample | -0.00 [-0.03, 0.03] |
| | KSADS Symptoms Anxiety (youth) - European sample | -0.01 [-0.04, 0.02] |
| | KSADS Symptoms OCD (caregiver) - European sample | 0.00 [-0.02, 0.03] |
| | UPPS Positive Urgency (youth) - European sample | 0.00 [-0.03, 0.03] |
| Pat et al. (2021) | P factor - European ancestry | 0.02 [-0.01, 0.05] |
| | P factor - African ancestry | -0.01 [-0.06, 0.04] |
| Riglin et al.(2020) | General psychopathology - age 13 | 0.01 [-0.02, 0.04] |
| | Emotional problems determined through factor analysis - age 13 | 0.02 [-0.01, 0.05] |
| | Behavioural problems determined through factor analysis - age 13 | -0.00 [-0.03, 0.03] |
| | Neurodevelopmental problems determined through factor analysis - age 13 | 0.00 [-0.02, 0.03] |
| | General psychopathology - age 7 | 0.03 [-0.00, 0.05] |
| | Emotional problems determined through factor analysis - age 7 | 0.00 [-0.02, 0.03] |
| | Behavioural problems determined through factor analysis - age 7 | -0.00 [-0.03, 0.03] |
| | Neurodevelopmental problems determined through factor analysis - age 7 | 0.00 [-0.02, 0.03] |
| Schlag et al. (2022) | ALSPAC Parent reported low prosociality - age 7 | 0.05 [ 0.02, 0.07] |
| | ALSPAC Parent reported low prosociality - age 10 | 0.03 [ 0.00, 0.06] |
| | ALSPAC Parent reported low prosociality - age 12 | 0.05 [ 0.02, 0.07] |
| | ALSPAC Parent reported low prosociality - age 13 | 0.01 [-0.02, 0.03] |
| | ALSPAC Parent reported low prosociality - age 17 | 0.01 [-0.02, 0.04] |

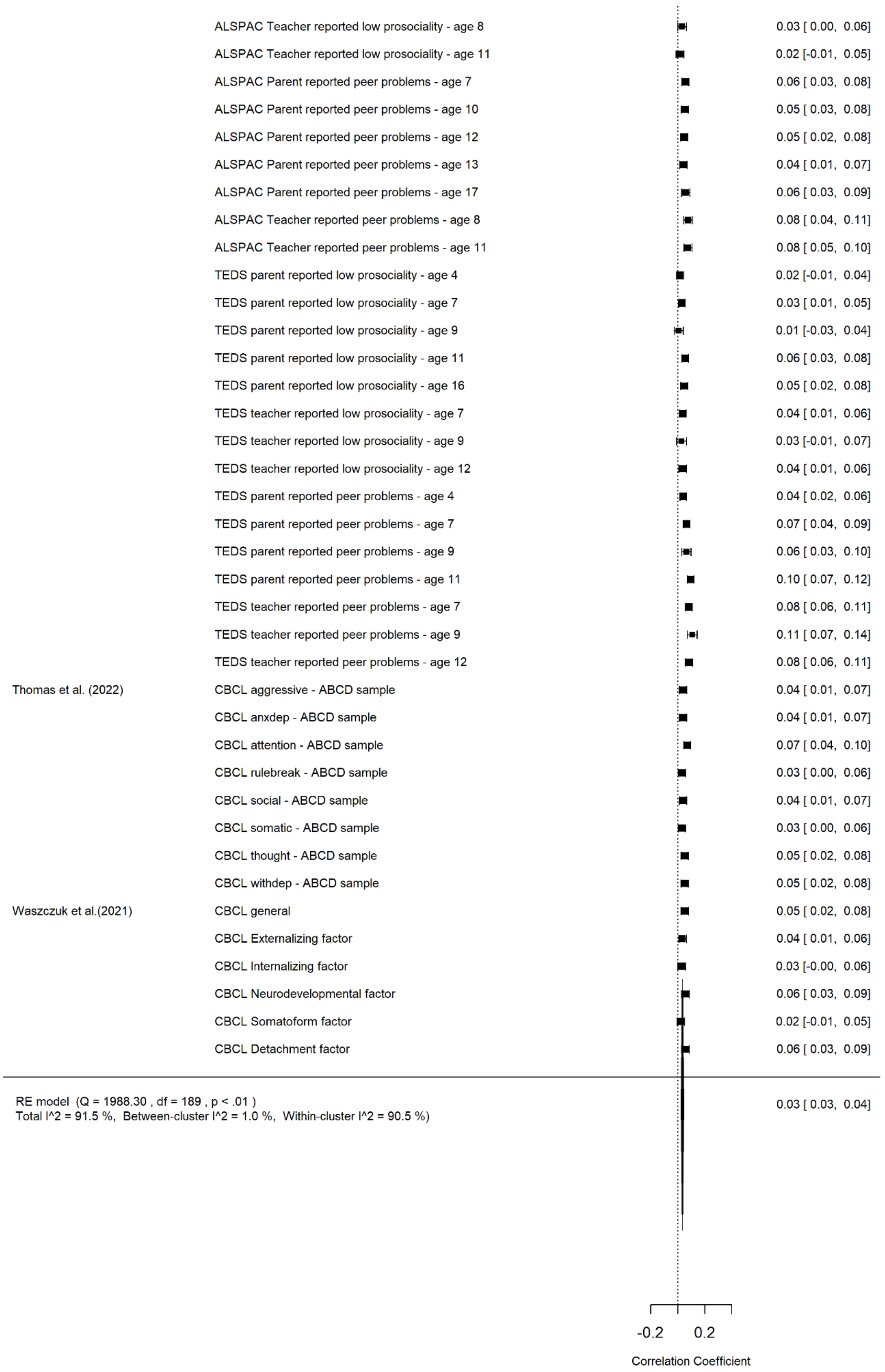

sFigure 8. Multi-level Meta-Analysis Results on the Association between Autism Polygenic Score and General Psychopathology.

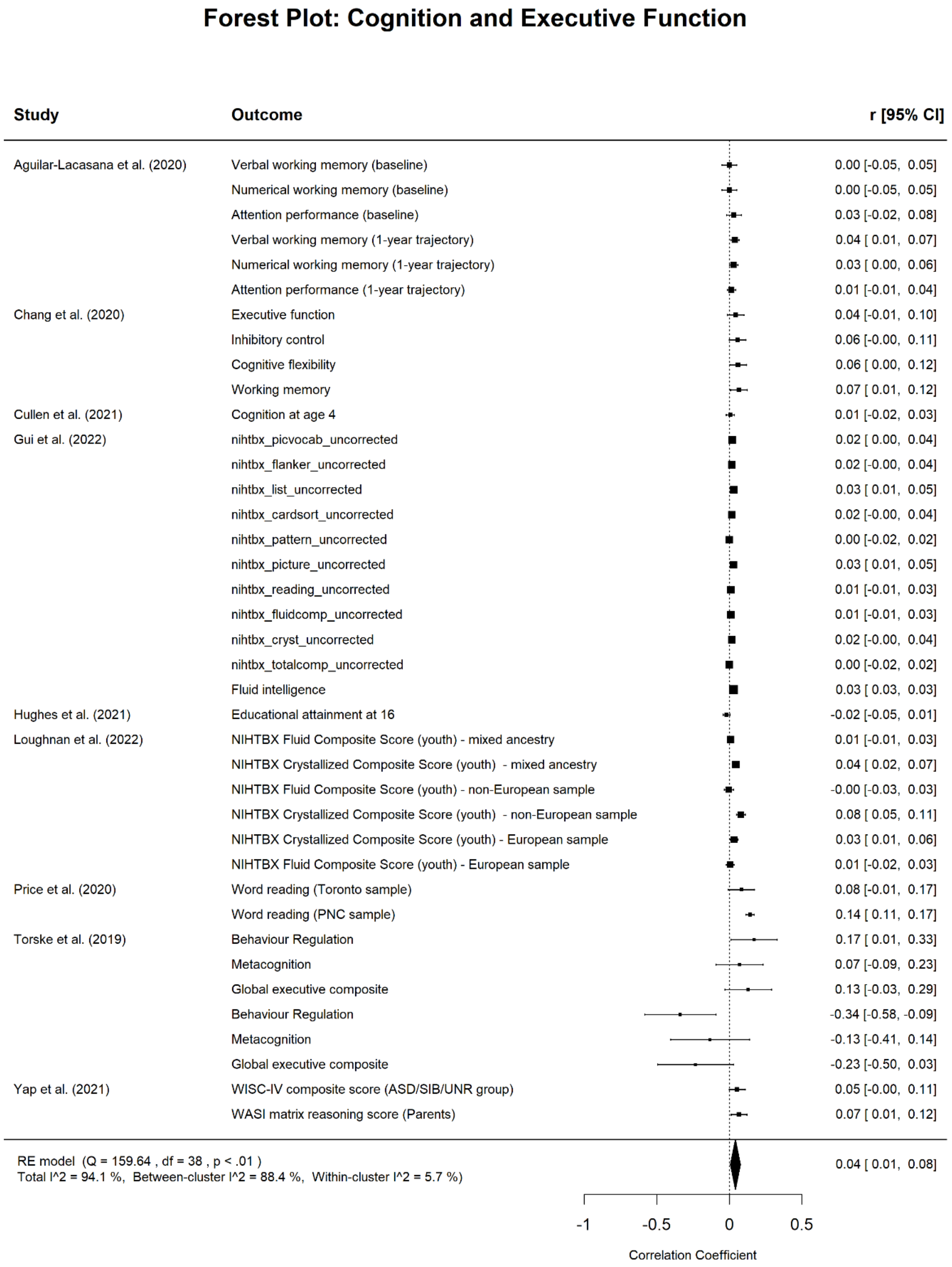

sFigure 9. Multi-level Meta-Analysis Results on the Association between Autism Polygenic Score and Cognition and Executive Function.

### Forest Plot: Physical Wellbeing

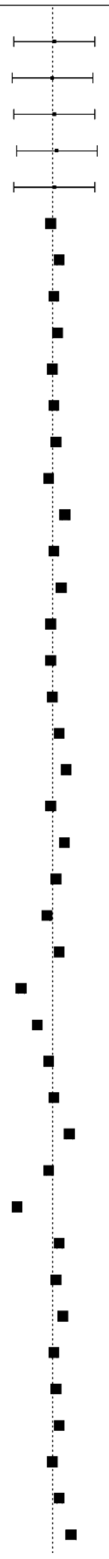

| Study | Outcome | r [95% CI] |
|---|---|---|
| Dennison et al. (2021) | Overall activity | 0.01 [-0.22, 0.24] |
| | Moderate activity | 0.00 [-0.23, 0.23] |
| | Walking activity | 0.01 [-0.22, 0.24] |
| | Sedentary activity | 0.02 [-0.20, 0.25] |
| | Sleep activity | 0.01 [-0.22, 0.24] |
| Havdahl et al. (2022) | Cigarette smoking (mother) | -0.01 [-0.03, 0.01] |
| | Alcohol consumption (mother) | 0.04 [ 0.02, 0.06] |
| | Binge drinking (mother) | 0.01 [-0.01, 0.03] |
| | Coffee consumption (mother) | 0.03 [ 0.01, 0.05] |
| | Binge coffee consumption (mother) | 0.00 [-0.02, 0.02] |
| | No supplements taken (mother) | 0.01 [-0.01, 0.03] |
| | Folate supplement before pregnancy (mother) | 0.02 [ 0.00, 0.04] |
| | Folate supplement during pregnancy (mother) | -0.02 [-0.04, -0.00] |
| | Body mass index before pregnancy (mother) | 0.07 [ 0.05, 0.09] |
| | Weight gain (mother) | 0.01 [-0.01, 0.03] |
| | Type 2 diabetes (mother) | 0.05 [ 0.03, 0.07] |
| | High blood pressure (mother) | -0.01 [-0.03, 0.01] |
| | Hyperthyroidism/hypothyroidism (mother) | -0.01 [-0.03, 0.01] |
| | Upper respiratory tract infections (mother) | 0.00 [-0.02, 0.02] |
| | Lower respiratory tract infections (mother) | 0.04 [ 0.02, 0.06] |
| | Urinary tract infection (mother) | 0.08 [ 0.06, 0.09] |
| | Fever (mother) | -0.01 [-0.03, 0.01] |
| | Asthma (mother) | 0.07 [ 0.05, 0.08] |
| | Psoriasis (mother) | 0.02 [ 0.00, 0.04] |
| | Type 1 diabetes (mother) | -0.03 [-0.05, -0.01] |
| | Other autoimmune disease (mother) | 0.04 [ 0.02, 0.06] |
| | Vaginal bleeding (mother) | -0.17 [-0.19, -0.16] |
| | Vitamin B12 insufficiency (mother) | -0.08 [-0.10, -0.07] |
| | Anemia/low hemoglobin during pregnancy (mother) | -0.02 [-0.04, -0.00] |
| | Pain (mother) | 0.01 [-0.01, 0.03] |
| | Migraine (mother) | 0.10 [ 0.08, 0.11] |
| | Headache (mother) | -0.02 [-0.04, -0.00] |
| | Epilepsy (mother) | -0.20 [-0.21, -0.18] |
| | Pain medication (mother) | 0.04 [ 0.02, 0.06] |
| | Paracetamol use (mother) | 0.02 [ 0.00, 0.04] |
| | Ibuprofen use (mother) | 0.06 [ 0.04, 0.07] |
| | Fever medication (mother) | 0.01 [-0.01, 0.03] |
| | Pain or fever medication (mother) | 0.02 [ 0.00, 0.04] |
| | Ever smoked (father) | 0.04 [ 0.02, 0.06] |
| | Cigarette smoking (father) | 0.00 [-0.02, 0.02] |
| | Ever drank alcohol (Father) | 0.04 [ 0.02, 0.06] |
| | Binge drinking before pregnancy (father) | 0.10 [ 0.08, 0.13] |

| | |
|---|---|
| No supplements taken (father) | 0.02 [-0.01, 0.05] |
| BMI (father) | -0.08 [-0.11, -0.05] |
| Diabetes (father) | 0.02 [ 0.00, 0.04] |
| High blood pressure (father) | -0.02 [-0.04, -0.00] |
| Asthma (father) | 0.03 [ 0.01, 0.05] |
| Psoriasis (father) | 0.02 [ 0.00, 0.04] |
| Other autoimmune disease (father) | 0.04 [ 0.02, 0.06] |
| Pain (father) | 0.00 [-0.02, 0.02] |
| Headache (father) | 0.03 [ 0.01, 0.05] |
| Migraine (father) | -0.01 [-0.03, 0.01] |
| Cigarette smoking - ever (mother) | 0.02 [ 0.00, 0.04] |
| Cigarette smoking - during pregnancy (mother) | 0.04 [ 0.02, 0.06] |
| Cigarette smoking - trimester 1 (mother) | -0.01 [-0.03, 0.01] |
| Cigarette smoking - trimester 2 (mother) | 0.01 [-0.01, 0.03] |
| Cigarette smoking - trimester 3 (mother) | 0.00 [-0.02, 0.02] |
| Alcohol consumption - ever (mother) | 0.07 [ 0.05, 0.08] |
| Alcohol consumption - during pregnancy (mother) | 0.04 [ 0.02, 0.06] |
| Alcohol consumption - trimester 1 (mother) | 0.05 [ 0.03, 0.07] |
| Alcohol consumption - trimester 2 (mother) | 0.07 [ 0.05, 0.08] |
| Alcohol consumption - trimester 3 (mother) | 0.06 [ 0.04, 0.08] |
| High blood pressure - before pregnancy (mother) | 0.06 [ 0.04, 0.07] |
| High blood pressure - during pregnancy (mother) | -0.01 [-0.03, 0.01] |
| High blood pressure - trimester 1 (mother) | 0.07 [ 0.05, 0.08] |
| High blood pressure - trimester 2 (mother) | -0.04 [-0.06, -0.02] |
| High blood pressure - trimester 3 (mother) | 0.00 [-0.02, 0.02] |
| Vaginal bleeding - during pregnancy (mother) | -0.17 [-0.19, -0.16] |
| Vaginal bleeding - trimester 1 (mother) | 0.00 [-0.02, 0.02] |
| Vaginal bleeding - trimester 2 (mother) | 0.10 [ 0.08, 0.11] |
| Vaginal bleeding - trimester 3 (mother) | 0.04 [ 0.02, 0.06] |
| Urinary tract infections - before pregnancy (mother) | 0.01 [-0.01, 0.03] |
| Urinary tract infections - during pregnancy (mother) | 0.08 [ 0.06, 0.09] |
| Urinary tract infections - trimester 1 (mother) | 0.08 [ 0.06, 0.09] |
| Urinary tract infections - trimester 2 (mother) | 0.11 [ 0.10, 0.13] |
| Urinary tract infections - trimester 3 (mother) | 0.07 [ 0.05, 0.08] |
| Fever - during pregnancy (mother) | -0.01 [-0.03, 0.01] |
| fever - Trimester 1 (mother) | 0.04 [ 0.02, 0.06] |
| fever - Trimester 2 (mother) | 0.00 [-0.02, 0.02] |
| Fever - Trimester 3 (mother) | -0.02 [-0.04, -0.00] |
| Pain - during pregnancy (mother) | 0.01 [-0.01, 0.03] |
| Pain - Trimester 1 (mother) | 0.03 [ 0.01, 0.05] |
| Pain - Trimester 2 (mother) | 0.01 [-0.01, 0.03] |
| Pain - Trimester 3 (mother) | 0.01 [-0.01, 0.03] |
| Migraine - ever (mother) | 0.05 [ 0.03, 0.07] |
| Migraine - before pregnancy (mother) | 0.05 [ 0.03, 0.07] |
| Migraine - after pregnancy (mother) | 0.10 [ 0.08, 0.11] |
| Paracetamol use - during pregnancy (mother) | 0.02 [ 0.00, 0.04] |
| Paracetamol use - trimester 1-2 (mother) | 0.01 [-0.01, 0.03] |
| Paracetamol use - trimester 2-3 (mother) | 0.00 [-0.02, 0.02] |
| Ibuprofen use - during pregnancy (mother) | 0.06 [ 0.04, 0.07] |
| Ibuprofen use - trimester 1-2 (mother) | 0.07 [ 0.05, 0.08] |
| Ibuprofen use - trimester 2-3 (mother) | -0.16 [-0.18, -0.15] |

| | | |
|---|---|---|
| Hunjan et al. (2021) | Alcohol intake | -0.00 [-0.01, 0.00] |
| | Calcium intake | 0.00 [-0.00, 0.01] |
| | Carbohydrate intake | 0.00 [-0.00, 0.01] |
| | Carotene intake | 0.00 [-0.00, 0.00] |
| | Fibre intake | 0.00 [-0.00, 0.01] |
| | Fat intake | 0.00 [ 0.00, 0.01] |
| | Folate intake | -0.00 [-0.01, 0.00] |
| | Food weight | -0.00 [-0.01, 0.00] |
| | Iron intake | 0.00 [-0.00, 0.01] |
| | Protein intake | 0.00 [-0.00, 0.01] |
| | Vitamin B12 intake | -0.00 [-0.01, 0.00] |
| | Vitamin B6 intake | -0.01 [-0.01, -0.00] |
| | Vitamin C intake | -0.00 [-0.01, 0.00] |
| | Vitamin D intake | -0.00 [-0.01, 0.00] |
| | Vitamin E intake | 0.00 [-0.00, 0.01] |
| Leppert et al. (2019) | self-reported smoking of mother during first semester | 0.04 [ 0.02, 0.06] |
| | self-reported smoking of mother during third semester | 0.06 [ 0.03, 0.08] |
| | self-reported alcohol consumption of mother during first semester | 0.00 [-0.02, 0.02] |
| | self-reported alcohol consumption of mother during third semester | 0.05 [ 0.02, 0.08] |
| | self-reported binge drinking of mother during first semester | 0.06 [ 0.04, 0.08] |
| | Mother taking iron supplements during pregnancy | 0.00 [-0.02, 0.02] |
| | Mother taking zinc supplements during pregnancy | 0.12 [ 0.10, 0.14] |
| | Mother taking folic acid supplements during pregnancy | 0.07 [ 0.04, 0.09] |
| | Mother taking vitamin supplements during pregnancy | 0.05 [ 0.03, 0.07] |
| | Mother's use of acetaminophen in early pregnancy | 0.00 [-0.02, 0.02] |
| | Mother's use of acetaminophen in late pregnancy | -0.01 [-0.03, 0.01] |
| | Mother's use of antidepressants | 0.11 [ 0.09, 0.14] |
| | Mother's prepregnancy BMI | -0.03 [-0.05, -0.01] |
| | Mother - Ever had diabetes | 0.14 [ 0.12, 0.16] |
| | Mother - Gestational diabetes | 0.17 [ 0.14, 0.19] |
| | Mother - Ever had hypertension | -0.06 [-0.09, -0.04] |
| | Mother - Gestational hypertension | -0.02 [-0.04, 0.00] |
| | Mother -Preeclampsia | 0.17 [ 0.14, 0.19] |
| | Mother - Vaginal bleeding during pregnancy | 0.02 [-0.00, 0.04] |
| | Mother - Any infection in pregnancy | 0.06 [ 0.03, 0.08] |
| | Mother - Ever had rheumatism | 0.17 [ 0.14, 0.19] |
| | Mother - Ever had psoriasis | 0.10 [ 0.08, 0.13] |
| | Mother - Bloodmarker Vitamin D | -0.01 [-0.04, 0.02] |
| | Mother - Bloodmarker selenium | 0.01 [-0.03, 0.05] |
| | Mother - Bloodmarker mercury | 0.02 [-0.02, 0.06] |
| | Mother - Bloodmarker cadmium | 0.01 [-0.03, 0.05] |
| | Mother - Bloodmarker lead | 0.05 [ 0.01, 0.09] |
| | Ceserean delivery - maternal PGS | 0.03 [ 0.01, 0.05] |
| | Low birth weight - maternal PGS | 0.00 [-0.02, 0.02] |
| | Preterm delivery - maternal PGS | -0.04 [-0.06, -0.02] |
| | Hypoxia - maternal PGS | 0.04 [ 0.01, 0.07] |
| | Low Apgar score - at 1 min - maternal PGS | 0.02 [-0.01, 0.05] |
| | Low Apgar score - at 5 min - maternal PGS | 0.04 [ 0.01, 0.07] |
| | Breastfeeding at 1 month old - maternal PGS | 0.05 [ 0.02, 0.07] |
| | Ceserean delivery - child PGS | -0.03 [-0.05, -0.01] |
| | Low birth weight - child PGS | -0.03 [-0.05, -0.01] |
| | Preterm delivery - child PGS | -0.06 [-0.08, -0.04] |
| | Hypoxia - child PGS | -0.01 [-0.04, 0.02] |

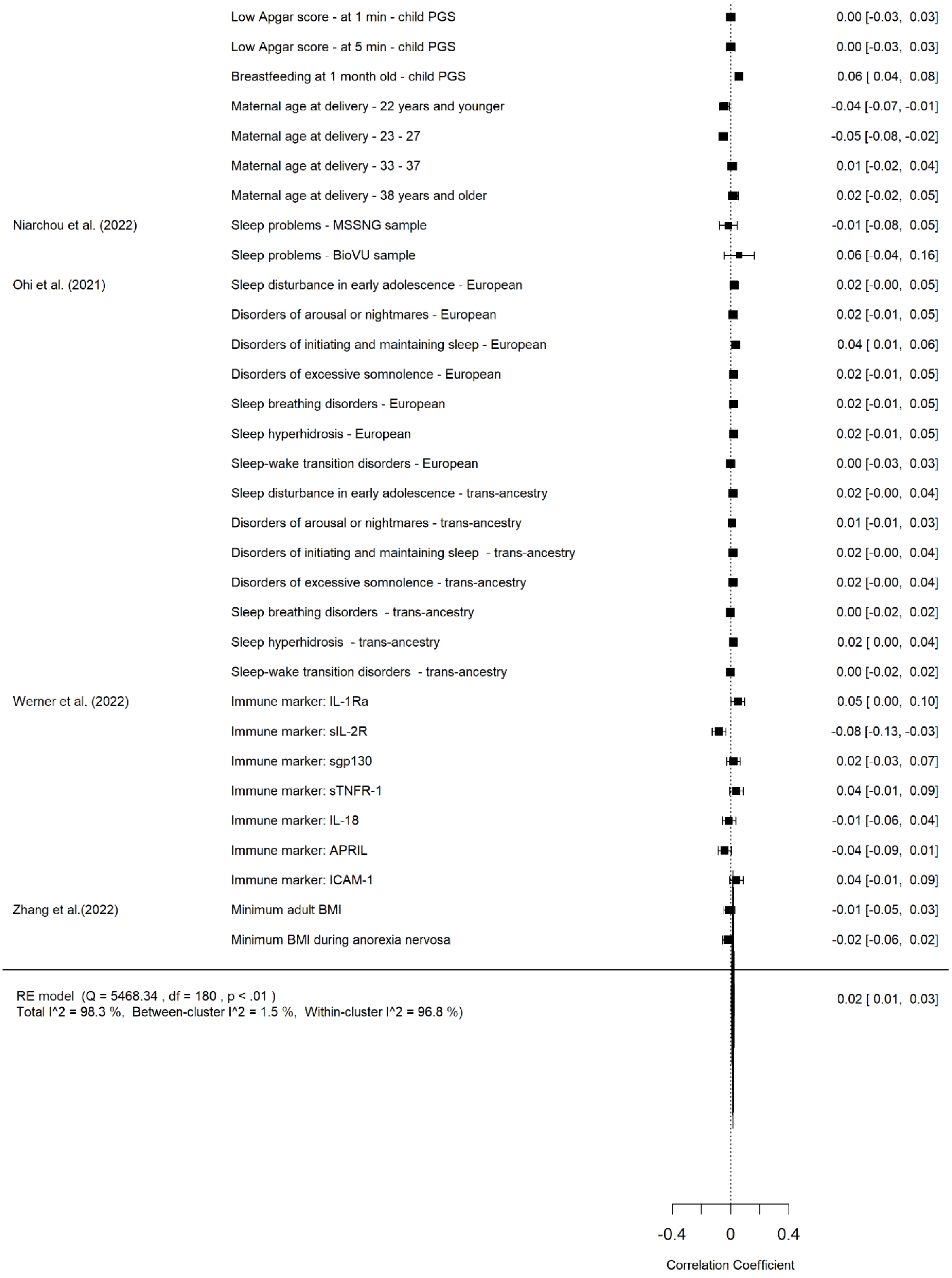

sFigure 10. Multi-level Meta-Analysis Results on the Association between Autism Polygenic Score and Physical Wellbeing.

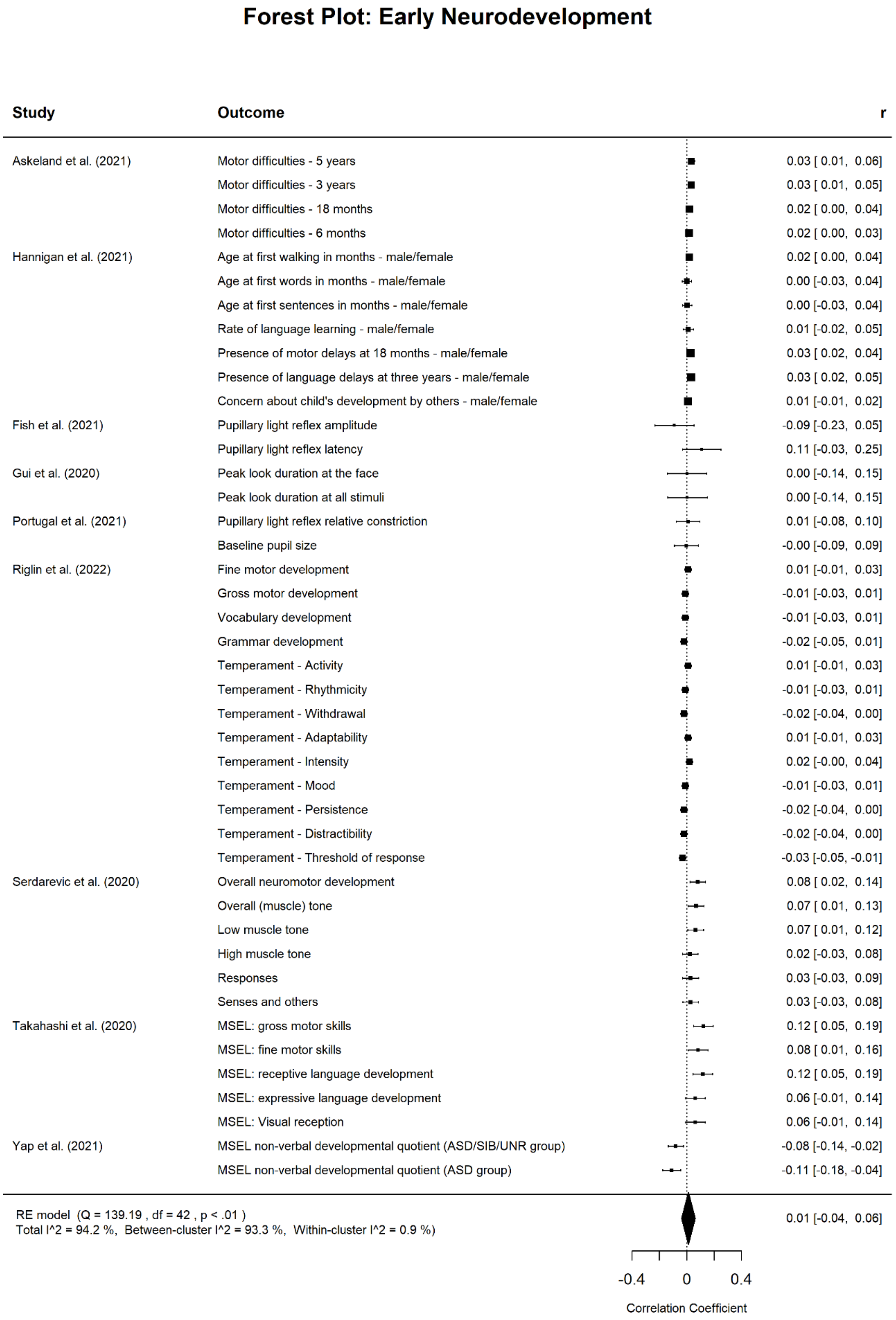

sFigure 11. Multi-level Meta-Analysis Results on the Association between Autism Polygenic Score and Early Neurodevelopment.

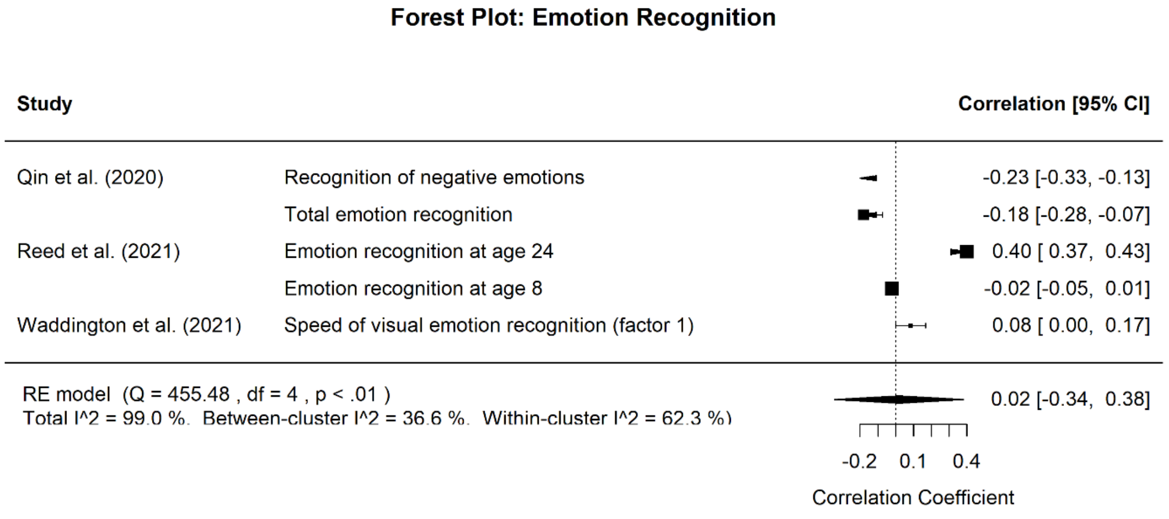

sFigure 12. Multi-level Meta-Analysis Results on the Association between Autism Polygenic Score and Emotion Recognition.

### Forest Plot: Brain Measures

| Study | Outcome | r [95% CI] |
|---|---|---|
| Alemany et al. (2021) | Left hemisphere, lateral occipital, gyrification | 0.02 [-0.04, 0.08] |
| | Left hemisphere, superior parietal, gyrification | -0.08 [-0.14, -0.02] |
| | Right hemisphere, inferior parietal, gyrification | 0.02 [-0.04, 0.08] |
| | Right hemisphere, superior frontal, cortical thickness | -0.03 [-0.09, 0.03] |
| | Left hemisphere, inferior temporal, surface area | -0.03 [-0.09, 0.03] |
| | Left hemisphere, rostral middle frontal, surface area | 0.04 [-0.02, 0.10] |
| Ecker et al. (2021) | Total neuroanatomical atypicality index (tAIs) | 0.11 [ 0.02, 0.20] |
| Gui et al. (2021) | N290 latency to face vs nonface stimuli | 0.20 [ 0.08, 0.33] |
| Jansen et al. (2019) | Global fractional anisotropy | 0.08 [ 0.02, 0.13] |
| | Global mean diffusivity | -0.04 [-0.09, 0.02] |
| | Left Superior longitudinal fasciculus (SLF) fractional anisotropy | 0.05 [-0.01, 0.11] |
| | Left Inferior longitudinal fasciculus (ILF) fractional anisotropy | 0.04 [-0.02, 0.10] |
| | Left Uncinate fasciculus (UNC) fractional anisotropy | 0.04 [-0.02, 0.10] |
| | Left Cingulum bundle cingulate (CGC) fractional anisotropy | 0.07 [ 0.02, 0.13] |
| | Left Corticospinal tract (CST) fractional anisotropy | 0.05 [-0.01, 0.11] |
| | Forceps major (FMA) fractional anisotropy | 0.03 [-0.03, 0.08] |
| | Forceps minor (FMI) fractional anisotropy | 0.04 [-0.02, 0.09] |
| | Right Superior longitudinal fasciculus (SLF) fractional anisotropy | 0.06 [ 0.00, 0.12] |
| | Right Inferior longitudinal fasciculus (ILF) fractional anisotropy | 0.04 [-0.01, 0.10] |
| | Right Uncinate fasciculus (UNC) fractional anisotropy | 0.05 [-0.01, 0.11] |
| | Right Cingulum bundle cingulate (CGC) fractional anisotropy | 0.05 [-0.00, 0.11] |
| | Right Corticospinal tract (CST) fractional anisotropy | 0.05 [-0.01, 0.11] |
| | Left Superior longitudinal fasciculus (SLF) mean diffusivity | -0.02 [-0.08, 0.03] |
| | Left Inferior longitudinal fasciculus (ILF) mean diffusivity | -0.00 [-0.06, 0.06] |
| | Left Uncinate fasciculus (UNC) mean diffusivity | -0.03 [-0.09, 0.03] |
| | Left Cingulum bundle cingulate (CGC) mean diffusivity | -0.02 [-0.08, 0.03] |
| | Left Corticospinal tract (CST) mean diffusivity | -0.02 [-0.08, 0.04] |
| | Forceps major (FMA) mean diffusivity | -0.01 [-0.07, 0.04] |
| | Forceps minor (FMI) mean diffusivity | -0.07 [-0.13, -0.01] |
| | Right Superior longitudinal fasciculus (SLF) mean diffusivity | -0.03 [-0.09, 0.03] |
| | Right Inferior longitudinal fasciculus (ILF) mean diffusivity | -0.04 [-0.10, 0.02] |
| | Right Uncinate fasciculus (UNC) mean diffusivity | -0.03 [-0.09, 0.03] |
| | Right Cingulum bundle cingulate (CGC) mean diffusivity | -0.03 [-0.08, 0.03] |
| | Right Corticospinal tract (CST) mean diffusivity | -0.01 [-0.07, 0.05] |

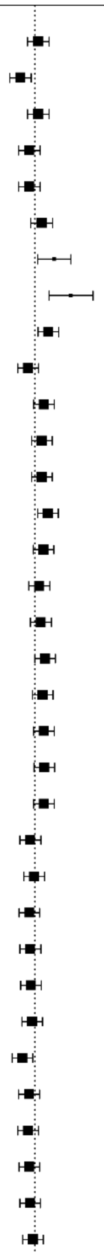

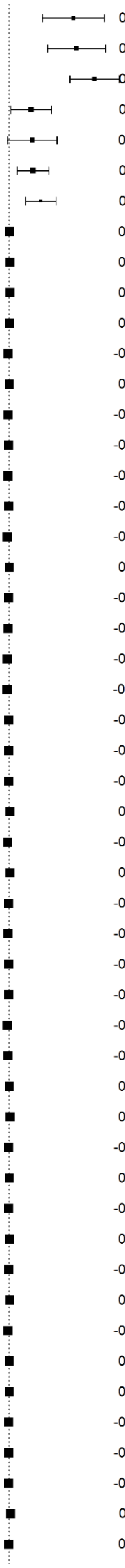

| | | |
|---|---|---|
| Lawrence et al. (2022) | Increased functional connectivity with increased PGS | 0.43 [ 0.22, 0.64] |
| | Increased functional connectivity with increased PGS | 0.45 [ 0.26, 0.64] |
| | Decreased functional connectivity with increased PGS | 0.57 [ 0.41, 0.73] |
| Mason et al. (2022) | N170 latency to face response (ASD cases) | 0.15 [ 0.01, 0.28] |
| | N170 latency to face response (TD) | 0.15 [-0.01, 0.32] |
| | N170 latency to face response (AAll) | 0.16 [ 0.06, 0.27] |
| Qin et al. (2020) | ALFF left amygdala | 0.21 [ 0.11, 0.31] |
| Sha et al. (2021) | Left area bankssts | 0.00 [-0.01, 0.01] |
| | Left area caudalanteriorcingulate | 0.01 [-0.00, 0.02] |
| | Left area cuneus | 0.01 [-0.01, 0.02] |
| | Left area entorhinal | 0.00 [-0.01, 0.02] |
| | Left area fusiform | -0.01 [-0.02, 0.01] |
| | Left area inferiorparietal | 0.00 [-0.01, 0.01] |
| | Left area inferiortemporal | -0.00 [-0.01, 0.01] |
| | Left area insula | -0.00 [-0.01, 0.01] |
| | Left area isthmuscingulate | -0.01 [-0.02, 0.00] |
| | Left area lateralorbitofrontal | -0.00 [-0.01, 0.01] |
| | Left area lingual | -0.01 [-0.02, 0.00] |
| | Left area medialorbitofrontal | 0.00 [-0.01, 0.01] |
| | Left area middletemporal | -0.00 [-0.01, 0.01] |
| | Left area paracentral | -0.00 [-0.02, 0.01] |
| | Left area parahippocampal | -0.01 [-0.02, 0.00] |
| | Left area parsopercularis | -0.01 [-0.02, -0.00] |
| | Left area parsorbitalis | -0.00 [-0.01, 0.01] |
| | Left area parstriangularis | -0.00 [-0.01, 0.01] |
| | Left area pericalcarine | -0.00 [-0.01, 0.01] |
| | Left area posteriorcingulate | 0.01 [-0.00, 0.02] |
| | Left area precuneus | -0.01 [-0.02, 0.00] |
| | Left area rostralanteriorcingulate | 0.01 [-0.00, 0.02] |
| | Left area rostralmiddlefrontal | -0.00 [-0.01, 0.01] |
| | Left area superiorfrontal | -0.01 [-0.02, 0.00] |
| | Left area superiorparietal | -0.00 [-0.01, 0.01] |
| | Left area superiortemporal | -0.00 [-0.01, 0.01] |
| | Left area supramarginal | -0.01 [-0.02, 0.00] |
| | Left area transversetemporal | -0.01 [-0.02, 0.00] |
| | Left thickness entorhinal | 0.00 [-0.01, 0.01] |
| | Left thickness inferiortemporal | 0.01 [-0.00, 0.02] |
| | Left thickness isthmuscingulate | -0.00 [-0.01, 0.01] |
| | Left thickness parahippocampal | 0.00 [-0.01, 0.01] |
| | Left thickness posteriorcingulate | -0.00 [-0.01, 0.01] |
| | Left thickness precentral | 0.00 [-0.01, 0.01] |
| | Left thickness rostralanteriorcingulate | -0.00 [-0.01, 0.01] |
| | Left thickness supramarginal | 0.01 [-0.01, 0.02] |
| | Left volume thalamus | -0.01 [-0.02, 0.00] |
| | Left volume caudate | 0.00 [-0.01, 0.01] |
| | Left volume putamen | 0.00 [-0.01, 0.01] |
| | Left volume pallidum | -0.00 [-0.01, 0.01] |
| | Left volume hippocampus | -0.00 [-0.01, 0.01] |
| | Left volume accumbens | -0.00 [-0.01, 0.01] |
| | Right area bankssts | 0.01 [ 0.00, 0.02] |
| | Right area caudalanteriorcingulate | 0.00 [-0.01, 0.01] |

| | |
|---|---|
| Right area cuneus | -0.00 [-0.01, 0.01] |
| Right area entorhinal | 0.01 [-0.00, 0.02] |
| Right area fusiform | -0.01 [-0.02, 0.00] |
| Right area inferiorparietal | 0.01 [-0.00, 0.02] |
| Right area inferiortemporal | -0.00 [-0.01, 0.01] |
| Right area insula | 0.00 [-0.01, 0.01] |
| Right area isthmuscingulate | -0.01 [-0.02, 0.01] |
| Right area lateralorbitofrontal | -0.00 [-0.01, 0.01] |
| Right area lingual | -0.01 [-0.02, 0.01] |
| Right area medialorbitofrontal | -0.00 [-0.01, 0.01] |
| Right area middletemporal | -0.00 [-0.01, 0.01] |
| Right area paracentral | 0.01 [-0.01, 0.02] |
| Right area parahippocampal | -0.00 [-0.01, 0.01] |
| Right area parsopercularis | -0.00 [-0.01, 0.01] |
| Right area parsorbitalis | -0.00 [-0.01, 0.01] |
| Right area parstriangularis | -0.00 [-0.01, 0.01] |
| Right area pericalcarine | -0.00 [-0.01, 0.01] |
| Right area posteriorcingulate | -0.00 [-0.01, 0.01] |
| Right area precuneus | -0.01 [-0.02, 0.00] |
| Right area rostralanteriorcingulate | 0.00 [-0.01, 0.01] |
| Right area rostralmiddlefrontal | -0.00 [-0.01, 0.01] |
| Right area superiorfrontal | -0.01 [-0.02, 0.00] |
| Right area superiorparietal | -0.00 [-0.01, 0.01] |
| Right area superiortemporal | -0.00 [-0.01, 0.01] |
| Right area supramarginal | -0.01 [-0.02, 0.00] |
| Right area transversetemporal | -0.01 [-0.02, -0.00] |
| Right thickness entorhinal | -0.01 [-0.02, 0.00] |
| Right thickness inferiortemporal | 0.00 [-0.01, 0.01] |
| Right thickness isthmuscingulate | -0.00 [-0.01, 0.01] |
| Right thickness parahippocampal | 0.01 [-0.00, 0.02] |
| Right thickness posteriorcingulate | -0.01 [-0.02, 0.00] |
| Right thickness precentral | 0.00 [-0.01, 0.01] |
| Right thickness rostralanteriorcingulate | -0.00 [-0.01, 0.01] |
| Right thickness supramarginal | 0.01 [-0.01, 0.02] |
| Right volume thalamus | -0.01 [-0.02, 0.00] |
| Right volume caudate | -0.00 [-0.01, 0.01] |
| Right volume putamen | -0.00 [-0.01, 0.01] |
| Right volume pallidum | 0.00 [-0.01, 0.01] |
| Right volume hippocampus | -0.00 [-0.01, 0.01] |
| Right volume accumbens | -0.00 [-0.02, 0.01] |
| Multivarate brain asymmetry | 0.03 [ 0.02, 0.04] |
| Regional asymmetry surface area bankssts | -0.20 [-0.21, -0.19] |
| Regional asymmetry surface area caudalanteriorcingulate | 0.15 [ 0.14, 0.16] |
| Regional asymmetry surface area cuneus | 0.36 [ 0.36, 0.37] |
| Regional asymmetry surface area entorhinal | -0.05 [-0.06, -0.04] |
| Regional asymmetry surface area fusiform | 0.06 [ 0.05, 0.07] |
| Regional asymmetry surface area inferiorparietal | -0.19 [-0.20, -0.18] |
| Regional asymmetry surface area inferiortemporal | -0.05 [-0.06, -0.03] |
| Regional asymmetry surface area insula | -0.06 [-0.07, -0.05] |
| Regional asymmetry surface area isthmuscingulate | -0.02 [-0.03, -0.01] |
| Regional asymmetry surface area lateralorbitofrontal | 0.02 [ 0.01, 0.03] |
| Regional asymmetry surface area lingual | -0.15 [-0.16, -0.14] |

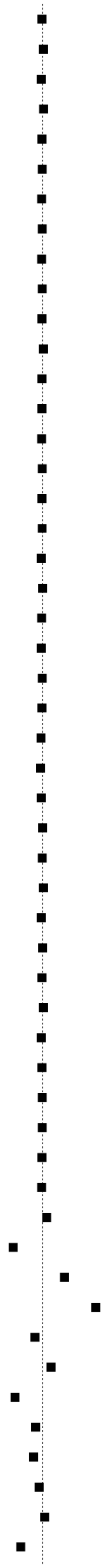

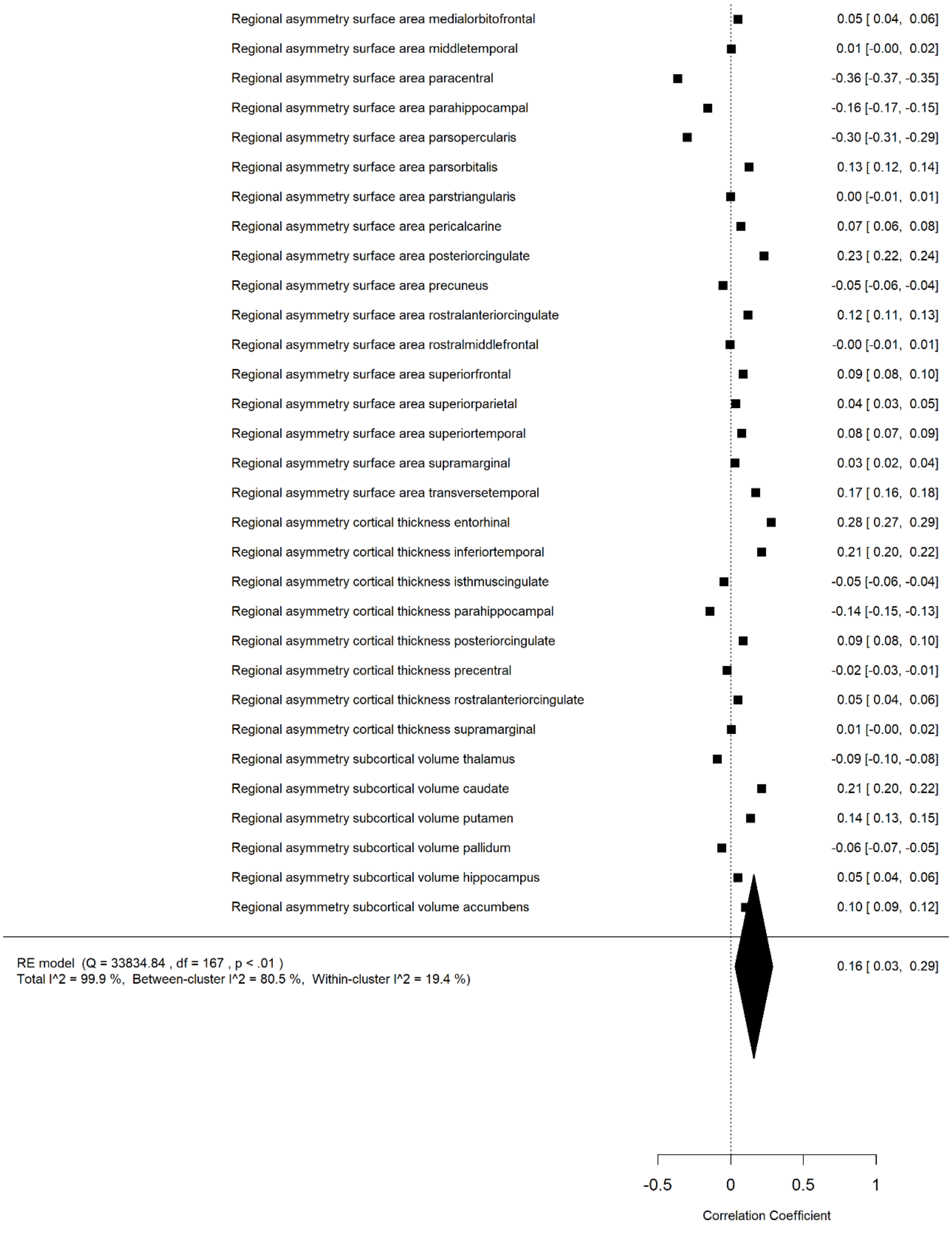

sFigure 13. Multi-level Meta-Analysis Results on the Association between Autism Polygenic Score and Brain Measures.

#### References

1. Gui Y, Zhou X, Wang Z, et al. Sex-specific genetic association between psychiatric disorders and cognition, behavior and brain imaging in children and adults. *Transl Psychiatry*. 2022;12(1):1-8. doi:10.1038/s41398-022-02041-6

2. Loughnan RJ, Palmer CE, Makowski C, et al. Unique prediction of developmental psychopathology from genetic and familial risk. *J Child Psychol Psychiatry*. 2022;63(12):1631-1643. doi:10.1111/jcpp.13649

3. Pat N, Riglin L, Anney R, et al. Motivation and Cognitive Abilities as Mediators Between Polygenic Scores and Psychopathology in Children. *J Am Acad Child Adolesc Psychiatry*. 2022;61(6):782-795.e3. doi:10.1016/j.jaac.2021.08.019

4. Riglin L, Thapar AK, Leppert B, et al. Using Genetics to Examine a General Liability to Childhood Psychopathology. *Behav Genet*. 2020;50(4):213-220. doi:10.1007/S10519-019-09985-4/TABLES/2

5. Schlag F, Allegrini AG, Buitelaar J, et al. Polygenic risk for mental disorder reveals distinct association profiles across social behaviour in the general population. *Mol Psychiatry*. 2022;27(3):1588-1598. doi:10.1038/s41380-021-01419-0

6. Thomas TR, Koomar T, Casten LG, Tener AJ, Bahl E, Michaelson JJ. Clinical autism subscales have common genetic liabilities that are heritable, pleiotropic, and generalizable to the general population. *Transl Psychiatry*. 2022;12(1):247. doi:10.1038/s41398-022-01982-2

7. Waszczuk MA, Miao J, Docherty AR, et al. General v. specific vulnerabilities: polygenic risk scores and higher-order psychopathology dimensions in the Adolescent Brain Cognitive Development (ABCD) Study. *Psychol Med*. Published online 2021:1-10. doi:10.1017/S0033291721003639

8. Achenbach TM. Achenbach System of Empirically Based Assessment (ASEBA). In: *The Encyclopedia of Clinical Psychology*. John Wiley & Sons, Ltd; 2015:1-8. doi:10.1002/9781118625392.wbecp150

9. Youngstrom EA, Findling RL, Danielson CK, Calabrese JR. Discriminative validity of parent report of hypomanic and depressive symptoms on the General Behavior Inventory. *Psychol Assess*. 2001;13(2):267-276. doi:10.1037/1040-3590.13.2.267

10. Whiteside SP, Lynam DR, Miller JD, Reynolds SK. Validation of the UPPS impulsive behaviour scale: a four-factor model of impulsivity. *Eur J Personal*. 2005;19(7):559-574. doi:10.1002/per.556

11. Carver CS, White TL. Behavioral inhibition, behavioral activation, and affective responses to impending reward and punishment: The BIS/BAS Scales. *J Pers Soc Psychol*. 1994;67(2):319-333. doi:10.1037/0022-3514.67.2.319

12. Kaufman J, Birmaher B, Brent D, et al. Schedule for Affective Disorders and Schizophrenia for School-Age Children-Present and Lifetime Version (K-SADS-PL): Initial Reliability and Validity Data. *J Am Acad Child Adolesc Psychiatry*. 1997;36(7):980-988. doi:10.1097/00004583-199707000-00021

13. Goodman R. The Strengths and Difficulties Questionnaire: A Research Note. *J Child Psychol Psychiatry*. 1997;38(5):581-586. doi:10.1111/j.1469-7610.1997.tb01545.x

14. Aguilar-Lacasaña S, Vilor-Tejedor N, Jansen PR, et al. Polygenic risk for ADHD and ASD and their relation with cognitive measures in school children. *Psychol Med*. 2022;52(7):1356-1364. doi:10.1017/S0033291720003189

15. Chang S, Yang L, Wang Y, Faraone SV. Shared polygenic risk for ADHD, executive dysfunction and other psychiatric disorders. *Transl Psychiatry 2020 101*. 2020;10(1):1-9. doi:10.1038/s41398-020-00872-9

16. Cullen H, Selzam S, Dimitrakopoulou K, Plomin R, Edwards AD. Greater genetic risk for adult psychiatric diseases increases vulnerability to adverse outcome after preterm birth. *Sci Rep 2021 111*. 2021;11(1):1-8. doi:10.1038/s41598-021-90045-5

17. Hughes A, Wade KH, Dickson M, et al. Common health conditions in childhood and adolescence, school absence, and educational attainment: Mendelian randomization study. *Npj Sci Learn 2021 61*. 2021;6(1):1-9. doi:10.1038/s41539-020-00080-6

18. Price KM, Wigg KG, Feng Y, et al. Genome-wide association study of word reading: Overlap with risk genes for neurodevelopmental disorders. *Genes Brain Behav*. 2020;19(6). doi:10.1111/GBB.12648

19. Torske T, Nærland T, Bettella F, et al. Autism spectrum disorder polygenic scores are associated with every day executive function in children admitted for clinical assessment. *Autism Res Off J Int Soc Autism Res*. 2020;13(2):207-220. doi:10.1002/AUR.2207

20. Yap CX, Alvares GA, Henders AK, et al. Analysis of common genetic variation and rare CNVs in the Australian Autism Biobank. *Mol Autism*. 2021;12(1):1-17. doi:10.1186/S13229-020-00407-5/FIGURES/3

21. Gershon RC, Wagster MV, Hendrie HC, Fox NA, Cook KF, Nowinski CJ. NIH Toolbox for Assessment of Neurological and Behavioral Function. *Neurology*. 2013;80(11 Supplement 3):S2-S6. doi:10.1212/WNL.0b013e3182872e5f

22. Dennison CA, Legge SE, Bracher-Smith M, et al. Association of genetic liability for psychiatric disorders with accelerometer-assessed physical activity in the UK Biobank. *PLOS ONE*. 2021;16(3):e0249189. doi:10.1371/JOURNAL.PONE.0249189

23. Havdahl A, Wootton RE, Leppert B, et al. Associations Between Pregnancy-Related Predisposing Factors for Offspring Neurodevelopmental Conditions and Parental Genetic Liability to Attention-Deficit/Hyperactivity Disorder, Autism, and Schizophrenia: The Norwegian Mother, Father and Child Cohort Study (MoBa). *JAMA Psychiatry*. 2022;79(8):799. doi:10.1001/jamapsychiatry.2022.1728

24. Hunjan AK, Hübel C, Lin Y, Eley TC, Breen G. Association between polygenic propensity for psychiatric disorders and nutrient intake. *Commun Biol 2021 41*. 2021;4(1):1-9. doi:10.1038/s42003-021-02469-4

25. Leppert B, Havdahl A, Riglin L, et al. Association of Maternal Neurodevelopmental Risk Alleles With Early-Life Exposures. *JAMA Psychiatry*. 2019;76(8):834-842. doi:10.1001/JAMAPSYCHIATRY.2019.0774

26. Niarchou M, Singer EV, Straub P, Malow BA, Davis LK. Investigating the genetic pathways of insomnia in Autism Spectrum Disorder. *Res Dev Disabil*. 2022;128:104299. doi:10.1016/j.ridd.2022.104299

27. Ohi K, Ochi R, Noda Y, et al. Polygenic risk scores for major psychiatric and neurodevelopmental disorders contribute to sleep disturbance in childhood: Adolescent Brain Cognitive Development (ABCD) Study. *Transl Psychiatry*. 2021;11(1):187. doi:10.1038/s41398-021-01308-8

28. Werner MCF, Wirgenes KV, Shadrin A, et al. Immune marker levels in severe mental disorders: associations with polygenic risk scores of related mental phenotypes and psoriasis. *Transl Psychiatry*. 2022;12(1):38. doi:10.1038/s41398-022-01811-6

29. Zhang R, Birgegård A, Fundín B, et al. Association of autism diagnosis and polygenic scores with eating disorder severity. *Eur Eat Disord Rev*. 2022;30(5):442-458. doi:10.1002/erv.2941

30. Askeland RB, Hannigan LJ, Ask H, et al. Early manifestations of genetic risk for neurodevelopmental disorders. *J Child Psychol Psychiatry*. 2021;63(7):810-819. doi:10.1111/JCPP.13528

31. Fish LA, Nyström P, Gliga T, et al. Development of the pupillary light reflex from 9 to 24 months: association with common autism spectrum disorder (ASD) genetic liability and 3-year ASD diagnosis. *J Child Psychol Psychiatry*. 2021;62(11):1308-1319. doi:10.1111/jcpp.13518

32. Gui A, Mason L, Gliga T, et al. Look duration at the face as a developmental endophenotype: elucidating pathways to autism and ADHD. *Dev Psychopathol*. 2020;32(4):1303-1322. doi:10.1017/S0954579420000930

33. Hannigan LJ, Askeland RB, Ask H, et al. Developmental milestones in early childhood and genetic liability to neurodevelopmental disorders. *Psychol Med*. 2023;53(5):1750-1758. doi:10.1017/S0033291721003330

34. Portugal AM, Taylor MJ, Viktorsson C, et al. Pupil size and pupillary light reflex in early infancy: heritability and link to genetic liability to schizophrenia. *J Child Psychol Psychiatry*. 2022;63(9):1068-1077. doi:10.1111/jcpp.13564

35. Riglin L, Tobarra-Sanchez E, Stergiakouli E, et al. Early manifestations of genetic liability for ADHD, autism and schizophrenia at ages 18 and 24 months. *JCPP Adv*. 2022;2(3):e12093. doi:10.1002/jcv2.12093

36. Serdarevic F, Tiemeier H, Jansen PR, et al. Polygenic Risk Scores for Developmental Disorders, Neuromotor Functioning During Infancy, and Autistic Traits in Childhood. *Biol Psychiatry*. 2020;87(2):132-138. doi:10.1016/J.BIOPSYCH.2019.06.006

37. Takahashi N, Harada T, Nishimura T, et al. Association of Genetic Risks With Autism Spectrum Disorder and Early Neurodevelopmental Delays Among Children Without Intellectual Disability. *JAMA Netw Open*. 2020;3(2):1921644. doi:10.1001/jamanetworkopen.2019.21644

38. Reed ZE, Mahedy L, Jackson A, et al. Examining the bidirectional association between emotion recognition and social autistic traits using observational and genetic analyses. *J Child Psychol Psychiatry*. 2021;62(11):1330-1338. doi:10.1111/JCPP.13395

39. Qin Y, Kang J, Jiao Z, et al. Polygenic risk for autism spectrum disorder affects left amygdala activity and negative emotion in schizophrenia. *Transl Psychiatry 2020 101*. 2020;10(1):1-12. doi:10.1038/s41398-020-01001-2

40. Waddington F, Franke B, Hartman C, Buitelaar JK, Rommelse N, Mota NR. A polygenic risk score analysis of ASD and ADHD across emotion recognition subtypes ADHD, ASD, emotion recognition, polygenic risk score, subtyping. *Am J Med Genet*. 2021;186:401-411. doi:10.1002/ajmg.b.32818

41. Ecker C, Pretzsch CM, Bletsch A, et al. Interindividual Differences in Cortical Thickness and Their Genomic Underpinnings in Autism Spectrum Disorder. *Am J Psychiatry*. 2022;179(3):242-254. doi:10.1176/appi.ajp.2021.20050630

42. Gui A, Meaburn EL, Tye C, Charman T, Johnson MH, Jones EJH. Association of Polygenic Liability for Autism With Face-Sensitive Cortical Responses From Infancy. *JAMA Pediatr*. 2021;175(9):968. doi:10.1001/jamapediatrics.2021.1338

43. Lawrence KE, Hernandez LM, Fuster E, et al. Impact of autism genetic risk on brain connectivity: a mechanism for the female protective effect. *Brain*. 2022;145(1):378-387. doi:10.1093/brain/awab204

44. Sha Z, Schijven D, Francks C. Patterns of brain asymmetry associated with polygenic risks for autism and schizophrenia implicate language and executive functions but not brain masculinization. *Mol Psychiatry*. 2021;26(12):7652-7660. doi:10.1038/s41380-021-01204-z

45. Mason L, Moessnang C, Chatham C, et al. Stratifying the autistic phenotype using electrophysiological indices of social perception. *Sci Transl Med*. 2022;14(658):eabf8987. doi:10.1126/scitranslmed.abf8987

46. Alemany S, Blok E, Jansen PR, Muetzel RL, White T. Brain morphology, autistic traits, and polygenic risk for autism: A population-based neuroimaging study. *Autism Res*. 2021;14(10):2085-2099. doi:10.1002/aur.2576

47. Jansen PR, Muetzel RL, Polderman TJC, et al. Polygenic Scores for Neuropsychiatric Traits and White Matter Microstructure in the Pediatric Population. *Biol Psychiatry Cogn Neurosci Neuroimaging*. 2019;4(3):243-250. doi:10.1016/j.bpsc.2018.07.010

48. Leppert B, Millard LAC, Riglin L, et al. A cross-disorder PRS-pheWAS of 5 major psychiatric disorders in UK Biobank. Zhu X, ed. *PLOS Genet*. 2020;16(5):e1008185. doi:10.1371/journal.pgen.1008185

49. Wendt FR, Carvalho CM, Pathak GA, Gelernter J, Polimanti R. Polygenic risk for autism spectrum disorder associates with anger recognition in a neurodevelopment-focused phenome-wide scan of unaffected youths from a population-based cohort. Williams SM, ed. *PLOS Genet*. 2020;16(9):e1009036. doi:10.1371/journal.pgen.1009036

50. Klei L, McClain LL, Mahjani B, et al. How rare and common risk variation jointly affect liability for autism spectrum disorder. *Mol Autism*. 2021;12(1):1-13. doi:10.1186/S13229-021-00466-2/FIGURES/5

51. Kalman JL, Loohuis LMO, Vreeker A, et al. Characterisation of age and polarity at onset in bipolar disorder. *Br J Psychiatry*. 2021;219(6):659-669. doi:10.1192/BJP.2021.102

52. Ohi K, Nishizawa D, Shimada T, et al. Polygenetic Risk Scores for Major Psychiatric Disorders Among Schizophrenia Patients, Their First-Degree Relatives, and Healthy Participants. *Int J Neuropsychopharmacol*. 2020;23(3):157-164. doi:10.1093/IJNP/PYZ073

53. Klein L, D'Urso S, Eapen V, Hwang LD, Lin PI. Exploring polygenic contributors to subgroups of comorbid conditions in autism spectrum disorder. *Sci Rep*. 2022;12(1):3416. doi:10.1038/s41598-022-07399-7

54. Maxwell JM, Coleman JRI, Breen G, Vassos E. Association Between Genetic Risk for Psychiatric Disorders and the Probability of Living in Urban Settings. *JAMA Psychiatry*. 2021;78(12):1355. doi:10.1001/jamapsychiatry.2021.2983

55. Hannon E, Schendel D, Ladd-Acosta C, et al. Elevated polygenic burden for autism is associated with differential DNA methylation at birth. *Genome Med*. 2018;10(1):19. doi:10.1186/s13073-018-0527-4

56. Peel AJ, Purves KL, Baldwin JR, et al. Genetic and early environmental predictors of adulthood self-reports of trauma. *Br J Psychiatry*. 2022;221(4):613-620. doi:10.1192/bjp.2021.207

57. Warrier V, Baron-Cohen S. Childhood trauma, life-time self-harm, and suicidal behaviour and ideation are associated with polygenic scores for autism. *Mol Psychiatry*. 2021;26(5):1670-1684. doi:10.1038/s41380-019-0550-x

58. Ratanatharathorn A, Koenen KC, Chibnik LB, Weisskopf MG, Rich-Edwards JW, Roberts AL. Polygenic risk for autism, attention-deficit hyperactivity disorder, schizophrenia, major depressive disorder, and neuroticism is associated with the experience of childhood abuse. *Mol Psychiatry*. 2021;26(5):1696-1705. doi:10.1038/s41380-020-00996-w

59. Khundrakpam B, Vainik U, Gong J, et al. Neural correlates of polygenic risk score for autism spectrum disorders in general population. *Brain Commun*. 2020;2(2):fcaa092. doi:10.1093/braincomms/fcaa092

60. Li D, Choque-Olsson N, Jiao H, et al. The influence of common polygenic risk and gene sets on social skills group training response in autism spectrum disorder. *Npj Genomic Med 2020 51*. 2020;5(1):1-8. doi:10.1038/s41525-020-00152-x

61. Morneau-Vaillancourt G, Andlauer TFM, Ouellet-Morin I, et al. Polygenic scores differentially predict developmental trajectories of subtypes of social withdrawal in childhood. *J Child Psychol Psychiatry*. 2021;62(11):1320-1329. doi:10.1111/jcpp.13459

62. Debost JCPG, Thorsteinsson E, Trabjerg B, et al. Genetic and psychosocial influence on the association between early childhood infections and later psychiatric disorders. *Acta Psychiatr Scand*. 2022;146(5):406-419. doi:10.1111/acps.13491

63. Strom NI, Smit DJA, Silzer T, et al. Meta-analysis of genome-wide association studies of hoarding symptoms in 27,651 individuals. *Transl Psychiatry*. 2022;12(1):1-8. doi:10.1038/s41398-022-02248-7

64. Riglin L, Leppert B, Langley K, et al. Investigating attention-deficit hyperactivity disorder and autism spectrum disorder traits in the general population: What happens in adult life? *J Child Psychol Psychiatry*. 2021;62(4):449-457. doi:10.1111/JCPP.13297

65. Nayar K, Sealock JM, Maltman N, et al. Elevated Polygenic Burden for Autism Spectrum Disorder Is Associated With the Broad Autism Phenotype in Mothers of Individuals With Autism Spectrum Disorder. *Biol Psychiatry*. 2021;89(5):476-485. doi:10.1016/J.BIOPSYCH.2020.08.029

66. Ding Y, Hou K, Xu Z, et al. Polygenic scoring accuracy varies across the genetic ancestry continuum. *Nature*. Published online May 17, 2023:1-8. doi:10.1038/s41586-023-06079-4

67. Grove J, Ripke S, Als TD, et al. Identification of common genetic risk variants for autism spectrum disorder. *Nat Genet 2019 513*. 2019;51(3):431-444. doi:10.1038/s41588-019-0344-8

68. Jansen AG, Dieleman GC, Jansen PR, Verhulst FC, Posthuma D, Polderman TJC. Psychiatric Polygenic Risk Scores as Predictor for Attention Deficit/Hyperactivity Disorder and Autism Spectrum Disorder in a Clinical Child and Adolescent Sample. *Behav Genet*. 2020;50(4):203-212. doi:10.1007/S10519-019-09965-8/FIGURES/1

69. Mattheisen M, Grove J, Als TD, et al. Identification of shared and differentiating genetic architecture for autism spectrum disorder, attention-deficit hyperactivity disorder and case subgroups. *Nat Genet*. 2022;54(10):1470-1478. doi:10.1038/s41588-022-01171-3

70. Schendel D, Munk Laursen T, Albiñana C, et al. Evaluating the interrelations between the autism polygenic score and psychiatric family history in risk for autism. *Autism Res*. 2022;15(1):171-182. doi:10.1002/AUR.2629

71. Trost B, Thiruvahindrapuram B, Chan AJS, et al. Genomic architecture of autism from comprehensive whole-genome sequence annotation. *Cell*. 2022;185(23):4409-4427.e18. doi:10.1016/j.cell.2022.10.009

72. Taylor MJ, Martin J, Lu Y, et al. Association of Genetic Risk Factors for Psychiatric Disorders and Traits of These Disorders in a Swedish Population Twin Sample. *JAMA Psychiatry*. 2019;76(3):280-289. doi:10.1001/JAMAPSYCHIATRY.2018.3652

73. Hjorthøj C, Uddin MJ, Wimberley T, et al. No evidence of associations between genetic liability for schizophrenia and development of cannabis use disorder. *Psychol Med*. 2021;51(3):479-484. doi:10.1017/S0033291719003362

74. Jansen AG, Jansen PR, Savage JE, et al. The predictive capacity of psychiatric and psychological polygenic risk scores for distinguishing cases in a child and adolescent

psychiatric sample from controls. *J Child Psychol Psychiatry*. 2021;62(9):1079-1089. doi:10.1111/JCPP.13370

75. Joo YY, Moon SY, Wang HH, et al. Association of Genome-Wide Polygenic Scores for Multiple Psychiatric and Common Traits in Preadolescent Youths at Risk of Suicide. *JAMA Netw Open*. 2022;5(2):e2148585. doi:10.1001/jamanetworkopen.2021.48585

76. Jørgensen CS, Horsdal HT, Rajagopal VM, et al. Identification of genetic loci associated with nocturnal enuresis: a genome-wide association study. *Lancet Child Adolesc Health*. 2021;5(3):201-209. doi:10.1016/S2352-4642(20)30350-3

77. Koomar T, Thomas TR, Pottschmidt NR, Lutter M, Michaelson JJ. Estimating the Prevalence and Genetic Risk Mechanisms of ARFID in a Large Autism Cohort. *Front Psychiatry*. 2021;12:849. doi:10.3389/FPSYT.2021.668297/BIBTEX

78. Legge SE, Jones HJ, Kendall KM, et al. Association of Genetic Liability to Psychotic Experiences With Neuropsychotic Disorders and Traits. *JAMA Psychiatry*. 2019;76(12):1256-1265. doi:10.1001/JAMAPSYCHIATRY.2019.2508

79. Legge SE, Cardno AG, Allardyce J, et al. Associations Between Schizophrenia Polygenic Liability, Symptom Dimensions, and Cognitive Ability in Schizophrenia. *JAMA Psychiatry*. 2021;78(10):1143-1151. doi:10.1001/JAMAPSYCHIATRY.2021.1961

80. Russell AE, Hemani G, Jones HJ, et al. An exploration of the genetic epidemiology of non-suicidal self-harm and suicide attempt. *BMC Psychiatry*. 2021;21(1):207. doi:10.1186/S12888-021-03216-Z/FIGURES/2
